# Nine-month, all-oral regimens for rifampin-resistant tuberculosis

**DOI:** 10.1101/2024.01.29.24301679

**Authors:** Lorenzo Guglielmetti, Uzma Khan, Gustavo E. Velásquez, Maelenn Gouillou, Amanzhan Abubakirov, Elisabeth Baudin, Elmira Berikova, Catherine Berry, Maryline Bonnet, Matteo Cellamare, Vijay Chavan, Vivian Cox, Zhanna Dakenova, Bouke Catherine de Jong, Gabriella Ferlazzo, Aydarkhan Karabayev, Ohanna Kirakosyan, Nana Kiria, Mikanda Kunda, Nathalie Lachenal, Leonid Lecca, Helen McIlleron, Ilaria Motta, Sergio Mucching-Toscano, Hebah Mushtaque, Payam Nahid, Lawrence Oyewusi, Samiran Panda, Sandip Patil, Patrick Phillips, Jimena Ruiz, Naseem Salahuddin, Epifanio Sanchez-Garavito, Kwonjune J. Seung, Eduardo Ticona, Lorenzo Trippa, Dante Vargas, Sean Wasserman, Michael L. Rich, Francis Varaine, Carole D. Mitnick

## Abstract

**Background:** After a history of poor treatments for rifampin-resistant tuberculosis (RR-TB), recent advances have resulted in shorter, more effective treatments. However, they are not available to everyone and have shortcomings, requiring additional treatment options.

**Methods:** endTB is an international, open-label, Phase 3 non-inferiority, randomized, controlled clinical trial to compare five 9-month all-oral regimens including bedaquiline (B), delamanid (D), linezolid (L), levofloxacin (Lfx) or moxifloxacin (M), clofazimine (C) and pyrazinamide (Z), to the standard (control) for treatment of fluoroquinolone-susceptible RR-TB. Participants were randomized to 9BLMZ, 9BCLLfxZ, 9BDLLfxZ, 9DCLLfxZ, 9DCMZ and control using Bayesian response-adaptive randomization. The primary outcome was favorable outcome at week 73 defined by two negative sputum culture results or by favorable bacteriologic, clinical and radiologic evolution. The non-inferiority margin was 12 percentage points.

**Results:** Of 754 randomized patients, 696 and 559 were included in the modified intention to treat (mITT) and per-protocol (PP) analyses, respectively. In mITT, the control had 80.7% favorable outcomes. Regimens 9BCLLfxZ [adjusted risk difference (aRD): 9.5% (95% confidence interval (CI), 0.4 to 18.6)], 9BLMZ [aRD: 8.8% (95%CI, −0.6 to 18.2)], and 9BDLLfxZ [3.9% (95%CI, −5.8 to 13.6)] were non-inferior in mITT and in PP. The proportion of participants experiencing grade 3 or higher adverse events was similar across the regimens. Grade 3 or higher hepatotoxicity occurred in 11.7% of the experimental regimens overall and in 7.1% of the control.

**Conclusions:** The endTB trial increases treatment options for RR-TB with three shortened, all-oral regimens that were non-inferior to a current well-performing standard of care.

ClinicalTrials.gov:NCT02754765

## Introduction

Tuberculosis resistant to rifampin (RR-TB), a key anti-tuberculosis drug, is a major global health threat. According to the World Health Organization (WHO), 410,000 people fall sick with RR-TB annually. Only 40% are diagnosed and treated, 65% of them successfully.^1^ Historically, inadequate response rates were largely due to the use of suboptimal 18- to 24-month regimens including injectables which conferred substantial toxicity.^2^ Recent advances, however, have markedly improved RR-TB treatment. New and re-purposed anti-tuberculosis drugs, supported by emerging clinical trial evidence, have led to the development of the all-oral 6-month regimen composed of bedaquiline, pretomanid, linezolid, and moxifloxacin (BPaLM),^3^ along with an alternative 9-11 month, 7-drug regimen.^4^ However, these breakthroughs are not available to everyone: BPaLM is not recommended for use in children or during pregnancy. Moreover, the 9-11-month regimen has substantial pill burden and significant toxicity; it also has inferior efficacy compared to the longer, conventional regimen.^5^

To optimize the use of newer and repurposed drugs and offer alternatives for patient-centered care, we conducted the endTB (Evaluating Newly Approved Drugs for Multidrug-resistant Tuberculosis) trial (ClinicalTrials.gov identifier NCT02754765). This Phase III clinical trial used Bayesian response-adaptive randomization^6,7^ to evaluate the efficacy and safety of five 9-month, all-oral treatment regimens compared to the evolving standard of care for fluoroquinolone-susceptible RR-TB.

## Methods

### Design and oversight

endTB is an international, multicenter, open-label Phase III, non-inferiority clinical trial conducted by the endTB consortium (Médecins Sans Frontières [MSF], Partners In Health [PIH], and Interactive Research and Development [IRD]). Full design (Figure S1) and implementation details are published^8^. The study was approved by institutional review/ethics boards at Harvard Medical School (HMS), IRD, Institute of Tropical Medicine (ITM), MSF, and at each participating site. All participants provided written informed consent.

Protocol committee members designed the trial, which was implemented in 7 countries by endTB partners (Tables S1-S2). The following groups provided oversight: Data Safety and Monitoring Board (DSMB), Scientific Advisory Committee, and Global TB Community Advisory Board (Tables S3-S5). Data were analyzed at Epicentre and validated for the primary efficacy endpoint by the University of California, San Francisco (UCSF). The MSF Pharmacovigilance Unit provided support for standardized recording, reporting, grading and classification of adverse events (Supplement). All authors contributed to writing and/or revision of the manuscript and vouch for the accuracy and completeness of the data and for the fidelity of the trial to the protocol. All study drugs were centrally purchased. The Consolidated Standards of Reporting Trials extension for adaptive design trials guided this trial report.^9^

### Participants

Individuals 15 years of age or older who had fluoroquinolone-susceptible, pulmonary RR-TB confirmed by WHO-endorsed rapid tests were enrolled at 12 sites in Georgia, India, Kazakhstan, Lesotho, Pakistan, Peru, and South Africa. Inclusion was irrespective of human immunodeficiency virus (HIV) serostatus or CD4 lymphocyte count. The trial excluded persons with baseline: pregnancy; elevated liver enzymes; uncorrectable electrolyte disorders; QT interval corrected by the Fridericia formula (QTcF) ≥450 msec or other cardiac risk factors for arrhythmia; resistance or prior exposure (≥30 days) to bedaquiline, delamanid, clofazimine, or linezolid; and ≥15 days treatment with any second-line anti-tuberculosis drug during the current TB episode.^8^ The Supplement details baseline eligibility criteria and retention in the study of participants who became pregnant.

### Randomization and treatment

The first 185 participants were randomized to one of six regimens using a fixed 1:1:1:1:1:1 ratio. Subsequently, assignment was made by Bayesian response-adaptive randomization based on 8-week culture results and 39-week efficacy outcomes. Details, including the rationale for the prior distribution used to generate the updated randomization lists, have been previously published.^6,7^ Randomization lists were updated monthly by a statistician who had no contact with trial participants or involvement in eligibility assessment. Assignment occurred through a centralized interactive randomization system. Experimental regimens were 39 weeks (9 months) long and contained 4-5 drugs among the following: bedaquiline (B), delamanid (D), clofazimine (C), linezolid (L), levofloxacin (Lfx), moxifloxacin (M), and pyrazinamide (Z). They were: 9BLMZ, 9BCLLfxZ, 9BDLLfxZ, 9DCLLfxZ, and 9DCMZ. Control group regimens reflected WHO Guidelines in effect during the trial.^5,10,11^ Treatment was administered 7 days/week, 6 under direct observation. In linezolid-containing experimental arms, linezolid dose was decreased at Week 16 or sooner if necessary to reduce toxicity. (Figure 1 and Tables S6-S7).

**Figure 1.**
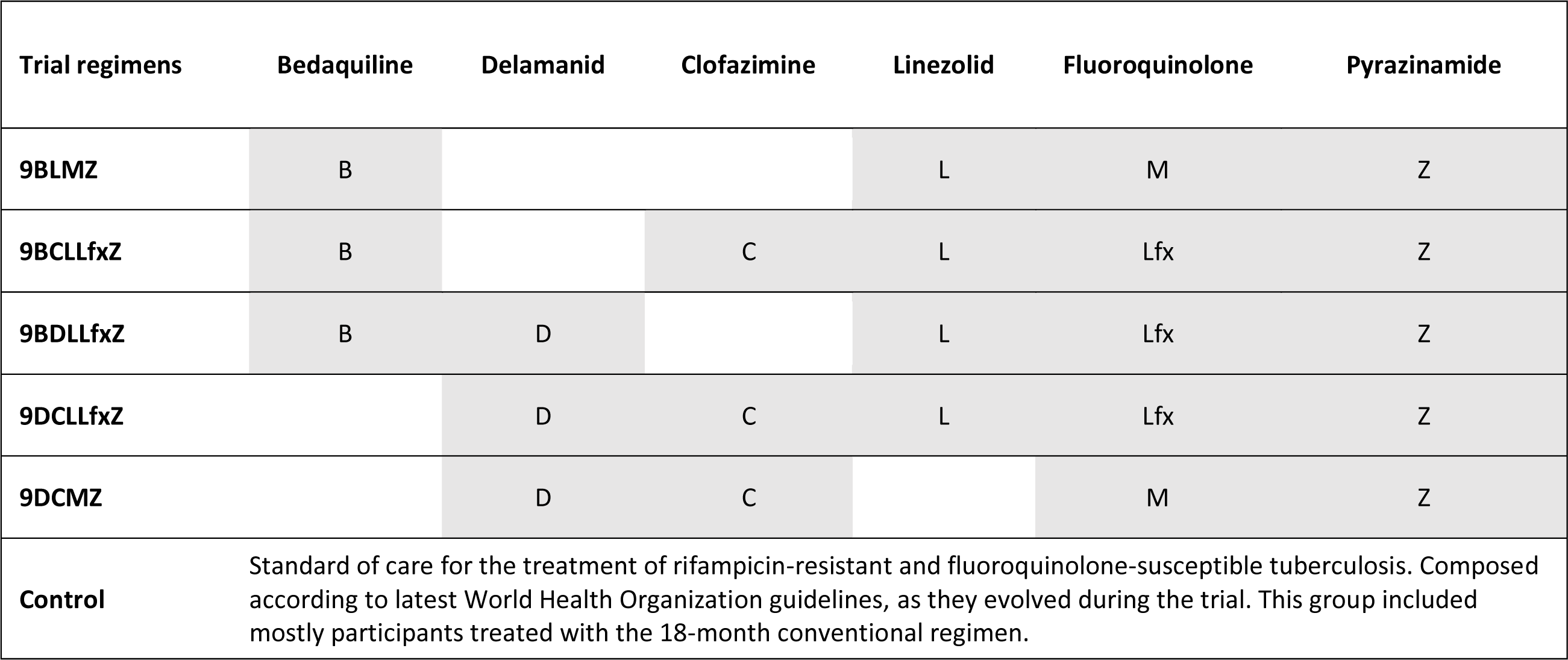
endTB Trial Experimental Regimen Composition.

### Procedures

Clinical, safety, and mycobacteriologic assessments occurred weekly until week 12, every 4 weeks until week 47, and every 6-8 weeks thereafter (Table S8). Standardized mycobacteriology tests were performed in designated, quality-controlled, trial-site laboratories; the Institute of Tropical Medicine supported site laboratories and performed additional testing. Procedures included smear microscopy and culture in Mycobacteria Growth Indicator Tube (MGIT) system at all laboratories and on solid Löwenstein-Jensen media at all laboratories except in South Africa. Phenotypic drug susceptibility (DST) testing was performed in MGIT for at least rifampin, fluoroquinolones. DST for bedaquiline, clofazimine, delamanid, and linezolid were gradually introduced. Mycobacteriology laboratory staff were blinded to treatment group assignments.

### Outcomes

Maximum follow-up was 104 weeks; follow-up ended when the final participant reached 73 weeks post-randomization. Favorable outcome at week 73 was the primary efficacy endpoint. It was established by the absence of an unfavorable outcome and either 1) two consecutive, negative cultures (one between weeks 65 and 73); or 2) favorable bacteriological, radiological, and clinical evolution. Unfavorable outcome was assigned in case of: death (from any cause); replacement/addition of one drug in the experimental arms or two drugs in the control arm; or initiation of new RR-TB treatment after the end of study treatment and before week 73. Classifications were similar for the secondary 104-week endpoint. Outcomes were adjudicated by the Clinical Advisory Committee (Supplement).

Safety outcomes were Grade 3 or higher adverse events (AEs), serious AEs (SAEs), death, discontinuation of at least one study drug due to AEs, and AEs of special interest (AESIs) defined as Grade 3 or higher: hepatotoxicity, hematologic toxicity, optic neuritis, peripheral neuropathy, or QTcF prolongation, all by week 73 (Supplement). AEs were graded by the site investigators according to the MSF Pharmacovigilance Unit Severity Scale.

### Analysis populations

Modified intention-to-treat (mITT) and per-protocol (PP) were co-primary analysis populations. The mITT population included all randomized participants who took at least one dose of study treatment (safety population) and who had a pre-randomization culture positive for *M. tuberculosis*. Participants with baseline phenotypic resistance to bedaquiline, clofazimine, delamanid, any fluoroquinolone, and/or linezolid were excluded. The PP population contains participants from the mITT population who: 1) completed a protocol-adherent course of treatment with 80% of expected doses taken within 120% of the regimen duration or did not complete because of treatment failure or death; and 2) did not receive more than 7 days of either a prohibited concomitant medication or a study drug not prescribed according to protocol. Other analysis populations are described in the Supplement.

### Statistical Analysis

Sample size assumptions included: week 73 favorable outcomes in 75% of participants in experimental groups, 70% of participants in the control, and relapse in 10%; 11% excluded in mITT and 10% in PP. A sample size of 750 afforded 80% power to establish non-inferiority (margin: −12% and one-sided type I error rate: 2.5%) of 3 experimental regimens in the mITT and 2 in the PP populations. In the efficacy analysis, we calculated the absolute between-group difference in the percentages of participants with favorable outcome at Week 73. If non-inferiority was demonstrated, superiority compared to the control was tested (p<0.05). To sequence comparisons, we used a hierarchical-testing approach (Supplement). Risk differences were estimated using a binomial regression model (generalized linear model for a binomial outcome with an identity link function). We adjusted for covariates using backward selection in pre-specified analyses (Supplement). Cox regression was used to estimate crude hazard ratios for time from randomization to unfavorable outcome and 95% confidence intervals (CI) for each experimental group. Schoenfeld residuals were used to test the proportional hazards assumption. Subgroup, sensitivity, and post-hoc analyses are described in the Supplement. We estimated the frequency of death, SAEs, AESIs and AEs of Grade 3 or higher by group. For Grade 3 or higher AEs, we also estimated the frequency of events related to a study drug. All analyses were performed in Stata version 17.0.

## Results

### Trial populations and baseline characteristics

Between February 2017 and October 2021, 1542 individuals underwent screening and 754 were randomized. Nine participants were excluded in the safety population (N=745) and 49 in the mITT population, which comprised 696 participants. The PP population included 559 participants (Figures 2 and S2).

Overall, 264 (37.9%) participants were female. Median age was 32.0 years, 25 (3.6%) were less than 18 years of age; 98 (14.1%) were living with HIV, 565 (81.2%) had sputum smear results graded 1+ or above, and 56.9% had cavitation on chest radiograph. Baseline demographic and clinical characteristics are provided in Tables 1 and S9.

Control group regimens contained at least five drugs at start in 118 (99.2%) participants. Most (114, 95.8%) were longer regimens and 97 (81.5%) included Groups A and B drugs in conformity with WHO 2022 recommendations^5^ (Tables S10-S11).

**Figure 2.**
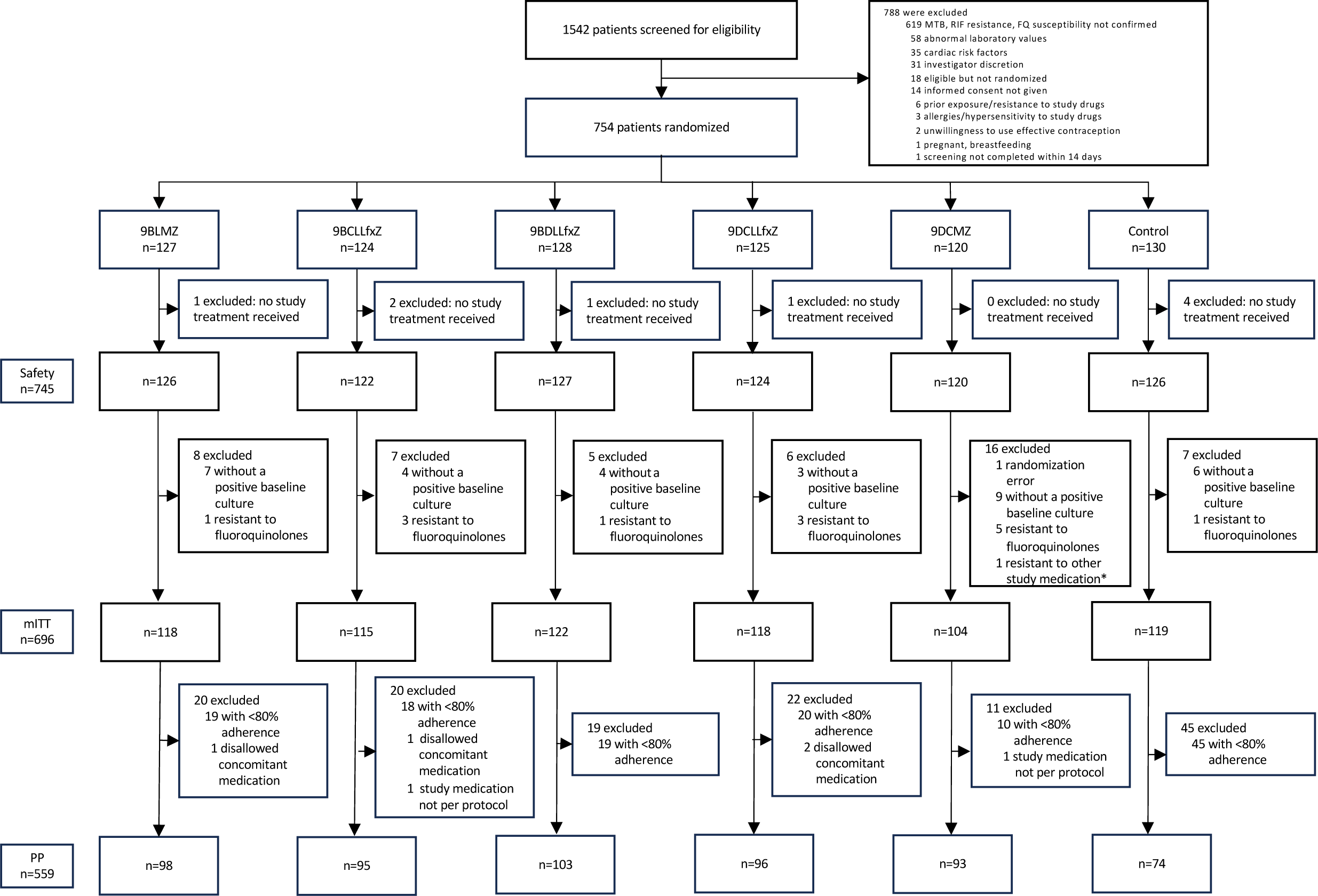
Participant flow diagram of study groups and analysis populations. MTB denotes *Mycobacterium tuberculosis*, FQ fluoroquinolone, mITT modified-intention-to-treat, PP per protocol *bedaquiline, clofazimine, delamanid and/or linezolid

**Table 1.** Baseline characteristics of participants in the mITT population. The modified-intention-to treat population included randomized participants with culture-positive, FQ-susceptible and rifampicin-resistant TB whose isolated *M. tuberculosis* strains were not determined to be resistant to bedaquiline, clofazimine, delamanid, fluoroquinolone, or linezolid. Participants who did not have a pre-treatment sputum culture positive for *M. tuberculosis* were also excluded from the modified-intention-to treat population. ECOG denotes eastern cooperative oncology group performance status, HIV human immunodeficiency virus, TB tuberculosis, HBsAg hepatitis surface antigen, and IQR interquartile range* Data on HIV, hepatitis B, hepatitis C, and diabetes were missing for one participant; ^†^data on CD4 count were unknown for 7 participants; ^‡^data on cavitation were unknown for 3 participants; ^£^extent of TB disease was defined as follows: limited = presence of lesions with slight to moderate density, but no cavitations, not exceeding the size of the apex of the lung; moderate = lesions present in one or both lungs, not exceeding a) scattered lesions of slight to moderate density that involve the total volume of one lung or partially involve both lungs, b) dense, confluent lesions that extend up to one third of the volume of one lung, and c) cavitation with a diameter of < 4 cm in any single cavity; extensive = lesions that are more extended than those defined as moderate; ^§^data on extent of TB disease, pyrazinamide and second-line injectable resistance were unknown for 5 participants; ^#^data on previous exposure to TB treatment were unknown for 25 participants; ^&^phenotypic drug susceptibility testing; ° second-line injectables are amikacin, capreomycin, and kanamycin.

| **Characteristic** | **9BLMZ**<br>**(N = 118)** | **9BCLLfxZ**<br>**(N = 115)** | **9BDLLfxZ**<br>**(N = 122)** | **9DCLLfxZ**<br>**(N = 118)** | **9DCMZ**<br>**(N = 104)** | **Control**<br>**(N = 119)** | **Total**<br>**(N = 696)** |
| --- | --- | --- | --- | --- | --- | --- | --- |
| Female sex – no. (%) | 41 (34.7%) | 37 (32.2%) | 55 (45.1%) | 38 (32.2%) | 45 (43.3%) | 48 (40.3%) | 264 (37.9%) |
| Median age, years (IQR)<br>[range] | 31.0<br>(25.0;41.0)<br>[15.0;69.0] | 38.0<br>(26.0;50.0)<br>[15.0;70.0] | 32.0<br>(22.0;45.0)<br>[15.0;70.0] | 30.5<br>(22.0;41.0)<br>[15.0;69.0] | 32.0<br>(23.5;46.0)<br>[15.0;71.0] | 31.0<br>(22.0;42.0)<br>[15.0;70.0] | 32.0<br>(23.0;44.0)<br>[15.0;71.0] |
| Study country – no. (%) |  |  |  |  |  |  |  |
| Georgia | 2 (1.7%) | 2 (1.7%) | 1 (0.8%) | 3 (2.5%) | 1 (1.0%) | 3 (2.5%) | 12 (1.7%) |
| India | 8 (6.8%) | 4 (3.5%) | 3 (2.5%) | 3 (2.5%) | 1 (1.0%) | 4 (3.4%) | 23 (3.3%) |
| Kazakhstan | 30 (25.4%) | 35 (30.4%) | 33 (27.0%) | 22 (18.6%) | 24 (23.1%) | 23 (19.3%) | 167 (24.0%) |
| Lesotho | 14 (11.9%) | 11 (9.6%) | 15 (12.3%) | 11 (9.3%) | 14 (13.5%) | 12 (10.1%) | 77 (11.1%) |
| Pakistan | 18 (15.3%) | 16 (13.9%) | 13 (10.7%) | 11 (9.3%) | 16 (15.4%) | 18 (15.1%) | 92 (13.2%) |
| Peru | 38 (32.2%) | 39 (33.9%) | 49 (40.2%) | 54 (45.8%) | 42 (40.4%) | 51 (42.9%) | 273 (39.2%) |
| South Africa | 8 (6.8%) | 8 (7.0%) | 8 (6.6%) | 14 (11.9%) | 6 (5.8%) | 8 (6.7%) | 52 (7.5%) |
| Median body mass index<br>(kg/m<sup>2</sup>) (IQR) | 19.9<br>(17.5;22.1) | 20.0<br>(18.4;23.6) | 20.9<br>(18.8;22.8) | 20.6<br>(18.1;23.6) | 19.9<br>(17.9;22.3) | 20.8<br>(17.6;23.0) | 20.4<br>(18.0;22.8) |
| ECOG – no. (n/%) |  |  |  |  |  |  |  |
| 0 | 42 (35.6%) | 35 (30.4%) | 51 (41.8%) | 47 (39.8%) | 35 (33.7%) | 43 (36.1%) | 253 (36.4%) |
| 1 | 55 (46.6%) | 62 (53.9%) | 53 (43.4%) | 54 (45.8%) | 52 (50.0%) | 63 (52.9%) | 339 (48.7%) |
| 2 | 17 (14.4%) | 15 (13.0%) | 12 (9.8%) | 16 (13.6%) | 15 (14.4%) | 11 (9.2%) | 86 (12.4%) |
| 3 | 4 (3.4%) | 3 (2.6%) | 6 (4.9%) | 1 (0.9%) | 2 (1.9%) | 2 (1.7%) | 18 (2.6%) |
| HIV, positive* – no. (%) | 15 (12.7%) | 14 (12.2%) | 17 (13.9%) | 18 (15.3%) | 15 (14.4%) | 19 (16.0%) | 98 (14.1%) |
| Median CD4 count among HIV-positive participants† (IQR) | 170.5<br>(41.0;505.0) | 190<br>(85.0;377.0) | 314.5<br>(157.0;478.5) | 328.5<br>(170.5;579.5) | 404.0<br>(143.0;643.0) | 269.0<br>(83.0;443.0) | 296.0<br>(118.0;497.0)<br>(N=91) |
| Antiretroviral treatment among HIV-positive participants | 10 (66.7%) | 8 (57.1%) | 9 (52.9%) | 12 (66.7%) | 11 (73.3%) | 11 (57.9%) | 61 (62.2%) |
| Hepatitis B, HbsAg positive* – no. (%) | 3 (2.5%) | 3 (2.6%) | 0 (0.0%) | 2 (1.7%) | 4 (3.8%) | 4 (3.4%) | 16 (2.3%) |
| Hepatitis C, positive* – no. (%) | 5 (4.2%) | 5 (4.3%) | 3 (2.5%) | 4 (3.4%) | 3 (2.9%) | 6 (5.0%) | 26 (3.7%) |
| Diabetes* – no. (%) | 19 (16.1%) | 18 (15.7%) | 20 (16.4%) | 16 (13.6%) | 16 (15.4%) | 15 (12.6%) | 104 (14.9%) |
| Smear result – no. (%) |  |  |  |  |  |  |  |
| Negative/Scanty | 20 (16.9%) | 19 (16.5%) | 31 (25.4%) | 24 (20.3%) | 18 (17.3%) | 19 (16.0%) | 131 (18.8%) |
| 1-2+ | 57 (48.3%) | 59 (51.3%) | 58 (47.5%) | 49 (41.5%) | 43 (41.3%) | 52 (43.7%) | 318 (45.7%) |
| 3+ | 41 (34.7%) | 37 (32.2%) | 33 (27.0%) | 45 (38.1%) | 43 (41.3%) | 48 (40.3%) | 247 (35.5%) |
| Cavitation‡ – no. (%) | 68 (57.6%) | 69 (60.0%) | 73 (59.8%) | 53 (44.9%) | 58 (55.8%) | 75 (63.0%) | 396 (56.9%) |
| Extent of TB disease<sup>£§</sup> – no. (%) |  |  |  |  |  |  |  |
| Limited | 21 (17.8%) | 14 (12.2%) | 18 (14.8%) | 23 (19.5%) | 20 (19.2%) | 18 (15.1%) | 114 (16.4%) |
| Moderate | 70 (59.3%) | 77 (67.0%) | 77 (63.1%) | 67 (56.8%) | 62 (59.6%) | 71 (59.7%) | 424 (60.9%) |
| Extensive | 27 (22.9%) | 24 (20.9%) | 26 (21.3%) | 25 (21.2%) | 22 (21.2%) | 29 (24.4%) | 153 (22.0%) |
| Prior exposure to TB treatment# - no. (%) |  |  |  |  |  |  |  |
| None | 76 (64.4%) | 67 (58.3%) | 78 (63.9%) | 80 (67.8%) | 70 (67.3%) | 74 (62.2%) | 445 (63.9%) |
| First-line drugs only | 20 (16.9%) | 23 (20.0%) | 27 (22.1%) | 25 (21.2%) | 22 (21.2%) | 31 (26.1%) | 148 (21.3%) |
| Other drugs | 15 (12.7%) | 19 (16.5%) | 15 (12.3%) | 7 (5.9%) | 11 (10.6%) | 11 (9.2%) | 78 (11.2%) |
| Pyrazinamide resistance<sup>§</sup> – no. (%) | 57 (48.3%) | 63 (54.8%) | 66 (54.1%) | 66 (55.9%) | 63 (60.6%) | 59 (49.6%) | 374 (53.7%) |
| Second-line injectable resistance<sup>°§</sup> – no. (%) | 14 (11.9%) | 18 (15.7%) | 15 (12.3%) | 13 (11.0%) | 13 (12.5%) | 16 (13.4%) | 89 (12.8%) |

### Efficacy results

In the primary outcome analysis of the control group, favorable outcomes occurred in 80.7% (95% CI, 72.4 to 87.3) in the mITT and in 95.9% (95%CI, 88.6 to 99.2) in the PP populations. Three experimental groups (9BLMZ, 9BCLLfxZ, and 9BDLLfxZ) were non-inferior to the control in both mITT and PP populations. In prespecified adjusted analyses, 9BCLLfxZ was superior to the control in mITT [adjusted risk difference (aRD): 9.5% (95%CI, 0.4 to 18.6)] and non-inferior in the PP [aRD: 0.3% (95%CI, - 6.1% to 6.7%). The 9BLMZ and 9BDLLfxZ groups were non-inferior to the control in the mITT [aRD: 8.8% (95%CI, −0.6 to 18.2) and 3.9% (95%CI, −5.8 to 13.6) respectively] and in the PP [aRD: 0.1% (95%CI, −6.2 to 6.4) and −2.9% (95%CI, −10.2 to 4.4) respectively] populations. The 9DCMZ group was non-inferior to the control in the mITT (aRD: 4.4% (95%CI, −5.7 to 14.5) and not non-inferior in the PP (aRD: −8.0% (95%CI, −16.2 to 0.2) populations. The 9DCLLfxZ group was not non-inferior to the control both in mITT and PP (Table 2, Figure 3).

Unfavorable outcomes due to positive culture results and unfavorable evolution occurred in 3.7% of the full cohort and in 10.2% of the 9DCLLfxZ group (Table 2). Loss to follow-up and consent withdrawal were more frequent in the control group than in any experimental regimen. Overall, recurrence occurred in 3 (0.4%) participants. Efficacy outcomes were similar at secondary endpoints, week 39 (Tables S12-S15) and week 104 (Figure 3).

Post-hoc adjusted analysis and sensitivity analyses at 73 weeks yield similar results to primary analysis (Tables S16-S18). Overall, treatment effects at 73 weeks did not differ importantly in subgroup analyses in the mITT population. Possible exceptions for some groups include: country, prior exposure to second-line anti-TB drugs, cavitation, HIV coinfection, and low body mass index. Outcomes generally improved, while relative treatment effect did not change meaningfully, over the study period; (Figures S3a-e).

In the 9BCLLfxZ group, time to unfavorable outcome was longer than in the control (p=0.039, log-rank test; hazard ratio=0.48 [95% CI: 0.23-0.98]), while the other groups were similar to the control (Figures S4a-e).

**Table 2.**
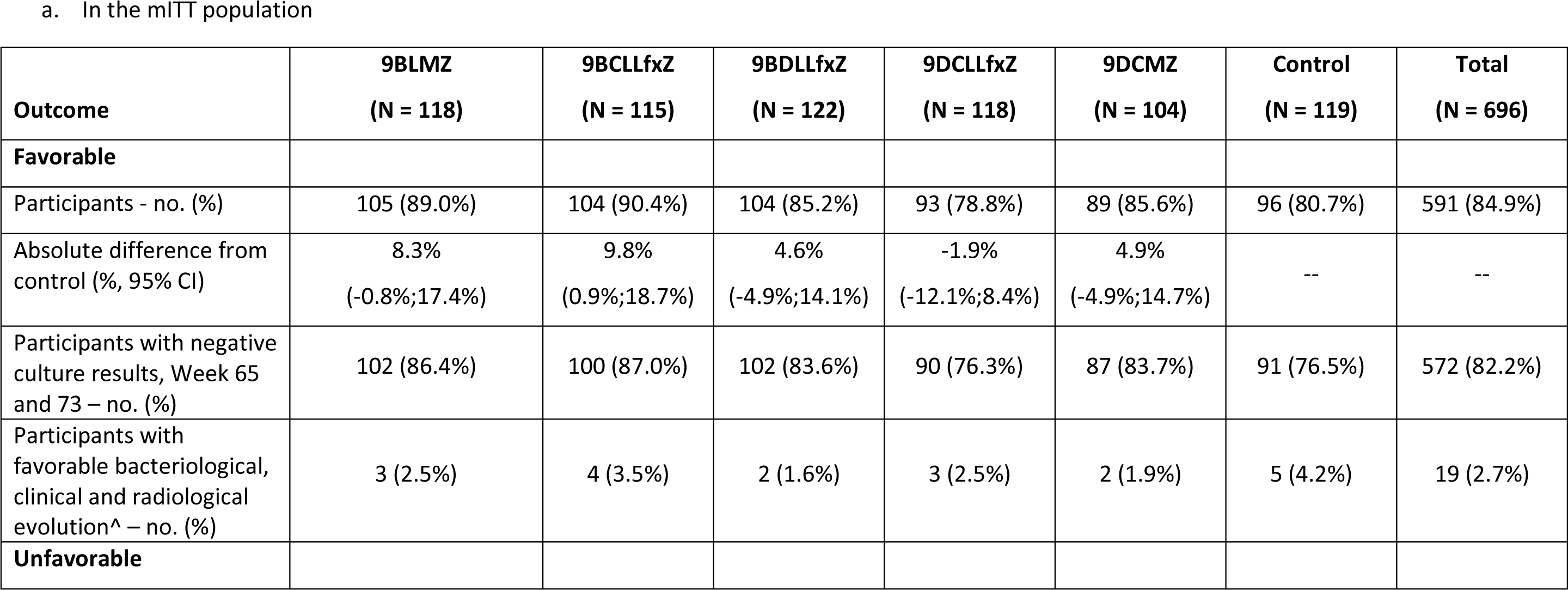
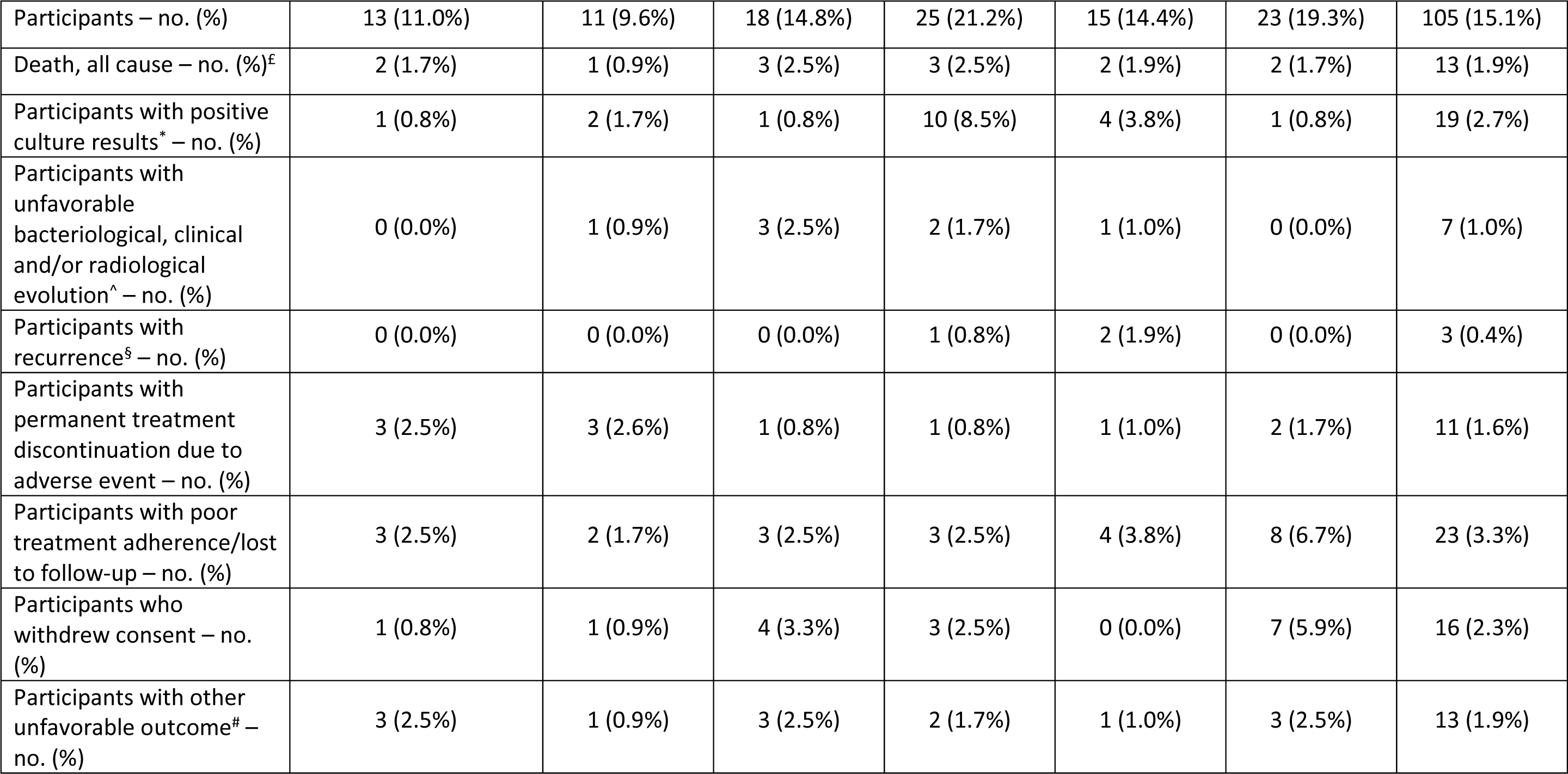
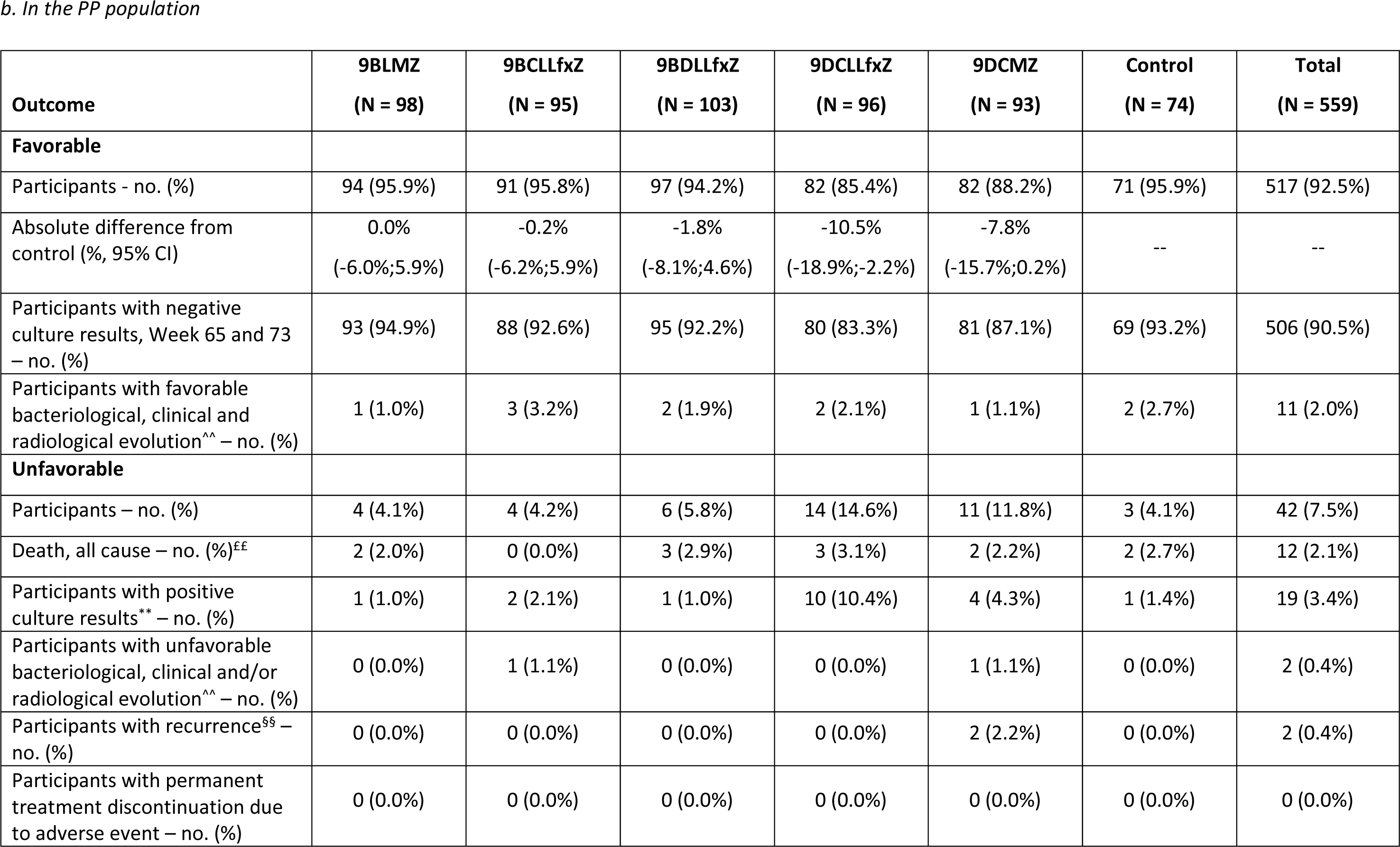
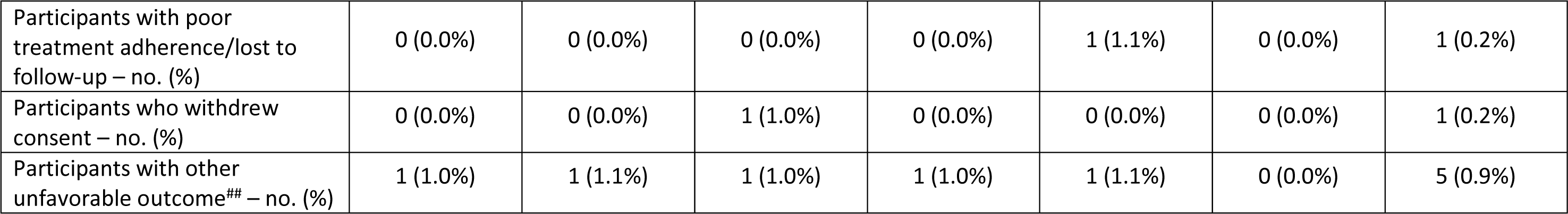
Primary efficacy outcomes at Week 73 in the modified-intention-to-treat and per-protocol populations. Part a presents outcomes at Week 73 in the modified-intention-to-treat population. The modified-intention-to treat population included randomized participants with culture-positive, FQ-susceptible and rifampicin-resistant TB whose isolated *M. tuberculosis* strains were not determined to be resistant to bedaquiline, clofazimine, delamanid, fluoroquinolone, or linezolid. Participants who did not have a pre-treatment sputum culture positive for *M. tuberculosis* were also excluded from the modified-intention-to treat population. ^ = participants with missing culture results from Week 65 to Week 73; ^£^=13 mITT participants experienced death as a treatment outcome, 1 participant in the safety population who was excluded from the mITT population also experienced death. 1 participant in the mITT population was assigned positive culture result as unfavorable outcome at 73 weeks and later died. * = participants who permanently discontinued treatment because of a positive sputum culture at Week 16 or later, or with a positive sputum culture at Week 65 or Week 73; § = participants with a positive sputum culture or who started a new treatment regimen after treatment completion; # = participants with other unfavorable outcome: not assessable after completing treatment (n=6), investigator’s judgement (n=4), pregnancy or breastfeeding (n=2), use of prohibited concomitant medication (n=1). Part b presents outcomes at Week 73 in the per-protocol population. The per-protocol population, was the modified-intention-to treat population excluding participants who, for reasons other than treatment failure or death, do not complete a protocol-adherent course of treatment. A protocol-adherent course of treatment was 80% of expected doses taken within 120% of the intended regimen duration. Participants who received more than 7 days of either a prohibited concomitant medication or a study drug not prescribed according to protocol were also be excluded from the per-protocol population. ^^^^participants with missing culture results from Week 65 to Week 73; ^££^=12 PP participants experienced death as a treatment outcome, 1 participant in the safety population who was excluded from the mITT population also experienced death. 1 participant in the mITT population was assigned positive culture result as unfavorable outcome at 73 weeks and later died; 1 death occurred in a participant in the mITT population who was excluded from PP population for receiving less than 80% of doses in 120% of treatment duration. **participants who permanently discontinued treatment because of a positive sputum culture at Week 16 or later, or with a positive sputum culture at Week 65 or Week 73; ^§§^participants with a positive sputum culture or who started a new treatment regimen after treatment completion; ^##^participants with other unfavorable outcome: not assessable after completing treatment (5).

**Figure 3.**
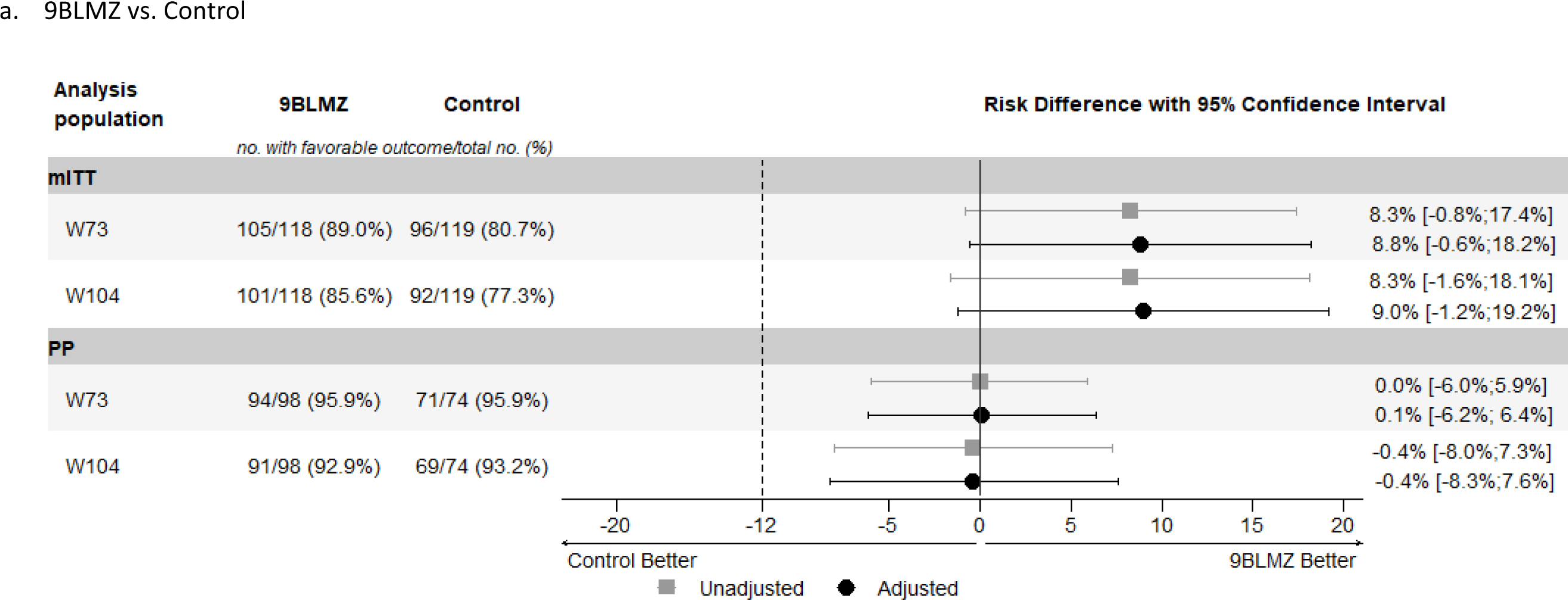
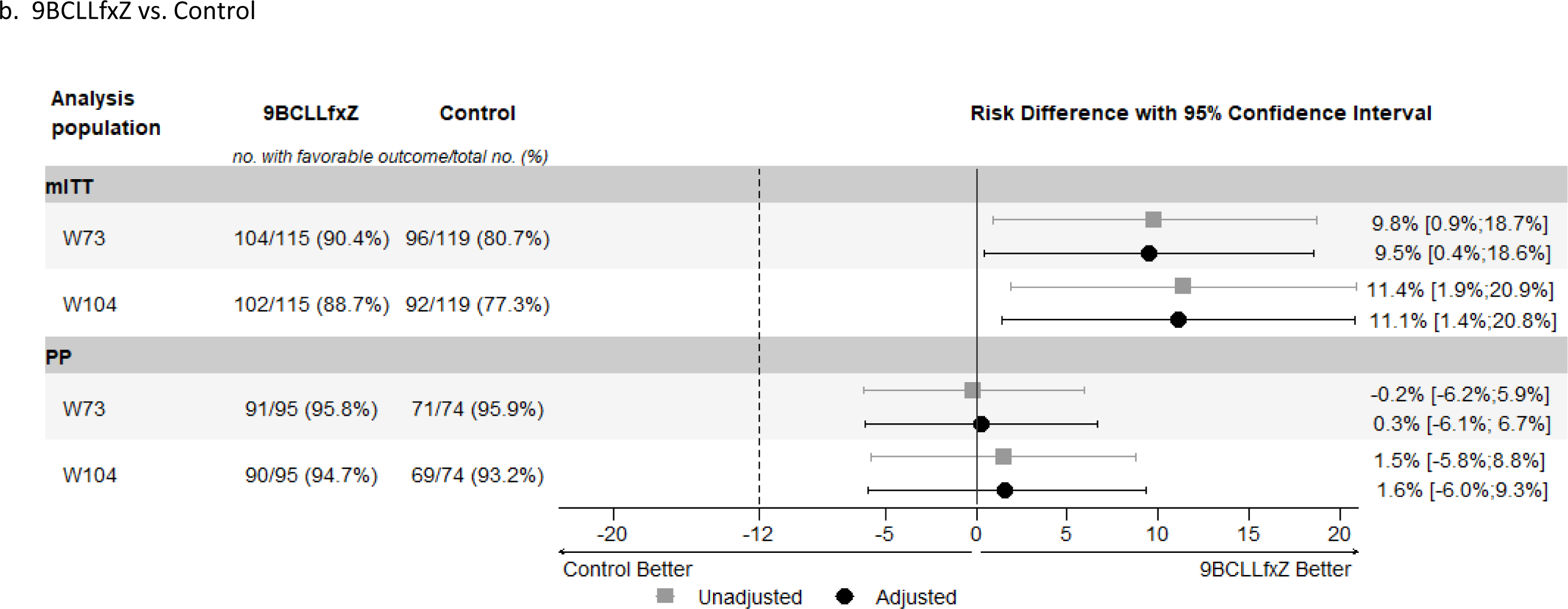
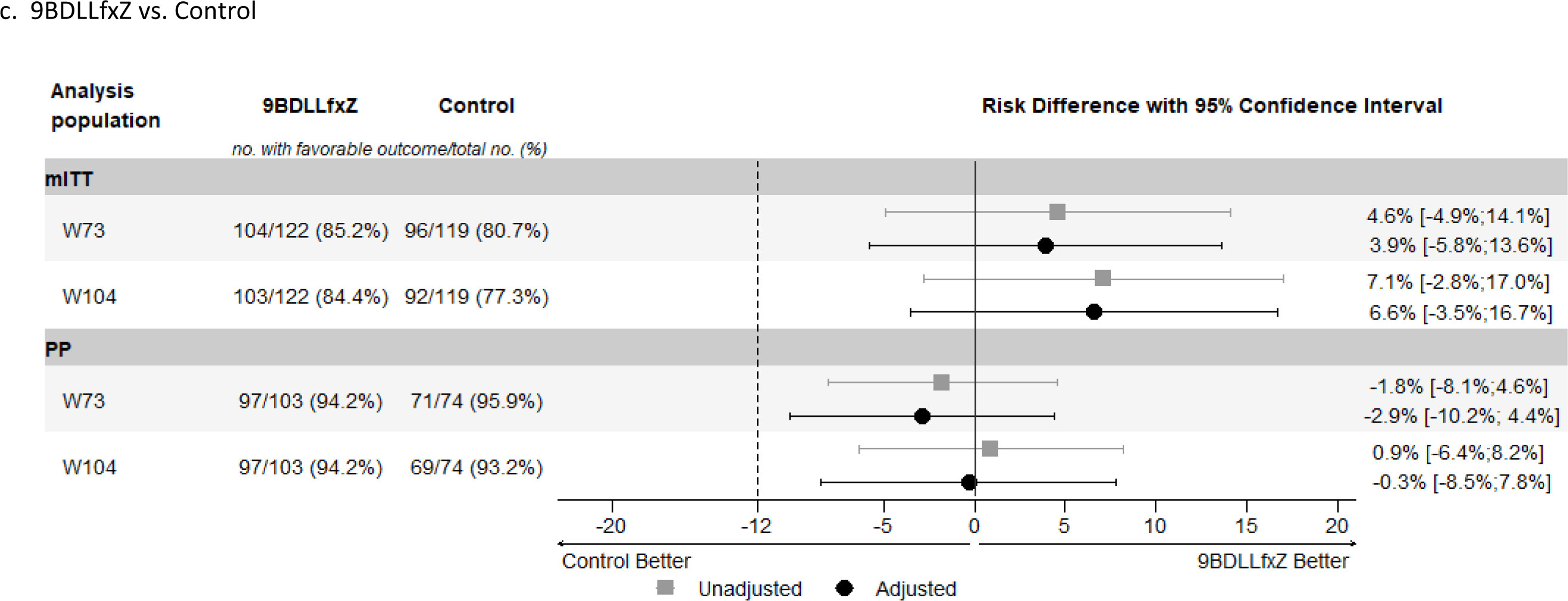
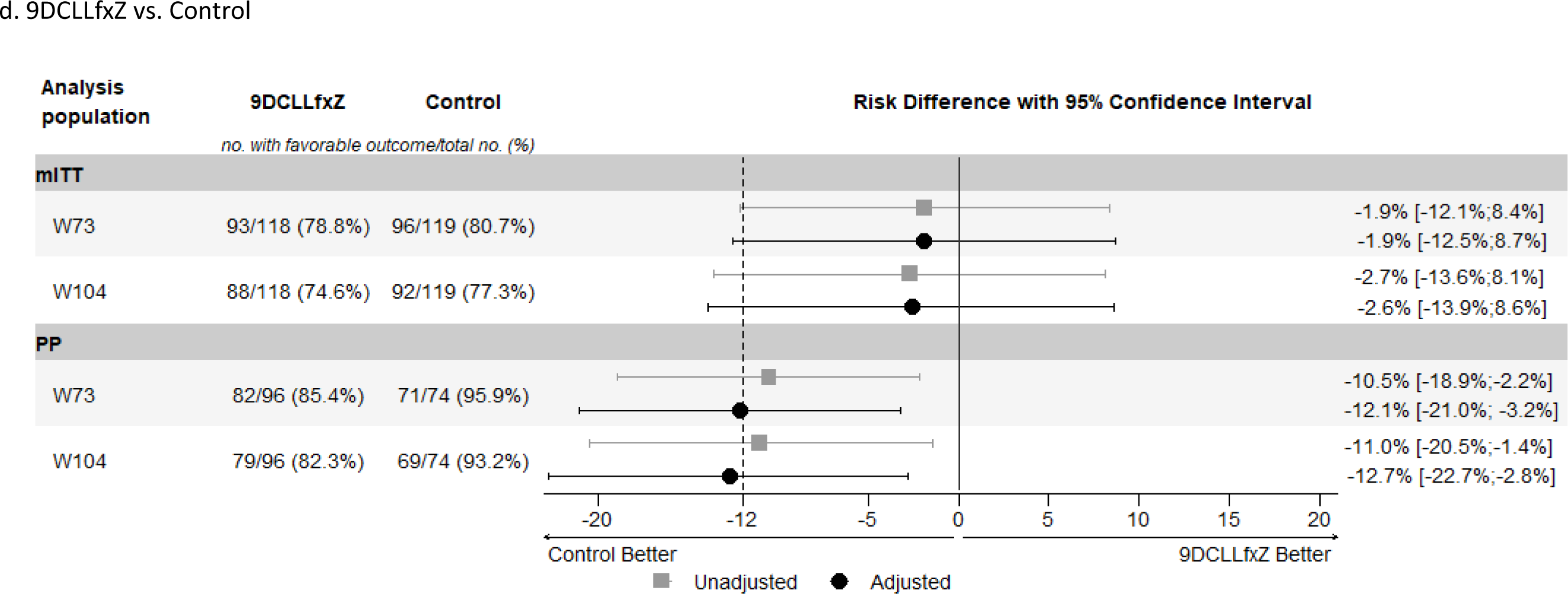
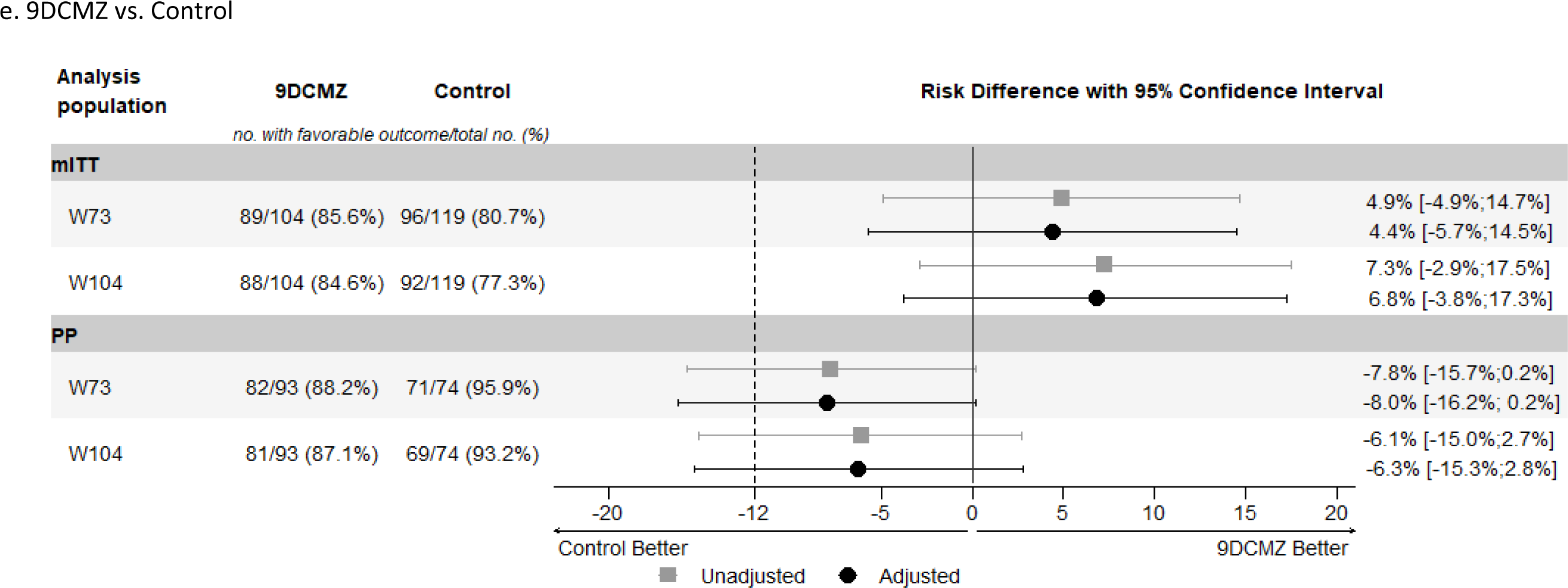
Primary and secondary efficacy analyses at Week 73 and Week 104, by experimental group versus control. Shows the results of the primary efficacy analysis in the modified-intention-to treat and the per-protocol analysis populations (a. 9BLMZ vs. control, b. 9BCLLfxZ vs. control, c. 9BDLLfxZ vs. control, d. 9DLLfxZ vs. control, e. 9DCMZ vs. control. The noninferiority margin of −12 percentage points is designated by the dashed vertical line. Participants were classified as having a favorable outcome at week 73 if one of the following was true: 1) their last two culture results were negative and were taken from sputum samples collected on separate visits, the latest between Week 65 and Week 73; 2) the last culture result (from a sputum sample collected between Weeks 65 and 73) was negative and either there was no other post-baseline culture result or the penultimate culture result was positive due to laboratory cross contamination; and bacteriological, radiological and clinical evolution is favorable; or 3) there was no culture result from a sputum sample collected between Week 65 and Week 73 or the result of that culture was positive due to laboratory cross contamination, and the most recent culture result was negative, and bacteriological, radiological and clinical evolution was favorable. Participants were classified as having a favorable outcome at week 104 if one of the following was true: 1) The last two cultures are negative and from sputum samples collected on separate visits, the latest between Week 97 and Week 104; 2) The last culture result (from a sputum sample collected between Week 97 and Week 104) was negative; and either there was no other post-baseline culture result or the penultimate culture result as positive due to laboratory cross contamination; and bacteriological, radiological and clinical evolution was favorable; or 3) there was no culture result from a sputum sample collected between Week 97 and Week 104 or the result of that culture was positive due to laboratory cross contamination; and the most recent culture result was negative; and bacteriological, radiological and clinical evolution was favorable. The modified-intention-to treat population included randomized participants with culture-positive, FQ-susceptible and rifampicin-resistant TB whose isolated *M. tuberculosis* strains were not determined to be resistant to bedaquiline, clofazimine, delamanid, fluoroquinolone, or linezolid. Participants who did not have a pre-treatment sputum culture positive for *M. tuberculosis* were also excluded from the modified-intention-to treat population. The per-protocol population was the modified-intention-to treat population excluding participants who, for reasons other than treatment failure or death, do not complete a protocol-adherent course of treatment. A protocol-adherent course of treatment was 80% of expected doses taken within 120% of the intended regimen duration. Participants who received more than 7 days of either a prohibited concomitant medication or a study drug not prescribed according to protocol were also be excluded from the per-protocol population. mITT (modified-intention-to-treat) analyses at weeks 73 and 104 were adjusted (prespecified) by: Hepatitis C, extent of disease; PP (per-protocol) analyses at weeks 73 and 104 were adjusted (prespecified) by: Age, BMI.

### Safety results

We report the number of participants in the safety population who experienced at least one of each safety event by Week 73 after randomization. The number with at least one Grade 3 or higher AE ranged from 54.8% (9BLMZ) to 61.4% (9BDLLfxZ) in experimental groups and was 62.7% in the control. SAE frequency was similar across groups: it ranged from 13.1% (9BCLLfxZ) to 16.7% (9DCMZ) of participants in experimental groups and was 16.7% in the control. Overall, death from any cause occurred in 15 (2.0%) participants by 73 weeks and 18 participants (2.4%) by 104 weeks; frequency was similar across treatment groups (Table 3). No deaths were considered related to study drugs (Table S26).

Among all Grade 3 or higher AEs and SAEs, 313/901 (34.7%) and 54/174 (31.0%), respectively were classified as related to study drugs. At least one AESI was reported in 23.9% of all participants: the most frequent, hepatotoxicity, occurred in 7.1% of the control and its frequency in experimental groups ranged from 6.3% in 9BDLLfxZ to 18.3% in 9BLMZ. Hematologic toxicity classified as an AESI occurred in 10.3% of control participants; in experimental groups, it ranged from 7.4% (9BCLLfxZ) to 10.5% (9DCLLfxZ). Peripheral neuropathy occurred in 4.8% in control and ranged from 2.4% (9DCLLfxZ) to 7.1% (9BDLLfxZ) in experimental groups. QTcF interval prolongation occurred exclusively in groups 9DCMZ (4.2%) and 9BCLLfxZ (3.3%). Other safety details, including drug discontinuations and deaths, are reported in Tables S20-S27. Ten (1.3%) participants became pregnant during study participation (Table S28).

**Table 3.** Safety analysis at Week 73 in the Safety population. Safety population is all randomized participants who had received at least one dose of study treatment. ^related is defined as at least a reasonable possibility to be caused by one or many drugs in the regimen; * QT interval corrected according to the Fridericia formula. ALT denotes alanine transaminase, AST aspartate aminotransferase

|  | **9BLMZ**<br>**(N = 126)** | **9BCLLfxZ**<br>**(N = 122)** | **9BDLLfxZ**<br>**(N = 127)** | **9DCLLfxZ**<br>**(N = 124)** | **9DCMZ**<br>**(N = 120)** | **Control**<br>**(N = 126)** | **Total**<br>**(N = 745)** |
| --- | --- | --- | --- | --- | --- | --- | --- |
| Participants with any adverse event – no. (%) | 126 (100.0%) | 122 (100.0%) | 127 (100.0%) | 124 (100.0%) | 120 (100.0%) | 125 (99.2%) | 744 (99.9%) |
| Grade 3 or higher adverse events |  |  |  |  |  |  |  |
| Participants with ≥1 event–no. (%) | 69 (54.8%) | 68 (55.7%) | 78 (61.4%) | 75 (60.5%) | 72 (60.0%) | 79 (62.7%) | 441 (59.2%) |
| No. of events | 136 | 166 | 144 | 148 | 148 | 163 | 901 |
| No. of events related to study drug(s) (% of all events)^ | 49 (36.3%) | 57 (34.3%) | 56 (38.9%) | 58 (40.2%) | 37 (25.0%) | 56 (34.4%) | 313 (34.7%) |
| Serious adverse events |  |  |  |  |  |  |  |
| Participants with ≥1 event–no. (%) | 18 (14.3%) | 16 (13.1%) | 20 (15.8%) | 18 (14.5%) | 20 (16.7%) | 21 (16.7%) | 113 (15.2%) |
| No. of events | 26 | 29 | 30 | 26 | 31 | 32 | 174 |
| No. of events related to study drug(s) (% of all events)^ | 7 (26.9%) | 11 (37.9%) | 11 (36.7%) | 11 (42.3%) | 6 (19.4%) | 8 (25.0%) | 54 (31.0%) |
| Death from any cause – no. (%) | 3 (2.4%) | 1 (0.8%) | 3 (2.4%) | 4 (3.2%) | 2 (1.7%) | 2 (1.6%) | 15 (2.0%) |
| Adverse event of special interest |  |  |  |  |  |  |  |
| Participants with ≥1 event–no. (%) | 35 (27.8%) | 33 (27.1%) | 25 (19.7%) | 33 (26.6%) | 26 (21.7%) | 26 (20.6%) | 178 (23.9%) |
| Participants with any Grade 3-4 increase in ALT or AST – no. (%) | 23 (18.3%) | 17 (13.9%) | 8 (6.3%) | 18 (14.5%) | 12 (10.0%) | 9 (7.1%) | 87 (11.7%) |
| Participants with any Grade 3-4 leukopenia, anemia, or thrombocytopenia – no. (%) | 11 (8.7%) | 9 (7.4%) | 10 (7.9%) | 13 (10.5%) | 9 (7.5%) | 13 (10.3%) | 65 (8.7%) |
| Participants with any Grade 3-4 peripheral neuropathy – no. (%) | 4 (3.2%) | 5 (4.1%) | 9 (7.1%) | 3 (2.4%) | 3 (2.5%) | 6 (4.8%) | 30 (4.0%) |
| Participants with any Grade 3-4 optic neuritis – no. (%) | 0 (0.0%) | 1 (0.8%) | 0 (0.0%) | 1 (0.8%) | 0 (0.0%) | 2 (1.6%) | 4 (0.5%) |
| Participants with any Grade 3-4 QT corrected* interval prolonged – no. (%) | 0 (0.0%) | 4 (3.3%) | 0 (0.0%) | 0 (0.0%) | 5 (4.2%) | 0 (0.0%) | 9 (1.2%) |
| Participants with permanent discontinuation of any drug due to adverse event – no. (%) | 29 (23.0%) | 32 (26.2%) | 41 (32.3%) | 32 (25.8%) | 22 (18.3%) | 54 (42.9%) | 210 (28.2%) |

## Discussion

This phase 3 trial identified three regimens (9BLMZ, 9BCLLfxZ, and 9BDLLfxZ) with robust evidence of non-inferior efficacy compared to the standard of care. 9BCLLfxZ was also superior to the control. These three regimens—and a fourth (9DCMZ) that was non-inferior only in the mITT population— produced favorable outcomes in more than 85% of participants at week 73.

Death was uncommon despite the substantial burden of comorbidities and cavitary disease. Grade 3 or higher AEs were frequent across all groups, but often unrelated to study drugs. Although the study was not powered for statistical comparison of safety outcomes, we observed some patterns. Grade 3 or higher hepatoxicity was more common in experimental groups, except in 9BDLLfxZ, than in the control. Pyrazinamide, included in all experimental groups and almost 50% of the control regimens, can cause elevated liver enzymes, as can bedaquiline, fluoroquinolones, and linezolid; this can be aggravated by alcohol use and active hepatitis B or C infection, which were present in the cohort.^12–14^ Linezolid-related toxicities were generally less frequent in experimental groups than in the control and this may reflect the safety benefit of routinely lowering the weekly dose of linezolid at 16 weeks or earlier.^15–18^ QTcF intervals of >500 ms were infrequent and occurred only in groups containing clofazimine and a second significant QT prolonger, bedaquiline or moxifloxacin. These results are consistent with emerging evidence about the safety of bedaquiline in combination with other QT-prolonging anti-TB drugs.^3,19,20^ Bayesian response-adaptive randomization was successful in identifying multiple non-inferior TB regimens in a single study. Randomization was ultimately relatively balanced because experimental regimens performed similarly to the control on the interim endpoints used to adjust probabilities. Improved surrogate markers for treatment response will permit increased efficiency of adaptive trials in _TB.21,22_

There are several limitations. Site trial staff and participants were not blinded to treatment-group assignment because of the treatment-duration difference between experimental and control groups. To mitigate risks of bias, we concealed treatment assignment and randomization probabilities from laboratory staff and central investigators. Bayesian adaptation and analysis for DSMB reports were performed by unblinded statisticians. During the trial enrollment period, WHO Guidelines changed twice. We incorporated these updates in trial guidance on the composition of control group regimens. However, the impact on regimen composition was modest because the initial trial guidance was already well aligned with current WHO recommendations. As a result, 81.5% of control regimens conformed with the most recent WHO recommendation.^5^

Strengths of the trial include its randomized, internally, concurrently controlled design, which is essential to high certainty of evidence for guidance.^23^ Other strengths include the consistency of the findings across populations, endpoints, and analyses. Moreover, the control group performance, 80.7% favorable outcome, was better than that reported in other recent studies.^3,4,24–26^ That non-inferiority could be established against this improved standard—and ruled out for 9DCLLfxZ at 78.8% favorable outcomes—provides confidence in the efficacy of the new regimens. High retention of participants— including in the control group—and completeness of study data indicate high-quality implementation. The trial included adolescents and retained pregnant women. The population was heterogeneous, representing 4 continents, a range of TB disease severity, and substantial burdens of important comorbidities, all contributing to generalizability of study results to the broader population of people affected by MDR/RR-TB.

These findings support the use of up to four new, all-oral, shorter MDR/RR-TB regimens in addition to BPaLM. The latter was recommended by WHO in 2022 for use in non-pregnant people of 14 years of age or older.^5^ 9BLMZ, 9BCLLfxZ, and 9BDLLfxZ could be used in nearly all adults, children, and pregnant women with fluoroquinolone-susceptible MDR/RR-TB; all drugs in the endTB regimens have pediatric formulations and are recommended regardless of age.^27,28^ Findings are also relevant to pregnant women: all drugs included in the non-inferior endTB regimens are considered to be acceptable for use during pregnancy.^5,29^ A fourth regimen, 9DCMZ, could be an alternative for people who have fluoroquinolone-susceptible MDR/RR-TB and contraindications to bedaquiline and linezolid. 9DCMZ was non-inferior in the mITT but not in the PP analysis. Most (81.5%) control group participants received bedaquiline and/or linezolid. Future work to assess the efficacy of this regimen relative to that of a contemporaneous linezolid- and bedaquiline-sparing comparator (in a population not able to receive these drugs) would be more informative.

Several implementation considerations arise. Country drug formularies can be vastly simplified, while still offering a range of treatment options. Further development of—and access to—rapid, reliable, resistance testing is essential both to optimize patient selection for these regimens and to detect emergence of resistance.^30,31^ Finally, this study underscores the need for diligent monitoring of liver enzymes to reduce risk.^5,13,28^ Hepatotoxicity is a known risk with many anti-TB drugs. endTB participants experienced a significant rate of hepatotoxicity, partly attributable to pyrazinamide’s inclusion in all experimental regimens. The same applies to linezolid: monitoring for known associated toxicities is essential to assure good outcomes. Conversely, QT interval prolongation monitoring could be optimized through risk-based strategies, focusing on persons receiving multiple QT-prolonging drugs or with arrhythmia risk factors.^32,33^

The endTB trial significantly increases treatment options for MDR/RR-TB for a broad range of patients with all-oral regimens that are shorter and simpler than—and non-inferior to—a well-performing standard of care.

## Supporting information

endTB Protocol v3.6

endTB SAP v2.0

## Data Availability

All data produced in the present study are available upon reasonable request to the authors.

