## Supplementary material for "Nine-month, all-oral regimens for rifampin-resistant tuberculosis": endTB Protocol v3.6

### (Evaluating Newly approved Drugs for multidrug-resistant TB)

Protocol Number: NCT02754765 (ClinicalTrials.gov)

Sponsor: Médecins Sans Frontières –France

Funding Sponsor: UNITAID

Principal Investigators: Carole Mitnick, Sc.D. and Lorenzo Guglielmetti, M.D.

Version Number: 3.6

Date: 20 July 2022

| Version number | Date |
|---|---|
| 1.0 | 21-Sep-2015 |
| 2.0 | 02-Feb-2016 |
| 2.1 | 07-Apr-2016 |
| 2.2 | 11-Aug-2016 |
| 2.3 | 05-Oct-2016 |
| 2.4 | 21-Nov-2016 |
| 2.5 | 09-Feb-2017 |
| 2.6 | 09-May-2017 |
| 3.0 | 12-Feb-2018 |
| 3.1 | 29-Aug-2018 |
| 3.2 | 23-Oct-2018 |
| 3.3 | 14-Feb-2019 |
| 3.4 | 31-Jul-2020 |
| 3.5 | 15-Dec-2020 |
| 3.6 | 20-Jul-2022 |

| Version number | Date | Summary of changes |
|---|---|---|
| 1.0 | 21-Sep-2015 | - Initial version |
| 2.0 | 02-Feb-2016 | - Increase of the sample size to 750 subjects<br>- Modifications to the experimental regimens endTB 1 and endTB 3<br>- Modifications of ECG monitoring frequency to weekly for the first 3 months<br>- Updates to Background Information and Scientific Rationale<br>- Modifications to the study eligibility criteria, study procedures, and the schedule of event<br>- Removal of the inclusion of pregnant women and women who are breastfeeding<br>- Discontinuation of study treatment when a female participant is found to be pregnant<br>- Updates to Concomitant Medications/Treatments<br>- Incorporation of comments from the Institutional Review Boards and Ethics Committees<br>- Other minor editorial and typographical updates and corrections |
| 2.1 | 07-Apr-2016 | - Change to the protocol title to clarify the approval status of the new drugs<br>- Updates to the investigator site information<br>- Modifications to the study outcome measures to ensure mutually exclusive classifications<br>- Clarification of the study procedures, and the schedule of events<br>- Addition of ethambutol to the list of Investigational Products<br>- Other minor editorial and typographical updates and corrections |
| 2.2 | 11-Aug-2016 | - Addition of the clinicaltrials.gov protocol number<br>- Change in study procedure from triplicate ECGs to duplicate ECGs<br>- Updates to the investigator site information<br>- Clarification of the study outcome measures<br>- Modification to the study exclusion criteria<br>- Clarification of the study procedures, visit window, and the schedule of events<br>- Clarification of pregnancy reporting for sites<br>- Other minor editorial and typographical updates and corrections |
| 2.3 | 05-Oct-2016 | - Change in study procedure from duplicate to triplicate ECGs for screening and baseline visits<br>- Modification of the study prescreening procedures<br>- Other minor editorial and typographical updates and correction |
| 2.4 | 21-Nov-2016 | - Clarification of subject exclusion criteria<br>- Other minor editorial and typographical updates and correction |
| 2.5 | 09-Feb-2017 | - Addition of South Africa as a study site<br>- Modification of control regimen duration to include shorter standardized regimen recently approved by the WHO<br>- Clarification of the study outcome measures<br>- Clarification of the study exclusion criteria<br>- Modification of the study prescreening procedures<br>- Clarification of the study visit window, and the schedule of events<br>- Change in study visit procedures at follow up visits, specifically Hepatitis B and C test, Chest X-ray, and Total Calcium<br>- Clarification of serious adverse event classification<br>- Other minor editorial and typographical updates and corrections |

| Version number | Date | Summary of changes (continued) |
|---|---|---|
| 2.6 | 09-May-2017 | - Administrative changes including, PI, Co-I, and study site address<br>- Modification of the study screening procedures<br>- Clarification of blood test<br>- Modification and clarification of the study exclusion criteria<br>- Clarification of the frequency of treatment delivery for the control group<br>- Other minor editorial and typographical updates and corrections |
| 3.0 | 12-Feb-2018 | Note: version not submitted to countries<br>- Eligibility for previous exposure to second-line drugs<br>- Addition of a randomization for linezolid dose-reduction strategy<br>- Possibility to measure CD4/viral load from screening<br>- Possibility to use previous chest X-ray, TSH, CD4/viral load and HbA1c results for baseline<br>- Update of time from pregnancy test date from treatment initiation<br>- Update of test repeat at screening<br>- Week 8 visit window flexibility<br>- Update of adverse event of special interest<br>- Update of safety follow up<br>- Update of secondary objectives and addition of exploratory objectives and related measures<br>- Randomization adaptation based on W8 culture negativity instead of culture conversion<br>- Administrative changes: addition of Pakistan as a study country, update of lead investigators, study site address and site PIs<br>- Other minor editorial and typographical updates and corrections |
| 3.1 | 29-Aug-2018 | Note: version not submitted to countries<br>- Continuation of treatment for patient with initial culture negative<br>- Addition of post-termination follow-up visits for patients withdrawn early<br>- Change in the follow-up duration (minimum W73 and up to W104)<br>- Possibility to measure TSH and HbA1c at screening visit<br>- Possibility to continue study treatment in patients becoming pregnant<br>- Update of drugs tested in DST (fluoroquinolones instead of ofloxacin)<br>- Update of publication policy<br>- Administrative changes: addition of a study site in Peru, change in recipient for SAE notification<br>- Other minor editorial and typographical updates and corrections |
| 3.2 | 23-Oct-2018 | - Reasons for Treatment Discontinuation and Withdrawal from Study: updated for changes due to WHO new guidance<br>- Clarification on bedaquiline loading dose in experimental regimens<br>- Clarification regarding patients becoming pregnant<br>- Revised the description of populations for sensitivity analysis<br>- Clarification regarding Event Validation Group<br>- Minor typographical corrections |
| 3.3 | 14-Feb-2019 | - Addition of amikacin in DST and genomic sequencing among genotyping methods<br>- Administrative update: update of site name in Kazakhstan, addition of India, deletion of Kyrgyzstan<br>- Other minor editorial and typographical updates and corrections |

| | | |
|---|---|---|
| 3.4 | 31-July-2020 | - Administrative changes (addition of Vietnam as a study country, addition of sites in India, Kazakhstan and Pakistan, update of IRBs, lead investigators, site PIs and sites)<br>- Modification of the study duration and of the estimated time to complete enrollment<br>- Change of birth control requirements to align with guidance from Clinical Trials Facilitation Group of European Union Medicines Agencies<br>- Change of exclusion criteria (deletion of albumin and modification of QRS complex duration to > = 120msec)<br>- Added possibility to perform study procedures at participants' homes or through remote visits to reduce risks during the COVID-19 outbreak<br>- Clarification on participation in another trial<br>- Possibility to use linked endTB-Q trial screening consent form and screening results for endTB<br>- Additional information about secondary linezolid dose randomization in randomization section<br>- Update of experimental regimen dosing according to weight<br>- Clarification on allowable screening sputum specimens collection and tests<br>- Addition of SARS-CoV-2 infection and COVID-19 as covariates in analysis<br>- Clarification on informed consent process: any witness must be able to read and write<br>- Addition of Week 104 endpoint to application of assessable population<br>- Modification of exploratory objective time point for linezolid dose reduction strategies (Week 104 instead of Week 73)<br>- Minor editorial and typographical updates and corrections<br>- Extraction of "Statement of Compliance" and "Institutions / Site Principal Investigators and Study Sites" from document body into appendices 2 and 3, respectively |
| 3.5 | 15-Dec-2020 | - Addition of the scientific rationale to drop the albumin exclusion criterion<br>- Update of publication, dissemination and access section |
| 3.6 | 20-Jul-2022 | - Addition of Week 73 as exploratory objective for efficacy endpoint across linezolid dose-reduction strategies<br>- Clarifications regarding definitions and handling of treatment discontinuation and withdrawal from study, including post-termination study visits<br>- Addition of optional MIC testing<br>- Text regarding levofloxacin posology adapted to drug dosing table for the 24-30 kg weight band<br>- Exclusion of participants with DST resistance result for bedaquiline, clofazimine, delamanid and/or linezolid from the mITT population<br>- Clarification that primary efficacy analyses will be conducted in both mITT and PP populations<br>- Sensitivity analyses: moved mention from analysis population to primary endpoint analyses section; removed incomplete description; and added referral to statistical analysis plan for full detail of sensitivity analyses<br>- Administrative update: deletion of Vietnam as a study country, update of IRBs, lead investigators, site PIs and sites<br>- One reference updated |

5.3.1 Randomization Procedures ... 53
5.3.2 Masking Procedures ... 54
5.3.3 Reasons for Treatment Discontinuation and Withdrawal from Study ... 55
5.3.4 Handling of Withdrawals and Discontinuations ... 56
5.3.5 Loss to Follow-up ... 56
5.3.6 Termination of Study ... 56
6 Study Treatment of Patients ... 57
6.1 Investigational Drugs ... 57
6.1.1 Bedaquiline (Be) ... 57
6.1.2 Delamanid (De) ... 57
6.1.3 Clofazimine (C) ... 57
6.1.4 Fluoroquinolones ... 58
6.1.5 Linezolid (Li) ... 58
6.1.6 Pyrazinamide (Z) ... 59
6.2 Trial Interventions ... 59
6.2.1 Experimental Treatment ... 59
6.2.2 Control Treatment ... 60
6.2.3 Modification of Study Intervention/Investigational Drug for a Participant ... 61
6.3 Medicines Supplies ... 61
6.4 Accountability Procedures for the Study Intervention/Investigational Drug(s) ... 62
6.5 Assessment of Subject Compliance with Study Intervention/ Investigational Drug ... 62
6.6 Concomitant Medications/Treatments ... 62
7 Study Schedule ... 64
7.1 Screening ... 64
7.1.1 Pre-screening ... 64
7.1.2 Screening ... 65
7.2 Baseline ... 66
7.3 Follow-up ... 67
7.4 Early Termination Visit ... 69
7.5 Post-termination follow-up visit(s) ... 69
8 Study Procedures/Evaluations ... 71
8.1 Clinical Evaluations ... 71
8.2 Laboratory Evaluations ... 72
8.2.1 Blood and Urine Tests ... 72
8.2.2 Tuberculosis Tests ... 73

11.4.1 Analysis Populations ... 90
11.4.2 Primary Endpoint Analyses ... 91
11.4.3 Data Handling Convention ... 92
12 Source Documents and Access to Source Data/Documents ... 93
13 Quality Control and Quality Assurance ... 94
14 Ethics/Protection of Human Subjects ... 95
14.1 Ethical Standard ... 95
14.2 Institutional Review Board ... 95
14.3 Informed Consent Process ... 95
14.3.1 Informed Consent/Assent Process (in Case of a Minor) ... 96
14.3.2 Consent for follow up of pregnancy in female partners of male participants ... 96
14.3.3 Special Populations ... 96
14.3.4 Subject Confidentiality ... 97
14.4 Study Discontinuation ... 97
14.5 Future Use of Stored Specimens ... 98
15 Data Handling and Record Keeping ... 99
15.1 Data Management Responsibilities ... 99
15.2 Data Capture Methods ... 99
15.3 Types of Data ... 99
15.4 Timing/Reports ... 100
15.5 Study Records Retention ... 100
15.6 Protocol Deviations ... 100
16 Trial Boards and Committees ... 101
16.1 Scientific Advisory Committee (SAC) ... 101
16.2 Global Tuberculosis Community Advisory Board (TB-CAB) ... 101
16.3 Data and Safety Monitoring Board (DSMB) ... 101
16.4 Clinical Advisory Committee (CAC) ... 102
16.5 Event Validation Group (EVG) ... 102
17 Publication, dissemination and access ... 103
18 REFERENCES ... 104

### LIST OF ABBREVIATIONS

| | |
|---|---|
| AE | Adverse Event |
| AESI | Adverse Event of Special Interest |
| ALT | Alanine Aminotransferase |
| ART | Anti-retroviral Therapy |
| AST | Aspartate Aminotransferase |
| ATP | Adenosine Triphosphate |
| Be, J, TMC, TMC207 | Bedaquiline |
| ßHCG | Beta Human Chorionic Gonadotropin |
| BID | Two times a day |
| BMI | Body Mass Index |
| C, CFZ | Clofazimine |
| CA++ | Calcium |
| CAC | Clinical Advisory Committee |
| CBC | Complete Blood Count |
| CD4 | CD4 Cell Count |
| CFR | Code of Federal Regulations |
| CFU | Colony Forming Unit |
| CI | Confidence interval |
| CRF | Case Report Form |
| CTCAE | Common Terminology Criteria for Adverse Events |
| CYP | Cytochrome P450 |
| De | Delamanid |
| DMID | Department of Microbiology and Infectious Diseases, National Institutes of Allergy and Infectious Diseases, National Institutes of Health |

| | |
|---|---|
| DNA | Deoxyribonucleic Acid |
| DR | Drug-Resistant |
| DS | Drug-Susceptible |
| DSMB | Data Safety and Monitoring Board |
| DST | Drug Susceptibility Test |
| DTP | Days to Positivity |
| EBA | Early Bactericidal Activity |
| ECG | Electrocardiogram |
| ECOG | Eastern Cooperative Oncology Group |
| eCRF | Electronic case report form |
| EMA | European Medicines Agency |
| EMB | Ethambutol |
| endTB | Study Name |
| EVG | Event Validation Group |
| ERB | Ethics Review Board |
| FDA | Food and Drug Administration |
| FQ | Fluoroquinolones |
| FQ-R | Fluoroquinolone-resistant |
| FQ-S | Fluoroquinolone-susceptible |
| GCP | Good Clinical Practice |
| H, INH | Isoniazid |
| HbA1c | Hemoglobin A1c |
| HBc | Hepatitis B Core |
| HBsAg | Hepatitis B Surface Antigen |
| HBV | Hepatitis B virus |

| | |
|---|---|
| HCV | Hepatitis C virus |
| HCVAb | Hepatitis C virus antibody |
| HIV | Human Immunodeficiency Virus |
| HIV+ | HIV seropositive |
| ICF | Informed Consent Form |
| ICH | International Council on Harmonization |
| IRB | Institutional Review Board |
| IRD | Interactive Research and Development |
| IVRS/IWRS | Interactive Voice Response System/ Interactive Web Response System |
| K+ | Potassium |
| KM | Kanamycin |
| Le, Lfx | Levofloxacin |
| Li, LZD | Linezolid |
| LJ | Löwenstein Jensen |
| LTFU | Loss to Follow Up |
| MAMS | Multi-Arm Multi-Stage |
| MDR-TB | Multidrug-resistant Tuberculosis |
| MedDRA | Medical Dictionary for Regulatory Activities |
| MG++ | Magnesium |
| MGIT | Mycobacterial Growth Indicator Tube |
| MIC | Minimal Inhibitory Concentration |
| mITT | Modified Intent-to-Treat |
| Mo, M, MXF | Moxifloxacin |
| MSF | Médecins Sans Frontières |
| N | Number (typically refers to subjects) |

| | |
|---|---|
| NRA | National Regulatory Authority |
| P, RPT | Rifapentine |
| Pa, PA-824 | Pretomanid |
| PAS | Para-aminosalicylic Acid |
| PI | Principal Investigator |
| PIH | Partners In Health |
| PP | Per Protocol |
| PR | Pulse Rate |
| PTO | Prothionamide |
| PT | Preferred Term |
| PV | Pharmacovigilance |
| QA | Quality Assurance |
| QC | Quality Control |
| QD | Once a day |
| QTcB | QT Interval Heart-Rate-Corrected by Bazett Formula |
| QTcF | QT Interval Heart-Rate-Corrected by Fridericia Formula |
| RIF, R | Rifampin, Rifampicin |
| RSI | Reference Safety Information |
| SAC | Scientific Advisory Committee |
| SAE | Serious Adverse Event |
| SAP | Statistical Analyses Plan |
| SD | Standard Deviation |
| SOC | System-Organ Class |
| SOP | Standard Operating Procedure |
| SUSAR | Suspected Unexpected Serious Adverse Reaction |

| | |
|---|---|
| TB | Tuberculosis |
| TSH | Thyroid-stimulating Hormone |
| TTP | Time to (culture) positivity |
| ULN | Upper Limit of Normal |
| WHO | World Health Organization |
| XDR-TB | Extensively Drug-resistant Tuberculosis |
| Z, PZA | Pyrazinamide |

### PROTOCOL SUMMARY

| | |
|---|---|
| **Title:** | endTB: Evaluating Newly approved Drugs for multidrug-resistant TB |
| **Phase:** | III |
| **Population:** | Adults and adolescents (≥15 years old) with pulmonary TB with *Mycobacterium tuberculosis (M. tuberculosis)* resistant to RIF and without resistance to fluoroquinolones |
| **Number of Countries:** | 7 (Georgia, India, Kazakhstan, Lesotho, Pakistan, Peru, South Africa) |
| **Sample Size:** | 750 subjects |
| **Study Duration:** | 5.5 years |
| **Participation Duration:** | Between 73 weeks and 104 weeks (17 and 24 months)<br><br>Study follow-up will end after the scheduled Week 73 for the last participant randomized (to be defined by the Sponsor). |
| **Description of Intervention:** | Multidrug treatment regimens for fluoroquinolone-susceptible, rifampin-resistant tuberculosis. Experimental regimens will contain bedaquiline and/or delamanid and up to 4 companion drugs and will be delivered for 39 weeks. Control-arm treatment may contain one of the following (bedaquiline or delamanid) and companion drugs, constructed and delivered according to local standard of care and consistent with WHO guidelines for the conventional or shorter regimen. |

| | |
|---|---|
| **Objectives:**<br>**Primary:**<br><br><br>**Secondary:** | To assess whether the efficacy of experimental regimens at Week 73 is non-inferior to that of the control.<br><br>1. To compare the efficacy of experimental regimens at Week 104 to that of the control.<br>2. To compare the frequency of and time to early treatment response (culture conversion) in experimental regimens to that of the control.<br>3. To compare the efficacy of experimental regimens at Week 39 to that of the control.<br>4. To compare, at Week 73 and Week 104, the proportion of patients who experienced failure or relapse in the experimental arms to that in the control arm.<br>5. To compare, at Week 73 and Week 104, the proportion of patients who died of any cause in the experimental arms to that in the control arm<br>6. To compare, at Week 73 and Week 104, the proportion of patients who experience grade 3 or higher Adverse Events (AEs) or Serious AEs (SAEs) of any grade in the experimental arms to that in the control arm.<br>7. To compare, at Week 73, the proportion of patients who experience Adverse Event of Special Interest (AESI) in experimental regimens to that in the control arm. |
| **Description of Study Design:** | This is a randomized, controlled, open-label, multi-country Phase III trial evaluating the efficacy of new combination regimens for treatment of MDR-TB. The study will enroll in parallel across 5 experimental and 1 standard-of-care control arms through adaptive randomization, which will be based on efficacy endpoints. In the experimental arms, treatment will be for 39 weeks. In the control arm, treatment will be delivered according to local practice (and WHO guidelines); duration may vary and will be approximately 86 weeks for the conventional regimen and 39-52 weeks for the standardized shorter regimen.<br>Trial participation in all arms will last at least until Week 73, and up to Week 104. |
| **Estimated Time to Complete Enrollment:** | 4 years |

### 1 KEY ROLES

**Individuals**[1]**:**

**Co-Principal Investigators:**

**Carole Mitnick, Sc.D.**
Department of Global Health & Social Medicine
Harvard Medical School
641 Huntington Ave.
Boston, MA 02115, USA
(617) 432-6018


**Lorenzo Guglielmetti, M.D.**
Sorbonne Université, UPMC Univ Paris 06, CR7, INSERM, U1135, Centre d'Immunologie et des Maladies Infectieuses,
91 Boulevard de l'hôpital, 75634 Paris Cedex 13, France
Medical Department
Médecins Sans Frontières
14/34 Avenue Jean-Jaurès,
75019 Paris, France


**Other Lead Investigators:**

**Elisabeth Baudin, M.Sc.**
Research Department
Epicentre
14/34 Avenue Jean-Jaurès,
75019 Paris, France


**Maryline Bonnet, M.D., Ph.D.**
Institut de Recherche pour le Développement (IRD)
UMI 233 TransVIHMI
UM – INSERM U1175
Epicentre
911 avenue Agropolis, BP 64501
34394 Montpellier cedex 5, France


**Michael Rich, M.D.**
Brigham and Women's Hospital & Partners In Health
641 Huntington Ave.
Boston, MA 02115, USA


**Bouke de Jong, M.D., Ph.D.**
Unit of Mycobacteriology
Institute of Tropical Medicine
Nationalestraat 155
2000 Antwerp, Belgium


**Francis Varaine, M.D.**
Medical Department
Médecins Sans Frontières
14/34 Avenue Jean-Jaurès,
75019 Paris, France


**Gabriella Ferlazzo, M.D.**
TB/HIV consultant
137 Rue de Hollerich

L-1741 Luxembourg


**Kwonjune Seung, M.D.**
Associate Physician
Division of Global Health Equity
Brigham and Women's Hospital
75 Francis St.
Boston, MA 02115, USA


**Payam Nahid, M.D., M.P.H.**
University of California, San Francisco
Division of Pulmonary & Critical Care Medicine
Zuckerberg San Francisco General Hospital
1001 Potrero Avenue, Room 5K1
San Francisco, CA 94110


**Gustavo Velásquez, M.D., M.P.H.**
Assistant Professor of Medicine
Division of HIV, Infectious Diseases, and Global Medicine
San Francisco General Hospital
University of California, San Francisco
San Francisco, CA


**Uzma Khan, M.D., M.S.**
Interactive Research and Development
WH # 13, Building 2,
Al Quoz 1, P.O. Box 51091,
Dubai, UAE


**Other Collaborators:**

**Central Data Center:**
Epicentre
14/34 Avenue Jean-Jaurès,
75019 Paris, France

**Central Laboratory:**
Unit of Mycobacteriology
Institute of Tropical Medicine
Nationalestraat 155
2000 Antwerp, Belgium

### 2 BACKGROUND INFORMATION AND SCIENTIFIC RATIONALE

#### 2.1 Background Information

Multidrug resistant tuberculosis (MDR-TB) is tuberculosis caused by *M. tuberculosis* strains that are resistant to the most potent anti-tuberculosis drugs in standard treatment, isoniazid and rifampin. MDR-TB is a global problem. A new shortened regimen offers hope to the subset of MDR-TB patients whose isolates are susceptible to fluoroquinolones, second-line injectables, and all other drugs in the regimen and who have not had prior exposure to second-line drugs.[2] The lack of a safe and effective MDR regimen that can be used in all patients is a major obstacle to delivering appropriate treatment to all patients with active MDR-TB disease. Of the 480,000 estimated, new MDR-TB patients in 2014, only 123,000 (26%) were actually diagnosed with the disease. Fewer, 111,000 (23%), ever received any treatment with second-line TB drugs.[3] Half or fewer are cured globally.[3] The consequences of this situation are dire: in at least one setting, 80% percent of patients in whom MDR-TB treatment failed died within three years.[4] And, each untreated or inadequately treated MDR-TB patient can infect 3-6 susceptible individuals each year. Those who do receive "appropriate" conventional treatment are exposed to 18-24 months of toxic, poorly tolerated treatment regimens[5] that result in frequent default.[6,7,8,9]

Toxicity occurs nearly universally in patients receiving standard-of-care treatment for MDR-TB. Common adverse events include: ototoxicity, liver and renal toxicity, gastrointestinal disorders, electrolyte imbalance, hypothyroidism, and neurotoxicity. Pivotal studies of two drugs, bedaquiline and delamanid, newly approved by stringent regulatory authorities for MDR-TB treatment provide the first opportunity for systematic, prospective reporting of toxicity to a regulatory standard. In the placebo arm of the bedaquiline study, 98% of participants experienced adverse events; 36% had a severe adverse event and 6% had to discontinue treatment.[10] In the placebo arm of the pivotal trial of delamanid, 94% of participants experienced adverse events after only 8 weeks of treatment, 9% of them severe.[11] Observational cohorts corroborate the high frequency of side effects, with estimates ranging from 73% to 79%. Specifically, 58-75% experienced nausea and vomiting, 9-17% hepatotoxicity, 16-19% hearing loss, 12-13% acute psychiatric disorders, 3-33% hypokalemia, 4-10% renal failure, and 4-8% neuropathy.[12,13] These numbers are likely underestimates as they are derived from retrospective studies, in which systematic, standard real-time recording and reporting of adverse events was not the norm. Some adverse events, like hypothyroidism, are only investigated in some settings. When thyroid stimulating hormone is monitored, hypothyroidism has been shown to affect 34% to 69% of patients.[14,15] Some of these adverse events, like acute liver and renal failure, psychosis and electrolyte imbalance (in particular hypokalemia), can be particularly severe and can cause (sudden) death.[16,17,18,19,20] For instance, in a cohort of 115 South African patients treated for drug-resistant tuberculosis, 6 died as a consequence of capreomycin-related adverse events: 5 cases of renal insufficiency and one severe hypokalemia.[21] Fluoroquinolones, a cornerstone of MDR-TB treatment, are known to prolong QT interval,[22] and less frequently to cause ventricular arrhythmias/dysrhythmias and cardiac arrest.[23] Moxifloxacin, favored for its increased activity,[24,25] has a greater impact on QT interval than levofloxacin, both after a single dose and after a short-term treatment course.[26,27]

This constellation of adverse events complicates management of MDR-TB treatment and frequently leads to suspension and replacement of the suspected drug(s) in conventional regimens.[28,29] In addition, treatment-related adverse events have been shown to be one of the major drivers of treatment default,[30] as well as to be associated with a lower rate of culture conversion.[21]

Returning to treatment efficacy, important subgroups could benefit even more than the general population affected by the disease from the availability of new, more effective treatment regimens. MDR-TB patients coinfected with HIV have abysmal treatment outcomes: in a large cohort of HIV-infected MDR-TB patients from Eastern Europe, only 22% experienced a successful treatment outcome, while 65% died, most of them during the first year after TB diagnosis.[31] In HIV-infected MDR-TB patients in South Africa, success was achieved in 48%; 19% died.[32] HIV infection has been shown to be a strong independent predictor of mortality in MDR-TB patients.[33] And the pill burden in co-infected patients is excessive: one such patient in Mozambique took 17 tablets, 3 capsules, 2 sachets and 1 intramuscular injection daily, for 8 months, followed by 17 tablets, 3 capsules, and 2 sachets for an additional year.

Another group with notably poor MDR-TB outcomes is adolescents. Although limited data are available, they signal a need for significant improvement. In a small cohort of adolescents with DR-TB from Khayelitsha, South Africa, 63.6% either died, failed therapy or were lost to follow up. In a smaller observational cohort of 11 adolescent patients from Mumbai, India with MDR-TB and HIV coinfection, more than 50% had poor outcomes.[34,35] One possible explanation is difficulty with compliance linked to the developmental characteristics of adolescence;[36] shorter, simpler regimens would likely represent an important improvement.

This desperate situation was due, in part, to a 50-year drought in anti-TB drug development. Finally, in 2013 and 2014, two new anti-TB drugs, bedaquiline and delamanid, were approved by stringent regulatory authorities for the treatment of MDR-TB. The pivotal trials of these drugs simply added each to a background regimen, improving interim and long-term outcomes, but retaining the toxic, long, conventional regimen.[10,11,37,38] Without incorporating these newly approved drugs into novel regimens, their impact on the burden of disease will be extremely limited. Only a few hundred patients have been treated with these drugs since their approval. The present effort, endTB aims to evaluate integration of these drugs in novel, 9-month, all oral, MDR-TB treatments.

Although there is risk inherent in testing new combinations, endTB's novel regimens have the potential to improve the current situation significantly. They could reduce treatment duration by more than 50%; eliminate injections and lighten the pill burden; and potentially increase efficacy without worsening toxicity. A shorter regimen with different therapeutic agents may be better tolerated, require less attention from health-care providers, and allow more patients to complete therapy, possibly with little increase in relapse. In addition, the implementation of injectable-sparing regimens would likely improve patient compliance and simplify the logistical burden on national tuberculosis programs, making treatment accessible to more than the 23% of patients who initiated treatment in 2014.[3]

These needs—poor outcomes and high toxicity of as well as limited access to MDR-TB treatment—drive the urgency of conducting studies to inform optimized individual and concomitant use of bedaquiline and delamanid. In particular, concomitant use has not yet been studied. It has, however, been advocated in individual cases and it would be an understandable option for clinicians to explore in light of the drawbacks to the conventional regimen.[39] It is urgent, therefore, to provide evidence to guide such decision-making so clinicians do not have to make clinical decisions blindly or with only expert opinion to guide them. This has been the state of MDR-TB treatment for decades. MDR-TB clinicians have had to use drugs with, at best, equivocal evidence of safety and efficacy (e.g., macrolides,[40] thioridazine[41]). MDR-TB regimens used to date have relied heavily on drugs such as aminoglycosides (ototoxicity, electrolyte imbalance, renal toxicity), cycloserine (psychiatric and central nervous toxicity), ethionamide (thyroid and GI toxicity) that may be toxic alone[42,43,44] or in combination.[45] These are rational decisions in light of the important risks associated with MDR-TB, yet they rely on expert opinion. Few of the drugs that are used off-label for MDR-TB have been optimized for the treatment of MDR-TB. In rare examples (fluoroquinolones and linezolid) any optimization is occurring after years of off-label use. For both, optimal dosing is only now being explored (ClinicalTrials.gov NCT02279875 and NCT01918397), despite reports of their use for MDR-TB for at least 10 years.[46,47] For these reasons, it is critical that carefully conducted studies generate evidence about safe and effective combinations of bedaquiline and delamanid, including appropriate companion drugs. Beginning to accumulate this evidence within endTB will reduce the inevitable gap from the time both drugs become available in programs to the moment that clinicians can make evidence-based decisions about incorporating these drugs in treatment regimens. Since both delamanid and bedaquiline will be available through the Global Drug Facility in 2016, the need is urgent for evidence about their joint use to inform clinical decision-making.

#### 2.2 Rationale

The goal of more effective, safer, and more accessible MDR-TB treatment globally can best be met with multiple treatment alternatives. This is because, among the ½ million new cases each year, there is significant heterogeneity in resistance patterns. In addition, there are comorbidities requiring concomitant treatments that may interact with drugs in a given regimen, and characteristics that determine toxicity and intolerability, as well as regional or temporal variability in drug availability.

Traditionally, new multidrug regimens are developed sequentially: researchers substitute a single new drug for an existing one or add a new drug to an existing regimen. Decades may pass before a new regimen is developed using this model.[48] endTB, along with a number of other regimen-development efforts, aims to shorten the time it takes to develop novel TB regimens, by simultaneously changing more than one drug in multidrug regimens. endTB aims to identify multiple new, effective regimens as rapidly as possible. endTB will examine five experimental regimens containing one or two new drugs in patients with fluoroquinolone-susceptible MDR-TB.

##### 2.2.1 Principles for Regimen Design in endTB

endTB aims to build on the studies that led to the approval of bedaquiline[10] and delamanid[11] to optimize the use of these two drugs along with other drugs used to treat MDR-TB. Specifically, endTB is testing 5 regimens that meet most or all of the following criteria:[49]

- Contain at least one new class of drug;
- Include regimens for MDR- and XDR-TB[i] strains;
- Contain three to five effective drugs, each from a different drug class;
- Be delivered orally;
- Have a simple dosing schedule;
- Have a good side-effect profile that allows limited monitoring;
- Last a maximum of 6 to 9 months;
- Have minimal interaction with antiretroviral drugs.

##### 2.2.2 Rationale for Choice of Drug Combinations

It is well established that optimal TB regimens are multidrug, containing agents that work against the different mycobacterial populations—actively replicating and dormant, intracellular and extracellular—present in active disease. Importantly, to optimize activity and minimize toxicity, the infecting isolate should be susceptible to (nearly) all of the drugs in a prescribed regimen. This reduces the total number of drugs, limiting the pill burden and avoiding toxicity. For this reason, endTB regimens will rely primarily on drugs to which there is limited prior population exposure: delamanid, bedaquiline, clofazimine, and linezolid. Two drug classes, to which prior exposure is more common will also be used because of their perceived importance to MDR-TB regimens. These are pyrazinamide and the late-generation fluoroquinolones, levofloxacin and moxifloxacin. One other potentially significant issue guiding regimen design in endTB is toxicity, specifically the QT-interval prolonging liability of several of the drugs: bedaquiline, clofazimine, moxifloxacin, and, to a lesser degree, delamanid and levofloxacin, have all been shown to prolong the QT interval. Although no increased frequency of clinical cardiac events has been documented with their use, either alone or in combination, endTB will exercise caution. The principles outlined above result in the following regimen selection strategy:

1. All regimens contain either bedaquiline or delamanid; one contains both;
2. All regimens contain pyrazinamide and a fluoroquinolone;
3. No regimen contains more than 5 or fewer than 4 drugs; at least 3 are considered likely effective;
4. No regimen contains more than 2 of the important QT-interval-prolonging drugs: clofazimine, bedaquiline, and moxifloxacin;
5. No regimen contains an injectable agent.

The five regimens selected are listed in Table 1. Pre-clinical and clinical evidence that supports the use of these combinations in shortened regimens is presented below, first drug by drug, then the combinations.

[i] Extensively drug-resistant TB, TB caused by MDR strains resistant also to at least one fluoroquinolone and second-line injectables

**Table 1 endTB Regimens**

| Trial Regimens | Bedaquiline | Delamanid | Clofazimine | Linezolid | Fluoroquinolone | Pyrazinamide |
|---|---|---|---|---|---|---|
| **endTB 1 BeLiMoZ** | Be | | | Li | Mo | Z |
| **endTB 2 BeCLiLeZ** | Be | | C | Li | Le | Z |
| **endTB 3 BeDeLiLeZ** | Be | De | | Li | Le | Z |
| **endTB 4 DeCLiLeZ** | | De | C | Li | Le | Z |
| **endTB 5 DeCMoZ** | | De | C | | Mo | Z |
| **endTB 6 Control** | Standard of care control, composed according to WHO Guidelines, including the possible use of De or Be. | | | | | |

Fluoroquinolones: Mo: moxifloxacin; Le: Levofloxacin.

Delamanid

Delamanid (OPC-67683 or Deltyba™) is a new agent derived from the nitrodihydro-imidazooxazole class of compounds. It inhibits the synthesis of mycolic acid, which is an exclusive component of the external wall of mycobacteria. Peak plasma levels are achieved 4-5 hours after administration. The half-life of delamanid is 38 hours after drug discontinuation. It is indicated for use against MDR-TB (EMA).

Delamanid has demonstrated *in vitro* and *in vivo* activity against both drug susceptible and drug resistant strains of *M. tuberculosis*.[50] Among nitroimidazoles, delamanid is thought to be more active and less prone to selection for resistant mutants than pretomanid and similar to TBA-354, two other nitroimidazoles under clinical development for TB.[51,52,53] In addition, delamanid showed activity in a chronic mouse model of TB, suggestive of sterilizing activity.[50] Although no published report documents delamanid's sterilizing activity in preventing relapse in mice, this characteristic is likely based on *in vitro* data and murine data on other nitroimidazoles in mouse models.[52]

Early bactericidal activity (EBA) of delamanid was demonstrated at all tested doses, 100-400 mg/day for 14 days.[54] Further clinical evidence for the efficacy of delamanid in MDR-TB treatment derives mainly from a Phase II trial in which 481 MDR-TB patients were randomized across nine countries to receive 100 mg delamanid, 200 mg delamanid, or placebo twice daily in combination with an optimized background regimen. Patients received the study treatment for 2 months. Higher sputum-culture conversion rates were observed in both the 100 mg delamanid group (45.5%) and 200 mg delamanid group (41.9%), compared to the placebo group (29.6%).[11] Longer-term results, which include roll-over into an open-label study, corroborate these findings. Nearly 90% of the 481 Phase II participants were included in the extension trial. Of these, 74.5% of participants who received delamanid for at least 6 months had favorable outcomes compared to only 55% of participants who received delamanid for 2 months or less. A strong mortality benefit was also observed.[38] Unpublished Phase III trial data show a modest improvement in culture conversion at 6 months. The median difference (6 days) was not statistically significant in the primary analysis (p=0.0562). The difference was greater (13 days), and statistically significant in two sensitivity analyses (p=0.028, 0.005). The study was not powered for long-term outcomes and these were very similar in both the experimental and control arms, surprisingly over 80% in both. A recently published case series of 19 children and adolescents with MDR-TB treated with regimens including delamanid given under compassionate use showed promising efficacy and tolerability results. Complementing likely sterilizing activity, delamanid appears to act early, when bacterial populations are large and actively replicating, resulting in demonstrated EBA as well as reduced time to culture conversion.

Bedaquiline

Bedaquiline fumarate (bedaquiline or Sirturo™) is a diarylquinoline anti-mycobacterial drug. It inhibits ATP synthesis, a novel method of action. The drug has a 5.5-month half-life. It is indicated for use against MDR-TB (US FDA, EMA).

*In vitro* activity of bedaquiline was demonstrated against *M. tuberculosis*, with a MIC <0.06 mcg/ml. It was found to be active both against replicating and dormant bacteria in *in vitro* studies.[55] Mouse studies revealed both early and sterilizing activity of bedaquiline in regimens containing first-line drug[56] and in novel combinations.[57]

The clinical efficacy of bedaquiline was demonstrated in two Phase II trials.[10,58,59] In the second, 160 patients were randomized to receive bedaquiline 400 mg, or placebo, daily for 2 weeks, followed by 200 mg bedaquiline, or placebo, 3 times per week for an additional 22 weeks.[10] Patients also received a background regimen containing various combinations of fluoroquinolones, aminoglycosides, pyrazinamide, ethionamide, ethambutol, and cycloserine/terizidone. Bedaquiline reduced the median time to culture conversion from 125 days (placebo group) to 83 days and increased the culture conversion rate at 24 weeks from 58% (placebo group) to 79%.[10] Pre-clinical and clinical data on bedaquiline support its contribution to bactericidal and sterilizing activity in a range of multidrug regimens.

Clofazimine

Clofazimine (Lamprene®) is a substituted riminophenazine derivative. It acts against mycobacteria by inhibiting replication and growth. It binds preferentially to mycobacterial DNA. Clofazimine accumulates in tissue; the half-life of clofazimine following repeated doses is estimated to be at least 70 days. It is indicated for use in leprosy.

In addition to its slow bactericidal effect, clofazimine has sterilizing activity against *M. tuberculosis*, demonstrated in mouse studies of first- and second-line multidrug regimens.[60,61]

Exciting results from a 9-month clofazimine-containing regimen first used in Bangladesh[62,63] renewed interest in clofazimine for the treatment of MDR-TB. Systematic reviews and meta-analyses of clofazimine for MDR-TB have reported pooled success rates similar to those generally reported for MDR-TB treatment: a systematic review of nine observational studies comprising 599 MDR-TB patients treated with clofazimine showed an overall success rate of 65% (95% CI:54%-76%).[64] Another review of twelve studies, comprising 3,489 patients across 10 countries showed an overall pooled success rate of 61.96% (95% CI: 52.79%-71.12%).[65] This is despite the fact that clofazimine is often reserved for patients with few treatment options who are at risk for worse than average treatment outcomes.

Equivocal evidence emerged from a recent EBA study. Clofazimine had no measurable activity alone or when added to a combination with bedaquiline, pretomanid and/or pyrazinamide at 14 days in 15 treatment-naïve, sputum smear-positive patients with pulmonary TB. A regimen containing pretomanid, pyrazinamide, and clofazimine, however, had activity that was neither distinguishable from the most active experimental regimen nor from the isoniazid-rifampin-ethambutol-pyrazinamide control.[66] Clofazimine is thought to require longer exposure to achieve maximum efficacy. At least one mouse pharmacokinetic/pharmacodynamics study shows dose-independent increases in plasma and tissue concentrations over the entire period of administration, 20 weeks.[67] An unpublished human simulation study also corroborates this. This study revealed that plasma concentrations of clofazimine dosed at 200 and 300 mg QD did not exceed the MIC99 until at least 3 weeks after initiation. They continued to rise as long as administered daily.[68] EBA studies might not, therefore, effectively capture the activity of clofazimine in humans. A randomized, controlled trial of 105 MDR-TB patients compared outcomes among participants receiving clofazimine 100 mg/day and participants not receiving clofazimine, as part of a 21-month multidrug regimen. Treatment success was reported in 76% of the clofazimine group and 53.8% in the control group (p=0.035). Furthermore, interim favorable endpoints assessed by the study (cavity closure and accelerated sputum culture conversion) occurred more frequently in the clofazimine arm.[69] Apparent benefits accruing to clofazimine in multidrug treatment may have more to do with sterilizing than with bactericidal activity.

<u>Linezolid</u>

Linezolid is a synthetic agent and a member of the oxazolidinone class of antibiotics.[70] Linezolid works by inhibiting ribosomal protein synthesis. It is rapidly absorbed (peak serum concentration reached 1-2 hours after administration); its average half-life is 5 hours in adults. It is licensed for gram-positive infections and has demonstrated activity against laboratory strains, as well as against clinical strains of pan-susceptible and resistant *M. tuberculosis*.[71,72,73] A series of mouse studies illustrated the bactericidal and sterilizing activity of linezolid (and other oxazolidinones) in multidrug treatment; the addition of linezolid to novel regimens, for as little as one month, reduced the frequency of relapse in mice after 3 months of treatment from 21 to 0%.[74]

There is a growing evidence from clinical studies that linezolid is effective against *M. tuberculosis*. MDR-TB case series have reported relatively high rates of treatment success using linezolid. A systematic review of 11 studies reporting outcomes of 148 MDR-TB patients treated with linezolid showed a pooled proportion of treatment success of 67.99% (95% CI: 58.0%-78.99%).[42] Another meta-analysis conducted on 12 studies reporting treatment outcome (cure, treatment completion, death, treatment failure) for 121 patients showed that 93.5% (100/107) MDR-TB cases achieved sputum culture conversion after treatment with individualized regimens containing linezolid and 81.8% (99/121) were successfully treated.[75] No significant difference in success was observed in either study comparing daily dose of linezolid (≤ 600 mg to > 600 mg).

In a clinical trial evaluating the addition of linezolid (600 mg QD) to a failing XDR-TB regimen, patients were randomly assigned to immediate or delayed (after 2 months) linezolid therapy, without other changes to their regimen. By 4 months, 79% (15/19) of patients in the immediate-start group, compared with 35% (7/20) of patients in the delayed-start group, had culture conversion (p=0.001). Overall, 87% of patients (34/39) had a negative sputum culture within 6 months after linezolid had been added to their drug regimen. It is worth noting that, despite the fact that linezolid was added as a single drug to failing regimens, failure was relatively infrequent (11% [4/38]). And, among the 30 patients who completed treatment, no relapse was detected up to 12 months post-treatment.[76]

In another clinical trial, 65 patients receiving multidrug treatment for XDR-TB were randomly assigned to linezolid or a control. The treatment success rate in the linezolid group was 69%, significantly higher than that in the control group (34.4%, p=0.004).[77] Linezolid, like bedaquiline and delamanid, may contribute both bactericidal and sterilizing activity, important in the context of shortened treatment for MDR-TB.

Fluoroquinolones

Fluoroquinolones, synthetic broad-spectrum antibacterial agents without a TB indication, inhibit DNA gyrase. Late-generation fluoroquinolones, moxifloxacin and levofloxacin, have established anti-TB activity.[78,79] Levofloxacin, the levo isomer of ofloxacin, is rapidly and, essentially, completely absorbed after oral administration. Peak plasma levels are achieved 3 hours after administration. The half-life of levofloxacin ranges from approximately 6 to 8 hours. Moxifloxacin is an 8-methoxyquinolone with a half-life of 11.5-15.6 hours that allows once-daily administration. The drug is rapidly absorbed and achieves high levels in tissues, including the lung.

A series of mouse studies revealed that moxifloxacin, in combination with first-line drugs, both shortened time to culture conversion and reduced risk of relapse, even with reduced treatment duration.[80,81] These results were extrapolated to confer a sterilizing benefit to moxifloxacin, capable of reducing standard treatment in humans by 2 months. Phase III clinical trials did not support such a benefit[82,83,84] and independent re-analysis of the pre-clinical and Phase II data suggest that moxifloxacin-containing first-line regimens could have been successfully shortened by 1 month instead of 2.[85] So, although the experience with fluoroquinolones for treatment shortening cautions about over-reliance on mouse and early-stage clinical trials,[86] it is reasonable to consider the late-generation fluoroquinolones as contributing to the activity and shortening potential of multidrug regimens.

The fluoroquinolones, especially the later-generation class members, are a cornerstone of treatment for MDR- and XDR-TB. In vitro, animal, and human studies all support the inclusion of this drug class for the treatment of resistant TB.[24,87] Later-generation fluoroquinolones have increased activity compared to early-generation and there is evidence that cross-resistance within the class is not complete.[88] Some works suggest that moxifloxacin is more active than levofloxacin against *M. tuberculosis,* though this is not conclusive.[87,89]

Pyrazinamide

Pyrazinamide is a unique anti-TB agent, whose mechanism of action remains unknown. It is a prodrug; once converted to pyrazinoic acid, it is active in acidic micro-environments and works against bacteria with low metabolic activity. Pyrazinamide is well absorbed from the gastrointestinal tract and attains peak plasma concentrations within 2 hours. Widely distributed in body tissues and fluids, pyrazinamide has a plasma half-life of 9 to 10 hours. Pyrazinamide is licensed for TB.

Pyrazinamide has potent sterilizing activity, demonstrated in the innovative Cornell mouse model in the 1950s.[90] Used during the initial phase of treatment, pyrazinamide, coupled with rifampin, was key to shortening 18-month anti-TB treatment.[91] British Medical Research Council studies demonstrated the dramatic sterilizing effects of pyrazinamide: relapse declined from 32% to 10% and two-month culture conversion increased by at least 10% when pyrazinamide was added to regimens containing streptomycin and isoniazid.[92]

Pyrazinamide is well established as a good companion drug for patients whose isolates are susceptible to this agent. In a recent, large meta-analysis of MDR-TB treatment, inclusion of pyrazinamide in the regimen was associated with improved outcomes.[8] Additional analyses of observational cohorts extend these findings, revealing the importance of pyrazinamide, especially when susceptibility is confirmed. In one study, pyrazinamide used in MDR-TB and XDR-TB patients whose isolates tested susceptible to pyrazinamide was associated with a nearly two-fold increase in probability of good outcomes.[93] In another, death was four times more likely in patients whose 5-drug regimen relied on pyrazinamide but whose isolate tested resistant when compared to patients with pyrazinamide-susceptible isolates receiving pyrazinamide in a 5-drug regimen.[94]

Pyrazinamide is a widely used anti-TB drug. And, pyrazinamide resistance is estimated to occur in 50% or more of MDR-TB patients.[92,95] Although progress has been made in the development of molecular tests for pyrazinamide resistance,[96] reliable information about isolate susceptibility to pyrazinamide is not generally available at the time a regimen is developed. Consequently, WHO recommends that pyrazinamide be universally included in MDR-TB regimens.[97] In acknowledgement of the important role pyrazinamide plays when active and the uncertainty about activity at the time of treatment initiation, all endTB experimental regimens contain pyrazinamide plus at least three other likely effective anti-TB agents.

*Efficacy of Shortened Treatment*

WHO approved a shortened regimen for treatment of selected MDR-TB patients in 2016.[2] This decision was supported by recent clinical studies. First, clinical treatment outcomes of MDR-TB patients on an effective, shortened regimen were reported in a long-term observational study in Bangladesh. 206 patients received a minimum of 9 months of treatment consisting of high-dose gatifloxacin, clofazimine, ethambutol, and pyrazinamide.[62] Treatment regimens also contained prothionamide, kanamycin and high-dose isoniazid for a minimum of 4 months. This cohort had a relapse-free cure rate of 87.9%.[62] More recent studies from Bangladesh and from Africa report similarly successful results with shortened, late-generation fluoroquinolone-containing regimens.[63,98,99] These regimens are being further evaluated in the STREAM study (ISRCTN78372190)[100] and several cohort studies in Africa and in Uzbekistan.

The impressive and consistent results of 9-month MDR-TB treatment in select populations has engendered confidence that regimens containing similar combinations may be significantly shortened, at least in populations with relatively low rates of resistance to second-line drugs. The hypothesis that these or other, shortened regimens could be effective in populations with more significant resistance profiles has yet to be tested. The achievement of success in such populations likely hinges on regimen composition that achieves similar early bactericidal and sterilizing activity to the “Bangladesh” regimen with drugs to which there is less population exposure. The endTB regimens aim to meet these goals. The components of such potentially successful, shortened regimens are described above; the rationale for their combination is presented below.

*Combinations*

A number of preclinical studies have been performed in the last several years that lend support to the combinations proposed here.[57,101,102] These studies suggest shortening potential of the new combinations by demonstrating effects on relapse-free survival, as well as on lung CFU counts during treatment. Combination studies have focused primarily on new anti-TB drug classes, bedaquiline and nitroimidazoles (delamanid, pretomanid, TBA-354) or oxazolidinones (linezolid sutezolid, posizolid). These drugs have been explored in combination with existing drugs, most often pyrazinamide, moxifloxacin, and rifapentine and in some cases with other second-line oral and injectable agents.

Bedaquiline-pyrazinamide combinations containing, as a third drug one of the following: moxifloxacin, linezolid, rifapentine, or clofazimine all demonstrated bactericidal activity at least equal to or greater than the standard first-line regimen on fully susceptible strains (Figure 1).[102] endTB regimens 1-3 (Table 1) build on this active nucleus of bedaquiline and pyrazinamide by adding a fluoroquinolone, levofloxacin (Le) or moxifloxacin (Mo), and a second drug with anti-TB activity. In the mouse studies below, addition of moxifloxacin to this nucleus (JZM, similar to endTB regimen 1) resulted in among the greatest declines in CFUs across the regimens—both between time 0 and month 1 and between months 1 and 2.

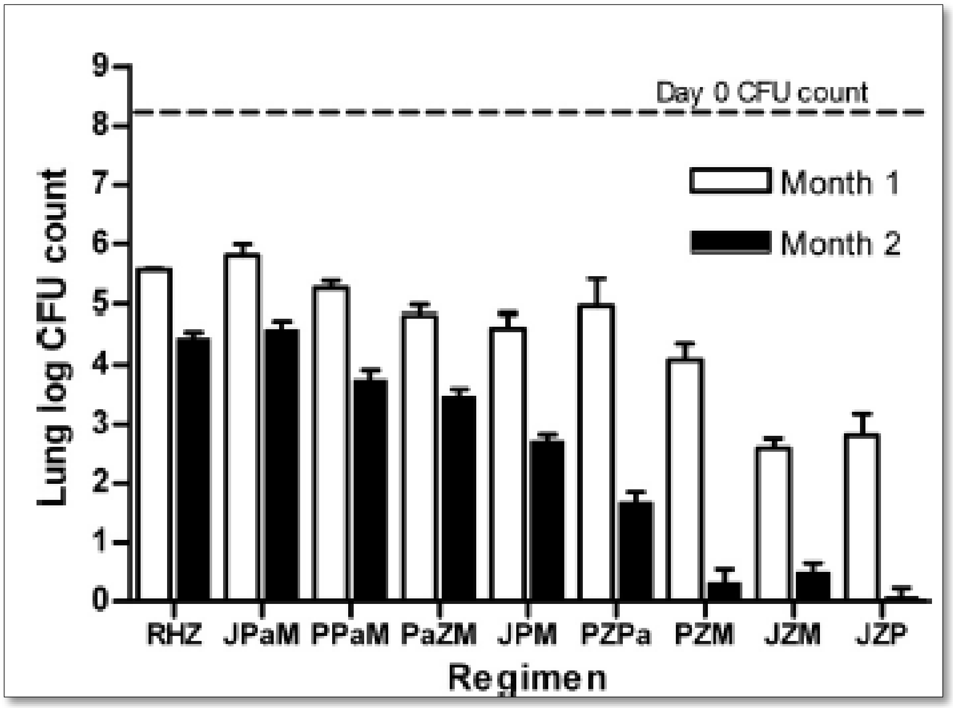

**Figure 1 Mean lung log10 CFU counts (±SD) after 1 and 2 months of treatment**

Abbreviations: R, rifampin; H, isoniazid; Z, pyrazinamide; J, bedaquiline; Pa, pretomanid; M, moxifloxacin; L, linezolid; P, rifapentine; C, clofazimine.[102]

In mice, several 3-drug regimens (including JMZ, JZL, JZC in Figure 2) prevented all relapse after 4 months of treatment and at least 93% of relapse after 3 months of treatment. In the case of JZC, there was no relapse after only 2 months of treatment.[102]

Benefits of clofazimine were further demonstrated in mouse experiments that compared a regimen containing drugs typically used in the treatment of MDR-TB with and without clofazimine, especially in the absence of the injectable agent after month 2.[61] This is supported by the documented efficacy of clofazimine *in vitro*[103] and in other mouse models.[104]

endTB regimens 3-5 all contain delamanid, a nitroimidazole with good clinical evidence for its efficacy against MDR- and XDR-TB.[11,38,105] Delamanid was selected as the nitroimidazole for several reasons. First, delamanid appears more potent than pretomanid and approximately equally potent to TBA-354.[51] Second, selection for delamanid-resistant mutants may be less common than selection for pretomanid-resistant mutants.[52] And, third, clinical development of delamanid was more advanced than for pretomanid and TBA-354, having achieved conditional approval by a stringent regulatory authority, at the time the decision was made. Lastly, the TB Alliance and MSF-Holland are conducting complementary trials with pretomanid while only a single trial in Korea aims to examine an all-oral delamanid-containing combination (NCT02619994).

Published pre-clinical studies that optimize delamanid-containing combinations are not available. More animal data exist for the use of the other nitroimidazoles in combination. Results from studies containing a different class member cannot be directly extrapolated to delamanid. Nevertheless, since delamanid is thought to be more active and less prone to selection for resistant mutants than pretomanid (and similar to TBA-354),[51,52] these studies provide complementary information for selection of regimens that adhere to the principles described above. Tasneen and colleagues showed consistent shortening potential in mice combining a nitroimidazole and an oxazolidinone. This combination is included in endTB regimens 3 and 4. The oxazolidinone used in endTB, linezolid, may be less active against TB (and more toxic)[106] than others under development. Linezolid, however, is approved for other indications, registered in many countries with important MDR-TB problems, and available for inclusion in the trial regimens, unlike the other oxazolidinones. Although Williams and colleagues found that a regimen containing a nitroimidazole, an oxazolidinone, and clofazimine had weak activity at months 1 and 2 and failed to prevent relapse after 4 months of treatment in mice,[57] the regimen studied contained neither pyrazinamide nor a fluoroquinolone which all endTB regimens contain. When active, pyrazinamide is one of the most sterilizing drugs used in TB treatment. And, Grosset and colleagues found less than 10% relapse among mice treated with clofazimine, pyrazinamide, and a fluoroquinolone (moxifloxacin) for at least 8 months.[61] This supports the selection of endTB regimen 5, notwithstanding the aforementioned limitations of the treatment duration in mice to predict that required in humans to prevent relapse.[86]

As with the other benefits accruing to nitroimidazole combinations, it is reasonable to expect that efficacy benefits found in pretomanid-bedaquiline combinations would be similar in delamanid-bedaquiline combinations. Although some antagonism has been reported with bedaquiline and nitroimidazole combinations,[52,102] its combination with pyrazinamide prevented more treatment relapses in an aerosol model of mice than did a regimen comprising isoniazid, rifampicin and pyrazinamide.[102] Similar results were found when pyrazinamide was replaced with an oxazolidinone, sutezolid or linezolid, as the companion drug in the bedaquiline-pretomanid combination.[57] Additional work by the same group reinforced the finding that a regimen containing bedaquiline, pretomanid, and an oxazolidinone was superior to the first-line combination. And, the addition of pyrazinamide further enhanced activity.[52] This animal work lends indirect support endTB regimen 3, that containing bedaquiline, delamanid, linezolid, pyrazinamide, and levofloxacin.

| Drug regimen | No. positive/total (%) after treatment for: | | | | |
|---|---|---|---|---|---|
| | 2 mo | 3 mo | 4 mo | 5 mo | 6 mo |
| RIF + PZA + INH[b] | ND[a] | ND | 7/14 (50) | 2/14 (14) | 0/14 (0) |
| RIF + PZA + PA-824 | ND | 15/15 (100) | 12/13 (92) | | |
| TMC + PZA | ND | 0/15 (0) | 0/15 (0) | | |
| TMC + PZA + PA-824 | ND | 1/15 (7) | 0/13 (0) | | |
| TMC + PZA + MXF | ND | 1/15 (7) | 0/15 (0) | | |
| TMC + PZA + LZD | ND | 0/15 (0) | 0/15 (0) | | |
| TMC + PZA + RIF | ND | 0/15 (0) | 0/15 (0) | | |
| TMC + PZA + RPT | 0/15 (0) | 0/14 (0) | 0/15 (0) | | |
| TMC + PZA + CFZ | 0/14 (0) | 1/14 (7) | 0/15 (0) | | |

[a] ND, not done.

**Figure 2 Proportions of mice with *M. tuberculosis*-positive cultures 3 months after completing treatment[102]**

The significantly shorter, novel regimens resulting in relapse-free cure in mice, coupled with the results of the new-drug trials, and the clinical evidence from the Bangladesh regimen all offer encouragement. The latter reminds us that the regimens that resulted in recommendations of 18-24 month treatment did not contain any fluoroquinolone,[107] or any drug with activity comparable to that of the late-generation fluoroquinolones that have been included in the successful studies of 9-month regimens for MDR-TB and that are used in endTB.

As noted, all endTB experimental regimens contain at least four drugs that are active against *M. tuberculosis*. This is consistent with current WHO guidance on the treatment of MDR-TB.[5] Two are drugs to which population exposure is limited (selected among bedaquiline, delamanid, linezolid, and clofazimine). All regimens also contain two drugs to which prior population exposure is extensive but either susceptibility is confirmed as part of inclusion (fluoroquinolone) or susceptibility is unknown at inclusion, but, when active, the effect on treatment shortening is pronounced (pyrazinamide).[108] In addition, the regimens avoid combination of more than two major QT-interval prolongers.

**endTB regimen 1 (BeLiMoZ)** contains 3 drugs with bactericidal activity and 2, plus pyrazinamide, with sterilizing activity (bedaquiline and linezolid). The 2 major QT-interval prolongers in the regimen are bedaquiline and moxifloxacin. That this four-drug regimen represents a viable option for shortened treatment is supported by previously described mouse studies (figure 2).[102] One of the 3-drug regimens that prevented all relapse after 4 months of treatment and at least 93% of relapse after 3 months of treatment is the nucleus of this regimen—bedaquiline, moxifloxacin, pyrazinamide—to which linezolid is added. There is no evidence of antagonism between moxifloxacin and linezolid, the drugs each used in one regimen. In fact, combinations of moxifloxacin, pyrazinamide with sutezolid (another oxazolidinone) had 2-month murine activity comparable to those drugs in combination with rifampin.[109]

**endTB regimen 2 (BeLiCLeZ)** changes regimen 1 by replacing moxifloxacin with levofloxacin (to reduce the QT liability) and clofazimine. It starts with the nucleus of bedaquiline, pyrazinamide, and clofazimine, which resulted in no relapse after only 2 months of treatment in the mouse.[102] To this, the regimen adds linezolid and levofloxacin. It contains at least 2 drugs with bactericidal activity (bedaquiline, levofloxacin, and linezolid) as well as at least 2 (beyond pyrazinamide) with some sterilizing activity (bedaquiline, linezolid, and clofazimine). The two major QT-interval prolongers are clofazimine and bedaquiline. Levofloxacin was substituted for moxifloxacin to reduce the QT liability.

**endTB regimen 3 (BeDeLiLeZ)** is the only regimen to contain both newly approved drugs, bedaquiline and delamanid. Like the others, it boasts at least 2 drugs with limited population exposure with both bactericidal and sterilizing activity. It has 1 major (bedaquiline) and 2 minor (delamanid and levofloxacin) QT-interval prolongers. No study data are available on the co-administration of bedaquiline and delamanid. The efficacy of a bedaquiline-nitroimidazole (pretomanid and TBA-354) combination is well supported in mice.[52] Evidence for the combination in humans is provided by two 14-day early bactericidal activity studies: (1) showing change in $\log_{10}$CFU similar to that of first-line treatment for drug-susceptible tuberculosis, at least in the first week,[110] and (2) demonstrating that a bedaquiline-pretomanid-pyrazinamide combination was at least as active the standard first-line therapy.[66]

The combination of bedaquiline and delamanid has been avoided largely because of concerns about additive (or worse) cardiotoxic effects (see section 2.3.1).

**endTB regimen 4 (DeLiCLeZ)** contains 3 drugs with some bactericidal activity (delamanid, linezolid, and levofloxacin) as well as 3 (+PZA) with treatment-shortening benefits (delamanid, linezolid and clofazimine). This 5-drug regimen contains 1 major (clofazimine) and 2 minor (delamanid and levofloxacin) QT-interval prolongers.

**endTB regimen 5 (DeCMoZ)** offers 2 drugs with some bactericidal activity (delamanid moxifloxacin) as well as 2 (+PZA) with treatment-shortening benefits (delamanid and clofazimine). One of two four drug, moxifloxacin-containing regimens, it may have a reduced toxicity profile compared to the five-drug regimens. It does, however, contain 2 major (clofazimine and moxifloxacin) and 1 minor (delamanid) QT-interval prolongers.

##### 2.2.3 Rationale for Study Design

*Non-Inferiority Study*

The current study aims to make the following improvements over the conventional regimen for MDR-TB:

1. Shorten treatment from 18-24 months to 9 months;
2. Eliminate the injectable and establish all-oral regimen(s);
3. Reduce the toxicity profile, including for patients co-infected with HIV;
4. Enhance treatment adherence and completion; and
5. Improve the efficacy.

Achieving points 1 - 4 above even without improving the efficacy of the current regimen would confer benefits to populations as well as to individual patients. These changes would likely improve prospects for expanding access to treatment beyond the 23% of MDR-TB patients reported to have received any treatment with second-line drugs.[2] Achieving these objectives may also improve compliance with treatment. Improved compliance, in turn, could translate into reduced frequency of loss-to-follow-up of patients on treatment. This frequency was most recently estimated at 16% in the 2012 treatment cohort reported to WHO.[3] Both these changes would have important epidemiological implications, which are reducing transmission of and morbidity and mortality from MDR-TB. In addition, shorter, less-toxic regimens could engender quality-of-life and economic benefits for patients who are able to return to activities of daily life sooner. In light of all these potential benefits *without* an improvement in efficacy, a non-inferiority design was selected for the endTB trial.

Non-inferiority margins are generally elected based on a known "consistent and reliable estimate of the efficacy of active treatment relative to placebo."[111] This is largely to avoid the phenomenon of biocreep in which the standard for efficacy of a new regimen declines over sequential non-inferiority trials, eventually, possibly becoming less efficacious than placebo.[112] In the case of MDR-TB as well as for many other bacterial infections, no such estimate exists. There are, however, some data on which to base the selection of the non-inferiority margin. The justification for the selection of the non-inferiority margin is in section 11.

*Adaptive Randomization*

The endTB trial has elected Bayesian adaptive, rather than fixed, randomization for this study. As with other methods, the sample size calculation in a Bayesian adaptive-randomization study is based on assumptions around statistical power, treatment response, effect size, and tolerance for type I error, made prior to the initiation of the study. Bayesian methods, which have been addressed in guidance by stringent regulatory authorities,[113] have been increasingly employed in oncology trials. In Bayesian outcome-adapted randomization, trials use 'real-time' information on treatment response and effect size to adjust the probability of randomization: it favors arms with greater treatment response. These and other adaptive trial designs (e.g., MAMS) are generally found to be more efficient (larger power/sample size ratio) than fixed-randomization designs. Adaptive designs "learn" from the trial and incorporate new information into its execution, refining the assumptions made before trial initiation. For these reasons, endTB opted to use an adaptive design. Bayesian adaptive randomization was selected in particular, because of evidence of its particular efficiency advantage in studies with more than one effective regimen.[114] Since endTB is exploring 5 experimental regimens, and aims to identify all of those that are effective, a design that permits doing this as efficiently as possible was chosen.

*Rationale for the timing of the primary endpoint evaluation*

The scheduled end of treatment in experimental arms is 39 weeks (i.e., 9 months). The primary endpoint will be measured at Week 73 post randomization, effectively 34 weeks after scheduled completion of the experimental regimens. Although very limited relapse has been reported among patients receiving the Bangladesh-type regimens,[62,63,98,99] recurrence due to relapse remains the primary concern associated with shortening regimens. Evidence from previous studies reveals that the greatest risk of recurrence due to relapse is in the period immediately following treatment completion.[115,116] And, it has been recommended to shorten follow-up in TB trials to a minimum of 6 months.[117] endTB has, therefore, elected to follow all participants on experimental regimens for at least 6 months (26 weeks) after completion of the regimen to evaluate potential relapse. Participants may take as long as 47 weeks to complete 39 weeks of treatment. The Week 73 endpoint permits 6-month evaluation of those participants who require the full 47 weeks.

*Rationale for randomization of reduced linezolid dose*

Those patients randomized to endTB regimens 1-4 will undergo linezolid dose-reduction either at Week 16 or after a linezolid-related AE requiring dose reduction, whichever is earlier. One prior clinical trial has evaluated the role of linezolid dose reduction, comparing a strategy of continuation of linezolid 600 mg daily to dose-reduction to 300 mg daily.[76] In the endTB trial, randomization of the reduced linezolid dose will allow investigation of the effect of these linezolid dose-reduction strategies on efficacy and toxicity, prospectively, in a large, heterogeneous population in a randomized controlled trial. Balanced randomization will mitigate bias, such as confounding by indication, which may be introduced if site clinicians choose the dose-reduction strategy.

#### 2.3 Potential Risks and Benefits

##### 2.3.1 Potential Risks

All endTB experimental regimens have a similar duration as the standardized shorter regimen recently recommended by the WHO[2], and they are substantially shorter than WHO conventional regimens.[5] There is, therefore, a potential for increased risk of treatment failure or relapse and emergence of drug resistance compared to the conventional regimen. Accumulating data on the use of shortened regimens containing no new drugs estimates the risk of failure on these regimens to be low (<2%).[62,63,98,99] endTB regimens do contain fewer drugs than those used in the cited studies. All endTB experimental regimens, however, include at least 3 drugs to which the bacilli are highly likely to respond, including at least one new, highly active drug. For the treatment of drug-susceptible TB, three drugs were demonstrated to be sufficient in patients whose isolates were susceptible to all the drugs; the fourth drug, ethambutol, was added later to provide additional coverage in case of baseline, undetected isoniazid resistance. Whether the endTB experimental regimens combine to confer the same treatment benefit in MDR-TB patients is unknown. Mouse models certainly support this hypothesis.[102,118] Nevertheless, to avert the risk that occult baseline resistance to drugs contained in endTB experimental regimens compromises treatment efficacy, the trial avoids using drugs that have high prior population exposure (isoniazid, ethambutol, thioamides, aminoglycosides; see section 2.2.2).

However, the possibility exists that one or more experimental regimens are insufficient to effect sustained cure. The alternative considered was adding more drugs; regimens 2, 3, and 4 contain 5 drugs, providing additional protection in case of resistance. Any other drugs that could be added confer considerable toxicity risks and may be compromised by substantial prior patient exposure. Increased efficacy, therefore, is not assured by adding these drugs to the experimental regimens. Increased toxicity and treatment complexity would, however, be virtually certain. Therefore, we have opted to exclude additional drugs and test simpler, less toxic regimens.

The risk of relapse after MDR-TB treatment is not well documented. However, the decision to adjust duration to 9 months is supported by emerging evidence from cohort studies that 9- and 12-month regimens without a new drug do not have a significantly increased risk of relapse.[62,63,98] In one study of treatment using the conventional regimen (18-24 months of treatment), 2 years of follow-up after standard treatment duration identified approximately 5% recurrent cases (no distinction could be made between reinfection and relapse).[116] Aung *et al.* estimated <1% relapse after 9-12 months of treatment in Bangladesh and a similarly small number of recurrent cases.[63] Piubello *et al.* recorded no relapse among 58 patients followed for 24 months after completion of a 12-14-month regimen, while 8.6% died during the follow up period.[98] None of these reports included regimens containing one of the new drugs and there is no evidence for the optimal duration of MDR-TB treatment containing a new drug. In the pivotal studies of delamanid and bedaquiline, study participants receiving standard background regimens plus delamanid or bedaquiline had improved the early and late treatment responses .[10,11,38,119] Incorporation of one or both of these new drugs may yield results similar to those reported with the shortened regimens. It is important to note that the recommended 20-month duration for conventional regimen is based largely on observational studies developed on a foundation of treatment without fluoroquinolones[107] and without a member of a new, highly active drug class (nitroimidazole and ATP-synthase inhibitor). The endTB experimental regimens contain both.

Emergence of additional drug resistance is estimated to be rare among patients who fail/relapse even after these shortened regimens.[63] The principles applied to avert failure of endTB regimens may also reduce risk of resistance amplification in the trial. Nevertheless, there are concerns about mutations in a single gene, *rv0678*, that may confer non-target based resistance to both bedaquiline and clofazimine, through a shared efflux pump.[120,121] This is a potential concern for one of the proposed regimens (regimen 2). The frequency of this event has yet to be established in the course of clinical care. Moreover, to date, there is no evidence that such mutations predict treatment failure. Specifically, the risk is unknown when the drugs are started simultaneously and accompanied by a likely active, late-generation fluoroquinolone, and there is no pre-existing evidence of the relevant mutation in the *rv0678* gene. To mitigate this risk, the endTB study population is limited to MDR-TB patients without molecular evidence of resistance to fluoroquinolones; and, regimen 2 contains 3 other drugs.

The main safety concerns for the experimental regimens are cardiotoxicity, hematologic disorders and neurotoxicity. The role of individual drugs in these risks, as well as the available information on the risk of combinations, is described below.

##### *Cardiotoxicity*

The primary indicator of potential cardiotoxicity is QT-interval prolongation; the correlation between QT-interval prolongation and cardiotoxicity, however, is limited. There have been no reported cardiac events attributable to the drugs used in treatment of MDR-TB. All 5 of the experimental endTB regimens and the control regimen contain drugs with QT prolonging effects (bedaquiline, clofazimine, moxifloxacin, and to a lesser extent, delamanid and levofloxacin) and/or drugs that promote pro-arrhythmic conditions (e.g., aminoglycosides).[122] This section focuses on the new drugs, bedaquiline and delamanid and their combination, and moxifloxacin; additional information on these drugs and other drugs used routinely in MDR-TB treatment that prolong the QT interval (fluoroquinolones, clofazimine) or increase the risk of arrhythmia (aminoglycosides, polypeptides) is available in the summary of product characteristics and/or package inserts provided by the Sponsor (Reference Safety Information).

Bedaquiline

Bedaquiline has established QT-prolonging effects. In one repeat-dose toxicity study in dogs, an increase (up to 16%) in QTc-interval was noted after 2 months of administration at a dose of 40 mg/kg/day. It is noteworthy that the bedaquiline plasma concentration was approximately 8-fold higher in dogs than in humans. In clinical studies, the mean observed increases in QT-interval corrected by Fridericia formula (QTcF) were: 9.3 ms at Week 2 and >10 ms at Week 8.[123] The largest increase in QTcF during 24 weeks of a bedaquiline + conventional regimen was observed at Week 18, reaching a mean increase of 15.7 ms.[10] There was no ventricular arrhythmia, torsade de pointes, or clinically significant dysrhythmia. Although excess deaths were reported in the bedaquiline-containing arm, none of the deaths was attributed to bedaquiline by the investigators. And, no association was found between QTc-interval prolongation and death in the clinical studies.[10,37]

As noted, endTB regimens contain other known QT-interval-prolonging drugs. The possibility exists that QT-interval prolongation and/or frequency of cardiotoxic events could be increased with the concurrent use of two or more such drugs. Existing data on this risk is limited, but derives mostly from the Phase II trials of one of these drugs with companion drugs included in optimized background regimens (OBR). When bedaquiline was used with clofazimine (and other, unspecified drugs) in 17 subjects, the QTcF increased by a mean of 31.9 ms while the increase was 12.3 ms in 177 subjects without concomitant clofazimine use. Prolongation of >500 ms occurred in 2 patients receiving bedaquiline and clofazimine; one had concurrent hypokalemia.[37]

In a clinical case series of 35 patients treated with bedaquiline, the QTcB (QT-interval corrected by Bazett formula) interval increased a median of 4.9 ms in patients who received bedaquiline combined with FQ; 7.3 ms when bedaquiline was combined with clofazimine, and -5.3 ms when not combined with either. bedaquiline was discontinued in 2(6%) patients due to persistent QTcB prolongation: one receiving moxifloxacin, and the other clofazimine and amiodarone. No cardiac arrhythmia or other clinical cardiac event occurred.[124]

Delamanid

Non-clinical data reveal that delamanid and/or its metabolites have the potential to affect cardiac repolarization via blockade of hERG potassium channels. In the Phase II trial of delamanid, the frequency of a prolonged QTcF interval was higher in both delamanid groups than in the placebo group: 13.1% in the delamanid 200 mg twice daily arm, 9.9% in the delamanid 100 mg twice daily arm, and 3.8% in the placebo arm. Among participants in the open-label trial, who received as much as 8 months of delamanid exposure, the proportion with QTcF prolongation did not increase, remaining under 10%. There was no ventricular arrhythmia, torsade de pointes, or clinically significant dysrhythmia. The authors conclude that QTcF prolongations occurring with delamanid exposure are "not associated with cardiac clinical manifestations.” [105]

In the delamanid studies, the combination of delamanid and levofloxacin, as part of the OBR, was explored: QTcF prolongation (defined as >500 ms) occurred in 3/94 patients receiving concomitant delamanid and levofloxacin and in 2/96 patients receiving placebo + levofloxacin. The magnitude of the increase from baseline was similar: 10.7 ms among those receiving both delamanid and levofloxacin compared to 12.1 ms among all participants who received delamanid for at least 1 month. Longer exposure achieved through continued delamanid administration in the open-label trial (208) did not change the proportion experiencing QTcF prolongation; it remained just under 10%. Lastly, there is no evidence that concomitant exposure to delamanid and clofazimine, although the latter was used rarely in the 204/208 studies, increases the cardiac risk of delamanid. QTcF prolongations occurring with delamanid exposure in 204/208 studies were not associated with cardiac clinical manifestations.[125]

In the trial, patients with low baseline serum albumin levels will not be excluded. This is in contrast with the EMA label for delamanid, but supported by increasing evidence.
The EMA-authorized label for delamanid includes serum albumin <2.8 g/dL as a contraindication. Moreover, it notes that "QTc prolongation is very closely correlated with the major delamanid metabolite DM-6705. Plasma albumin and CYP3A4 regulate the formation and metabolism of DM-6705 respectively and the potential risk for low albumin levels to increase the risk of QTc prolongation. In a clinical study, the presence of hypoalbuminaemia was associated with an increased risk of prolongation of the QTc interval in delamanid treated patients. Patients who commence delamanid with serum albumin <3.4 g/dL or experience a fall in serum albumin into this range during treatment should receive very frequent monitoring of ECGs throughout the full delamanid treatment period. " The referenced study is the Phase II pivotal trial.
However, recent results from a large Phase III trial did not reveal increased QTcF prolongation among those with baseline albumin <2.8 g/dL who received delamanid + OBR over the course of investigational exposure.[126] While participants who received delamanid and had baseline albumin of <2.8 g/dL had a maximum QTcF change of 10.8 ms (by week 9), participants with higher albumin (>2.8 g/dL) in the delamanid arm had a similar maximum QTcF change, 9.5 ms by week 8. And, participants with baseline albumin of <2.8 g/dL who received placebo had a higher maximum QTcF change, 14.5 ms (by week 11).
Further evidence is provided by another recent trial, ACTG 5343, which examined change in QTcF prolongation among patients who received bedaquiline + OBR, delamanid + OBR (N=25), or bedaquiline + delamanid + OBR (N=24) for 6 months with ECGs done at baseline, week 8, and every two weeks from 8-24 weeks.[127] Maximum mean change in QTcF was higher in the bedaquiline + delamanid arm, but no QTcF intervals >500 ms were observed in either arm. No overall pattern could be discerned between baseline albumin, delamanid exposure, and QTcF prolongation. While the maximum mean change was higher in the <2.8 g/dL group than in the ≥2.8 g/dL group in the bedaquiline + delamanid arm, it was lower in the delamanid arm. Numbers of participants with albumin <2.8 g/dL were extremely low in both arms.

In summary, data that examine the association of QTcF interval prolongation and delamanid exposure do not support exclusion of patients with baseline albumin of <2.8 g/dL from the trial. Rather, there is significant evidence from that albumin increases with appropriate anti-TB therapy, including in MDR-TB patients.[128–130] And, given the intensive electrocardiogram monitoring schedule for all study participants, any increased risk associated with inclusion of patients with lower albumin will be promptly identified and managed according to study procedures.

Combination of bedaquiline and delamanid

The safety profile of bedaquiline combined with delamanid (as in endTB regimen 3) has not previously been studied. Experience with bedaquiline and pretomanid, as well as with other combinations, might be informative. In the early bactericidal activity (EBA) study of bedaquiline and pretomanid no corrected QT interval prolongation greater than 500 ms occurred.[110] One subject receiving pretomanid, moxifloxacin, and pyrazinamide experienced prolongation of greater than 60 ms increase from baseline; no subjects receiving bedaquiline and pretomanid experienced this prolongation.[110] In the 8-week study, no QTc (F and B) prolongation greater than 500 ms occurred nor did any clinical cardiac events. Increases of 60 ms or greater QTcB were reported in 13.3%, and QTcF in 26.7%, of patients receiving bedaquiline, pretomanid, and clofazimine, at least twice than the proportion recorded in the clofazimine only arm (6.7% QTcB and F).[66,110] Bedaquiline and pretomanid (and linezolid) are currently being combined in a study for a duration of at least 6 months (NCT02333799); toxicity data are not yet available.

Two case reports of XDR-TB patients treated with the combination of bedaquiline and delamanid have been published and lend further, even if anecdotal, evidence. In one case, a pulmonary XDR-TB patient, the association was well tolerated and no QT prolongation occurred.[131] In the second case, QT interval prolongation was observed during XDR-TB treatment with a drug combination including clofazimine, bedaquiline and delamanid; following the QT increase, bedaquiline was interrupted and re-introduced after 5 days. No QTcF>500 ms was recorded, nor any cardiac arrhythmia occurred.[132] A small case series offers additional evidence on the association of these two drugs. Three MDR-TB patients received bedaquiline and delamanid for 1, 3, and 6 months respectively; two additional patients were switched from bedaquiline to delamanid without a washout period, effectively exposed to the two drugs simultaneously in light of the lengthy half-life of bedaquiline. So far, no ≥60 ms QTc prolongation, >500 ms QTc values or clinical cardiac events have been observed in this group of patients (M. Jachym, personal communication).

In sum, QT prolongation reveals is observed when bedaquiline or delamanid is used in conjunction with other QT-prolonging anti-TB drugs. To date, however, no cardiac events have been reported among patients receiving either bedaquiline or delamanid. Moreover, although very limited data are available on their concurrent use, nothing in that experience cautions against testing combinations in carefully monitored settings.

ACTG 5343, the National Institute of Allergy and Infectious Diseases (NIAID)-sponsored combination study of bedaquiline and delamanid, was slated to provide the first evidence from a well-conducted trial of the safety and efficacy of delamanid and bedaquiline in combination. That trial, however, has encountered significant delays since it was first proposed in 2014. It is now scheduled to start by mid-2016. Even if enrollment is completed in 6 months, with 6 months of treatment and 24 months of follow-up, results will not be available before mid-2019. Moreover, ACTG 5343 will enroll only 84 participants, 1/3 of whom will be exposed to bedaquiline and delamanid in combination. Although endTB will not report results prior to ACTG 5343, beginning contemporaneously will provide data on safety of the combination in at least as many subjects (possibly 2-3 times more) as ACTG 5343.

Moxifloxacin

Moxifloxacin, while significantly prolonging the QT interval,[87] has a relatively safe cardiac profile[133] with clinical cardiac toxicities occurring in only 0.1% of more than 10,600 patients receiving oral moxifloxacin (and in 0.1% among comparators); no cases of torsades de pointes were reported among the 10,600 patients exposed to oral moxifloxacin.[133] In a registry in the United States, only one case of torsades de pointes was notified out of more than one million patients treated with moxifloxacin.[134] In another study, torsades de pointes were notified less than once per million of prescriptions of levofloxacin, while no cases were reported in association with moxifloxacin after 1.4 million prescriptions.[135] Moxifloxacin activity against *M. tuberculosis* lends justification for its use in MDR-TB patients, despite the potential cardiac liability. Additionally, another late-generation fluoroquinolone, gatifloxacin used in higher than standard doses, has been part of a clofazimine-containing regimen for MDR-TB. The excellent results, exceeding 80% success, further argue for the assumption of risk attendant to the combination of clofazimine and late-generation fluoroquinolones.[62,63,99]

Risk mitigation

To balance the potential cumulative cardiotoxicity risks with the potential efficacy benefits of the combination regimens, the protocol limits to two the number of major QT prolongers without other pro-arrhythmic drugs used in the regimens. Exceptions to this principle include a limited number of control arm regimens, which may include more than two of the following: bedaquiline, clofazimine, moxifloxacin,[134] and an aminoglycoside/polypeptide.

Monitoring of all study participants includes intensive, electrocardiogram (ECG) monitoring (see Schedule of Events, Appendix 1). Monitoring frequency is reduced after an initial period in accordance with data from the observational cohorts of bedaquiline-treated patient in South Africa[136] and France[137] in which the maximum increase in QTc interval was recorded after two months of bedaquiline administration. In studies C208 and C209, QTcF continued to increase through 24 weeks of treatment: the mean prolongation, however, was between 10 and 15 ms at both 12 and 24 weeks.[37]

*Hematologic disorders and peripheral neuropathies*

Linezolid-containing endTB regimens, both experimental and control, carry the risk of hematologic disorders (anemia, thrombocytopenia, myelosuppression, and leukopenia), optic and peripheral neuropathy. The hematologic disorders are reversible and dose related. Peripheral and optic neuropathy can be irreversible. The systematic review and meta-analysis published by Sotgiu et al.[75]found that almost 60% of patients receiving linezolid experienced AEs including: anemia (38%), peripheral neuropathy (46%), gastrointestinal disorders (17%), optic neuritis (13%), and thrombocytopenia (12%). The frequency of AEs was significantly lower when a daily linezolid dose of 600 mg or less was used, instead of more than 600 mg. Differences were apparent for anemia (22% vs 60%), peripheral neuropathy (40% vs 57%) and optic neuritis (10% vs 17%).[138] This finding was confirmed by another systematic review and meta-analysis showing that the frequency of adverse events leading to treatment discontinuation was reduced by half among patients receiving ≤600 mg (29%) compared to those receiving >600 mg daily linezolid dose (61%).[42] Good success and a lower frequency of AEs were reported with linezolid 300 mg once daily in a case series of 21 M/XDR-TB patients[139] and in a retrospective study of 51 M/XDR-TB patients.[140] Another cohort of 34 MDR-TB patients in South Africa, slightly more than half HIV-coinfected, was started on a linezolid dose of 600 mg (with the exception of 2 patients with BMI <14 who received 300 mg/day). Fourteen experienced adverse events. With dose reductions, tolerability was similarly improved: seven continued linezolid treatment at their starting dose, six improved after reduction from 600 to 300 mg daily linezolid dose and only one had to stop the treatment.[141] In a clinical trial, 33 participants initially randomized to receive either immediate or delayed linezolid 600 mg daily underwent a second randomization, implemented after sputum smear conversion or receipt of linezolid for 4 months. Participants were randomized to receive linezolid dosed at 600 mg daily or at 300 mg daily.[76,142] Remaining at 600 mg daily was associated with an increased hazard of linezolid-related AEs compared to dose-reduction to 300 mg daily (hazard ratio of 2.7).[76,142] The frequency of linezolid-AEs was 88% in those who remained at 600 mg daily and 69% in those dose-reduced to 300 mg daily.

Alternatives considered to avoid the toxicity associated with linezolid include using new oxazolidinones (e.g., sutezolid, posizolid), which may cause less toxicity. These experimental agents, however, are not yet available for inclusion in a Phase III trial of combination regimens and will not be available for routine use for MDR-TB treatment for some years. Another promising alternative to dose reduction is intermittent (thrice weekly) linezolid treatment, which has been shown to reduce side effects, compared to daily dosing, while achieving good outcomes and satisfactory drug concentrations in the blood in a small cohort of patients.[143] This approach is supported by pre-clinical and clinical data, summarized by Nuermberger[144], that illustrate that trough concentrations ($C_{min}$) or time during which linezolid concentrations are likely to inhibit mitochondrial protein synthesis predict linezolid toxicity.

In light of the relatively well-established anti-TB activity of linezolid,[42,75,76,77] including unpublished reports of linezolid efficacy in humans and mice when delivered for periods of four months or less,[145] the emphasis in endTB will be on mitigating toxicity. This will address an important gap in our understanding of dosing strategies that may reduce linezolid-related toxicity and permit its use with less toxicity monitoring. This will be accomplished by a secondary randomization (1:1) of participants in the linezolid-containing experimental arms (endTB regimens 1-4) to either of 300 mg daily or 600 mg, thrice weekly, at Week 16 (or earlier if indicated by toxicity) of daily 600 mg dosing. In a recent, unpublished EBA study, the daily increase in time to positivity in MGIT between 300 mg daily dose and the 600 mg daily dose had overlapping confidence intervals;[145] as noted above, the former dose has been provisionally linked with reduced toxicity. As summarized above, both strategies may contribute to reduced toxicity after the efficacy benefits of 600 mg daily have been realized. Since available efficacy data support both dosing strategies, there is equipoise about the comparison. Participants will be closely monitored with routine complete blood counts, vision examinations, and screening for peripheral neuropathy (see Schedule of Events, Appendix 1). Dosing will begin at 600 mg /day and will be adjusted at Week 16, or earlier if indicated. This could result in a reduction in neurological toxicity usually occurring after 5 months of daily treatment. The impact may be less pronounced on hematological toxicity, which is usually reported early in the treatment and was reported in a systematic review and meta-analysis to occur in 17.9% of patients who received

≤600 mg daily dosing.[140 ,76]

*Other risks*

Additional concern comes from the lack of data regarding the safety of the specific drug combinations in the experimental arms. However, useful data can be extrapolated from the published randomized clinical trials. In those studies, bedaquiline was given in association with background regimens frequently containing a fluoroquinolone and pyrazinamide while only a small subgroup of patients received clofazimine and linezolid.[10,37] In the main registration Phase IIb trial, study C208, a greater number of deaths was recorded in the bedaquiline arm than in the placebo-controlled arm; no causality link was found between the administration of bedaquiline and the deaths.[10] In addition, this high casualty rate was not confirmed by study C209, an open-label uncontrolled trial testing bedaquiline plus standard treatment regimen for MDR/XDR-TB.[37] In the published cohorts of patients receiving bedaquiline in clinical practice, this drug has been mostly administered in association with linezolid and, in South Africa, with clofazimine, with overall good tolerability and a low death rate.[136,147] Similarly, randomized controlled trials 204 and 208 featured the addition of delamanid to treatment regimens that usually contained pyrazinamide and a fluoroquinolone; rarely did regimens include Group 5 drugs. The safety profile of the reported combinations was reassuring.[11,38]

HIV-coinfected patients represent a population with special risks, in particular the risk of interactions between antiretrovirals and new drugs. The use of lopinavir and ritonavir (a CYP3A4 inhibitor) was shown to increase the exposure to bedaquiline and delamanid.[148,149] The co-administration of these drugs will therefore be contraindicated. In addition, the use of efavirenz (a CYP3A4 inducer) in bedaquiline-containing arms will be disallowed.[150,151] Encouragingly, in the South African group of HIV-coinfected individuals, HIV treatment and MDR-TB drugs were used concurrently without adversely affecting outcomes.[136]

##### 2.3.2 Benefits

All study participants will receive treatment for MDR-TB. Treatment will be well supervised and informed by monitoring and patient support enhanced compared to that routinely provided in most MDR-TB treatment programs.

There are several benefits of the design. First, the design provides the possibility of identifying more than one non-inferior regimen. If more than one regimen is demonstrated to be non-inferior to the current standard, this would provide options for programs and individual clinicians when faced with programmatic (i.e., lack of registration, stock outs) or clinical (i.e., prior exposure/resistance, drug intolerance, concomitant medications) circumstances that demand flexibility in treatment. Second, randomization to the control arm does not preclude possible receipt of a new drug; study participants who are randomized to the control arm and who have an indication for bedaquiline or delamanid, may receive a new drug as part of the control regimen. Lastly, implementation of the study itself, in public-sector facilities, has potential additional short- and long-term benefits. Public-sector facilities often struggle with timely diagnosis, consistent clinical and bacteriologic monitoring, documentation and management of adverse events (AEs), and incorporation of innovation. Implementation of this pragmatic study will support study sites in all of these challenges. Measures implemented as part of this protocol—rapid molecular testing for resistance; systems for clinical accompaniment, transport and processing of high-quality sputum samples; systematic recording of all AEs including SAEs and AESIs, routine ECG monitoring, and expedited reporting of SUSARs—will improve quality of care for study participants and have the potential to improve care for those treated in these facilities in the future.

The study will include HIV-infected patients regardless of their immunologic status. The evidence supporting the use of the new drugs in HIV-positive patients is scarce, as published clinical trials included only a few HIV co-infected patients.[10,37,38] However, the cohort of patients treated with bedaquiline through compassionate use in South Africa included a high proportion (59%) of HIV-infected patients; the safety profile of treatment was promising in this subgroup.[136]

A further benefit of the present effort is the inclusion of a small number of adolescents, which fulfills Principle number 13 of the Declaration of Helsinki for medical research involving human subjects. This principle states that "Groups that are underrepresented in medical research should be provided appropriate access to participation in research." [152]

Finally, the adaptive-randomization design formally uses all available information to increase the probability of participant assignment to effective arms, enhancing benefits of study participation over fixed-randomization designs.[153] Moreover, the adaptive randomization affords greater trial efficiency than fixed randomization, reducing the study sample size by at least 20%; efficiency gains are greater in cases where the difference in response between the experimental and control arms is smaller. This has possible benefits for cost and time to trial completion as well as for risk undertaken by study participants.

### 3 OBJECTIVES

#### 3.1 Study Objectives

##### 3.1.1 Primary Objective

The primary objective is to assess whether the efficacy of experimental regimens at Week 73 is non-inferior to that of the control.

##### 3.1.2 Secondary Objectives

1. To compare the efficacy of experimental regimens at Week 104 to that of the control
2. To compare the frequency of and time to early treatment response (culture conversion) in experimental regimens to that of the control
3. To compare the efficacy of experimental regimens at Week 39 to that of the control
4. To compare, at Week 73 and Week 104, the proportion of patients who experienced failure or relapse in the experimental arms to that in the control arm
5. To compare, at Week 73 and Week 104, the proportion of patients who died of any cause in the experimental arms to that in the control arm
6. To compare, at Week 73 and Week 104, the proportion of patients who experience grade 3 or higher AEs or SAEs of any grade in the experimental arms to that in the control arm
7. To compare, at Week 73 and Week 104, the proportion of patients who experience AESIs in experimental regimens to that in the control arm

##### 3.1.3 Exploratory Objectives

1. To compare safety and efficacy endpoints across experimental arms, e.g., arms containing bedaquiline vs. delamanid (regimen 2 vs. 4); arms containing clofazimine & levofloxacin vs. arms containing moxifloxacin (regimen 2 vs. 1); arm containing bedaquiline and delamanid vs. arms containing one of the drugs and clofazimine (regimen 3 vs. 4 and regimen 3 vs. 2)
2. To evaluate conversion endpoints as potential surrogate markers for unfavorable outcome
3. To compare the activity of regimens between patients with strains that are resistant to important drugs in those regimens (e.g., pyrazinamide) and patients with strains that are sensitive
4. To estimate the frequency of drug resistance amplification among treatment failures and relapses occurring by Week 104
5. To compare treatment adherence in the experimental arms to that in the control and across regimens
6. To compare the frequency of and time to severe linezolid-related toxicity between linezolid dose-reduction strategies.
7. To compare, at Week 73 and Week 104, efficacy endpoints across linezolid dose-reduction strategies: 300 mg daily or 600 mg thrice weekly.

#### 3.2 Study Outcome Measures

##### 3.2.1 Primary Outcome Measures

The primary efficacy outcome is the proportion of participants with favorable outcome at Week 73.

A participant's outcome will be classified as favorable at Week 73 if the outcome is not classified as unfavorable, and one of the following is true:

- The last two culture results are negative. These two cultures must be taken from sputum samples collected on separate visits, the latest between Week 65 and Week 73;
- The last culture result (from a sputum sample collected between Weeks 65 and 73) is negative; and either there is no other post-baseline culture result or the penultimate culture result is positive due to laboratory cross contamination; and bacteriological, radiological and clinical evolution is favorable;
- There is no culture result from a sputum sample collected between Week 65 and Week 73 or the result of that culture is positive due to laboratory cross contamination; and the most recent culture result is negative; and bacteriological, radiological and clinical evolution is favorable.

A participant's outcome will be classified as unfavorable at Week 73 in case of any of the following:

1. Replacement or addition of one or more investigational drugs in an experimental arm or in the control arm if using the shortened regimen (failure);
2. Replacement or addition of two or more investigational drugs in the control arm if using the conventional regimen (failure);
3. Initiation of a new MDR-TB treatment regimen after the end of the allocated study regimen and before Week 73 (relapse);
4. Death from any cause;
5. At least one of the last two cultures, the latest being from a sputum sample collected between Week 65 and Week 73, is positive in the absence of evidence of laboratory cross contamination (failure/relapse);
6. The last culture result (from a sputum sample collected between Week 65 and Week 73) is negative;

   ***AND***
   - there is no other post-baseline culture result or the penultimate culture is positive due to laboratory cross contamination; and bacteriological, radiological or clinical evolution is unfavorable (failure/relapse);

7. There is no culture result from a sputum sample collected between Week 65 and Week 73 or it is positive due to laboratory cross contamination;

***AND***
- the most recent culture is negative; and bacteriological, radiological or clinical evolution is unfavorable (failure/relapse); or,
-the most recent culture result is positive in the absence of laboratory cross contamination

8. The outcome is not assessable because there is no culture result from a sputum sample collected between Week 65 and Week 73 or it is positive due to laboratory cross contamination;

***AND***
- there is no other post-baseline culture result or the most recent culture is positive due to laboratory cross contamination; or,

- the most recent culture is negative and bacteriological, radiological and clinical evolution is not assessable;

9. Previously classified as unfavorable in the present study.[ii]

The procedures for outcome assignment and evaluation of the bacteriological, radiological and clinical evolution will be elaborated in SOPs.

###### 3.2.2 Secondary Outcome Measures

*Efficacy:*

1. Proportion of participants with favorable outcome at Week 104:

- A participant's outcome will be classified as favorable at Week 104 if the outcome is not classified as unfavorable, and one of the following is true:

    a) The last two cultures are negative. These two cultures must be from sputum samples collected on separate visits, the latest between Week 97 and Week 104;
    b) The last culture result (from a sputum sample collected between Week 97 and Week 104) is negative; and either there is no other post-baseline culture result or the penultimate culture result is positive due to laboratory cross contamination; and bacteriological, radiological and clinical evolution is favorable;
    c) There is no culture result from a sputum sample collected between Week 97 and Week 104 or the result of that culture is positive due to laboratory cross contamination; and the most recent culture result is negative; and

[ii] Exception: A participant whose outcome was unfavorable because it was unassessable at Week 39 is eligible for re-evaluation at Week 73.

bacteriological, radiological and clinical evolution is favorable.

- A participant's outcome will be classified as unfavorable at Week 104 in case of any of the following:

  a) Replacement or addition of one or more investigational drugs in the experimental arm or in the control arm if using the shortened regimen (failure);
  b) Replacement or addition of two or more investigational drugs in the control arm if using the conventional regimen (failure);
  c) Initiation of a new MDR-TB treatment regimen after the end of the allocated study regimen and before Week 104 (relapse);
  d) Death from any cause;
  e) At least one of the last two cultures, the latest being from a sputum sample collected between Week 97 and Week 104, is positive in the absence of evidence of laboratory cross contamination (failure/relapse);
  f) The last culture result (from a sputum sample collected between Week 97 and Week 104) is negative;

     ***AND***
     - there is no other post-baseline culture result or the penultimate culture is positive due to laboratory cross contamination; and bacteriological, radiological or clinical evolution is unfavorable (failure/relapse).

  g) There is no culture result from a sputum sample collected between Week 97 and Week 104 or it is positive due to laboratory cross contamination;

     ***AND***
     - the most recent culture is negative; and bacteriological, radiological or clinical evolution is unfavorable (failure/relapse);

  h) There is no culture result from a sputum sample collected between Week 97 and Week 104 or it is positive due to laboratory cross contamination;

     ***AND***
     - there is no other post-baseline culture or it is positive; or,

     - the most recent culture is negative and bacteriological, radiological and clinical evolution is not assessable.

  i) Previously classified as unfavorable in the present study;[iii]
  j) Loss to follow-up.[iv]

---

[iii] Exception: A participant whose outcome is unfavorable because it is unassessable at Week 73 is eligible for re-evaluation at Week 104.
[iv] Loss to follow-up is defined in section Loss to Follow-up5.3.5.

2. Early treatment response:

   a) Proportion of patients with culture conversion assessed in MGIT system (and LJ where possible): 2 consecutive negative cultures from specimens collected at 2 different visits; if there is a missing or contaminated culture between 2 negatives, the definition of conversion is still met;
   b) Time to culture conversion: assessed in MGIT system (and LJ where possible): time from treatment initiation to first of 2 consecutive negative cultures; if there is a missing or contaminated culture between 2 negatives, the definition of conversion is still met;
   c) Change in time to positivity (TTP) in MGIT over 8 weeks.

3. Proportion of participants with favorable outcome at Week 39:

- A participant's outcome will be classified as favorable at Week 39 if all culture results from samples collected between Week 36 and Week 39 are negative and the outcome is not classified as unfavorable.

A participant's outcome will be classified as unfavorable at Week 39 in case of:

   a) In the experimental arm or in the control arm if using the shortened regimen, addition or replacement of one or more drugs;
   b) In the control arm, if using the conventional regimen, addition or replacement of two or more drugs;
   c) Death from any cause;
   d) At least one culture result (from a sample collected between Week 36 and Week 39) is positive;
   e) The patient is not assessable because the last available culture result is from a sample collected before Week 36.

4. At Week 73 and Week 104, proportion of participants with a treatment failure or relapse (defined in section 3.2.1 and 3.2.2).

*Safety:*

1. At Week 73 and Week 104, the proportion of patients who died of any cause;
2. The proportion of participants with grade 3 or greater AEs and serious adverse events (SAEs) of any grade by Week 73 and Week 104;
3. The proportion of patients with AESIs (as defined in section 9.2.5) by Week 73 and Week 104.

##### 3.2.3 Exploratory Outcome Measures

1. Safety and efficacy endpoints as defined in sections 3.2.1 and 3.2.2;
2. Culture conversion endpoints as defined in section 3.2.2 and favorable/unfavorable outcomes as defined in sections 3.2.1 and 3.2.2;
3. Drug resistance amplification: defined as any change in baseline DST, from susceptible to resistant, on identical strain;
4. Adherence is defined as completion of 80% of the prescribed doses (for this purpose, a dose is defined as all the study medications at the prescribed dose for that particular day);
5. Severe linezolid-related toxicity is defined as grade 3 or higher linezolid-related AEs (leukopenia, anemia, thrombocytopenia, peripheral neuropathy, and optic neuropathy), SAEs, and AEs requiring linezolid discontinuation.

### 4 STUDY DESIGN

This is a randomized, controlled, open-label, multi-country Phase III trial evaluating the efficacy of new combination regimens for treatment of MDR-TB. The study will enroll in parallel across 5 experimental and 1 standard-of-care control arms. Randomization will be outcome adapted using Bayesian interim analysis of efficacy endpoints. Trial participation in all arms will last at least until Week 73 and up to Week 104. Study follow-up will end after the scheduled Week 73 for the last participant randomized (to be defined by the Sponsor).

In the experimental arms, treatment will be for 39 weeks and post-treatment follow up for an additional 65 weeks. In the control arm, treatment will be delivered according to local practice (and WHO guidelines); duration will be variable, for approximately 86 weeks for the conventional regimen and 39 to 52 weeks for the standardized shorter regimen.

Experimental arms will be those presented in Table 1:

1. bedaquiline-linezolid-moxifloxacin-pyrazinamide (BeLiMoZ)
2. bedaquiline-clofazimine-linezolid-levofloxacin-pyrazinamide (BeLiCLeZ)
3. bedaquiline-delamanid-linezolid-levofloxacin-pyrazinamide (BeDeLiLeZ)
4. delamanid-clofazimine-linezolid-levofloxacin-pyrazinamide (DeLiCLeZ)
5. delamanid-clofazimine-moxifloxacin-pyrazinamide (DeCMoZ)
6. Control arm will be local standard of care consistent with WHO guidelines. Conventional or shorter standardized treatment regimens may be used, in compliance with WHO recommendations. Conventional treatment regimens may include delamanid for endTB participants who have a higher risk of poor outcomes; it may include bedaquiline or delamanid in endTB participants who have drug resistance patterns or toxicity (e.g., ototoxicity) that preclude development of a likely effective regimen without bedaquiline or delamanid.[154,155] Shortened regimens should be composed as recommended by WHO and used only in patients who meet the WHO recommendations for their use.

#### 4.1 Study Population

The study population will comprise adults and adolescents (≥15 years) with pulmonary tuberculosis suspected or confirmed to be resistant to rifampin in the catchment areas served by IRD, PIH and MSF affiliates in at least 7 countries: enrollment will begin in Georgia, India, Kazakhstan, Lesotho, Pakistan, Peru, and South Africa and may expand to include one or more other sites. These sites have been selected for their MDR-TB burden (Table 2), which we anticipate will permit enrollment to be completed in approximately 30 months.

#### 4.2 Treatment Delivery and Duration

Oral drugs will be delivered 7 days/week during the treatment phase in both experimental and control arms. Treatment with injectable drugs in the control arm will be delivered at least 6 days/week, according to local practices. All doses will be directly observed by designated study staff, except on occasions where provisions are made for treatment support by other, trained individuals. The scheduled duration of experimental regimens is 39 weeks. Control treatment duration will be according to the local standard and consistent with WHO guidance. Control treatment will be administered by study staff until the planned end of treatment date for the study participant or until the end of study follow-up, whichever occurs first. Treatment will continue after completion of the study period according to local protocol and at the discretion of local physicians.

**Table 2 MDR-TB Burden in endTB Sites (2015 and 2016)**

| | MDR-TB patients notified and treated in study catchment areas | |
|---|---|---|
| | **2015** | **2016** |
| **Georgia** | 189 | 177 |
| **India** | Not available | Not available |
| **Kazakhstan** | 315 | 301 |
| **Lesotho** | 203 | 240 |
| **Pakistan** | 211 | 221 |
| **Peru** | 437 | 439 |
| **South Africa** | 118 | 120 |
| **Vietnam** | Not available | Not available |

#### 4.3 Assessment of Study Objectives and Measurement of Endpoints

Primary and secondary outcome measures are enumerated in sections 3.2.1 and 3.2.2.

##### 4.3.1 Organization of Evaluations

Study visits will occur at designated study facilities in each site. Treatment delivery/administration and sputum specimen collection may occur in participants' homes or other locales acceptable to study participants and staff. Other study procedures may occur in these places or through remote visits to reduce risks to study staff and participants during the COVID-19 outbreak. A single designated microbiology laboratory for each study site will perform all study microbiology on sputum specimens collected within the study framework. Routine hematology and blood chemistry will be performed in designated clinical labs for each site. Unscheduled laboratory tests may be performed in other clinical laboratories, as needed for participant safety. Chest X-rays and ECG will be performed in designated facilities and read, for clinical purposes, by study physicians or designated personnel.

### 5 STUDY ENROLLMENT AND WITHDRAWAL

Male or female patients with pulmonary TB who are age 15 and older and who have (risk factors for or) documented rifampin-resistant TB but do not have known fluoroquinolone-resistant TB may be consented and screened. Screening consent for the linked endTB-Q trial (clinicaltrials.gov registry NCT03896685) may be used to permit the use of the screening information for endTB. Prospective participants will be identified in inpatient or outpatient facilities in the study catchment areas. Patients who agree to be evaluated for the study will be referred to study staff for consent and screening. Consent from a legal representative will be additionally requested for patients considered minor per local laws. Study staff will be in regular contact with facility staff to permit prompt referral. Consistent with the epidemiology of drug-resistant TB in some study sites, the number of males identified and randomized may exceed the number of females.

#### 5.1 Subject Inclusion Criteria

A patient will be eligible for randomization if s/he:

1. Has documented pulmonary tuberculosis due to strains of *M. tuberculosis* resistant to rifampin (RIF) and susceptible to fluoroquinolones, diagnosed by validated rapid molecular test;
2. Is ≥ 15 years of age;
3. Is willing to use contraception: pre-menopausal women or women whose last menstrual period was within the preceding year, who have not been sterilized, must agree to use contraception[v] unless their partner has had a vasectomy; men who have not had a vasectomy must agree to use condoms;
4. Provides informed consent for study participation; additionally a legal representative of patients considered minor per local laws should also provide consent;
5. Lives in a dwelling that can be located by study staff and expects to remain in the area for the duration of the study.

#### 5.2 Subject Exclusion Criteria

A patient will not be eligible for randomization if s/he:

1. Has known allergies or hypersensitivity to any of the investigational drugs;
2. Is known to be pregnant or is unwilling or unable to stop breast-feeding an infant;
3. Is unable to comply with treatment or follow-up schedule;
4. Has any condition (social or medical) which, in the opinion of the site principal investigator, would make study participant unsafe;
5. a. Has had exposure (intake of the drug for 30 days or more) in the past five years to bedaquiline, delamanid, linezolid, or clofazimine, or has proven or likely resistance to

[v] Allowed methods are defined in a study SOP.

bedaquiline, delamanid, linezolid, or clofazimine (e.g., household contact of a DR-TB index case who died or experienced treatment failure after treatment containing bedaquiline, delamanid, linezolid, or clofazimine or had resistance to one of the listed drugs); exposure to other anti-TB drugs is not a reason for exclusion.
b. Has received second-line drugs for 15 days or more prior to screening visit date in the current MDR/RR-TB treatment episode. Exceptions include: (1) patients whose treatment has failed according to the WHO definition[156] and who are being considered for a new treatment regimen; (2) patients starting a new treatment regimen after having been "lost to follow-up" according to the WHO definition[154] and, (3) patients in whom treatment failure is suspected (but not confirmed according to WHO definition), who are being considered for a new treatment regimen, and for whom the Clinical Advisory Committee (CAC) consultation establishes eligibility.

6. Has one or more of the following:
   - Hemoglobin ≤ 7.9 g/dL;
   - Uncorrectable electrolytes disorders:
     - Total Calcium < 7.0 mg/dL (1.75 mmol/L);
     - Potassium < 3.0 mEq/L (3.0 mmol/L) or ≥6.0 mEq/L (6.0 mmol/L);
     - Magnesium < 0.9 mEq/L (0.45 mmol/L);
   - Serum creatinine > 3 x ULN;
   - Aspartate Aminotransferase (AST) or Alanine Aminotransferase (ALT) ≥ 3 x ULN;
   - Total bilirubin ≥ 3 x ULN;
   - Unless otherwise specified, Grade 4 result as defined by the MSF Severity Scale on any of the screening laboratory tests.
7. Has cardiac risk factors defined as:
   - An arithmetic average of the two ECGs with highest QTcF intervals of greater than or equal to 450 ms. Retesting to reassess eligibility will be allowed using an unscheduled visit during the screening phase;
   - Evidence of ventricular pre-excitation (e.g., Wolff Parkinson White syndrome);
   - Electrocardiographic evidence of either:
     - Complete left bundle branch block or right bundle branch block; OR
     - Incomplete left bundle branch block or right bundle branch block and QRS complex duration greater than or equal to 120 msec on at least one ECG;
   - Having a pacemaker implant;
   - Congestive heart failure;
   - Evidence of second- or third-degree heart block;
   - Bradycardia as defined by sinus rate less than 50 bpm;
   - Personal or family history of Long QT Syndrome;
   - Personal history of arrhythmic cardiac disease, with the exception of sinus arrhythmia;
   - Personal history of syncope (i.e. cardiac syncope not including syncope due to vasovagal or epileptic causes).

8. Concurrent participation in another trial of any medication used or being studied for TB treatment, as defined in cited documents.[vi,vii]

9. Is taking any medication that is contraindicated with the medicines in the trial regimen which cannot be stopped (with or without replacement) or requires a wash-out period longer than 2 weeks (see section 6.6).

Patients found not to be eligible for endTB due to abnormalities may be reevaluated within the screening window (up to 14 days after screening consent signature date). If any ineligibility persists, or if for any other reasons a patient has not been randomized within the 14-day screening window, then s/he becomes ineligible for randomization. S/he may be rescreened once. Patients who are rescreened should be newly consented and given a new screening number.

All patients who are not eligible for the endTB trial will be managed according to local routine practice or may enroll in other studies. Results of screening for endTB, and for the linked endTB-Q trial, may be used interchangeably. Separate screening for the two trials is not required.

#### 5.3 Treatment Assignment Procedures

The study will use Bayesian, outcome-adaptive randomization to assign treatment. The probability of assignment to each experimental arm will vary according to interim and end-of-treatment outcomes of participants previously assigned in the trial.

##### 5.3.1 Randomization Procedures

*Bayesian adaptive randomization principle:*

The trial will begin with participants being randomly assigned, in equal numbers, to each of the 6 regimens. The Bayesian adaptation will use two preliminary outcomes, Week 8 culture negativity and favorable outcome at Week 39 (described in section 3.2.2), and will estimate the treatment success (favorable outcome) at Week 39 (TS-39) given treatment success (culture conversion) at Week 8 (TS-8) using the relation:

P(TS-39) =
P(TS-39 | positive TS-8)*P(positive TS-8) + P(positive TS-39 | negative TS-8)*P(negative TS-8)

Then the Bayesian model will use observed data to calculate the posterior probability of a positive treatment effect (PTE) for each experimental regimen, defined as:

---

[vi] WHO consolidated guidelines on drug-resistant tuberculosis treatment. Geneva: World Health Organization; 2019. Licence: CC BY-NC-SA 3.0 IGO. Any drug mentioned in Table 2.1 or its footnotes.

[vii] Guidelines for treatment of drug-susceptible tuberculosis and patient care, 2017 update. Geneva: World Health Organization; 2017. Licence: CC BY-NC-SA 3.0 IGO. Any drug mentioned as part of a recommended regimen for drug-susceptible TB.

P (PTE arm a) = P(positive TS-39 in arm a > positive TS-39 in control)

Thus, the probability of randomizing the $i^{th}$ patient to the experimental regimen a will be calculated as follows:

P (patient $i^{th}$ to the regimen a) $= \frac{P(PTE\ arm\ a)}{\sum P(PTE)}$

The probability of assignment to the control arm will match the probability of assignment to the most effective experimental arm.

*Adaptive randomization procedure:*

The randomization list will be generated and updated by a central study staff member (statistician or data manager) who has no direct contact with the trial participants or involvement with the assessment for eligibility in the trial.

Once a participant is eligible for the study and his/her details are entered into the clinical trial database, the investigator on site responsible for the randomization will receive the participant inclusion number and the allocated regimen through a centralized interactive randomization system (IVRS/IWRS).

Adaptation will start once approximately 30 participants have been randomized to each regimen. Subsequent interim analyses to adapt the randomization on Weeks 8 and 39 outcomes will be run approximately every 4 weeks.

Updated randomization lists will then be generated, reflecting the new assignment probabilities. If during the conduct of the study, an experimental regimen is dropped from the study (for any reason described in section 5.3.3), then the list will be updated accordingly.

*Secondary linezolid dose randomization:*

A blocked randomization list will be generated by the Sponsor, using blocks of varying size. Randomization will be stratified by site and balanced 1:1 between the two linezolid dose-reduction strategies.

Participants randomized to the experimental arm will undergo linezolid dose-reduction randomization either at Week 16 or after a linezolid-related AE requiring dose reduction, whichever is earlier.

##### 5.3.2 Masking Procedures

The study is an open label study; the regimens will not be masked. Nevertheless, access to the regimen assignment information will be limited at central and site levels. Procedures will be developed to minimize the bias introduced in endpoint assessment, clinical management, adaptation of randomization, and by review of interim results.

##### 5.3.3 Reasons for Treatment Discontinuation and Withdrawal from Study

*Treatment discontinuation*

**Experimental arm and shortened regimen in the control arm**

Study treatment discontinuation is defined as permanent discontinuation of two or more investigational drugs, or addition or replacement of one or more investigational drugs.

**Control arm, conventional regimen**

Study treatment discontinuation is defined as addition or replacement of two or more drugs. Addition or replacement of two or more drugs to the control arm in order to conform to a new WHO guidance—rather than in response to emerging participant data—might be accepted without qualifying as study treatment discontinuation. This will be assessed on a case-by-case basis by the Clinical Advisory Committee.

**Control arm, conventional & shortened regimens**

Changes within a drug class (i.e., replacing one member of the fluoroquinolone, thioamide, or aminoglycoside/polypeptide class with another member of the same class) do not constitute a replacement.

**For all arms, regimens**

Modification of dose or frequency or temporary suspension of one or more investigational drugs is not considered to be study treatment discontinuation.

The clinical investigator will be discouraged from changing or restarting treatment without evidence of treatment failure or recurrence of MDR-TB. As soon as the clinical investigator has any indication of a treatment failure, recurrence, or serious toxicity, s/he may modify the regimen as required to protect the well-being of the study participant. S/he should also contact the Clinical Advisory Committee (CAC) to review the case and consider other actions. Any permanent treatment change requires advice from the CAC. This applies to all scenarios below as well as other scenarios in which permanent treatment change is considered by an investigator.

Permanent treatment discontinuation guidance due to AEs is described in section 9.4.1. Permanent discontinuation of investigational drug(s) or regimen is also strongly advised if any of the following occurs:

- An enrolled patient becomes pregnant or begins breastfeeding an infant, and has other treatment options with reasonable chance of success;
- The patient requests to stop study treatment.

Permanent discontinuation of investigational drug(s) or regimen may be considered on a case-by-case basis in the following circumstances:

- Requirement for prohibited concomitant medications;
- Any condition (social or medical) which, in the opinion of the site principal investigator, would make study participant unsafe;
- Indications of treatment failure including positive culture at Week 16 or later.

*Withdrawal from the study*

A participant will be withdrawn from the study if:

- Participant's initial strain is determined by confirmatory resistance testing to be resistant to fluoroquinolone (late exclusion);
- Participant withdraws consent.

###### 5.3.4 Handling of Withdrawals and Discontinuations

The Investigator must make every reasonable effort to keep each patient on his/her allocated regimen and in follow-up for the whole duration of the study. Exceptions include participants who permanently discontinue study treatment prior to Week 73 and participants who withdraw. Participants who permanently discontinue study treatment prior to Week 73 will complete only Early Termination and Post-termination follow-up visit(s), as described in sections 7.4 and 7.5, respectively. Any patient withdrawn from the study will be encouraged to complete an Early Termination Visit, as described in section 7.4 and7.5.

These patients will be followed for safety as described in section 9.6.

If consent is withdrawn, data collected prior to withdrawal of consent will be analyzed with complete data sets as is appropriate (e.g., time to sputum negativity for a patient who withdraws after becoming culture negative).

###### 5.3.5 Loss to Follow-up

Participants for whom study staff have no information and whom they have been unable to contact, in-person or by phone for more than 14 weeks before the last study visit per study schedule will be considered lost to follow-up. The "loss to follow-up" designation will be made after the last study visit per study schedule.

###### 5.3.6 Termination of Study

The study may be terminated by the Sponsor, one of the IRBs, or local regulatory authorities. Reasons for termination of one or more arms by Sponsor include the identification of unacceptable toxicities or unequivocal efficacy results. Such a decision may be recommended by the DSMB and would be implemented in consultation with the investigators and the Scientific Advisory Committee (SAC).

### 6 STUDY TREATMENT OF PATIENTS

#### 6.1 Investigational Drugs

The study will use a combination of marketed anti-TB drugs in each of the 6 treatment arms. Each individual anti-TB drug in the experimental regimens is described below; additional information on the anti-mycobacterial activity of each, alone and in combination, is in section 2. The control regimen will use anti-TB drugs according to local practice and consistent with WHO guidance. Summary of product characteristics and/or package inserts for all investigational drugs and WHO interim guidance documents on bedaquiline and delamanid will be made available to investigators.

##### 6.1.1 Bedaquiline (Be)

Bedaquiline fumarate (bedaquiline or Sirturo™) is a diarylquinoline anti-mycobacterial drug. It inhibits ATP synthesis, a novel method of action. The drug has a 5.5-month half-life. CYP3A4 is the major CYP isoenzyme involved in the metabolism of bedaquiline. In December 2012, the United States Food and Drug Administration conditionally approved the use of bedaquiline as part of combination therapy for adults (≥ 18 years) with pulmonary MDR-TB when no other alternatives are available.[157] Each Sirturo™ tablet for oral administration contains 120.89 mg of bedaquiline fumarate drug substance; equivalent to 100 mg bedaquiline per tablet. The recommended dosage for Sirturo™ is 400 mg once daily for 2 weeks followed by 200 mg 3 times per week for 22 weeks with food. In the study, participants who are randomized to receive bedaquiline will be dosed 200 mg once daily for 2 weeks followed by 100 mg 3 times per week for 37 weeks if they weigh between 24 to 30 kg, or 400 mg once daily for 2 weeks followed by 200 mg 3 times per week for 37 weeks if they weigh more than 30 kg.

##### 6.1.2 Delamanid (De)

Delamanid (OPC-67683 or Deltyba™) is an agent derived from the nitrodihydro-imidazooxazole class of compounds and inhibits the synthesis of mycolic acid, which is an exclusive component of the external wall of mycobacteria. Peak plasma levels are achieved 4-5 hours after administration. The half-life of delamanid is 38 hours after drug discontinuation. Delamanid received a conditional marketing authorization by the European Medicines Agency (EMA) in November 2013 for the treatment of adult patients with pulmonary MDR-TB when an effective treatment regimen cannot otherwise be devised for reasons of resistance or tolerability.[158] Each Deltyba™ tablet for oral administration contains 50 mg delamanid. The recommended dosage for delamanid is 100 mg twice daily for 24 weeks. In the study, participants who are randomized to receive delamanid will be dosed 50 mg twice daily (if they weigh between 24 to 30 kg) or 100 mg twice daily (if they weigh more than 30 kg) for 39 weeks.

##### 6.1.3 Clofazimine (C)

Clofazimine (Lamprene®) is a substituted riminophenazine derivative. It acts against mycobacteria by inhibiting mycobacterial replication and growth, and binds preferentially to mycobacterial DNA. In addition to its slow bactericidal effect, clofazimine has anti-inflammatory properties. Clofazimine is indicated as part of combination therapy in treating leprosy. Clofazimine is retained in the human body for a long time. The half-life of clofazimine following repeated doses is estimated to be at least 70 days. The recommended dosage for clofazimine for adults and adolescents (50-70 kg) is 300 mg once a month under supervision plus 50 mg daily as self-medication when used in combination with dapsone and rifampicin. In the study, participants who are randomized to receive clofazimine will be dosed 100 mg once daily for 39 weeks.

##### 6.1.4 Fluoroquinolones

*Levofloxacin (Le):*

Levofloxacin, a synthetic broad-spectrum antibacterial agent, is the levo isomer of ofloxacin. Levofloxacin is rapidly and, essentially, completely absorbed after oral administration. Peak plasma levels are achieved 3 hours after administration. The half-life of levofloxacin ranges from approximately 6 to 8 hours. Elimination occurs mainly via excretion of unmetabolized drug in the urine. The recommended dose for levofloxacin in treating nosocomial pneumonia is 750 mg every 24 hours for a duration of 7 to 14 days. In the study, participants who are randomized to receive levofloxacin will be dosed 750 mg once daily if they weigh between 24 to 45 kg or 1000 mg once daily if they weigh 46 kg or more. The treatment duration will be 39 weeks.

*Moxifloxacin (Mo):*

Moxifloxacin, also a synthetic antibacterial agent, has a half-life of 11.5-15.6 hours that allows it to be given once a day. The drug is rapidly absorbed and achieves high levels in tissues, including the lung. Approximately 52% of the oral dose is metabolized via glucuronide and sulfate conjugation. The cytochrome P450 system is not involved in metabolism. Moxifloxacin is excreted as either unchanged drug or metabolites in urine and in feces. The recommended dose for moxifloxacin is one 400 mg tablets every 24 hours. This dosage will be used for participants who are randomized to receive moxifloxacin in this study. The treatment duration will be 39 weeks.

##### 6.1.5 Linezolid (Li)

Linezolid is a synthetic antibacterial agent member of the oxazolidinone class of antibiotics, which has clinical utility in the treatment of infections caused by aerobic gram-positive bacteria.[70] Linezolid works by inhibiting ribosomal protein synthesis, a mechanism of action different from that of other antibacterial agents; therefore, cross-resistance between linezolid and other classes of antibiotics is unlikely. It is rapidly absorbed (peak serum concentration reached 1-2 hours after administration), highly bioavailable, given orally, and hepatically metabolized without involvement of the cytochrome P450 system. Its average half-life is 5 hours in adults. The recommended dosage for linezolid in treating adults and adolescents (> 12 years of age) with community-acquired or nosocomial pneumonia is 600 mg (IV or oral administration) every 12 hours for 10 to 14 consecutive days. In the study, participants who are randomized to receive linezolid will start at 300 mg daily if they weigh between 24 to 30 kg or at 600 mg daily if they weigh more than 30 kg. They will undergo a second, balanced randomization to establish the dose-reduction strategy between 300 mg daily or 600 mg thrice weekly. The dose will be reduced routinely at Week 16 or sooner if necessary to reduce toxicity related to linezolid. The treatment duration will be 39 weeks.

##### 6.1.6 Pyrazinamide (Z)

Pyrazinamide, an analog of nicotinamide, is a unique anti-TB agent. Although its mechanism of action remains unknown, pyrazinamide has potent sterilizing activity and is used during the initial phase of anti-TB treatment with the goal of shortening therapy. Pyrazinamide is well absorbed from the gastrointestinal tract and attains peak plasma concentrations within 2 hours. Widely distributed in body tissues and fluids, pyrazinamide has a plasma half-life of 9 to 10 hours in patients with normal renal and hepatic function. Dosage recommendations for pyrazinamide vary across countries. Usual dose of pyrazinamide is 15 to 30 mg/kg once daily. Alternatively, pyrazinamide could also be taken twice weekly on a dose of 50 to 75 mg/kg. In the study, all participants who are randomized to the experimental regimens will receive pyrazinamide once daily on a dose of 22 to 35 mg/kg for 39 weeks.

#### 6.2 Trial Interventions

##### 6.2.1 Experimental Treatment

The experimental regimens are endTB 1 (BeLiMoZ), endTB 2 (BeLiCLeZ), endTB 3 (BeDeLiLeZ), endTB 4 (DeLiCLeZ), and endTB 5 (DeCMoZ) (see Table 1). Subjects who are randomized to an experimental arm will receive treatment for 39 weeks and post-treatment follow-up for 65 weeks. Participants may take as long as 47 weeks to complete all doses of a 39-week treatment regimen. Dosing of experimental regimens will be oral and weight based, as described below in Table 3.

**Table 3 Experimental Regimen Dosing by Weight Bands**

| Drug* | Weight Band (kg) | | | | | |
|---|---|---|---|---|---|---|
| | 24-30 | >30-35 | >35-45 | >45-55 | >55-70 | >70 |
| **Bedaquiline (Be)** | 200 mg QD for 2 weeks** followed by 100 mg 3 times/week | 400 mg QD x 2 weeks*** followed by 200 mg 3x/week | | | | |
| **Delamanid (De)** | 50 mg BID | 100 mg BID | | | | |
| **Moxifloxacin (Mo)** | 400 mg | | | | | |
| **Levofloxacin (Le)** | 750 mg | | | 1000 mg | | |
| **Linezolid (Li)******** | 300 mg QD for up to Week 16 (followed by 300 mg QD or 600 mg 3x/week) | 600 mg QD for up to Week 16 (followed by 300 mg QD or 600 mg 3x/week) | | | | |
| **Clofazimine (C)** | 100 mg | | | | | |
| **Pyrazinamide (Z)** | 800 mg | | 1200 mg | 1600 mg | | 2000 mg |

* Dosing is once a day unless otherwise indicated.
** The intensive phase of bedaquiline treatment for the 24-30 kg weight band comprises 2 weeks of 200 mg QD. If the patient is already receiving bedaquiline treatment, bedaquiline within the experimental regimen should be dosed as a continuation of that treatment (i.e., only remaining doses of 2-week intensive phase should be administered and intensive phase should not be restarted if it was already complete at time of study treatment initiation).
*** The intensive phase of bedaquiline treatment for participants weighing more than 30 kg comprises 2 weeks of 400 mg QD. If the patient is already receiving bedaquiline treatment, bedaquiline within the experimental regimen should be dosed as a continuation of that treatment (i.e., only remaining doses of 2-week intensive phase should be administered and intensive phase should not be restarted if it was already complete at time of study treatment initiation).
**** Linezolid dosing will be routinely modified at Week 16 or sooner if necessary to reduce toxicity related to linezolid. The modification will entail either decreased (300 mg daily) or intermittent (600 mg 3x/week) dosing as defined by balanced randomization.

##### 6.2.2 Control Treatment

The control regimen (endTB 6 in Table 1) is the locally used standard of care MDR-TB regimen that is consistent with WHO guidance. endTB-specific guidance for regimen development and dosing is provided in the trial SOP. Subjects who are randomized to the standard-of-care control arm will receive study treatment until the planned end of treatment date for the study participant or until the end of study follow-up, whichever happens earlier. Treatment will continue after completion of the study period according to local protocol and at the discretion of local physicians.

###### 6.2.3 Modification of Study Intervention/Investigational Drug for a Participant

Certain events or conditions may necessitate modification, temporary suspension, or permanent discontinuation of the study regimen. Any time a study regimen is temporarily suspended because of clinical AE or laboratory abnormality, it will be restarted as soon as the participant's condition permits (see section 9.4.1). Participants who experience temporary study regimen suspension remain "on study" and will be followed until study completion.

##### 6.3 Medicines Supplies

The MSF procurement center, MSF Logistique (Bordeaux, France) will acquire all investigational drugs for the trial. If necessary, other procurement mechanisms meeting similar standards may also be used to acquire investigational drugs. Investigational drugs to be procured include:

- Amikacin
- Bedaquiline
- Capreomycin
- Clofazimine
- Cycloserine
- Delamanid
- Ethambutol
- Ethionamide
- Isoniazid
- Kanamycin
- Levofloxacin
- Linezolid
- Moxifloxacin
- PAS
- Prothionamide
- Pyrazinamide
- Terizidone

All investigational drug packages will be labeled with specific endTB clinical trial stickers. They will only be used for the purpose of the trial. Summary of product characteristics and/or package inserts for all investigational drugs, will be provided to the sites through a document entitled Reference Safety Information (RSI). Investigational drug formulation, packaging, labeling and storage requirements from manufacturers are described in the RSI, whereas labelling and storage requirements specific to the trial are detailed in the endTB Pharmacy Manual.

#### 6.4 Accountability Procedures for the Study Intervention/Investigational Drug(s)

The site pharmacist will inventory and acknowledge receipt of all shipments of the investigational drugs. S/he will keep accurate records of the quantities of the investigational drugs dispensed and used by each subject. At the conclusion of the study, documentation of medications received will be stored with the study documents.

#### 6.5 Assessment of Subject Compliance with Study Intervention/ Investigational Drug

Adherence to investigational drugs will be assessed by regular review of each participant's treatment documentation and unused investigational drug supply.

#### 6.6 Concomitant Medications/Treatments

The following medication categories are disallowed, for each specific anti-tuberculosis drug, during the treatment and up to 1 month after the end of treatment:

<u>Bedaquiline</u>

1. Moderate and strong CYP3A4 inhibitors.

2. Moderate and strong CYP3A4 inducers.

<u>Delamanid</u>

1. Strong CYP3A4 inducers.

<u>Linezolid</u>

1. Any medicinal product that inhibits monoamine oxidases A or B.
2. Tricyclic antidepressants.
3. Selective serotonin reuptake inhibitors.
4. Selective serotonin/norepinephrine reuptake inhibitor.
5. Triptans.
6. Other serotoninergic agents.

Further information is available in the corresponding standard operating procedure (SOP).
For concomitant medications that have the potential to prolong the QT interval or are associated with torsades de pointes, clinical investigators will be instructed in the SOP to avoid the drug in regimens with other QT prolonging drug(s) and if not possible to use with caution.

Clinical investigators will be instructed in the SOP on other concomitant medications that should only be used with particular caution with investigational drugs.

There are no additional absolute contraindications on medications delivered concomitantly with clofazimine, pyrazinamide, moxifloxacin, levofloxacin, kanamycin, amikacin, capreomycin, ethionamide, prothionamide, cycloserine, isoniazid and PAS.

### 7 STUDY SCHEDULE

Procedures and evaluations are fully detailed in section 8. A full visit schedule/schedule of events is provided in Appendix 1. Participants should complete evaluations according to the visit schedule. Visit windows are as follows:

- Randomization should occur no more than 14 days after screening consent signature date;
- Treatment should be initiated as quickly as possible. The only exceptions to this are cases where the washout of concomitant medications requires additional time, up to a maximum of 5 days. The procedure for these cases will be elaborated in SOPs;
- +/- 2 days for Weeks 1 to 12 of the study;
- +/- 7 days for Weeks 16 to 36 of the study;
- +/- 14 days for Weeks 39 to 65 and Weeks 81 to 97 of the study;
- +/-30 days for Weeks 73 and 104 of the study.

In the protocol, study week numbers refer to the named week and its respective window, e.g., "Week 39" includes the 14-day periods before and after week 39.

Week numbers are calculated based on the day of randomization, assuming that study regimen starts on that day. If treatment initiation is delayed, the Week 8 visit window may be revised to be 8 weeks (+/-2 days) after study treatment initiation.

During the COVID-19 outbreak, efforts should be made to safely conduct all study visits and procedures. When this is not possible, priority should be placed on visits and procedures required to protect participant safety, ensure completion of treatment, and assessment of study outcome. Certain procedures require close physical contact. When such contact is not feasible, replacement by remote assessments (e.g., BPNS replaced by subjective peripheral neuropathy assessment), is preferable to not performing the procedure.

#### 7.1 Screening

##### 7.1.1 Pre-screening

As part of the routine care in country, some or all of the following procedures may be completed for routine care and information may be available for review:

- Basic demographics;
- Sputum smear microscopy;
- Sputum culture;
- Phenotypic DST results;
- Rapid molecular testing for resistance to at least RIF.

Results of the above procedures may be used to identify potentially eligible patients to whom participation in the screening process can be proposed.

##### 7.1.2 Screening

Study information will be provided to patients and screening consent obtained from patients who agree to be screened for the study. Screening consent will be obtained prior to any trial-specific evaluation.

The following procedures will be completed after the participant has provided screening consent:

- Medical history;
- Inclusion and exclusion criteria, as listed in sections 5.1 and 5.2, respectively, will be assessed;
- Physical examination (including vital signs);
- Concomitant medicine evaluation (consideration for wash-out period and/or replacement medicine if needed) including anti-TB treatment;
- Pregnancy test (urine or serum) for women of childbearing age;
- Complete blood count (CBC);
- ALT and AST, total and direct bilirubin, albumin, creatinine, electrolyte testing;
- Lab tests: TSH and HbA1c (or 2 fasting blood glucose tests if HbA1c cannot be done); – optionally, documented results of TSH (performed less than 4 weeks prior to screening visit date) and HbA1c (performed less than 3 months prior to screening visit date) may substitute for screening tests. Investigators are encouraged to repeat TSH and/or HbA1c if they deem it useful for patient management or to have a reliable pre-treatment result;
- Hepatitis serology will be offered (seropositivity is not a contraindication for study participation), unless a documented result is available from less than one month before screening visit date;
- HIV serology will be offered, unless patient is known to be HIV positive or a documented negative result is available from less than one month before screening visit date;
- CD4 count and HIV viral load will be performed if the patient is known or found to be HIV positive. Optionally, documented results of CD4 count (performed less than six months prior to screening visit date) and HIV viral load (performed less than 4 weeks prior to screening visit date) may substitute for screening tests. Investigators are encouraged to repeat CD4 count and HIV viral load if they deem it useful for patient management or to have a reliable pre-treatment result.
- ECG;
- Collection of 3 sputum specimens to perform[viii]:
    - Validated rapid molecular test for (at least) RIF resistance (one RIF-resistant result from the designated study lab is required prior to randomization);
    - Validated rapid molecular test for fluoroquinolone resistance (one result showing

[viii] All study participants require the following performed by a designated study lab: validated molecular tests indicating RIF resistance and FQ susceptibility, as well as smear, culture, and phenotypic DST. Smear, culture, and phenotypic DST must be performed by designated study lab on screening or baseline samples. Molecular tests do not need to be repeated at screening or baseline if they were performed by a designated study lab on pre-screening samples collected no more than 3 weeks prior to screening visit.

FQ susceptibility from the designated study lab is required prior to randomization);
    - Smear microscopy;
    - Culture on MGIT (and LJ, where possible);
    - Phenotypic drug-susceptibility testing (DST) will be performed and genotyping, genomic sequencing (see section 8.2.2) may be performed on collected specimens and isolated strains.
- Strain storage for future research related to TB (if optional consent is granted).

#### 7.2 Baseline

Full study information will be provided to patients and baseline informed consent obtained from patients who agree to participate in the study. Baseline informed consent will be obtained prior to any of the following procedures.

The following procedures will be completed during the baseline visit (Visit 2):

- Review eligibility criteria;
- Demographics;
- Interval Medical history;
- Physical exam (including vital signs);
- TB symptom assessment;
- ECOG assessment;
- Neurologic exam;
- Ophthalmologic exam;
- Mental health assessment;
- Concomitant medicine review: this includes ART for HIV co-infected patients. All HIV co-infected participants will be eligible for ART. ART regimens will be adjusted as necessary to facilitate study inclusion;
- Lab tests: TSH and HbA1c (or 2 fasting blood glucose tests if HbA1c cannot be done), if not completed at screening;
- Hepatitis serology (if not completed at screening);
- CD4 count and HIV viral load, if patient is HIV positive (if not completed at screening);
- Serum pregnancy test (women of childbearing age). Blood specimen leading to negative pregnancy test result should be drawn within 72 hours prior to treatment initiation (test can be repeated if needed);
- If complete screening microbiological and molecular TB testing results[viii] have not been obtained, 2 sputum specimens should be collected and tested prior to randomization:
- Audiometry;
- ECG;
- Chest X-ray – not necessary if results judged adequate are available from imaging performed less than 3 weeks prior to baseline visit date;
- Adherence counseling;
- Optional consent for strain storage for future research related to TB.

Once the results of all baseline procedures necessary for eligibility assessment are available, and eligibility is confirmed, patient will be randomized through the IVRS/IWRS system according to procedures in Randomization Procedures. Treatment should be initiated as quickly as possible.

Baseline procedures that are not necessary for eligibility assessment can be run in parallel to randomization and study regimen initiation.

#### 7.3 Follow-up

Additional safety follow-up (after last study visit) may be indicated as described in section 9.6.

*Weeks 1 to 47 (Visits 3-23):*

Visits will occur as shown in the schedule of event in Appendix 1. All visits during this interval will include the following:

- Interval medical history;
- Physical examinations (including vital signs);
- TB symptom assessment;
- Adherence counseling;
- Treatment compliance review;
- Concomitant medicine review;
- AE symptom review;
- ECG.

Additionally, at Week 4, 8, 12, 16, 20, 24, 28, 32, 36, 39, 43, and 47, visits will include:

- Neurologic exam;
- Ophthalmologic exam;
- CBC;
- AST and ALT, total and direct bilirubin, creatinine;
- Total calcium, potassium, magnesium;
- Audiometry.

And, at Week 2, 4, 8, 12, 16, 20, 24, 28, 32, 36, 39, 43, and 47, two sputum specimens will be collected for:

- Sputum smear microscopy;
- Culture on MGIT (and LJ, where possible).

In addition, the following procedures will be completed at the specified visits:

- ECOG assessment: Week 39;
- TSH: Week 24 and 47;
- HbA1c (if abnormal at screening/baseline): Week 24 and 47;
- CD4 and viral load (if HIV positive at screening or after): Week 24 and 47;
- Pregnancy test (women of childbearing age, urine or serum) if needed;
- If culture positive at or after Week 16: Phenotypic DST, validated rapid molecular test

for fluoroquinolone, and genotyping or genomic sequencing for discrimination between relapse and reinfection;

- Chest X-ray: Week 8 and 39;
- For participants on the linezolid-containing experimental arms (endTB regimens 1-4), randomization of linezolid dose reduction at Week 16, or earlier if indicated.

*After Week 47:*

Visits will occur at Week 53, 59, 65, 73, 81, 89, 97, and 104 (study visits W81-W104 might not happen if end of study follow-up has already occurred).

All visits will include:

- Interval medical history;
- Physical examination (including vital signs);
- TB symptom assessment;
- Concomitant medicine review;
- Adherence counseling and treatment compliance review for patients still on treatment;
- AE symptom review;
- Collection of 2 sputum specimens for:
    - Sputum smear microscopy;
    - Culture on MGIT (and LJ, where possible).
    - If culture positive: phenotypic DST, validated rapid molecular test for fluoroquinolone, and genotyping or genomic sequencing for discrimination between relapse and re-infection;

At Week 73 and 104, visit will include:

- ECOG assessment;
- Neurologic exam;
- Ophthalmologic exam;
- Mental health assessment;
- Audiometry;
- Chest X-ray.

At Week 73 and Week 104, at least one interpretable (positive or negative) culture result is required. If not available, a repeat sputum sample should be collected as soon as possible and cultured on MGIT (and LJ, where possible).

In addition, the following procedures will be completed at the specified visits:

- CBC: Week 104;
- HbA1c (if abnormal at screening/baseline): Week 73;
- CD4 and viral load (if HIV positive at screening or after): Week 73;
- Pregnancy test (women of childbearing age, urine or serum) if needed;
- ECG: Week 53 and 73.

#### 7.4 Early Termination Visit

Any patient withdrawn from the study before the last study visit per study schedule or who permanently discontinues study treatment is encouraged to complete an Early Termination Visit. The following procedures will be completed at early termination visits, if possible:

- Interval medical history;
- Physical examination (including vital signs);
- TB symptom assessment;
- ECOG assessment;
- Neurologic exam;
- Ophthalmologic exam;
- Mental health assessment;
- Concomitant medicine review;
- AE symptom review;
- CBC;
- AST and ALT, total and direct bilirubin, creatinine, TSH,
- Total calcium, potassium, magnesium;
- Pregnancy test (urine or serum) for women of child-bearing age;
- Audiometry;
- ECG;
- Chest X-ray, according to clinical judgement, and in any case if no adequate results are available from imaging performed less than 3 weeks prior to the early termination visit date;
- Based on previous results available, HbA1c or fasting blood glucose (if abnormal at screening/baseline) and CD4 and viral load (if patient is HIV positive) tests may be indicated;
- Collect two sputum specimens for:
  - Sputum smear microscopy;
  - Culture on MGIT (and LJ, where possible);
  - If culture positive: phenotypic DST and genotyping or genomic sequencing (for discrimination between relapse and re-infection).

#### 7.5 Post-termination follow-up visit(s)

Any participant who permanently discontinues study treatment prior to Week 73 5.3.4will complete post-termination follow-up visit(s):

- If the patient permanently discontinues treatment prior to Week 39, (s)he will attend the post-termination follow-up visits at Week 39 and Week 73.

- If the patient permanently discontinues treatment prior to Week 73, (s)he will attend the post-termination follow-up visit at Week 73.

If the patient is withdrawn or permanently discontinues treatment between Week 73 and Week 104, the early termination visit is the last study visit.

The following procedures will be completed at the post-termination follow-up visit(s), if possible:

- Interval medical history;
- Physical examination (including vital signs);
- TB symptom assessment;
- ECOG assessment;
- Concomitant medicine review;
- AE symptom review;
- Chest X-ray, according to clinical judgment, and in any case if no adequate results are available from imaging performed less than 3 weeks prior to the post-termination follow-up visit date;
- Based on previous results available: HbA1c or fasting blood glucose (if abnormal at screening/baseline) and CD4 and viral load (if patient is HIV positive) tests may be indicated.
- Collect two sputum specimens for:
    - Sputum smear microscopy;
    - Culture on MGIT (and LJ, where possible);
    - If culture positive: phenotypic DST and genotyping or genomic sequencing (for discrimination between relapse and re-infection).

### 8 STUDY PROCEDURES/EVALUATIONS

#### 8.1 Clinical Evaluations

*Medical history*:

Medical history will be obtained from patient interview with medical staff at initial screening. The medical history must include all relevant diagnoses occurring within the past 30 days before screening visit date. In addition, the following diagnoses should be recorded if they have occurred at any time during the patient's history: TB, HIV, Hepatitis B or C, arrhythmia, or diabetes mellitus. Any allergies to medications must also be documented. A list of current medications must also be elicited during this medical history, including any medications taken in the last 30 days before screening visit date. Patients will be asked to provide a medication history for any TB or anti-retroviral therapy (ART) ever taken. Patients will also be asked about their consumption of alcoholic beverages and/or recreational drugs.

*Interval medical history:*

Patients will be asked about any changes in their medical history since the last study visit.

*Physical examination:*

The physical examination at baseline visit will comprise vital signs (blood pressure, respiratory rate, temperature, peripheral pulse, oxygen saturation if patient is dyspneic), height (done at baseline visit; may be repeated in 15-17 year olds), weight and review of organ systems, as described in the SOP. This exam at all follow-up visits will also be driven by any previously identified or new signs or symptoms. Evidence of AEs will be obtained by interview and examination.

*TB symptom assessment*:

Patients will be asked about the following: fever, weight loss, cough, and hemoptysis.

*ECOG performance status assessment*:

Standard scale will be used for assessment.

*Mental health assessment:*

Standard scale(s) will be used to detect anxiety and depressive disorders.

*Neurologic examination*:

Neurologic examination will include detection of peripheral neuropathies.

*Ophthalmologic examination*:

Visual acuity and color perception assessment will be performed.

*Concomitant medicine review***:**

Patients will be asked whether they have taken any concomitant medication since the last study visit.

*Adherence counseling:*

At each visit, patients will be counseled about the importance of taking their medications, in spite of discomforts, and the dangers of developing further resistance if they fail to do so. Patients will be informed by study clinicians and nurses of possible AEs.

*Adverse events review:*

Patients will be asked whether they have experienced any adverse event since the last study visit.

*Treatment compliance*:

Taken and missed doses will be recorded in real time.

#### 8.2 Laboratory Evaluations

##### 8.2.1 Blood and Urine Tests

These laboratory tests will be performed at visits as described in section 7. Results from prior tests may be substituted as stipulated in sections 7.1 and 7.2.

*Complete Blood Count:*

Hematology will include a complete blood count with differential, including white blood cell count, hemoglobin, hematocrit, and platelet count.

*Blood Chemistry:*

Blood chemistry tests include the following:

- Electrolytes include magnesium and potassium and total calcium;
- HbA1c (2 fasting glucose if HbA1c cannot be done);
- Renal function tests: creatinine; a creatinine clearance will be calculated using Cockcroft-Gault formula;
- ALT, AST, total and direct bilirubin;
- Albumin and

- TSH.

*HIV and related tests:*

Testing methodologies will conform to national guidelines, and to relevant standards for high- or low-HIV prevalence settings. Refusal to be HIV tested will not preclude study participation. HIV seronegativity is not a criterion for inclusion.

CD4 and HIV viral load will be assessed for patients known or found to be HIV infected.

*Hepatitis B and C test:*

Hepatitis serologies should include tests for: Hepatitis B (Hepatitis B Core Antibody [anti-HBc total] and Hepatitis B Surface Antigen [HBsAg]) and hepatitis C (Hepatitis C Antibody [HCVAb]). Refusal to be tested for hepatitis serology will not preclude study participation.

*Pregnancy test:*

For women of childbearing age, β-HCG (beta human chorionic gonadotropin) will be assessed in a validated serum test for baseline, and in a validated serum or urine test with a sensitivity of at least 20 mIU/mL for all other timepoints.

##### 8.2.2 Tuberculosis Tests

The mycobacterial procedures will be detailed in the endTB analytical plan and relevant SOPs.

*Smear-microscopy:*

Three sputum specimens will be collected (preferably, at least one at home in early morning and at least one spontaneously expectorated at the health facility) for smear microscopy at screening and two sputum specimens will be collected during specified follow-up visits. Ziehl-Neelsen or auramine staining methods will be performed at designated study lab. Laboratories should participate in quality control programs with reference laboratories.

*Mycobacterium tuberculosis culture:*

Culture will be performed in liquid medium (BACTEC960 also known as MGIT), and if possible, in solid LJ medium. Tests will be performed at designated study lab, either local reference (regional or national) or established research laboratory.

*Drug susceptibility testing (DST):*

DST to antituberculosis drugs (preferably at least: isoniazid, rifampicin, ethambutol, pyrazinamide, amikacin, capreomycin, kanamycin, fluoroquinolone(s)) will be performed locally according to the site laboratory SOP and international standards.

When indicated, DST to new drugs such as linezolid, clofazimine, bedaquiline, and delamanid will be done at the supranational TB laboratory at the Tropical Institute of Medicine (ITM) in Antwerp (Belgium) after shipment of the strain. If local, quality-assured laboratories develop validated capacity to perform tests in standardized fashion, local testing to these drugs may also be performed.

ITM may also perform phenotypic DST to other antituberculosis drugs. ITM may perform or outsource genomic sequencing to identify resistance-conferring mutations and/or heteroresistance. MIC testing may be performed either on solid or liquid medium according to procedures validated in ITM for isolates shipped to ITM.

*Validated rapid molecular test for (at least) RIF resistance:*

The endTB analytical plan specifies the type(s) of validated rapid molecular tests and procedures to be used for this trial.

*Validated rapid molecular test for FQ resistance:*

The endTB analytical plan specifies the type(s) of validated rapid molecular tests and procedures to be used for this trial.

*Genotyping*

Genotyping (or sequencing) of *M. tuberculosis* will be performed to distinguish reinfection from relapse in participants whose sputum cultures are positive at any time after Week 16. Genotyping (or sequencing) will be performed according to SOPs at the endTB reference lab, ITM.

###### 8.2.3 Special Assays or Procedures

*Audiometry:*

Pure tone air conduction audiometry will be conducted in a standardized fashion according to validated procedures.

*Electrocardiogram:*

Standard 12-lead ECGs will be performed at specified time points during the study (in triplicate at screening and baseline; and in duplicate [at least] at follow up visits, at any early termination visit, and at unscheduled visits at which potential cardiac toxicity is evaluated) by a site investigator or designated personnel according to the SOP. The delegated site personnel will read each ECG in real time and will record the results. All measurements will be corrected using the Fridericia formula (QTcF = QT / 3√RR). ECG tracings will be kept with the patient's medical records as source document. Initial clinical decision-making will rely on the mean of the locally calculated QTcF intervals, according to the algorithm specified in the SOP; follow up medical decision-making will be supported by central reading.

*Chest X-ray:*

Full-size frontal (posteroanterior) chest X-ray will be performed on site. A specific itemized chest X-ray evaluation sheet will be used to assess the findings and quality of the X-ray.

#### 8.3 Specimen Preparation, Handling, Shipping, and Storage

Specimens and strains will be prepared, handled, shipped and stored according to procedures elaborated in the endTB analytical plan. Specimen and strain storage will be delegated by the Sponsor to either local study labs or to the Institute of Tropical Medicine (ITM) in Antwerp. Specimens and strains will be stored for up to 5 years after the publication of the result of the trial to permit any additional trial-related analyses. Future storage and use of strains is detailed in section 14.5.

### 9 ASSESSMENT OF SAFETY AND MANAGEMENT OF ADVERSE EVENTS

#### 9.1 Specification of Safety Parameters/Endpoints

Safety endpoints are listed in sections 3.1.2 and 3.1.3.

#### 9.2 Adverse Events

##### 9.2.1 Definition of Adverse Events

An AE is any untoward medical occurrence in a patient or clinical investigation subject after administration of an investigational drug. It does not necessarily have a causal relationship with the study treatment. An AE can therefore be any unfavorable and unintended sign (including an abnormal laboratory finding), symptom, or disease temporally associated with the use of a medicinal (investigational) product, whether or not related to the medicinal (investigational) product. If there is any doubt as to whether a clinical observation is an AE, the event should be reported.

An AE is any medical event reported by the patient even if the event occurred between two study visits and is resolved when reported by the patient. Any medical condition that is present at the time that the patient is screened is considered as baseline and is not reportable as an AE. However, if a pre-existing condition deteriorates at any time during the study (e.g. increase of severity), it should be recorded as an AE.

An adverse reaction refers to all untoward and unintended responses to an investigational medicinal product related to any dose administered. Response in this context means that a causal relationship between the event and the investigational drug is at least a reasonable possibility.

An AE or suspected adverse reaction is considered “unexpected” if it is not consistent in nature, severity and/or outcome with the current safety knowledge on the product(s) as detailed in the Reference Safety Information, supplied by the Sponsor.

##### 9.2.2 Assessment of AEs

Study participants will be monitored and assessed clinically for new AEs at all scheduled study visits (see schedule of events in Appendix 1). At each visit the clinical investigator will also systematically assess the evolution and outcome of previously recorded AEs.

Daily visits for supervised treatment will provide additional opportunities for symptomatic screening and referral for unscheduled visits for potential AEs. Laboratory screening for hematologic and biochemical abnormalities and electrocardiogram for monitoring of the QT length will be conducted at specific study visits during treatment and after treatment completion.

Severity of all AEs will be graded and relationship to study product will be evaluated.

##### 9.2.3 Severity of AEs

All AEs will be assessed for severity by the study clinicians. A standardized MSF Severity Grading Scale has been developed mainly following the Division of Microbiology and Infectious Diseases (DMID) adult toxicity tables (November 2007) and the Common Terminology Criteria for Adverse Events (CTCAE) (June 2010). Each AE will be assigned a grade 1 to 4.

For events not included in the protocol-defined grading systems, the following general definitions from grades 1 to 4 will be applied to classify event severity:

- **GRADE 1 Mild** Transient or mild discomfort (<48 hours); no medical intervention/therapy required
- **GRADE 2 Moderate** Mild to moderate limitation in activity - some assistance may be needed; no or minimal medical intervention/therapy required
- **GRADE 3 Severe** Marked limitation in activity, some assistance usually required; medical intervention/therapy required, hospitalizations possible
- **GRADE 4 Life-threatening** Extreme limitation in activity, significant assistance required; significant medical intervention/therapy required, hospitalization or hospice care probable

Changes in the severity of an AE should be documented to allow an assessment of the duration of the event at each level of intensity. AEs characterized as intermittent require documentation of onset and duration of each episode.

##### 9.2.4 Relationship to Study Products

The assessment of an AE's relationship to the regimen drugs will be the responsibility of the site clinical investigator. The clinical investigator will assess the relationship to regimen drugs for all AEs using the terms: related or not related. To help assess, the following guidance for relatedness is used:

- Related: there is a reasonable possibility that the AE may be related to the drug(s);
- Not related: there is **no** reasonable possibility that the AE is related to the drug(s).

##### 9.2.5 Adverse Events of Special Interest (AESI)

An AESI is a medical concern specific to the investigational medicinal product(s) for which close monitoring is required. In this study, the following AEs, regardless of their seriousness or causal relationship to treatment, are considered of interest:

- Grade 3 or above “electrocardiogram QT corrected interval prolonged”;
- Grade 3 or above leukopenia, anemia or thrombocytopenia;
- Grade 3 or above peripheral neuropathy;
- Grade 3 or above optic neuritis;

- Grade 3 or above increase in alanine aminotransferase (ALT) or aspartate aminotransferase (AST).

#### 9.3 Serious Adverse Events (SAEs)

An SAE is defined as an AE meeting at least one of the following conditions:

- An event leading to death.
- A life-threatening event (defined as a subject at immediate risk of death at the time of the event; it does not refer to an event which hypothetically might have caused death if it were more severe).
- An event requiring hospitalization or prolongation of existing hospitalization. The following will not meet the definition of SAE: hospitalization for elective surgery planned prior to subject enrollment; admission to remove barriers to care in the ambulatory environment; admission to perform ECG per protocol; hospitalization for infection control or smear positivity; or visit to the hospital (e.g. emergency room) that lasted less than 24 hours and did not result in admission.
- An event resulting in a persistent or significant incapacity or substantial disruption of the ability to conduct normal life functions.
- An event resulting in congenital anomaly or birth defect.
- Any other important medical event that may not result in death, be immediately life-threatening, or require hospitalization may be considered a serious adverse experience when, based upon appropriate medical judgment, the event may jeopardize the subject and may require medical or surgical intervention to prevent one of the outcomes listed above. Examples of such medical events include allergic bronchospasm requiring intensive treatment in an emergency room or at home, blood dyscrasias or convulsions that do not result in inpatient hospitalization, or the development of drug dependency or drug abuse.

Suspected transmission of an infectious agent (pathogenic or nonpathogenic) via the investigational drug(s) should always be considered an SAE. All deaths, regardless of the cause (e.g. disease progression), are considered as SAEs. If the cause of the death is not immediately assessable or is not known (e.g., investigator learns of participant death from the participant's family), the death is still a reportable SAE. All reasonable efforts should be made to clarify the cause of the death. New or worsening TB related signs and symptoms that meet SAE criteria are recorded and reported as SAEs.

A Grade 4 AE or laboratory abnormality according to the severity scale does not necessarily indicate an SAE, unless the occurrence meets one or more of the above definitions.

#### 9.4 Procedures to be Followed in the Event of Abnormal Laboratory Test Values, Abnormal Clinical Findings or Incidental Events

Treatment must be discontinued for safety reasons if any clinical AE, laboratory abnormality, intercurrent illness, or other medical condition or situation occurs such that continued exposure to treatment would not be in the best interest of the subject.

##### 9.4.1 Management of Adverse Events

Additional guidance for the management of adverse events will be provided in an SOP. Adverse events will be managed as described below except if stated differently in the SOP.

Severity Grade 1:

Participants who develop a Grade 1 AE will generally continue investigational drugs without alteration of the dosage and will be managed at the discretion of the site clinical investigator. A grade 1 AE may require closer monitoring with unscheduled visits and additional laboratory tests.

Severity Grade 2:

Participants who develop a Grade 2 AE will generally continue investigational drugs without alteration of the dosage and will be managed at the discretion of the site clinical investigator. A grade 2 AE may require closer monitoring with unscheduled visits and additional laboratory tests.

Severity Grade 3:

If the AE is considered by the site clinical investigator as not related to the investigational drugs, dosing may continue and the AE may be managed at the discretion of the site clinical investigator with closer monitoring.

Grade 3 AE defined by the clinical investigator as related to investigational drugs or of unknown etiology may result in modification or withholding of investigational drugs. The participant will be managed at the discretion of the site clinical investigator and reevaluated approximately weekly until improvement, resolution, or stabilization of the AE, as determined by the site clinical investigator.

Any Grade 3 AE considered as related to the investigational drugs that does not resolve within 14 days may result in permanent modification or discontinuation of one or more investigational drugs.

Severity Grade 4:

If the AE is considered by the site clinical investigator as not related to the investigational drugs, participant may continue study treatment only if the site clinical investigator considers that the potential benefit of study treatment outweighs the risk, otherwise participants will have all investigational drugs withheld. The participant will be managed at the discretion of the site investigator and reevaluated approximately weekly until improvement, resolution, or stabilization of the AE, as determined by the site clinical investigator. If the AE resolves within 14 days the study treatment may be restarted.

If the AE is considered by the site clinical investigator as related to the investigational drugs or of unknown etiology the participants will be permanently discontinued from one or more investigational drugs.

##### 9.4.2 Overdose

An overdose is defined as the administration of a quantity of a medicinal product given per administration or cumulatively which is above the maximum recommended dose according to the authorized product information or other in-use references (trial protocol or WHO guidelines). The determination of an overdose will be left to the discretion of the clinical investigator, based on the total dose administered, the emergence of any clinical signs and symptoms suggestive of a toxic administration, as well as his own clinical judgment as it applies to each individual case.

The patient may be hospitalized for observation and appropriate supportive treatment administered. The medical consequences of overdose should be managed as described in the section 9.4.1 above.

##### 9.4.3 Pregnancy

Pregnancy must be avoided during study participation due to the identified, potential or unknown risks for the offspring caused by investigational drug(s) exposure. To be eligible, women and male partners of women of childbearing age must be willing to use contraception. If despite all precautions, a patient (or his partner) is found to be pregnant while being treated with the investigational drugs or during the safety follow-up period, pregnancy should be notified to the pharmacovigilance (PV) unit and followed-up until an outcome is known (see section 9.5.7). Of note, only pregnancies considered by the clinical investigator to be related to the investigational medicinal products (i.e. resulting from a drug-drug interaction with the contraceptives) are considered as AEs. Permanent discontinuation of the study treatment is strongly advised immediately upon knowledge of pregnancy in a female participant. Participants in whom study treatment is discontinued for pregnancy will be referred to receive the local, standard of MDR-TB treatment for pregnant women.

#### 9.5 Reporting Procedures

##### 9.5.1 Adverse Event Reporting

AE recording and reporting in all patients regardless of the study treatment arm starts with the initiation of study treatment.

All AEs including local and systemic reactions will be immediately recorded by the clinical investigator (or his/her designee).

The duration of the safety follow-up is described in section 9.6.

Event description, time of onset, clinician's assessment of severity, relationship to study product assessed by clinical investigator, and time of resolution/stabilization of the event will be recorded and data will be entered in the clinical database. Each AE will be coded to a Preferred Term (PT) and primary associated System-Organ Class (SOC) according to the Medical Dictionary for Regulatory Activities (MedDRA) dictionary. All AEs occurring while on study will be documented appropriately regardless of relationship and all AEs will be followed to adequate resolution even if this extends beyond the study-reporting period (see section 9.6).

##### 9.5.2 Serious Adverse Events

The exact same process as for AEs applies (see section 9.5.1). In addition, SAEs are immediately notified, **within 24 hours of SAE awareness,** by the site principal investigator (or designee) to the Sponsor as described below (section 9.5.3) using an SAE report form (supplied by the Sponsor). In case of death, all efforts will be made to identify the cause of patient's death and collect the necessary level of information allowing assessment of the causal relationship with the investigational treatment. Causality assessment will be established for all SAEs taking into account the relationship's plausibility, temporal relationship, consistency, prior knowledge on the drugs, specificity, etc. This takes into account individual patient and cumulative safety data (including from external sources such as publications and from the PV unit database).

##### 9.5.3 Timelines, Recipients and Notification period

***Immediate notification of SAEs to Sponsor***

The site principal investigator will notify immediately (within 24 hours of awareness at the site) all SAEs to the Sponsor pharmacovigilance (PV) unit using the following email address:

***Expedited reporting of Unexpected Serious Adverse Reactions to National Regulatory Authorities (NRAs) and local Institutional Review Boards (IRBs)***

All SAEs deemed related to one or more investigational medicinal product(s) and considered unexpected with the use of such products are reportable to NRAs and IRBs:

- Within 7 calendar days after first knowledge by the Sponsor for fatal and life-threatening Unexpected Adverse Reactions,
- Within 15 calendar days after first knowledge by the Sponsor for all other Serious Unexpected Adverse Reactions.

The PV unit is responsible for reporting individual cases to NRAs and to the principal investigators, who must report the same to the local IRBs under their responsibilities.

All other SAEs are reported in an Annual Safety Report prepared by the PV unit unless there are specific local regulations from IRBs or NRA for a more frequent or prompt reporting.

##### 9.5.4 Notification Procedure to the Sponsor

*Initial notification:*

The initial SAE notification must include at least the following information: the patient trial identification number, the description of SAE, the trial product, and the name and signature of investigator who notifies the SAE. All SAEs will be reported on an SAE report form, dated and signed, with due care being paid to the severity grading and relationship to the trial product.

The investigator should assess, if possible, the diagnosis of all SAE. Diagnosis, or if not available, syndrome should be reported whenever possible. When several symptoms are linked to one pathology, they will be reported using the same SAE report form.

*Complementary (follow-up) notification:*

The resolution or worsening of the event will be notified following the same procedures described for the initial notification. Any change in the initial outcome (e.g. resolved, back to previous status, worsening) and any relevant complementary information on the case (e.g. final diagnosis, test results) should be reported. A new SAE occurring after complete resolution should generally be declared as a separate SAE. Follow-up of SAE should continue also after completion of study visits (see section 9.6).

All documents and information transmitted to the PV unit must be anonymized.

##### 9.5.5 Adverse Events of Special Interest (AESI)

AESIs, regardless of their causal relationship with study treatment, will also be recorded in the CRF and reported to the PV unit using the report forms and time frame described for SAEs.

Only AESIs that fulfill the criteria for expedited reporting, i.e., serious, unexpected, and related to investigational medicinal product(s), will be reported to IRBs and NRAs as detailed in section 9.5.4.

##### 9.5.6 Overdose

Any identified overdose must be recorded in the CRF together with its medical consequences (if any) and must be reported by the site principal investigator (or designee) to the PV unit in an expedited manner (within 24 hours of awareness) using an SAE report form (supplied by the Sponsor).

Only overdoses associated with AEs that fulfill the criteria for expedited reporting, i.e. serious, unexpected, and related to investigational medicinal products(s), will be reported to IRBs and NRAs as detailed in section 9.5.4.

##### 9.5.7 Reporting of Pregnancy

Any pregnancy occurring in a study participant (or the female partner of a study participant) during the trial will be reported to the PV unit by the clinical investigator using a dedicated pregnancy report form within 24 hours of pregnancy awareness. In addition, for any pregnancy occurring in a study participant, the dates of the positive pregnancy test result and pregnancy event report should also be recorded and entered in the clinical database.

The site investigator will follow the subject until pregnancy outcome is known (e.g. elective abortion, miscarriage, live birth) and notify the outcome to the PV unit. Infants born from exposed pregnancies should be followed-up at least at 6 and 12 months of age and assessed for safety (section 9.6).

Any fetus/child anomaly or birth defect detected and any other serious pregnancy consequences for the mother or the fetus/child (per definition in section 9.3) should be reported immediately upon awareness (within 24 hours) to the PV unit on an SAE report form.

Pregnancies associated with AEs that fulfill the criteria for expedited reporting, i.e. serious, unexpected, and related to investigational medicinal products(s), will be reported to IRBs and NRAs as detailed in section 9.5.4.

Further guidance on the management of pregnancy information is provided in the pregnancy report form completion guideline supplied by the Sponsor.

#### 9.6 Type and Duration of Safety Follow-up

*While the patient is in the study:*

The type of follow-up will depend on the severity and seriousness of the AE. The clinical investigator will determine need for hospitalization based on the clinical condition and level of health management required. In such situation, the clinical investigator will be in a close contact with the ward medical staff and will visit the patient approximately once a week. For outpatient care, the frequency of unscheduled visits to the trial site for clinical and laboratory monitoring will depend on the severity of the AE. When necessary, participants will be referred to specialists.

As a general rule, all AEs / SAEs should be followed-up until stabilization or resolution. Resolution of an AE is defined as the return to pretreatment status or stabilization of the condition with the expectation that it will remain chronic.

*Post study safety follow-up:*

The clinical investigator must appropriately refer all patients with AEs/AESIs/SAEs ongoing at the time of last study visit.

Last study visit may correspond to:

- Last study visit per study schedule;

- In case of withdrawal or permanent treatment discontinuation: early termination visit or post-termination follow-up visit, whichever is later (see section 7.5).

If possible, the Sponsor PV unit should be informed of any change/outcome in AESIs/SAEs (see Table 4).

The other AEs do not require post-study safety follow-up and should be given an outcome (e.g. not resolved) at the time of last study visit.

Pregnancies should be followed up until an outcome is known and the infants born from exposed pregnancies are assessed for safety at 6 and12 months of age.

**Table 4 Post study safety follow-up**

| Post study safety follow-up | |
|---|---|
| Participant status | AESI/SAE follow-up |
| Completes last study visit per study schedule but has ongoing AESI/SAEs | Followed for outcome for 4 additional weeks or until resolution/stabilization (whichever is later). |
| Is withdrawn from the study and has ongoing AESI/SAEs | Followed for outcome for 8 additional weeks or until resolution/stabilization (whichever is later). |
| Participant status | Pregnancy/infant follow-up |
| Participant becomes pregnant during the study | Followed until pregnancy outcome is known and infant at 6 and 12 months of age (as applicable). |
| Participant's partner becomes pregnant or has delivered a baby <12 months before the last study visit of her partner. | |

#### 9.7 Halting Rules

Halting rules for safety reasons will be detailed in the DSMB charter, as described in section 9.8.

#### 9.8 Safety Oversight

Safety oversight will be under the direction of the DSMB. This will include the review of the halting rules proposed by the trial investigators. The DSMB will review safety and efficacy data on each arm of the study at least semi-annually. The DSMB will review aggregate safety data to detect any increased rate of occurrence of serious suspected adverse reactions. The DSMB will also receive listings and/or reports of SAEs, AESIs, and pregnancies notified since the last DSMB meeting. The DSMB will advise the trial Sponsor of its findings. See section 16.3 for more information about the DSMB.

### 10 CLINICAL MONITORING

The purposes of trial monitoring are to verify that:

- The rights and well-being of human subjects are protected.
- The reported trial data are accurate, complete, and verifiable from source documents.
- The conduct of the trial is in compliance with the currently approved protocol/amendment(s), with ICH GCP, and with the applicable regulatory requirement(s).

A detailed endTB Monitoring Plan will be developed to specify the responsibilities and qualifications of monitors, central monitoring procedures, and site monitoring procedures. Central monitoring of data will be conducted by CRF review, with appropriate range and consistency checks programmed into the database. The Trial Master File will be stored and maintained by the study Sponsor throughout the trial. All trial specific documents will be centrally tracked and copies obtained from the sites for all communication with regulatory bodies.

The study will contract external monitor(s) who will conduct regular site visits to review documentation including, but not limited to: written informed consent documents and corresponding source documents (e.g. original patients' records and laboratory data), CRFs, SAE reports, and the site regulatory binders. Participating investigators must agree to allow trial-related monitoring and audits, ethics committee review and regulatory inspections by providing direct access to source data/documents, including electronic records, as required. Patient consent for this is obtained as part of the trial consent process. Study monitor(s) will meet with site principal investigators to discuss any problems and actions to be taken and will document visit findings and discussions in a formal report. All visit reports will be submitted to the study Sponsor.

### 11 STATISTICAL CONSIDERATIONS

#### 11.1 Study Hypotheses

The non-inferiority of an experimental arm compared to the control will be established if the difference in favorable outcome (proportion of participants with favorable outcome in the experimental arm minus proportion of participants with favorable outcome in the control arm) at Week 73 is greater than the lower equivalence margin, i.e. if the lower bound of the one-sided 97.5% CI around the treatment difference is greater than this threshold. The non-inferiority margin is 12%. In other terms, if $p_E$ and $p_C$ are the percentages of participants with favorable outcome at Week 73 in the experimental and control arms respectively, the null and alternative hypotheses to be tested for each experimental arm are:

$H_0$ : $p_E - p_C < -12$ %

$H_A$: $p_E - p_C \geq -12$ %

#### 11.2 Sample Size Considerations

*Primary efficacy analysis*

The sample size was estimated through simulations that considered the following parameters: early efficacy response at Week 8; end of treatment response frequency at Week 39; primary outcome frequency at Week 73 for each experimental arm and the control arm; expected number of effective arms among the five experimental; the type I error; and the non-inferiority margin.

We assumed an early efficacy response proportion (defined as the proportion of participants having an 8-week culture negativity) of 40% in the experimental arms, compared to the control of 30%.[11,159] Sample size estimates were calculated for 75% favorable outcome at Week 73 in the non-inferior arms, resulting from two response rate scenarios: 75% favorable outcomes at Week 73 after no relapse and after 10% relapse between Week 39 and Week 73. Favorable outcome frequency in the control arm was assumed to be 5% lower than the frequency of success in non-inferior arms, 70% at Week 73 and 70% or 80% at Week 39. Simulations varied the number of expected non-inferior arms from 1 to 3 among the 5 experimental arms. Also, simulations explored non-inferiority margins of 10 and 12%. The type I error was set at 2.5% (one-sided).

With a 12% non-inferiority margin, a sample size of 750 randomized participants provides power greater than 80% to demonstrate the non-inferiority of at least 1 (and up to 3) experimental regimens in the modified intent-to-treat (mITT) and up to 2 in the per-protocol (PP) populations. This assumes a proportion of favorable outcome at Week 73 of at least 75% in the experimental regimens and 70% in the control, with up to 10% of relapse between Weeks 39 and 73. The populations of analyses are described in section 11.4.2. Our simulations assumed a loss of 11% of subjects between the randomized population and mITT population and an additional loss of 10% between the mITT and PP populations.

The 8-week efficacy response assumptions are derived from the 8-week treatment response in the experimental arms in the Phase II delamanid and bedaquiline studies. In the delamanid study, 2-month sputum culture conversion in liquid medium occurred in more than 40% of participants in the active arms[11] and in almost 30% in the control arm. In the bedaquiline study, the frequency of two-month conversion in liquid medium was 48% in the active arm and 9% in the control arm.[119]

The 70% favorable outcome at Week 73 in the control arm is derived from existing data about the current conventional regimen and the enhanced standard afforded by the control in endTB. The control arm in endTB improves on the present conventional regimen by permitting inclusion of delamanid or bedaquiline in regimens for patients with an indication for one of those drugs.[154,] The response in the control arm at Week 73 is, therefore, be expected to be in the range of reported response in those who have not received new drugs and the response observed in those who have received new drugs. This ranges from a low of 32% at approximately 2 years after randomization in the placebo arm in the bedaquiline study[10] to a high of 74.5% for participants who received 6 months of delamanid.[38] In between are a series of estimates: 54% in the individual-patient data meta-analysis of patients with no new drugs;[8] 55% in the placebo arm in the delamanid study[38](some received up to two months of delamanid); 58% for participants who received 6 months of bedaquiline;[10] and 62% in recent meta-analyses of observational studies of patients who received no new drugs.[6,9] At 70%, the expected rate of response at Week 73 in the control group in endTB is considerably better than the response rates in both the current conventional regimen and the bedaquiline arm (58%) if slightly worse than among those receiving 6 months of delamanid (74.5%).

The 75% response at Week 73 in the experimental arm also reflects available data. As noted, the best reported response using a standard-duration treatment with a new drug is 74.5% among those receiving delamanid. A series of observational studies of short-course MDR-TB treatment are reporting treatment success of greater than 80%.[62,63,98,99] The likely efficacy of the endTB regimens relative to the short-course regimens previously reported is unknown. Adhering to the principle of conservatism (i.e., assumptions that would decrease the expected difference between the experimental and control arms and increase the study sample size), we opted for a Week 73 response at the lower end of the range from existing data, closer to the expected response in the control.

One of the stated objectives of the study is to identify as many effective regimens as possible. It is not possible to estimate from previous evidence how many of the five regimens are likely to be effective. However, we opted to have sufficient sample size to assure 80% power to detect if more than half (3 of 5) of the regimens are non-inferior. This does allow the possibility that the study is underpowered if all 5 experimental regimens are non-inferior, but with a modest treatment effect. Sample size increases to detect 5 non-inferior regimens were not justifiable. We elected a 12% non-inferiority margin for several reasons. First, the comparator in endTB reflects an important improvement over the conventional regimen by including at least one new drug. The expected proportion of favorable outcomes in the control arm reflects almost a 10% improvement over the current standard experimental regimens. The concern about biocreep is, therefore, mitigated. Second, relative to the control arm (and the current conventional regimen), the experimental regimens would result in a significantly reduced pill burden and treatment duration, expected better tolerability, and expected increased adherence achieved by reducing the treatment duration from approximately 86 weeks (in the control), to 39 weeks (in the experimental arms). Third, if the control is as effective as assumed for the sample size estimations (70%), any non-inferior regimen could be concluded, with 97.5% confidence to have a favorable outcome frequency of at least 58%. If treatment success is 75% in the control arm, for any non-inferior experimental regimen, we would be able to conclude with 97.5% confidence that response is 63% or greater. These are the absolute lowest bounds of regimens that would be declared non-inferior in the context of this study. Both are superior to the proportion of successful outcomes on MDR-TB reported by WHO (48%) and to treatment success in the placebo arms in both the delamanid (55%) and bedaquiline (32%) trials. And, the response in one or more experimental arms may be even greater than the 75% estimated; this could result in a lower bound of treatment effect that is greater than the 12% margin. Taken together with the shortened, simplified dosing, this could be considered an improvement that programs or physicians opt to implement for some or all patients. Lastly, we note that the 12% margin was used in one other novel-regimen study, the STAND study, which was vetted by both the US FDA and the EMA. It was primarily a study of a new regimen for drug-susceptible TB. Arguably, since current treatment for drug-susceptible TB is very effective and well tolerated, there would be little tolerance for a new regimen with any decrease in efficacy. In contrast, the control in endTB will be approximately 20 months in length and likely poorly tolerated. A modest loss in efficacy could be accepted in exchange for easier delivery, shorter duration, and improved tolerability.

The sample size calculations were done in R (programming language), using bootstrapping methods for the simulations.

*Exploratory analysis of toxicity related to linezolid dose reduction*

For the second randomization assigning the linezolid dose-reduction strategy, we assumed a sample size of 280 patients randomized to endTB regimens 1-4, which is a conservative estimate representing the absolute minimum from prior simulations.[160] Using the proportions of linezolid-related AEs from published studies, [76,42] we assumed that the proportion of AEs observed in the 300 mg daily arm would be 69% as previously reported in the only other clinical trial to investigate the 300 mg dose,[76] and 52% for the 600 mg thrice weekly group. The latter estimate is extrapolated, conservatively, from the meta-analysis among patients receiving linezolid doses lower than 600 mg daily. It exceeds the upper limit of the 95% CI reported for AEs in that study: 34% (95% CI, 23-46%).[42] Based on these effect sizes, and assuming 10% of patients are lost to follow-up, a time-to-event analysis with a sample size of $n$ = 280 patients randomized 1:1 to the two linezolid dosing strategies would have 80% power to detect a hazard ratio of 1.43 for severe linezolid-related toxicity with 300 mg daily compared to 600 mg thrice weekly in a two-sided, two-sample log-rank test with 5% significance.

#### 11.3 Planned Interim Analyses

Interim analyses will be performed on both early (culture negativity at Week 8) and end-of-treatment outcomes (at Week 39) in order to adapt the randomization. They will occur first when approximately 30 participants have been randomized to each arm, and then approximately every 4 weeks.

Stopping rules for futility will be defined, based on Week 73 treatment response (See section 11.3.2).

Independent of this efficacy review, regular reviews of the efficacy and safety data will be performed by the DSMB.

##### 11.3.1 Safety Review

The DSMB will regularly review the trial data, especially on enrolment of the participants, as well as efficacy and safety of study treatments. Details of the review and stopping rules will be described in the DSMB charter (see sections 9.7 and 16.3).

##### 11.3.2 Efficacy Review

Further efficacy review may inform dropping of regimen(s) for futility. Stopping an arm early may be indicated if the posterior probability of a regimen being non-inferior to the control at Week 73 falls below a predefined threshold of 5%.

Conversely, if the posterior probability is at least 95% that a regimen is superior to the control at Week 73, stopping for efficacy will be considered. This may trigger a review by the DSMB to decide if this superior regimen should be stopped (i.e. no more participants are randomized to this regimen) or if the evidence is to be balanced with the safety data.

#### 11.4 Final Analysis Plan

In this section, analysis populations, analyses of primary and main secondary endpoints as well as main data conventions will be described. A more detailed statistical analysis plan (SAP) will be provided separately and finalized before the database lock. Despite a Bayesian approach to the interim analyses, a frequentist approach will be used for final analyses on all endpoints.

##### 11.4.1 Analysis Populations

The safety population will include all enrolled participants who had received at least one dose of study treatment (exposed). Safety analyses will be based on the treatment actually received after inclusion (as treated).

The first efficacy population will be the mITT population, containing all randomized participants with culture-positive, FQ-susceptible and rifampicin-resistant TB. Exclusion from the mITT population will occur if screening/baseline DST results from the designated study lab indicate resistance (using a test deemed to be reliable by the trial reference lab, ITM) to bedaquiline, clofazimine, delamanid, fluoroquinolone, and/or linezolid. Participants whose sputum culture is not positive will be excluded from the mITT population. Participants without any post baseline data will be also excluded from the mITT population.

Participants from the mITT population will be analyzed in the arm to which they were randomly allocated (as randomized).

The second efficacy population, the PP population, is the same as the mITT population with the exclusion of patients who, for reasons other than treatment failure or death, do not complete a protocol-adherent course of treatment. A participant will be considered to have completed a protocol-adherent course of treatment if they have taken 80% of expected doses within 120% of the regimen duration. Participants who receive more than 7 days of either a prohibited concomitant medication or an IP not prescribed according to protocol will also be excluded from the PP population.

##### 11.4.2 Primary Endpoint Analyses

Analysis of the primary endpoint will compare the proportions of participants with a favorable outcome at Week 73 (as defined in section 3.2.1) between each experimental arm and the internal control. For all pairwise comparisons, a two-sided 95% confidence interval of the difference will be estimated; a 2-sided 95% CI of the proportion of participants with a favorable outcome will be also be provided for all regimens.

The non-inferiority of an experimental arm compared to the control will be established if the difference in favorable outcome (proportion of participants with favorable outcome in the experimental arm minus proportion of participants with favorable outcome in the control arm) at Week 73 is greater than the lower equivalence margin, i.e. if the lower bound of the one-sided 97.5% CI—which corresponds to the lower bound of the 2-sided 95% CI—is greater than or equal to -12%.

The main primary efficacy analyses will be performed on both mITT and PP populations for the non-inferiority comparisons.

As multiple experimental regimens will be compared to one active control, the type I error will be controlled by ordering the non–inferiority comparisons, experimental regimens will be ordered by decreased proportions of favorable outcome. A fixed sequence approach will be considered: the regimen with the highest proportion of favorable outcomes will be compared to the control. If non-inferiority is concluded, then a comparison between the control and the experimental regimen having the second highest proportion of favorable outcomes will be performed, and so on as long as non-inferiority is concluded. Once the non-inferiority is not demonstrated, the comparisons will stop. All the previous comparisons will have been done at the full alpha level of 2.5%.

No additional adjustments will be made to control the type I error.

Adjusted analyses on the primary endpoint will be also performed by controlling for covariates including: country, presence of comorbidities (including SARS-CoV-2 infection and COVID-19), degree of resistance, prior exposure to TB treatment, extent of disease (based on chest X-ray), and BMI. Kaplan-Meier estimates of probability of favorable outcomes will also be generated for the mITT population.

Non-inferiority analyses may be performed in other populations. These and other sensitivity analyses will be detailed in the statistical analysis plan.

##### 11.4.3 Data Handling Convention

In principle, data collected within 2 weeks before or after planned visits will be included in the analysis of the endpoint. There are two exceptions, at Weeks 73 and 104 when the window may be extended to 30 days after the visit. This exception applies to participants who are missing either data on evolution or culture results from samples collected between weeks 65 and 73. If data on evolution in the 30 days after Week 73 or culture results from samples collected in the 30 days after Week 73 are available, they will inform outcome classification. Similarly, the window will be extended to 30 days after the Week 104 visit for participants who are missing either data on evolution or culture results from samples collected between weeks 97 and 104. If data on evolution in the 30 days after Week 104 or culture results from samples collected in the 30 days after Week 104 are available, they will inform outcome classification.

### 12 Source Documents and Access to Source Data/Documents

Appropriate medical and research records will be maintained for this trial, in compliance with ICH E6 GCP, Section 4.9 and regulatory and institutional requirements for the protection of confidentiality of subjects.

The following individuals and groups will have access to study records:

- Members of the study team.
- IRBs that review the study (including IRB members, staff, and legal counsel).
- Regulatory agencies in study site countries.
- Study Monitor(s).

Authorized representatives of the Sponsor, and regulatory agencies indicated above will be permitted to examine (and when required by applicable law, to copy) clinical records for the purposes of quality assurance reviews, audits and evaluation of the study safety and progress.

Source data are all information, original records of clinical findings, observations, or other activities in a clinical trial necessary for the reconstruction and evaluation of the trial. Examples of these original documents and data records include, but are not limited to, hospital records, clinical and office charts, laboratory notes, X-rays, and subject files and records kept at the laboratory.

### 13 QUALITY CONTROL AND QUALITY ASSURANCE

SOPs for quality management will be developed and detailed in a separate Quality Management Plan.

The executing institutions are responsible for conducting routine quality assurance (QA) and quality control (QC) activities to internally monitor study progress, protocol compliance and accuracy of data in relation to source documentation. The site Principal Investigator will provide direct access to all trial-related sites, source data/documents, and reports for the purpose of monitoring and auditing by the Sponsor, and inspection by local and regulatory authorities.

External clinical monitors will verify that the clinical trial is conducted and data are generated, documented (recorded), and reported in compliance with the protocol, GCP, and the applicable regulatory requirements. Clinical monitoring reports will be submitted to the Sponsor.

The data centers at each site will implement quality control procedures beginning with the data entry system and will generate data quality control checks that will be run with the database. Any missing data or data anomalies will be communicated to the local study team for clarification and resolution.

The site Principal Investigator will ensure all study personnel are appropriately trained and that applicable training documentation is maintained on site. All key study staff will be trained and certified in good clinical practices.

### 14 ETHICS/PROTECTION OF HUMAN SUBJECTS

The protection of human research subjects will be assured throughout the study execution and reporting.

#### 14.1 Ethical Standard

The investigator will ensure that this study is conducted in full conformity with the current revision of the Declaration of Helsinki (October 2013) [152] or with the International Conference for Harmonization Good Clinical Practice (ICH-GCP) [161] regulations and guidelines, whichever affords the greater protection to the subject. In addition, this study will be conducted in accordance with local standards.

#### 14.2 Institutional Review Board

Before they are placed into use, the study protocol and informed consent forms (ICFs) will be reviewed and approved by IRBs supervising all study sites and ethics authorities that oversee research conducted by Harvard Medical School/PIH and MSF/Epicentre. Any amendments to the protocol or consent materials will be reviewed and approved by the IRBs before they are submitted to local authorities and ethics committees for approval.

#### 14.3 Informed Consent Process

Informed consent is a process that will be initiated prior to the individual's agreeing to participate in the study and will continue throughout the individual's study participation. Extensive discussion of risks and possible benefits of screening and therapy will be provided by study staff to potential subjects (and, with permission from potential subjects, their families). Potential subjects will receive counseling about study objectives and procedures, potential toxicities, and the informed consent process. Consent forms describing in detail study screening, study interventions & products, study procedures and risks will be given to the subject and written documentation of informed consent will be required prior to screening and to starting intervention/administering study product. Consent forms will be IRB approved, will contain all the elements required by the ICH E6 Guidelines for Good Clinical Practice.

Additional elements required by local regulations and written translations in the native language might have to be generated at local level. English versions will have to be made available for IRB and Sponsor's review upon request.

The subject will be asked to read and review the document. Upon reviewing the document, the person obtaining consent will explain the research study to the subject and answer any questions that may arise. For subjects who speak and understand the language used in the consent document, but are unable to read or write, all of the information in the consent form will be communicated verbally, in the presence of an adult witness who can read and write and who is not a member of the study team; informed consent requires the signature or mark of the subject. The subject will sign or mark the informed consent document prior to any procedures being done or data being recorded specifically for the study. The subjects will have the opportunity to discuss the study with their surrogates or think about it prior to agreeing to participate. The subjects may withdraw consent at any time throughout the course of the trial. A copy of the signed and dated informed consent document will be given to the subjects for their records. The rights and welfare of the subjects will be protected by emphasizing, to subjects, that the quality of their medical care will not be adversely affected if they decline to participate in this study. The informed consent process will be recorded in the subject's clinical chart.

During the informed consent process, the subjects receive information about anticipated payment and/or expenses to the subject for study participation. Value and structure of compensation (e.g., money, food supplements) are determined in conjunction with local authorities in each study site. The details are described in each informed consent form. Investigational drugs and exams will be provided free of charge throughout the study. All exams are free of charge including for patients still followed for safety after withdrawal or the end of study. Subjects will not incur any costs as a result of their participation.

In an emergency, specialist consultation, treatment, and/or hospitalization costs will be covered by the trial. For non-urgent situations, specialist consultations fees for concomitant disease may be covered. Resulting treatment will not be systematically covered by the trial when the disease is not directly related to the trial intervention.

###### 14.3.1 Informed Consent/Assent Process (in Case of a Minor)

For participants considered minor per local laws, between 15 and 17 years old (or any different legal age for majority), participant's assent will be obtained in addition to the consent of the parent or guardian. For subjects who speak and understand the language used in the assent document, but are unable to read or write, all of the information in the assent form will be communicated verbally, in the presence of an adult witness who can read and write and who is not a member of the study team; assent requires the signature or mark of the subject. The subject will sign or mark the assent document prior to any procedures being done or data being recorded specifically for the study. The same process will be used for consent by a parent/guardian who is not able to read or write.

###### 14.3.2 Consent for follow up of pregnancy in female partners of male participants

In case of pregnancy in the female partner of a male participant, consent will be sought for follow-up of the pregnancy, including of the child at least 6 and 12 months after delivery.

###### 14.3.3 Special Populations

*a. Children*

Children younger than 15 years old will be excluded from this study because there is limited information on the safety and tolerability of the experimental regimens among children. Minors 15-17 years old will be included, if approved by local regulators, as they generally experience tuberculosis and metabolize anti-TB drugs like adults. If their safety among adults is confirmed with this study, research examining safety and tolerability among children will be considered.

*b. Women*

Women who are pregnant or breastfeeding at screening will not be included in this study because of the lack of evidence on the safety and tolerability of these regimens on fetal development.

Based on strong recommendation from the Data Safety and Monitoring Board, the Global TB Community Advisory Board, scientific publications and draft FDA guidance,[162–171] women becoming pregnant during study participation and whose pregnancy is not terminated may remain on study treatment, if all the following conditions are met:

- the clinical trial insurance policy subscribed in the country covers pregnant participant, including damage to and loss of the fetus;
- the local authorities and ethics committee(s) approve;

AND

- the participant is at least 18 years of age;
- in the individual participant, the site PI, according to his/her clinical judgment, considers that the expected benefits of continuing the treatment outweigh the risks of ongoing fetal exposure;
- the CAC has reviewed the Site PI's recommendation;
- the participant is informed of the therapeutic options, potential risks and expected benefits and her separate specific consent is obtained.

##### 14.3.4 Subject Confidentiality

This study will be performed in accordance with the ICH guidelines for GCP and local national standards for protection of privacy of identifiable health information. Confidentiality will be guaranteed by a suitable coding system, which will anonymize all patient data. Patients will be given a study identification number. All study records will be managed in a secure and confidential fashion. Study records will be maintained in locked cabinets, and computer records will be password protected. Access to study records will be restricted to specified team members and entities mentioned in section 12. Disclosure to other parties is strictly prohibited. Study team members are subject to the obligation of professional secrecy. Methods for secure data handling are detailed below in section 15 and in the Data Management Plan.

#### 14.4 Study Discontinuation

In the event that the study is discontinued (e.g. by the investigator, the Sponsor, the DSMB, and/or regulatory authorities), subjects on study phase treatment will continue to receive TB treatment in accordance with national guidelines. Treatment will be offered free of charge, as guaranteed through the public health systems.

#### 14.5 Future Use of Stored Specimens

This section refers only to use of specimens and strains of *M. tuberculosis* not specified in the current protocol.

Subjects (and their legal representatives as applicable) will be asked during the endTB consent process to provide written informed consent for their strains to be stored and analyzed for any future studies. A subject may consent to participation in the study without consenting to future use of stored strains. Relevant IRBs will oversee any future research using these specimens.

Specimens and strains may be stored in a specimen or strain repository or bank for up to 20 years. These specimens and strains will be used only for purposes that are consistent with improving diagnosis (including resistance testing) and treatment of TB. The specific conditions governing strain and specimen banking will be detailed in agreements that comply with intellectual property standards of the Sponsor. All necessary permissions will be obtained before exporting any specimen or strain out of the participating countries according to national regulations in the countries concerned.

### 15 DATA HANDLING AND RECORD KEEPING

#### 15.1 Data Management Responsibilities

Data will be collected and entered into an electronic Case Report Form (eCRF). Each participant will be assigned a unique study ID. eCRFs will be checked for accuracy and completeness at the site by staff designated for data quality control; checks for consistency will be implemented at the data entry level on site and centrally after data entry.

Procedures to ensure good quality data will be detailed in the Data Management Plan, including the details of the following:

- eCRFs versions and completion guidelines;
- Database structure, dictionary, data entry system and validation of the database;
- Methods of data collection;
- Type of data entry;
- Data preparation before entry onto electronic system;
- Data entry validation, data cleaning, data validation plan and data clarification form generation;
- Data flow;
- Database audit trail;
- Database lock;
- Data review checks to support monitoring;
- SAE reconciliation;
- IVRS/IWRS data reconciliation;
- ECG data reconciliation;
- Security/backup;
- Electronic data transfer rules;
- Archiving and security arrangements;
- Coding conventions.

#### 15.2 Data Capture Methods

Data collected on site will be entered locally in an eCRF, a web-based system that is GCP compliant. The clinical data will be entered directly from the source documents, or transcribed on study-specific forms before data entry into the eCRF.

#### 15.3 Types of Data

Data for this study will include demographics; physical examination (including neuro, ophthalmologic, vital signs, and performance status); investigational drugs; review of TB symptoms, concomitant medications and AEs; adherence; safety laboratory, microbiologic, molecular lab results and outcome measures.

Data will be managed centrally at the Study Data Management Center (Epicentre). Additionally, data from the SAE/pregnancy reports will also be entered in a separate database at the centralized MSF Pharmacovigilance Unit.

#### 15.4 Timing/Reports

Regular data review and data cleaning for quality control will be organized in a blinded way. The DSMB will receive blinded and unblinded reports organized as detailed in the DSMB charter.

#### 15.5 Study Records Retention

Study documents will be retained for a minimum of 10 years after study completion. An SOP will be developed to detail the conditions and specifications of the storage during this period, at the site and Sponsor's level.

#### 15.6 Protocol Deviations

No deviations from, or changes to, the protocol should be implemented without prior written IRB, regulatory authority approval/favorable opinion and approval from the Sponsor. The investigator, or designated site staff, is responsible for documenting and explaining any deviations from the protocol. As a result of deviations, corrective actions are to be developed by the sites and implemented promptly.

It is the responsibility of the sites to use continuous vigilance to identify and report deviations according to the guidelines of the Sponsor. All deviations from the protocol must be addressed in study subject source documents. Protocol deviations must be sent to the Sponsor; dissemination to the local IRB will be in accordance with local regulations.

### 16 TRIAL BOARDS AND COMMITTEES

#### 16.1 Scientific Advisory Committee (SAC)

The SAC is an external and independent committee that will provide expertise on the relevance and scientific validity of the trial design and implementation. It will review and advise on the relevance of the studies' objectives within the context of the DR-TB research landscape, the appropriateness of the study design and methods, the safety of the proposed regimens, the feasibility to meet the stated objectives of the project, the complementarity of the proposed trial with other ongoing or planned external trials, trial progress upon receiving semi-annual project reports, and any important scientific decisions or changes made by the trial teams during the course of the trial (e.g., major protocol amendment) or based on the reports from the DSMB. The committee will consist of at least one each of the following: clinical TB expert, clinical trial statistician with expertise in Bayesian and other adaptive designs, TB clinical trials expert, clinical trial expert with expertise in testing drug combinations from a different medical field (e.g., HIV), and a mycobacteriologist. Meetings will be held at least annually to discuss the trial update reports.

#### 16.2 Global Tuberculosis Community Advisory Board (TB-CAB)

Initiated by the Treatment Action Group in 2011 to catalyze research-literate community activism, TB-CAB acts "in an advisory capacity to institutions conducting clinical trials of new TB drugs, treatment regimens, diagnostics, and vaccines, and to provide input on study design early access, regulatory approval, postmarketing, and implementation strategies".[172] The TB CAB comprises HIV and TB activists from both high- and low-TB-burden countries around the world. Although the TB-CAB is not a committee specific to endTB, endTB has sought and incorporated input from the TB-CAB throughout the development of the protocol. TB-CAB will be updated and consulted for input at similar intervals to the SAC.

#### 16.3 Data and Safety Monitoring Board (DSMB)

The DSMB is an independent consultative board that will provide general advice on the progress of the trial, including the rate of inclusions, the quality of follow-up, the overall rate of drug-related AEs, changes in biological markers, the overall incidence of primary outcomes, and the number of subjects needed. They will advise as to the continuation of the study according to the protocol dependent of safety and efficacy data available only to the DSMB; this advice may include the recommendation that one or more study arm is dropped or modified based on emerging data. The DSMB will operate under the rules of a charter approved by the Sponsor which will be written at the organizational meeting of the DSMB. The charter will describe the following: composition (likely an MDR TB clinician, a cardiologist, a statistician, and a mycobacteriologist); member responsibilities; frequency of reviews; and stopping/halting rules. The DSMB may also conduct and interpret intermediate analyses, if necessary. Please also see section 9.7.

#### 16.4 Clinical Advisory Committee (CAC)

The CAC is composed of clinical experts representing various disciplines and specialties. It serves as an advisory body to endTB site investigators by providing consultation regarding subject eligibility, case management, medical monitoring, and permanent treatment discontinuation. Where the CAC lacks relevant expertise for a particular issue facing the committee, the CAC may propose recruitment of an additional ad hoc member to serve this function. Members of the CAC can at any time convene an ad-hoc meeting, virtually or in-person, in response to consultation requests from the sites.

#### 16.5 Event Validation Group (EVG)

A committee, Event Validation Group (EVG), consisting mainly of clinical experts, will be responsible for applicable outcome event validation reported by each site. Functioning of EVG will be detailed in an SOP.

### 17 PUBLICATION, DISSEMINATION AND ACCESS

Results of this study will be submitted for publication in a peer-reviewed scientific journal. The study PI(s) will retain the primary responsibility for and make every effort to expedite the preparation of publications.

Authorship will be extended to the following individuals: study investigators, site PIs, and other individuals having major contribution to the study design, implementation, data analysis, and preparation of the written manuscript following International Committee of Medical Journal Editors criteria.[173]

Results will be disseminated to study participants. Dissemination methods of results may include one or more of the following: summary of findings letter distributed to study participants and/or open community meetings in the settings in which the trial is implemented. Final dissemination approaches will be developed in conjunction with the Global TB Community Advisory Board, and local Community Advisory Boards, where possible. Results will also be shared with normative bodies that define policy and practice guidance.

De-identified data may be shared for guideline development and for other efforts to improve tuberculosis treatment and diagnostics.

Results will be used to facilitate post-trial access in trial settings and elsewhere for any experimental regimens found to be non-inferior to the standard of care control. All drugs included in the experimental regimens are already available for purchase through the Global Drug Facility, which procures drugs of assured quality at the best available price for low- and middle-income countries. endTB partners will continue to advocate at global level for lower prices of the drugs comprising non-inferior endTB regimens.

New, non-inferior regimens will result in a tremendous simplification of the management of MDR/RR-TB; this will make treatment easier for practitioners to administer, programs to decentralize and consequently make treatment more accessible to patients. Non-inferior endTB regimens can also address one of the major reasons for the limited access to treatment for MDR/RR-TB: high drug costs due to fragmentation of market across many different products and formulations. By identifying priority regimens that comprise four or five drugs, the results of endTB could support increased size, stability, and consolidation of the market, conceivably making the market more attractive to competitive producers and resulting in reduced price.

154. World Health Organization. *The use of delamanid in the treatment of multidrug-resistant tuberculosis interim policy guidance*. (World Health Organization, 2014).

155. World Health Organisation. *Companion handbook to the WHO guidelines for the programmatic management of drug-resistant tuberculosis*. (World Health Organization, 2015).

161. ICH International Conference on Harmonization - Integrated Addendum to ICH E6(R1): Guideline for Good Clinical Practice E6(R2). (2016).

162. Gupta, A. *et al.* Toward Earlier Inclusion of Pregnant and Postpartum Women in Tuberculosis Drug Trials: Consensus Statements From an International Expert Panel. *Clin. Infect. Dis.* **62**, 761–769 (2016).

163. Council for International Organizations of Medical Sciences (CIOMS). International Ethical Guidelines for Health-related Research Involving Humans, Fourth Edition. Geneva. (2016).

164. Jaspard, M. *et al.* Bedaquiline and Linezolid for Extensively Drug-Resistant Tuberculosis in Pregnant Woman. *Emerg. Infect. Dis.* **23**, 1731–1732 (2017).

165. U.S. Department of Health and Human Services, Food and Drug Administration & Center for Drug Evaluation and Research (CDER). Guidance for Industry Pulmonary Tuberculosis: Developing Drugs for Treatment.

166. NIH. Task Force on Research Specific to Pregnant Women and Lactating Women (PRGLAC). *https://www.nichd.nih.gov/About/Advisory/PRGLAC*.

167. Fauci, A. & Bianchi, D. W. NIAIDS. Refining the Maternal and Pediatric Research Agenda: 2020 and Beyond. *Httpswwwniaidnihgovresearchrefining-Matern.--Pediatr.-Res.-Agenda-2020--Beyond*.

168. Treatment Action Group. It's Time to Deliver - Including Pregnant and Lactating Women in Clinical Research. *Httpwwwtreatmentactiongrouporgcontentwebinar--Pregnant--Lact.-Women-Clin.-Res.*

### APPENDICES

**Appendix 1: Schedule of Events**

**Appendix 2: Statement of Compliance**

**Appendix 3: Institutions / Site Principal Investigators and Study Sites**

### Appendix 1: endTB Clinical Trial Schedule of Events

| | | Treatment | | | | | | | | | | | | | | | | | | | | Follow-Up | | | | | | | | | | | | |
|---|---|---|---|---|---|---|---|---|---|---|---|---|---|---|---|---|---|---|---|---|---|---|---|---|---|---|---|---|---|---|---|---|---|---|
| | Screening | Baseline | W1 | W2 | W3 | W4 | W5 | W6 | W7 | W8 | W9 | W 10 | W 11 | W 12 | W 16 | W 20 | W 24 | W 28 | W 32 | W 36 | W39 | W43 | W47 | W53 | W59 | W65 | W73 | W81 | W89 | W97 | W 104 | Sub total (minimal count) | Early Termination | Post-termination follow-up |
| Visit | 1 | 2 | 3 | 4 | 5 | 6 | 7 | 8 | 9 | 10 | 11 | 12 | 13 | 14 | 15 | 16 | 17 | 18 | 19 | 20 | 21 | 22 | 23 | 24 | 25 | 26 | 27 | 28 | 29 | 30 | 31 | | | |
| Window Period | | | +/-2 | +/-2 | +/-2 | +/-2 | +/-2 | +/-2 | +/-2 | +/-2[#] | +/-2 | +/-2 | +/-2 | +/-2 | +/-7 | +/-7 | +/-7 | +/-7 | +/-7 | +/-7 | +/-14 | +/-14 | +/-14 | +/-14 | +/-14 | +/-14 | +/-30 | +/-14 | +/-14 | +/-14 | +/-30 | | | |
| Eligibility | | | | | | | | | | | | | | | | | | | | | | | | | | | | | | | | | | |
| Subject Consent | X | X | | | | | | | | | | | | | | | | | | | | | | | | | | | | | | 2 | | |
| Demographics | X | X | | | | | | | | | | | | | | | | | | | | | | | | | | | | | | 2 | | |
| Medical History | X | | | | | | | | | | | | | | | | | | | | | | | | | | | | | | | 1 | | |
| Inclusion/ Exclusion | X | X | | | | | | | | | | | | | | | | | | | | | | | | | | | | | | 2 | | |
| Clinical Evaluation | | | | | | | | | | | | | | | | | | | | | | | | | | | | | | | | | | |
| Vital Signs | X | X | X | X | X | X | X | X | X | X | X | X | X | X | X | X | X | X | X | X | X | X | X | X | X | X | X | X | X | X | X | 31 | X | X |
| Interval Medical History | | X | X | X | X | X | X | X | X | X | X | X | X | X | X | X | X | X | X | X | X | X | X | X | X | X | X | X | X | X | X | 30 | X | X |
| Physical Exam | X | X | X | X | X | X | X | X | X | X | X | X | X | X | X | X | X | X | X | X | X | X | X | X | X | X | X | X | X | X | X | 31 | X | X |
| TB Symptom Assessment | | X | X | X | X | X | X | X | X | X | X | X | X | X | X | X | X | X | X | X | X | X | X | X | X | X | X | X | X | X | X | 30 | X | X |
| ECOG Assessment | | X | | | | | | | | | | | | | | | | | | | X | | | | | | X | | | | X | 4 | X | X |
| Neurologic Exam | | X | | | | X | | | | X | | | | X | X | X | X | X | X | X | X | X | X | | | | X | | | | X | 15 | X | |
| Ophthalmologic Exam | | X | | | | X | | | | X | | | | X | X | X | X | X | X | X | X | X | X | | | | X | | | | X | 15 | X | |
| Mental Health Assessment | | X | | | | | | | | | | | | | | | | | | | | | | | | | X | | | | X | 3 | X | |
| Treatment | | | | | | | | | | | | | | | | | | | | | | | | | | | | | | | | | | |
| Randomization | | X* | | | | | | | | | | | | | X** | | | | | | | | | | | | | | | | | (1) | | |
| Treatment Compliance | | | X | X | X | X | X | X | X | X | X | X | X | X | X | X | X | X | X | X | X | X[C] | X[C] | X[C] | X[C] | X[C] | X[C] | X[C] | X[C] | X[C] | X[C] | 19 | X[C] | |
| Adherence Counseling | | X | X | X | X | X | X | X | X | X | X | X | X | X | X | X | X | X | X | X | X | X[C] | X[C] | X[C] | X[C] | X[C] | X[C] | X[C] | X[C] | X[C] | | 20 | | |
| Concomitant Medication Review | X | X | X | X | X | X | X | X | X | X | X | X | X | X | X | X | X | X | X | X | X | X | X | X | X | X | X | X | X | X | X | 31 | X | X |
| AE Review | | | X | X | X | X | X | X | X | X | X | X | X | X | X | X | X | X | X | X | X | X | X | X | X | X | X | X | X | X | X | 29 | X | X |
| Laboratory Testing | | | | | | | | | | | | | | | | | | | | | | | | | | | | | | | | | | |
| CBC | X[1] | | | | | X | | | | X | | | | X | X | X | X | X | X | X | X | X | X | | | | | | | | X | 14 | X | |
| Total Ca++, K+, Mg++ | X[1] | | | | | X | | | | X | | | | X | X | X | X | X | X | X | X | X | X | | | | | | | | | 13 | X | |

| | Screening | Baseline | W1 | W2 | W3 | W4 | W5 | W6 | W7 | W8 | W 9 | W 10 | W 11 | W 12 | W 16 | W 20 | W 24 | W 28 | W 32 | W 36 | W39 | W 43 | W 47 | W 53 | W 59 | W 65 | W 73 | W 81 | W 89 | W 97 | W 104 | Sub total (minimal count) | Early Termination | Post-termination follow-up |
|---|---|---|---|---|---|---|---|---|---|---|---|---|---|---|---|---|---|---|---|---|---|---|---|---|---|---|---|---|---|---|---|---|---|---|
| Visit | 1 | 2 | 3 | 4 | 5 | 6 | 7 | 8 | 9 | 10 | 11 | 12 | 13 | 14 | 15 | 16 | 17 | 18 | 19 | 20 | 21 | 22 | 23 | 24 | 25 | 26 | 27 | 28 | 29 | 30 | 31 | | | |
| Window Period | | | +/- 2 | +/- 2 | +/- 2 | +/- 2 | +/- 2 | +/- 2 | +/- 2 | +/- 2# | +/- 2 | +/- 2 | +/- 2 | +/- 2 | +/- 7 | +/- 7 | +/- 7 | +/- 7 | +/- 7 | +/- 7 | +/- 14 | +/- 14 | +/- 14 | +/- 14 | +/- 14 | +/- 14 | +/- 30 | +/- 14 | +/- 14 | +/- 14 | +/- 30 | | | |
| Creatinine | X[1] | | | | | X | | | | X | | | | X | X | X | X | X | X | X | X | X | X | | | | | | | | | 13 | X | |
| AST and ALT[2] | X[1] | | | | | X | | | | X | | | | X | X | X | X | X | X | X | X | X | X | | | | | | | | | 13 | X | |
| Total and Direct Bilirubin | X[1] | | | | | X | | | | X | | | | X | X | X | X | X | X | X | X | X | X | | | | | | | | | 13 | X | |
| Albumin | X[1] | | | | | | | | | | | | | | | | | | | | | | | | | | | | | | | 1 | | |
| TSH | X[3] | X[8] | | | | | | | | | | | | | | | X | | | | | | X | | | | | | | | | 3 | X | |
| HbA1c | X[4] | X[8] | | | | | | | | | | | | | | | X[5] | | | | | | X[5] | | | | X[5] | | | | | (1) | X[5, 17] | X[5, 17] |
| HIV test | X[6] | | | | | | | | | | | | | | | | | | | | | | | | | | | | | | | 1 | | |
| CD4 count and viral load (in HIV+) | X[7] | X[8] | | | | | | | | | | | | | | | X[9] | | | | | | X[9] | | | | X[9] | | | | | (0) | X[9,17] | X[9,17] |
| Hepatitis B serology (anti-HBc total and HbsAg) | X | X[8] | | | | | | | | | | | | | | | | | | | | | | | | | | | | | | 1 | | |
| Hepatitis C Serology: HCVAb | X | X[8] | | | | | | | | | | | | | | | | | | | | | | | | | | | | | | 1 | | |
| Pregnancy test[11] | X | X[10] | | | | | | | | | | | | | | | | | | | | | | | | | | | | | | (2) | X | |
| Sputum Specimen Testing | | | | | | | | | | | | | | | | | | | | | | | | | | | | | | | | | | |
| Smear Microscopy | X | | | X | | X | | | | X | | | | X | X | X | X | X | X | X | X | X | X | X | X | X | X | X | X | X | X | 22 | X | X |
| Culture (LJ & MGIT) | X | | | X | | X | | | | X | | | | X | X | X | X | X | X | X | X | X | X | X | X | X | X | X | X | X | X | 22 | X | X |
| Rapid Molecular Test for RIF Resistance | X[12] | X[8] | | | | | | | | | | | | | | | | | | | | | | | | | | | | | | 1 | | |
| Rapid molecular test for FQ resistance | X[12] | X[8] | | | | | | | | | | | | | X[13] | | | | | | | | | | | | | | | | | (1) | | |
| DST (conventional 1st and 2nd line) | X | | | | | | | | | | | | | | X[13,14] | | | | | | | | | | | | | | | | | (1) | X[17] | X[17] |
| DST (new drugs) | X[14] | | | | | | | | | | | | | | X[14] | | | | | | | | | | | | | | | | | (0) | X[17] | X[17] |
| Genotyping[14] | X | | | | | | | | | | | | | | X | | | | | | | | | | | | | | | | | (0) | X[17] | X[17] |
| Specimen Storage | X[15] | | | | | | | | | | | | | | X[15] | | | | | | | | | | | | | | | | | (1) | X | |
| Special Assays or Procedures | | | | | | | | | | | | | | | | | | | | | | | | | | | | | | | | | | |
| Audiometry | | X | | | | X | | | | X | | | | X | X | X | X | X | X | X | X | X | X | | | | X | | | | X | 15 | X | |
| ECG[1] | X[1] | X | X | X | X | X | X | X | X | X | X | X | X | X | X | X | X | X | X | X | X | X | X | X | | | X | | | | | 25 | X | |
| Chest X-ray | | X[16] | | | | | | | | X | | | | | | | | | | | X | | | | | | X | | | | X | 5 | X[17] | X[17] |

#. W8 visit window should occur within 8 weeks (+/-2 days) of study treatment initiation.

*: Randomization for treatment regimen assignment.

**: Only for patients receiving linezolid, randomization to linezolid dose reduction strategy may occur at W16 or earlier (concomitant to dose reduction decision).

1. Test with abnormal result at screening visit may be repeated within screening window.
2. ALT/AST tests to be symptom driven after Week 47.
3. Optionally, documented results of TSH (performed less than 4 weeks prior to screening visit date) may substitute for screening/baseline test. Investigators are encouraged to repeat TSH if they deem it useful for patient management or to have a reliable pre-treatment result.
4. Optionally, documented results of HbA1c (performed less than 3 months prior to screening visit date) may substitute for screening/baseline test. Investigators are encouraged to repeat HbA1c if they deem it useful for patient management or to have a reliable pre-treatment result.
5. HbA1c test (or 2 fasting blood glucose test if HbA1c cannot be done) to be repeated every 6 months only if abnormal at screening/baseline.
6. HIV serology will be offered, unless patient is known to be positive or a documented negative result is available from less than one month before screening visit date.
7. CD4 count and HIV viral load will be performed if the patient is known or found to be HIV positive. Optionally, documented results of CD4 count (performed less than six months prior to screening visit date) and viral load (performed less than 4 weeks prior to screening visit date) may substitute for screening test. Investigators are encouraged to repeat CD4 count/viral load if they deem it useful for patient management or to have a reliable pre-treatment result.
8. Complete only if not done at screening.
9. CD4 count and HIV viral load to be monitored every 6 months in HIV-positive patients.
10. Serum pregnancy test at baseline visit may be repeated if the blood specimen was not drawn within 72 hours prior to treatment start.
11. When required, pregnancy tests may be performed on urine or serum samples after baseline visit until the end of study treatment.
12. Rapid molecular test results for RIF and FQ resistance, performed at designated study lab not older than 3 weeks prior to screening visit date have to be available before randomization.
13. Done locally on first positive culture or any culture reversion at or after Week 16.
14. Done at ITM on first positive culture or any culture reversion at or after Week 16 and on corresponding screening/baseline strain.
15. Store the screening/baseline sample and isolate from the corresponding culture. When possible, store samples and isolates from subsequent positive cultures for study (and future research) purposes).

16. Not necessary if previous adequate imaging results within 3 weeks prior to baseline visit date are available.
17. May be indicated, based on previous results available.

C. Only for participants still on prescription.

#### Appendix 2: Statement of Compliance

This trial will be conducted with human subjects oversight from the following Institutional Review Boards (IRBs):

- Harvard Medical School Institutional Review Board
- Médecins Sans Frontières Ethics Review Board (ERB)
- Institute of Tropical Medicine Institutional Review Board
- Interactive Research and Development Institutional Review Board
- Ethics Committees of the Hospital Nacional Hipólito Unanue, Peru
- Ethics Committee of the National Center for Tuberculosis and Lung Diseases of Georgia
- National Health Research Ethics Committee of Ministry of Health of Lesotho
- Central Bioethics Committee of the Ministry of Healthcare and Wealth of the Republic of Kazakhstan
- University of Cape Town Human Research Ethics Committee, South Africa
- National Bioethics Committee – Pakistan Health Research Council (PHRC), Pakistan
- Ethics Committee of the National AIDS Research Institute (NARI), Indian Council of Medical Research, Pune, India.
- Institutional Ethics Committee, Foundation for Medical Research Mumbai, India.

The study will be carried out in accordance with the following guidelines and regulations:

- Compliance with the International Conference on Harmonization (ICH) E6: Good Clinical Practice (GCP): Consolidated Guideline and the applicable regulatory requirements.
- UNITAID Terms of Award
- Regulations of Clinical Trials in all implementing countries

All key personnel (all individuals responsible for the design and conduct of this study) have completed Human Research Subjects Protection Training. Study implementation is also guided by Good Participatory Practices in TB Trials. [1]

#### Appendix 3: Institutions / Site Principal Investigators and Study Sites

| | | |
|---|---|---|
| **Peru:**<br>Dr. Leonid Lecca<br>Socios En Salud Sucursal Peru<br>Jr. Emeterio Perez, 310, San Martin de Porres, Lima 31, Peru<br><br>Dr. Dante Elmo Vargas Vásquez<br>Centro de Investigación del Hospital Nacional Hipólito Unanue<br>Av Cesar Vallejo 1390, El Augustino, Lima 10, Peru<br><br>Dr. Epifanio Sánchez Garavito<br>Centro de Investigación de Enfermedades Neumológicas del Hospital Nacional Sergio Bernales<br>Av. Túpac Amaru Km 14.5, Comas, Lima 7, Peru | **Kazakhstan:**<br>Dr. Elmira Berikova<br>National Scientific Center of Phthisiopulmonology of the MOH of the Republic of Kazakhstan<br>5 Bekhozhin Street, Almaty, Kazakhstan 050010<br><br>Dr. Dakenova Zhanna<br>State Municipal Enterprise on the right of economic management “City Centre of Phthisiopulmonology”<br>of Nur-sultan city’s administration.<br>Zheleznodorozhniy residential area,<br>A1 street, Building 5<br>Nur-Sultan city<br>020000, Kazakhstan | **India:**<br>Dr. Samiran Panda<br>ICMR-National AIDS Research Center<br>Plot No. 73, G-block, MIDC, Bhosari, Pune, 411026<br>Maharashtra, India<br><br>Dr. Chetankumar Jain<br>MSF clinic, 3rd floor, Pitruchaaya, Sanghavi Corporate Park, Off B.K.S.D. Marg, Govandi East, Mumbai 400 088, India<br><br>Dr. Sandip Patil<br>ICMR-National AIDS Research Center<br>Site: Aundh Chest Hospital,<br>New Sangavi, Pune<br><br>Dr. Padmaja Jogewar<br>Joint Director (NCD), Directorate of Health Services, Maharashtra, India |
| **Lesotho:**<br>Dr Kunda Kwabisha Mikanda<br>Botshabelo MDR-TB Hospital<br>Botshabelo, Next to NHTC, Maseru<br>Private Bag A391<br>Maseru, 0100, Lesotho | **Pakistan:**<br>Dr. Naseem Salahuddin<br>The Indus Hospital<br>Plot C-76, Sector 31/5<br>Opposite Korangi Crossing Road<br>Karachi, Pakistan<br><br>Dr. Naseem Salahuddin<br>Institute of Chest Disease, Kotri, District Jamshoro, Sindh, Pakistan<br><br>Dr. Naseem Salahuddin PMDT site of Civil Hospital Mirpurkhas.<br>Mirpur Khas, Sindh, Pakistan | |
| **Georgia:**<br>Dr. Nana Kiria<br>National Center for Tuberculosis and Lung Diseases<br>Adjara (Achara) str. N8,<br>Tbilisi 0101, Georgia | **South Africa:**<br>Dr. Sean Wasserman<br>Corner of Lansdowne & Basil Zondi Road,<br>Khayelitsha, Cape Town,<br>7784 South Africa | |
