## Supplementary material for "Nine-month, all-oral regimens for rifampin-resistant tuberculosis": endTB SAP v2.0

### Statistical Analysis Plan

#### endTB (Evaluating Newly approved Drugs for multidrug-resistant TB)

Protocol Number: NCT02754765 (Clinicaltrials.gov)

| **SAP** | **Effective Date**: 23-Jun-2023 |
|---|---|
| **Version Number and Date**: 2.0, 22 June 2023 | |

| **Revision** | |
|---|---|
| 1.0 Date 04-Jan-2021 | Initial version |
| 2.0 Date 22-Jun-2023 | Adaptations to reflect protocol version 3.6 (20Jul2022)<br>Update definition of study populations<br>Reorganization of endpoints and analyses' numbering to match objectives (8)<br>Addition of definitions: loss to follow-up (9.3), recurrence, relpase and reinfection (9.4), SAE (9.9)<br>Update of Culture results (9.5), TTP (9.7), adherence (9.8) definitions<br>Precision added on statistical analyses used: risk difference using a binomial regression model, adjusted analyses (10.3.1,10.3.2, 10.3.3, 10.4.4)<br>Multiple imputation of missing values moved to sensitivity analyses (10.3).3.<br>Addition of subgroup analysis section (10.3.1.1)<br>Clarification on sensitivity analyses (10.3.1.2, 10.3.2, 10.3.3)<br>Word change: recurrence instead of relapse<br>Addition of Appendix 12.2 AE categories |

| **Name and function** | **Date** (DD-MMM-YYYY) | **Signature** |
|---|---|---|
| *Authors* | | |
| Maelenn Gouillou, Biostatistician | | |
| *Reviewed by* | | |
| Maryline Bonnet, Co-Investigator | | |
| Elisabeth Baudin, Co-Investigator, Biostatistician | | |
| *Approved by* | | |
| Lorenzo Guglielmetti, Co-Principal Investigator | | |
| Carole Mitnick, Co-Principal Investigator | | |

*LIST OF ABBREVIATIONSs* 4
1. *INTRODUCTION* 5
2. *STUDY OBJECTIVES* 5
3. *STUDY DESIGN* 5
4. *SAMPLE SIZE CALCULATION* 6
5. *TREATMENT GROUPS* 6
6. *STUDY PLAN* 7
7. *STUDY ANALYSIS POPULATIONS* 11
**7.1 Efficacy populations** 11
7.1.1 Modified Intent To Treat population 11
7.1.2 Per Protocol Population 11
7.1.3 Other efficacy Populations 11
**7.2 Safety population** 12
8. *ASSESSEMENT OF OBJECTIVES* 12
**8.1 Efficacy endpoints** 12
8.1.1. Primary efficacy endpoint 12
8.1.2 Secondary efficacy endpoints 13
8.1.3 Exploratory efficacy endpoints 15
**8.2 Safety endpoints** 15
**8.3 Summary** 16
9. *DEFINITIONS* 18
**9.1 Scheduled visits, visit window definition, and unscheduled visits** 18
**9.2 Data Handling Convention for Week 73 and 104 outcomes** 18
**9.3 Definition of loss to follow-up** 18
**9.4 Definition of recurrence, relapse, and reinfection** 19
**9.5 Definition of a culture result** 19
**9.6 Definition of an AFB smear result** 19
**9.7 Definition of time to positivity (TTP) in MGIT** 19
**9.8 Definition of adherence** 20
**9.9 Definition of an adverse event** 20
**9.10 Definition of a serious adverse event** 21
**9.11 Definition of an adverse event of special interest** 21
**9.12 Definition of non-TB microbiology laboratory variables** 21
**9.13 Definition of clinical signs, symptoms, evaluations** 21
**9.14 Definition of severe linezolid-related toxicity** 22
*10.STATISTICAL METHODS AND DATA ANALYSIS* 22
**10.1 General rules** 22
**10.2 Data summaries/description** 22
10.2.1 Patient/Participant disposition 22
10.2.2 Demographic and baseline characteristics 23
10.2.3 Regimen characteristics (control arm) 24
10.2.4 Adherence 24
**10.3 Efficacy analyses** 24
10.3.1 Analysis of the primary efficacy endpoint (PO) 24
10.3.2 Analysis of the secondary efficacy endpoints 26
10.3.3 Analysis of the exploratory efficacy endpoints 27
**10.4 Safety analyses** 28
10.4.1 Analysis of adverse events 29
10.4.2 Analysis of laboratory variables 29
10.4.3 Analyses of clinical signs and assessments 29
10.4.4 Analysis of the exploratory safety endpoints 30

**10.5 Major protocol deviations 30**

***11.INTERIM ANALYSIS 30***

***12.APPENDIX*** *Error! Bookmark not defined.*

**12.1 Study flow diagram 31**

**12.2 AE categories 32**

#### LIST OF ABBREVIATIONS

| | |
|---|---|
| AE | Adverse Event |
| AESI | Adverse Event of Special Interest |
| ALT | Alanine Aminotransferase |
| AST | Aspartate Aminotransferase |
| Be | Bedaquiline |
| BMI | Body Mass Index |
| C | Clofazimine |
| CA++ | Calcium |
| CBC | Complete Blood Count |
| CD4 | CD4 Cell Count |
| CI | Confidence Interval |
| De | Delamanid |
| DR | Drug-resistant |
| DS | Drug-Susceptible |
| DSMB | Data and Safety Monitoring Board |
| DST | Drug Susceptibility Test |
| ECG | Electrocardiogram |
| ECOG | Eastern Cooperative Oncology Group |
| EEO | Exploratory Efficacy Objective |
| ESO | Exploratory Safety Objective |
| endTB | Study name |
| EVG | Event Validation Group |
| FQ | Fluoroquinolones |
| HbA1c | Hemoglobin A1c |
| HBc | Hepatitis B Core |
| HBsAg | Hepatitis B Surface Antigen |
| HBV | Hepatitis B virus |
| HCV | Hepatitis C virus |
| HCVAb | Hepatitis C virus antibody |
| HIV | Human Immunodeficiency Virus |
| ITT | Intent-to-treat |
| IWRS | Interactive Randomization System |
| IP | Investigational Product |
| K+ | Potassium |
| Le | Levofloxacin |
| Li | Linezolid |
| LJ | Löwenstein Jensen |
| LTFU | Loss to Follow Up |
| MDR-TB | Multidrug-resistant Tuberculosis |
| MedDRA | Medical Dictionary for Regulatory Activities |
| MG++ | Magnesium |
| MGIT | Mycobacterial Growth Indicator Tube |
| mITT | Modified Intent-to-Treat |
| Mo | Moxyfloxacin |
| MTB | Mycobacterium Tuberculosis |
| PI | Principal Investigator |
| PO | Primary Objective |
| PP | Per-protocol |
| PT | Preferred Term |
| QTcF | QT Interval Heart-Rate-Corrected by Fridericia Formula |
| RIF | Rifampicin |
| SAE | Serious Adverse Event |
| SAP | Statistical Analysis Plan |
| SCC | Sputum culture conversion? Or stable culture conversion? |
| SD | Standard Deviation |
| SEO | Secondary Efficacy Objective |
| SSO | Secondary Safety Objective |
| SOC | System Organ Class |
| SOP | Standard operating procedure |
| TB | Tuberculosis |
| TSH | Thyroid-stimulating Hormone |
| TTP | Time to (culture) positivity |
| WHO | World Health Organization |
| Z | Pyrazinamide |

#### 1. INTRODUCTION

This standard methodology document, the statistical analysis plan, explains the rules and conventions to be used in the presentation and analysis of efficacy and safety results for the clinical study “endTB, evaluating newly approved drugs for multidrug resistant TB”. It describes the data and endpoints to be summarized and analyzed, including specifics of the statistical analyses to be performed. SAP version 2.0 was based on protocol version 3.6 dated July 20, 2022. Any changes made after the approval of this document will be recorded in the cover page and in the appropriate section of the clinical study report. Any post hoc analyses that are performed will be detailed in the same section of the clinical study report.

#### 2. STUDY OBJECTIVES

The primary objective of the study is to assess whether the efficacy of experimental regimens at Week 73 is non-inferior to that of the control (PO).

The secondary objectives are:

1. To compare the efficacy of experimental regimens at Week 104 to that of the control (SEO1).
2. To compare the frequency of and time to early treatment response (culture conversion) in experimental regimens to that of the control (SEO2).
3. To compare the efficacy of experimental regimens at Week 39 to that of the control (SEO3).
4. To compare, at Week 73 and Week 104, the proportion of participants who experienced failure or recurrence in the experimental arms to that in the control arm (SEO4).
5. To compare, at Week 73 and Week 104, the proportion of participants who died of any cause in the experimental arms to that in the control arm (SSO5).
6. To compare, at Week 73 and Week 104, the proportion of participants who experience grade 3 or higher AEs or SAEs of any grade in the experimental arms to that in the control arm (SSO6).
7. To compare, at Weeks 73 and 104, the proportion of participants who experience AESIs in experimental regimens to that in the control arm (SSO7).

The exploratory objectives are:

1. To compare efficacy and safety endpoints across experimental arms, e.g., arms containing bedaquiline vs. delamanid (regimen 2 vs. 4); arms containing clofazimine & levofloxacin vs. arms containing moxifloxacin (regimen 2 vs. 1); arms containing linezolid vs. arms containing the same drugs and no linezolid (regimen 4 vs. 5); arms containing delamanid vs. arms containing the same drugs and no delamanid (regimen 1 vs. 3); arms containing bedaquiline and delamanid vs. arms containing one of the drugs and clofazimine (regimen 3 vs. 4 and regimen 3 vs. 2) (EEO1, ESO2).
2. To evaluate conversion endpoints as potential surrogate markers for unfavorable outcome (EEO3).
3. To compare the efficacy of regimens between participants with strains that are resistant to pyrazinamide and participants with strains that are sensitive (EEO4).
4. To estimate the frequency of drug resistance amplification among treatment failures and relapses occurring by Week 104 (EEO5).
5. To compare treatment adherence in the experimental arms to that in the control and across regimens (EEO6).
6. To compare the frequency of and time to severe linezolid-related toxicity between linezolid dose-reduction strategies (ESO7).
7. To compare, at Week 73 and Week 104, efficacy endpoints across linezolid dose-reduction strategies: 300 mg daily or 600 mg thrice weekly (EEO8).

#### 3. STUDY DESIGN

This is a Phase III, randomized, controlled, open-label, non-inferiority, multi-country trial, evaluating the efficacy of new combination regimens for treatment of MDR-TB. The study enrolls in parallel across 5 experimental and 1 standard-of-care control arms. Randomization is outcome adapted using Bayesian interim analysis of efficacy endpoints and centralized, using an interactive randomization system (IWRS). Trial participation in all arms is at least until Week 73 and up to Week 104. Study follow-up ends after the scheduled Week 73 visit for the last participant randomized (hybrid follow-up). In the experimental arms, treatment is for 39 weeks and post-treatment follow up for up to an additional 65 weeks (as defined above). In the control arm, treatment is delivered according to local practice (and WHO guidelines); duration is variable, typically lasting approximately 86 weeks for the conventional regimen and 39 to 52 weeks for the standardized shorter regimen.

Participants randomized to endTB regimens 1-4 (all linezolid-containing experimental arms) had undergone linezolid dose-reduction either at Week 16 or after a linezolid-related AE requiring dose reduction, whichever was earlier. The starting dose of linezolid was 600 mg once daily. The secondary, fixed, balanced randomization stratified by site reduced the dose to 300 mg once daily or 600 mg thrice weekly. Participants received the reduced linezolid dose until the end of the experimental regimen or until linezolid was permanently discontinued for toxicity.

#### 4. SAMPLE SIZE CALCULATION

The sample size was estimated through simulations that considered the following parameters: early efficacy response at Week 8, end of treatment response frequency at Week 39, primary outcome frequency at Week 73 for each experimental arm and the control arm, expected number of effective arms among the five experimental, the type I error, and the non-inferiority margin[1].

We assumed an early efficacy response proportion (defined as the proportion of participants having an 8-week culture negativity) of 40% in the experimental arms, compared to the control of 30%. Sample size simulations used estimates of 75% favorable outcome at 73 weeks in the non-inferior arms, resulting from two response rate scenarios: 75% favorable outcomes at 73 weeks after no relapse and after 10% relapse between 39 and 73 weeks. Favorable outcome frequency in the control arm was assumed to be 5% lower than in non-inferior experimental arms, 70% at 73 weeks and 70% or 80% at 39 weeks. Simulations varied the number of expected non-inferior arms from 1 to 3 among the 5 experimental arms. Also, simulations explored non-inferiority margins of 10 and 12%. The type I error was set at 2.5% (one-sided).

With a 12% non-inferiority margin, a sample size of 750 randomized participants provides power greater than 80% to demonstrate the non-inferiority of at least 1 (and up to 3) experimental regimens in the modified intent-to-treat (mITT) and up to 2 in the per-protocol (PP) populations. This assumes a proportion of favorable outcome at week 73 of 75% in the experimental regimens and 70% in the control, with up to 10% of relapse between 39 and 73 weeks. The analysis populations are described in section 7. Our simulations assumed a loss of 11% of subjects from the randomized population to the mITT population and an additional loss of 10% from the mITT to the PP populations.

*Exploratory analysis of toxicity related to linezolid dose reduction*

For the secondary randomization of linezolid dose-reduction strategy, we assumed a sample size of 280 participants randomized to endTB regimens 1-4, which is a conservative estimate representing the absolute minimum from prior simulations. We assumed that the proportion of AEs observed in the 300 mg daily arm would be 69% as previously reported in the only other clinical trial to investigate the 300 mg dose,[2] and 52% for the 600 mg thrice weekly group. The latter estimate is extrapolated, conservatively, from a meta-analysis among patients receiving linezolid doses lower than 600 mg daily. It exceeds the upper limit of the 95% CI reported for AEs in that study: 34% (95% CI, 23-46%).[3] Based on these effect sizes, and assuming 10% of participants are not assessable, a time-to-event analysis with a sample size of n = 280 participants randomized 1:1 to the two linezolid dosing strategies would have 80% power to detect a hazard ratio of 1.43 for severe linezolid-related toxicity with 300 mg daily compared to 600 mg thrice weekly in a two-sided, two-sample log-rank test with 5% significance.

#### 5. TREATMENT GROUPS

The control regimen uses anti-TB drugs according to local practice and consistent with WHO guidance. Changes in control regimen guidance over time will be described, with the list of changes, date each started, affected countries, and implications for analysis.

The 5 experimental regimens are a combination of marketed anti-TB drugs. Each individual anti-TB drug in the experimental regimens is described in Table 1:

**Table 1: Experimental endTB regimen**

| Trial Regimens | Bedaquiline | Delamanid | Clofazimine | Linezolid | Fluoroquinolone | Pyrazinamide |
|---|---|---|---|---|---|---|
| **endTB 1 BeLiMoZ** | Be | | | Li | Mo | Z |
| **endTB 2 BeCLiLeZ** | Be | | C | Li | Le | Z |
| **endTB 3 BeDeLiLeZ** | Be | De | | Li | Le | Z |
| **endTB 4 DeCLiLeZ** | | De | C | Li | Le | Z |
| **endTB 5 DeCMoZ** | | De | C | | Mo | Z |
| **endTB 6 Control** | Standard of care control, composed according to WHO Guidelines, including the possible use of De or Be. | | | | | |

Fluoroquinolones = Mo: moxifloxacin; Le: levofloxacin.

Treatment duration in the experimental groups is 39 weeks and up to 104 weeks in the control group.

#### 6. STUDY PLAN

Table 2 is the Schedule of Events, which details the timing and content of all assessments performed during the study.

**Table 2: Schedule of Events**

| | | Treatment | | | | | | | | | | | | | | | | | | | | Follow-Up | | | | | | | | | | | | |
|---|---|---|---|---|---|---|---|---|---|---|---|---|---|---|---|---|---|---|---|---|---|---|---|---|---|---|---|---|---|---|---|---|---|---|
| | Screening | Baseline | W1 | W2 | W3 | W4 | W5 | W6 | W7 | W8 | W9 | W 10 | W 11 | W 12 | W 16 | W 20 | W 24 | W 28 | W 32 | W 36 | W39 | W43 | W47 | W53 | W59 | W65 | W73 | W81 | W89 | W97 | W 104 | Sub total (minimal count) | Early Termination | Post-termination follow-up |
| Visit | 1 | 2 | 3 | 4 | 5 | 6 | 7 | 8 | 9 | 10 | 11 | 12 | 13 | 14 | 15 | 16 | 17 | 18 | 19 | 20 | 21 | 22 | 23 | 24 | 25 | 26 | 27 | 28 | 29 | 30 | 31 | | | |
| Window Period (+/- days) | | | +/-2 | +/-2 | +/-2 | +/-2 | +/- 2 | +/-2 | +/-2 | +/- 2[#] | +/-2 | +/-2 | +/-2 | +/-2 | +/-7 | +/-7 | +/-7 | +/-7 | +/-7 | +/-7 | +/-14 | +/- 14 | +/- 14 | +/- 14 | +/- 14 | +/- 14 | +/- 30 | +/- 14 | +/- 14 | +/- 14 | +/- 30 | | | |
| Eligibility | | | | | | | | | | | | | | | | | | | | | | | | | | | | | | | | | | |
| Subject Consent | X | X | | | | | | | | | | | | | | | | | | | | | | | | | | | | | | 2 | | |
| Demographics | X | X | | | | | | | | | | | | | | | | | | | | | | | | | | | | | | 2 | | |
| Medical History | X | | | | | | | | | | | | | | | | | | | | | | | | | | | | | | | 1 | | |
| Inclusion/ Exclusion | X | X | | | | | | | | | | | | | | | | | | | | | | | | | | | | | | 2 | | |
| Clinical Evaluation | | | | | | | | | | | | | | | | | | | | | | | | | | | | | | | | | | |
| Vital Signs | X | X | X | X | X | X | X | X | X | X | X | X | X | X | X | X | X | X | X | X | X | X | X | X | X | X | X | X | X | X | X | 31 | X | X |
| Interval Medical History | | X | X | X | X | X | X | X | X | X | X | X | X | X | X | X | X | X | X | X | X | X | X | X | X | X | X | X | X | X | X | 30 | X | X |
| Physical Exam | X | X | X | X | X | X | X | X | X | X | X | X | X | X | X | X | X | X | X | X | X | X | X | X | X | X | X | X | X | X | X | 31 | X | X |
| TB Symptom Assessment | | X | X | X | X | X | X | X | X | X | X | X | X | X | X | X | X | X | X | X | X | X | X | X | X | X | X | X | X | X | X | 30 | X | X |
| ECOG Assessment | | X | | | | | | | | | | | | | | | | | | | X | | | | | | X | | | | X | 4 | X | *X* |
| Neurologic Exam | | X | | | | X | | | | X | | | | X | X | X | X | X | X | X | X | X | X | | | | X | | | | X | 15 | X | |
| Ophthalmologic Exam | | X | | | | X | | | | X | | | | X | X | X | X | X | X | X | X | X | X | | | | X | | | | X | 15 | X | |
| Mental Health Assessment | | X | | | | | | | | | | | | | | | | | | | | | | | | | X | | | | X | 3 | X | |
| Treatment | | | | | | | | | | | | | | | | | | | | | | | | | | | | | | | | | | |
| Randomization | | X* | | | | | | | | | | | | | X** | | | | | | | | | | | | | | | | | (1) | | |
| Treatment Compliance | | | X | X | X | X | X | X | X | X | X | X | X | X | X | X | X | X | X | X | X | X[C] | X[C] | X[C] | X[C] | X[C] | X[C] | X[C] | X[C] | X[C] | X[C] | 19 | X[C] | |
| Adherence Counseling | | X | X | X | X | X | X | X | X | X | X | X | X | X | X | X | X | X | X | X | X | X[C] | X[C] | X[C] | X[C] | X[C] | X[C] | X[C] | X[C] | X[C] | | 20 | | |
| Concomitant Medication Review | X | X | X | X | X | X | X | X | X | X | X | X | X | X | X | X | X | X | X | X | X | X | X | X | X | X | X | X | X | X | X | 31 | X | X |
| AE Review | | | X | X | X | X | X | X | X | X | X | X | X | X | X | X | X | X | X | X | X | X | X | X | X | X | X | X | X | X | X | 29 | X | X |
| Laboratory Testing | | | | | | | | | | | | | | | | | | | | | | | | | | | | | | | | | | |
| CBC | X[1] | | | | | X | | | | X | | | | X | X | X | X | X | X | X | X | X | X | | | | | | | | X | 14 | X | |
| Total Ca++, K+, Mg++ | X[1] | | | | | X | | | | X | | | | X | X | X | X | X | X | X | X | X | X | | | | | | | | | 13 | X | |

| | Screening | Baseline | W1 | W2 | W3 | W4 | W5 | W6 | W7 | W8 | W 9 | W 10 | W 11 | W 12 | W 16 | W 20 | W 24 | W 28 | W 32 | W 36 | W39 | W 43 | W 47 | W 53 | W 59 | W 65 | W 73 | W 81 | W 89 | W 97 | W 104 | Sub total (minimal count) | Early Termination | Post-termination follow-up |
|---|---|---|---|---|---|---|---|---|---|---|---|---|---|---|---|---|---|---|---|---|---|---|---|---|---|---|---|---|---|---|---|---|---|---|
| Visit | 1 | 2 | 3 | 4 | 5 | 6 | 7 | 8 | 9 | 10 | 11 | 12 | 13 | 14 | 15 | 16 | 17 | 18 | 19 | 20 | 21 | 22 | 23 | 24 | 25 | 26 | 27 | 28 | 29 | 30 | 31 | | | |
| Window Period | | | +/-2 | +/-2 | +/-2 | +/-2 | +/-2 | +/-2 | +/-2 | +/-2[#] | +/-2 | +/-2 | +/-2 | +/-2 | +/-7 | +/-7 | +/-7 | +/-7 | +/-7 | +/-7 | +/-14 | +/-14 | +/-14 | +/-14 | +/-14 | +/-14 | +/-30 | +/-14 | +/-14 | +/-14 | +/-30 | | | |
| Creatinine | X[1] | | | | | X | | | | X | | | | X | X | X | X | X | X | X | X | X | X | | | | | | | | | 13 | X | |
| AST and ALT[2] | X[1] | | | | | X | | | | X | | | | X | X | X | X | X | X | X | X | X | X | | | | | | | | | 13 | X | |
| Total and Direct Bilirubin | X[1] | | | | | X | | | | X | | | | X | X | X | X | X | X | X | X | X | X | | | | | | | | | 13 | X | |
| Albumin | X[1] | | | | | | | | | | | | | | | | | | | | | | | | | | | | | | | 1 | | |
| TSH | X[3] | X[8] | | | | | | | | | | | | | | | X | | | | | | X | | | | | | | | | 3 | X | |
| HbA1C | X[4] | X[8] | | | | | | | | | | | | | | | X[5] | | | | | | X[5] | | | | X[5] | | | | | (1) | X[5, 17] | X[5, 17] |
| HIV test | X[6] | | | | | | | | | | | | | | | | | | | | | | | | | | | | | | | 1 | | |
| CD4 count and viral load (in HIV+) | X[7] | X[8] | | | | | | | | | | | | | | | X[9] | | | | | | X[9] | | | | X[9] | | | | | (0) | X[9,17] | X[9,17] |
| Hepatitis B serology ( anti-HBc total and HbsAg) | X | X[8] | | | | | | | | | | | | | | | | | | | | | | | | | | | | | | 1 | | |
| Hepatitis C Serology: HCVAb | X | X[8] | | | | | | | | | | | | | | | | | | | | | | | | | | | | | | 1 | | |
| Pregnancy test[11] | X | X[10] | | | | | | | | | | | | | | | | | | | | | | | | | | | | | | (2) | X | |
| **Sputum Specimen Testing** | | | | | | | | | | | | | | | | | | | | | | | | | | | | | | | | | | |
| Smear Microscopy | X | | | X | | X | | | | X | | | | X | X | X | X | X | X | X | X | X | X | X | X | X | X | X | X | X | X | 22 | X | X |
| Culture (LJ & MGIT) | X | | | X | | X | | | | X | | | | X | X | X | X | X | X | X | X | X | X | X | X | X | X | X | X | X | X | 22 | X | X |
| Rapid Molecular Test for RIF Resistance | X[12] | X[8] | | | | | | | | | | | | | | | | | | | | | | | | | | | | | | 1 | | |
| Rapid molecular test for FQ resistance | X[12] | X[8] | | | | | | | | | | | | | X[13] | | | | | | | | | | | | | | | | | (1) | | |
| DST (conventional 1st and 2nd line) | X[14] | | | | | | | | | | | | | | X[13,14] | | | | | | | | | | | | | | | | | (1) | X[17] | X[17] |
| DST (new drugs) | X[14] | | | | | | | | | | | | | | X[14] | | | | | | | | | | | | | | | | | (0) | X[17] | X[17] |
| Genotyping[14] | X | | | | | | | | | | | | | | X | | | | | | | | | | | | | | | | | (0) | X[17] | X[17] |
| Specimen Storage | X[15] | | | | | | | | | | | | | | X[15] | | | | | | | | | | | | | | | | | (1) | X | |
| **Special Assays or Procedures** | | | | | | | | | | | | | | | | | | | | | | | | | | | | | | | | | | |
| Audiometry | | X | | | | X | | | | X | | | | X | X | X | X | X | X | X | X | X | X | | | | X | | | | X | 15 | X | |
| ECG[1] | X[1] | X | X | X | X | X | X | X | X | X | X | X | X | X | X | X | X | X | X | X | X | X | X | X | | | X | | | | | 25 | X | |
| Chest X-ray | | X[16] | | | | | | | | X | | | | | | | | | | | X | | | | | | X | | | | X | 5 | X[17] | X[17] |

#. W8 visit window should occur within 8 weeks (+/-2 days) of study treatment initiation.

*: Randomization for treatment regimen assignment.

**: Only for participants receiving linezolid, randomization to linezolid dose reduction strategy may occur at W16 or earlier (concomitant to dose reduction decision).

1. Test with abnormal result at screening visit may be repeated within screening window.
2. ALT/AST tests to be symptom driven after Week 47.
3. Optionally, documented results of TSH (performed less than 4 weeks prior to screening visit date) may substitute for screening/baseline test. Investigators are encouraged to repeat TSH if they deem it useful for patient management or to have a reliable pre-treatment result.
4. Optionally, documented results of HbA1c (performed less than 3 months prior to screening visit date) may substitute for screening/baseline test. Investigators are encouraged to repeat HbA1c if they deem it useful for patient management or to have a reliable pre-treatment result.
5. HbA1c test (or 2 fasting blood glucose tests if HbA1c cannot be done) to be repeated every 6 months only if abnormal at screening/baseline.
6. HIV serology will be offered, unless patient is known to be positive or a documented negative result is available from less than one month before screening visit date.
7. CD4 count and HIV viral load will be performed if the patient is known or found to be HIV positive. Optionally, documented results of CD4 count (performed less than six months prior to screening visit date) and viral load (performed less than 4 weeks prior to screening visit date) may substitute for screening test. Investigators are encouraged to repeat CD4 count/viral load if they deem it useful for patient management or to have a reliable pre-treatment result.
8. Complete only if not done at screening.
9. CD4 count and HIV viral load to be monitored every 6 months in HIV-positive participants.
10. Serum pregnancy test at baseline visit may be repeated if the blood specimen was not drawn within 72 hours prior to treatment start.
11. When required, pregnancy tests may be performed on urine or serum samples after baseline visit until the end of study treatment.
12. Rapid molecular test results for RIF and FQ resistance, performed at designated study lab not older than 3 weeks prior to screening visit date have to be available before randomization.
13. Done locally on first positive culture or any culture reversion at or after Week 16.
14. Done at ITM on first positive culture or any culture reversion at or after Week 16 and on corresponding screening/baseline strain.
15. Store the screening/baseline sample and isolate from the corresponding culture. When possible, store samples and isolates from subsequent positive cultures for study (and future research) purposes).
16. Not necessary if previous adequate imaging results within 3 weeks prior to baseline visit date are available.
17. May be indicated, based on previous results available.

C. Only for participants still on prescription.

### 7. STUDY ANALYSIS POPULATIONS

#### 7.1 Efficacy populations

##### *7.1.1 Modified Intent To Treat population*

The modified intent-to-treat population (mITT) will contain all randomized participants with culture-positive, FQ-susceptible and rifampicin-resistant TB at baseline (or screening). FQ susceptibility and rifampicin resistance are diagnosed by validated rapid molecular test performed by a designated study laboratory. Exclusion from the mITT population will occur if any of the following occur:

1) Any DST results performed on any screening or baseline sample at the designated study lab indicate resistance to bedaquiline, clofazimine, delamanid, fluoroquinolone and/or linezolid
2) There is no positive sputum culture before randomization
3) There is no post baseline data (i.e. no study visits performed, no reported events occurred after randomization).

Participants in the mITT population will be analyzed in the arm to which they were randomly allocated by the IWRS (as randomized) regardless of received treatment.

##### *7.1.2 Per Protocol Population*

The per protocol (PP) population will include participants in the mITT population, except participants who, for reasons other than treatment failure or death, do not complete a protocol-adherent course of treatment. A protocol-adherent course of treatment contains 80% of expected doses within 120% of the nominal regimen duration. Receipt of either a prohibited concomitant medication or an IP not prescribed according to protocol for more than 7 days constitutes a non-protocol-adherent course of treatment and results in exclusion from the PP population. No other deviations resulted in exclusion from PP.

##### *7.1.3 Other efficacy Populations*

Additional efficacy populations will be defined for sensitivity analyses for the Week 73 and Week 104 endpoints. Use of these populations is specified in section 10.3:

1) <u>Assessable</u>: this population will include participants from the PP population whose outcomes were classified as favorable or unfavorable, excluding those whose outcomes were unfavorable because they were not assessable (per section 8.1). Also excluded from this population are participants who experienced voluntary withdrawal, loss to follow up (LTFU) as defined in section 9.3, or confirmed reinfection. Distinct populations will be created for analyses of Week 73 and Week 104 endpoints.

2) <u>All-culture mITT</u>: this population will comprise the mITT population plus participants with baseline culture-negative TB. Participants in the All-culture mITT population will be analyzed in the arm to which they were randomly allocated by the IWRS (as randomized) regardless of received treatment.

3) <u>All-DST mITT:</u> this population will comprise the mITT population plus participants who had screening/baseline DST results from the designated laboratory indicating resistance (using a test deemed to be reliable by the trial reference laboratory, ITM) to bedaquiline, clofazimine, delamanid, and/or linezolid. Participants in the All-DST mITT population will be analyzed in the arm to which they were randomly allocated by the IWRS (as randomized) regardless of received treatment.

##### 7.2 Safety population

The safety population will include all enrolled participants who received at least one dose of study treatment (exposed) of any drug. Safety analyses will be based on the observed treatment regimen received (as treated) regardless of randomization.

#### 8. ASSESSEMENT OF OBJECTIVES

Each endpoint references the abbreviation assigned to the corresponding objective above (in section 2).

##### 8.1 Efficacy endpoints

###### *8.1.1. Primary efficacy endpoint*

The primary efficacy endpoint used for the primary objective (PO) is a favorable outcome at Week 73.

A favorable outcome can only be established if all possible unfavorable outcomes are ruled out.
Unfavorable outcome is established by any of the following situations:

- Replacement or addition of one or more investigational drugs in an experimental arm or in the control arm if using the shortened regimen (failure);
- Replacement or addition of two or more investigational drugs in the control arm if using the conventional regimen (failure);[i]
- Initiation of a new MDR-TB treatment regimen after the end of the allocated study regimen and before Week 73 (recurrence);
- Death from any cause (death);
- At least one of the last two TB cultures, the latest being from a sputum sample collected between Week 65 and Week 73, is positive in the absence of evidence of laboratory cross contamination, as defined in section 9.5 (failure/recurrence);
- The most recent (i.e., last) TB culture result (from a sputum sample collected between Week 65 and Week 73) is negative; AND there is no other post-baseline culture result or the penultimate culture is positive due to laboratory cross contamination; and bacteriological, radiological or clinical evolution is unfavorable (failure/recurrence);
- There is no TB culture result from a sputum sample collected between Week 65 and Week 73 or it is positive due to laboratory cross contamination; AND the most recent (i.e., last) culture is negative; and bacteriological, radiological or clinical evolution is unfavorable or, the most recent (i.e. last) culture result is positive in the absence of laboratory cross contamination (failure/recurrence);
- There is no culture result from a sputum sample collected between Week 65 and Week 73 or it is positive due to laboratory cross contamination; AND there is no other post-baseline culture result or the most recent culture is positive due to laboratory cross contamination; or, the most recent culture is negative and bacteriological, radiological and clinical evolution is not assessable (not assessable);
- Previously classified as unfavorable in the present study. Prior assignment of unfavorable, due only to being not assessable at Week 39, is insufficient to establish unfavorable outcome at

[i] Addition or replacement of two or more drugs to the control arm in order to conform to a new WHO treatment guidance —rather than in response to emerging participant data — does not lead to establish an unfavourable outcome.

Week 73. If unfavorable was assigned at Week 39 because the outcome was not assessable at that timepoint, a new Week 73 outcome is assigned.

If an unfavorable outcome has not been assigned at week 73 and one of the following conditions is met, the outcome is favorable:

- The last two culture results are negative. These two cultures must be from sputum samples collected on separate visits, the latest between Week 65 and Week 73;
- The last culture result (from a sputum sample collected between Week 65 and Week 73) is negative; and either there is no other post-baseline culture result or the penultimate culture result is positive due to laboratory cross contamination; and bacteriological, radiological and clinical evolution is favorable;
- There is no culture result from a sputum sample collected between Week 65 and Week 73 or the result of that culture is positive due to laboratory cross contamination; and the most recent culture result is negative; and bacteriological, radiological and clinical evolution is favorable.

The Week 73 efficacy endpoint will not be determined from raw data at the time of analysis. It will have been previously assigned through the event-validation process (according to the above definitions and described in SOP SP-031-CT), validated, and entered into the study database. The validated outcome will be used for the analysis.

The primary efficacy endpoint will also be used for the exploratory objectives EEO1, EEO3, EEO4 and EEO8.

###### *8.1.2 Secondary efficacy endpoints*

The main secondary efficacy endpoints are:

1. Favorable outcome at Week 104 (used for SEO1).

A favorable outcome can only be established if all possible unfavorable outcomes are ruled out.
Unfavorable outcome is established by any of the following situations:

- Replacement or addition of one or more investigational drugs in the experimental arm or in the control arm if using the shortened regimen (failure);
- Replacement or addition of two or more investigational drugs in the control arm if using the conventional regimen (failure);
- Initiation of a new MDR-TB treatment regimen after the end of the allocated study regimen and before Week 104 (recurrence);
- Death from any cause (death);
- At least one of the last two cultures, the latest being from a sputum sample collected between Week 97 and Week 104, is positive in the absence of evidence of laboratory cross contamination (failure/recurrence);
- The last culture result (from a sputum sample collected between Week 97 and Week 104) is negative and there is no other post-baseline culture result or the penultimate culture is positive due to laboratory cross contamination; and bacteriological, radiological or clinical evolution is unfavorable (failure/recurrence);
- There is no culture result from a sputum sample collected between Week 97 and Week 104 or it is positive due to laboratory cross contamination and the most recent culture is negative; and bacteriological, radiological or clinical evolution is unfavorable (failure/recurrence);
- There is no culture result from a sputum sample collected between Week 97 and Week 104 or it is positive due to laboratory cross contamination and there is no other post-

baseline culture or it is positive or the most recent culture is negative and bacteriological, radiological and clinical evolution is not assessable (not assessable);
- Previously classified as unfavorable in the present study[ii] ;
- Lost to follow-up, as defined in section 9.3.

If an unfavorable outcome has not been assigned and one of the following conditions is met, the outcome is favorable:
- The last two cultures are negative. These two cultures must be from: sputum samples collected on separate visits, the latest between Week 97 and Week 104;
- The last culture result (from a sputum sample collected between Week 97 and Week 104) is negative; and either there is no other post-baseline culture result or the penultimate culture result is positive due to laboratory cross contamination; and bacteriological, radiological and clinical evolution is favorable;
- There is no culture result from a sputum sample collected between Week 97 and Week 104 or the result of that culture is positive due to laboratory cross contamination; and the most recent culture result is negative; and bacteriological, radiological and clinical evolution is favorable.

The Week 104 efficacy endpoint will not be determined from raw data at the time of analysis. It will have been previously assigned through the event-validation process (according to the above definitions and described in SOP SP-031-CT), validated, and entered into the study database. The validated outcome will be used for the analysis.

Favorable outcome at Week 104 will also be used for the exploratory objective EEO8.

2. Culture conversion endpoints (used for SEO2):
   a. Initial sputum culture conversion (SCC) by Week 8. This is defined as 2 consecutive negative sputum culture results from specimens collected at 2 different visits, the first being at or before Week 8. If there is a single missing or contaminated (not AFB/contaminated) culture between 2 negatives, the definition of conversion is still met.
   b. Time to initial culture conversion. This is defined as the interval between the date of study treatment initiation and the date of collection of sputum resulting in the first of at least 2 consecutive negative cultures (as defined above). If there is a missing or contaminated (not AFB/contaminated) culture between 2 negatives, the definition of conversion is still met.
   c. Change in time to MGIT culture positivity (TTP) from baseline through 8 weeks (defined in section 9.7).
   Participants who discontinue study participation or die before conversion remain in the denominator of those eligible for conversion and count as not having converted. Participants with only contaminated or missing results will be counted as not having converted.

3. Favorable outcome at Week 39 (used for SEO3).
A favorable outcome can only be established if all possible unfavorable outcomes are ruled out. Unfavorable outcome is established by any of the following situations:
- In the experimental arm or in the control arm if using the shortened regimen, addition or replacement of one or more drugs;
- In the control arm, if using the conventional regimen, addition or replacement of two or more

[ii] Prior assignment of unfavorable, due only to being not assessable at Week 73 is insufficient to establish unfavorable outcome at Week 104. If unfavorable was assigned at Week 73 because the outcome was not assessable at that timepoint, a new Week 104 outcome is assigned.

drugs;
- Death from any cause;
- At least one culture result (from a sample collected between Week 36 and Week 39) is positive;
- The outcome is not assessable because the last available culture result is from a sample collected before Week 36.

If an unfavorable outcome has not been assigned and all culture results from samples collected between Week 36 and Week 39 are negative, the outcome is favorable..

The Week 39 efficacy endpoint will not be calculated from raw data at the time of analysis. It will have been previously assigned through the event-validation process (according to the above definitions and described in SOP SP-031-CT), validated, and entered into the study database. The validated outcome will be used for the analysis.

4. Treatment failure or recurrence (used for SEO4):
   a. at Week 73 (as defined in list of unfavorable outcomes in section 8.1.1).
   b. at Week 104 (as defined in list of unfavorable outcomes in section 8.1.2).

##### *8.1.3 Exploratory efficacy endpoints*

The additional exploratory efficacy endpoints are:
- Drug resistance amplification by week 104. This is defined as change in any DST result from susceptible on baseline DST to resistant on any post-baseline DST, on identical strain[iii]: used for EEO5.
- Treatment adherence of ≥80% as defined in section 9.8: used for EEO6.

#### 8.2 Safety endpoints

1. Death from any cause (SSO5).

2. Occurrence of (SSO6 and ESO2):
   a. AE of grade 3 or higher as defined in section 9.9 by Week 73.
   b. AE of grade 3 or higher as defined in section 9.9 by Week 104.
   c. SAE as defined in section 9.10 by Week 73.
   d. SAE as defined in section 9.10 by Week 104.
   e. AE of grade 3 or higher or SAE by Week 73.
   f. AE of grade 3 or higher or SAE by Week 104.

3. Occurrence of (SSO7):
   a. AESI as defined in section 9.11 by Week 73.
   b. AESI as defined in section 9.11 by Week 104.

4. Severe linezolid-related toxicity (as described in section 9.14) (ESO7):
   a. Occurrence of severe linezolid-related toxicity
   b. Time to first severe linezolid-related toxicity.

[iii] Data related to drug resistance amplification and relapse outcomes will be provided by ITM.

#### 8.3 Summary

| N° | Objective | Endpoint |
|---|---|---|
| | **Primary Efficacy** | |
| **PO** | To assess whether the efficacy of experimental regimens at Week 73 is non-inferior to that of the control | Favorable outcome at Week 73 |
| | **Secondary efficacy** | |
| **SEO1** | To compare the efficacy of experimental regimens at Week 104 to that of the control. | Favorable outcome at Week 104 |
| **SEO2** | To compare the frequency of and time to early treatment response (culture conversion) in experimental regimens to that of the control. | Culture conversion endpoints:<br>a. Initial sputum culture conversion (SCC) by Week 8.<br>b. Time to initial culture conversion<br>c. Change in time to MGIT culture positivity (TTP) from baseline through 8 weeks. |
| **SEO3** | To compare the efficacy of experimental regimens at Week 39 to that of the control. | Favorable outcome at Week 39 |
| **SEO4** | To compare, at Week 73 and Week 104, the proportion of participants who experienced failure or recurrence in the experimental arms to that in the control arm | Treatment failure or recurrence:<br>a. at Week 73<br>b. at Week 104 |
| | **Exploratory efficacy** | |
| **EEO1** | To compare efficacy endpoints across experimental arms | Favorable outcome at Week 73 |
| **EEO3** | To evaluate conversion endpoints as potential surrogate markers for unfavorable outcome | Favorable outcome at Week 73<br>Culture conversion-related endpoints for unfavorable outcome at week 73 |
| **EEO4** | To compare the efficacy of regimens between participants with strains that are resistant to pyrazinamide and participants with strains that are sensitive | Favorable outcome at Week 73 |
| **EEO5** | To estimate the frequency of drug resistance amplification among treatment failures and relapses occurring by Week 104 | Drug resistance amplification by week 104 |
| **EEO6** | To compare treatment adherence in the experimental arms to that in the control and across regimens | Treatment adherence of ≥80% |
| **EEO8** | To compare, at Week 73 and Week 104, efficacy endpoints across linezolid dose-reduction strategies: 300 mg daily or 600 mg thrice weekly | Favorable outcome at Week 73<br>Favorable outcome at Week 104 |
| | **Safety** | |
| **SSO5** | To compare, at Week 73 and Week 104, the proportion of participants who died of any cause in the experimental arms to that in the control arm | Death from any cause |

| | | |
|---|---|---|
| **SSO6** | To compare, at Week 73 and Week 104, the proportion of participants who experience grade 3 or higher AEs or SAEs of any grade in the experimental arms to that in the control arm | Occurrence of :<br>a. AE of grade 3 or higher by Week 73.<br>b. AE of grade 3 or higher by Week 104.<br>c. SAE by Week 73.<br>d. SAE by Week 104.<br>e. AE of grade 3 or higher or SAE by Week 73.<br>f. AE of grade 3 or higher or SAE by Week 104 |
| **SSO7** | To compare, at Weeks 73 and 104, the proportion of participants who experience AESIs in experimental regimens to that in the control arm | Occurrence of:<br>a. AESI by Week 73.<br>b. AESI by Week 104. |
| **ESO2** | To compare safety endpoints across experimental arms | Occurrence of :<br>a. AE of grade 3 or higher by Week 73.<br>b. AE of grade 3 or higher by Week 104.<br>c. SAE as defined by Week 73.<br>d. SAE as defined by Week 104.<br>e. AE of grade 3 or higher or SAE by Week 73.<br>f. AE of grade 3 or higher or SAE by Week 104 |
| **ESO7** | To compare the frequency of and time to severe linezolid-related toxicity between linezolid dose-reduction strategies | Severe linezolid-related toxicity:<br>a. Occurrence of severe linezolid-related toxicity<br>b. Time to first severe linezolid-related toxicity. |

### 9. DEFINITIONS

#### 9.1 Scheduled visits, visit window definition, and unscheduled visits

Scheduled study visits occur at screening, baseline (Week 0), at weekly intervals until Week 12, then at 4-weekly intervals until Week 36 and then at Week 39, 43, 47, 53, 59, 65, 73, 81, 89, 97 and 104, calculated from date of randomization. Each scheduled visit had a predefined allowable window of days before and after the target scheduled date during which a protocol-conforming study visit could occur.

For the purpose of analysis, the scheduled visit results will be used preferentially. If the scheduled visit did not occur, or if the scheduled visit result is missing, the value measured at the date closest to the target scheduled visit date and within the window period (as defined in Table 2) will be used for analyses.

If the scheduled visit result is missing and there is no result from an unscheduled visit in the window period, the result for that visit will be classified as missing. Exceptions are described for outcome assignment at Weeks 73 and 104 in section 9.2.

The following results from evaluations at pre-screening visits are also allowable to characterize participants at randomization and establish eligibility for analysis populations:

- TSH performed less than 4 weeks prior to screening visit date,
- HbA1c performed less than 3 months prior to screening visit date,
- HBV, HCV and HIV serology performed less than one month before screening visit date,
- CD4 count performed less than six months prior to screening visit date,
- HIV viral load performed less than 4 weeks prior to screening visit date
- Molecular tests for rifampicin and fluoroquinolone resistance performed by a designated study lab on pre-screening samples collected no more than 3 weeks prior to screening visit,
- Chest X-ray of adequate quality performed less than 3 weeks prior to baseline visit date.

For longitudinal analyses (e.g., change in time to positivity), all available values (from scheduled and unscheduled visits) will be used.

#### 9.2 Data Handling Convention for Week 73 and 104 outcomes

For assignment of outcomes at Weeks 73 and 104, data collected up to 30 days after the close of the window period around the study visit (i.e., up to 60 days after the target visit date) are included among those used to inform outcome classification. This applies to data on clinical, radiological, and bacteriological evolution, culture results, and death.

For the Week 73 endpoint, data outside the window period are only used if relevant results were not available between 65 to 73 Weeks. For the 104 Week endpoint, these data are only used if relevant results were not available between 97 to 104 Weeks.

#### 9.3 Definition of loss to follow-up

Participants who a) did not complete their scheduled final study visit and b) for whom study staff have no information and whom they have been unable to contact, in-person or by phone for more than 14 weeks before the last study visit per study schedule will be considered lost to follow-up.
The "loss to follow-up" designation is made only after the scheduled final study visit.

#### 9.4 Definition of recurrence, relapse, and reinfection

Recurrence is a composite outcome of relapse and reinfection:
- Relapse is recurrence that occurs after the end of the assigned study treatment and in the absence of genotypic evidence of a new infection.
- Reinfection is recurrence that occurs during or after study treatment and in the presence of genotypic evidence of a new infection.

In primary analysis, recurrences (relapse and reinfection) will be considered unfavorable outcomes. Sensitivity analysis is discussed in section 0.

#### 9.5 Definition of a culture result

A culture result is called positive for TB if *Mycobacterium tuberculosis* (MTB) grows in MGIT or LJ medium. If more than one culture result is available from sputum collected at a single visit, results will be collapsed into a single (positive-dominant) result for the purposes of all analyses as follows:
1) **Positive**, if culture result from sample A and/or sample B is positive for MTB.
2) **Negative**, if no culture result is positive for MTB and sample A and/or sample B is negative.
3) **Contaminated,** if culture results from sample A and/or sample B are contaminated or AFB/Contaminated and no culture result is negative or positive for MTB.
4) **Missing**, if no sputum sample was collected or no result is available from all samples collected at a single visit.

Outcome assignment relied on available genotypic evidence and consulted the study reference laboratory to establish or rule out cross contamination.

#### 9.6 Definition of an AFB smear result

A smear result is called positive if it is graded as 'scanty' or 1+ or more.
If more than one smear result is available from sputum collected at the same visit, results will be collapsed into a single (positive-dominant) smear result for the purposes of all analyses, with the following overall result:
1) **Positive**, if at least one of the smear results is positive; positive smears will be further classified as scanty, 1+, 2+, 3+ in accordance with the highest reported value among smear results a given visit;
2) **Negative,** if at least one of the smear results is negative and none of the additional smear results are positive;
3) **Missing,** if no sputum sample was collected or no result is available from all samples collected at a single visit.

#### 9.7 Definition of time to positivity (TTP) in MGIT

Time to positivity (TTP) in MGIT is "time to detection" of *M. tuberculosis* in the BACTEC 960 system. It is reported in days and hours.
- When MTB is not detected within 41 days and 23 hours, the MGIT culture result is assumed to be negative and the resulting TTP set to the maximum of 42 days.
- When a MGIT culture result is positive with a TTP greater than 41 days and 23 hours, the MGIT culture is considered negative and TTP is set to 42 days.
- When a MGIT culture result is contaminated or missing, the value for TTP is set to missing.

If more than TTP result is available from sputum collected at the same visit, results will be collapsed into a single TTP result for the purposes of analyses, with the following overall result:

- If both TTP results are positive, the mean value of 2 TTP is used.
- If only one is positive, the TTP from the positive sample is used.
- If both are negative, TTP is set to 42.
- If one is negative and the other is neither negative nor positive, then TTP is set to 42.

#### 9.8 Definition of adherence

Treatment adherence is defined as the percentage of total doses taken compared to the expected number of doses to be taken, based on assigned regimen. It does not discriminate among reasons for missed intakes. For each drug expected number of doses is calculated using: expected duration, number of daily doses, and the number of times per week at treatment initiation.

The following formula will be applied for each participant:

$$\frac{\sum_{i=1}^{n}(Total\ doses\ taken\ drug\ i)}{\sum_{i=1}^{n}\left(Number\ of\ daily\ doses\ drug\ i * Number\ of\ days\ per\ week\ drug\ i * (Expected\ duration\ drug\ i/7)\right)} \times 100$$

where:

- n= number of drugs prescribed at treatment initiation
- Total doses taken drug *i* = number of times the drug *i* was taken under DOT + number of times drug *i* was self-administered
- Number of daily doses drug *i* = number of prescribed doses in assigned regimen per day
- Number of days per week drug *i* = number of prescribed days of administration per week
- Expected duration drug *i* = expected number of days that the participant will receive drug *i* based on study protocol for experimental arms and WHO recommendations for control arm (see below).

The expected duration for each drug will be:

- In experimental arms: 273 days (9 months) for all drugs;
- In conventional (longer) control arm: 547 days (18 months) for oral drugs; 156 days (6 months, 6 times per week) for injectable drugs;
- In control arm with shorter MDR-TB regimen: 182 days (6 months) for bedaquiline; 273 days (9 months) for levofloxacin or moxifloxacin, clofazimine, pyrazinamide, ethambutol; 121 days (4 months) for ethionamide, isoniazid.

According to the study protocol, participants may take as long as 47 weeks (329 days) to complete 39 weeks of treatment in the experimental arms: doses taken after 47 weeks (329 days) have elapsed since randomization will be excluded from the adherence calculation.

Participants will be considered adherent to the study treatment if adherence is at least 80%.

#### 9.9 Definition of an adverse event

AE is any untoward medical occurrence in a study participant after administration of an investigational drug. It does not necessarily have a causal relationship with the study treatment. A pre-existing condition that deteriorates at any time during the study (e.g., increase of severity) is an AE.

All events are graded according to v.5.0 of the MSF Severity Scale. Grade 3 events are generally those which result in marked limitation in activity. Some assistance and medical intervention/therapy are usually required, hospitalization is possible.

#### 9.10 Definition of a serious adverse event

An SAE is defined as an AE meeting at least one of the following conditions:

- An event leading to death.
- A life-threatening event (defined as a subject at immediate risk of death at the time of the event; it does not refer to an event which hypothetically might have caused death if it were more severe).
- An event requiring hospitalization or prolongation of existing hospitalization. The following will not meet the definition of SAE: hospitalization for elective surgery planned prior to subject enrollment; admission to remove barriers to care in the ambulatory environment; admission to perform ECG per protocol; hospitalization for infection control or smear positivity; or visit to the hospital (e.g. emergency room) that lasted less than 24 hours and did not result in admission.
- An event resulting in a persistent or significant incapacity or substantial disruption of the ability to conduct normal life functions.
- An event resulting in congenital anomaly or birth defect.
- Any other important medical event that may not result in death, be immediately life-threatening, or require hospitalization may be considered a serious adverse experience when, based upon appropriate medical judgment, the event may jeopardize the subject and may require medical or surgical intervention to prevent one of the outcomes listed above. Examples of such medical events include allergic bronchospasm requiring intensive treatment in an emergency room or at home, blood dyscrasias or convulsions that do not result in inpatient hospitalization, or the development of drug dependency or drug abuse.

#### 9.11 Definition of an adverse event of special interest

An AE of special interest (AESI) is a medical concern specific to the investigational medicinal product(s) for which close monitoring is required. In this study, the following AEs, regardless of their seriousness or causal relationship to treatment, are considered of interest:

- Grade 3 or above “electrocardiogram QT corrected interval prolonged”;
- Grade 3 or above leukopenia, anemia or thrombocytopenia;
- Grade 3 or above peripheral neuropathy;
- Grade 3 or above optic neuritis;
- Grade 3 or above increase in alanine aminotransferase (ALT) or aspartate aminotransferase (AST).

#### 9.12 Definition of non-TB microbiology laboratory variables

Laboratory variables routinely collected are: complete blood cell count (including WBC count, neutrophils, lymphocytes, eosinophils, hemoglobin, hematocrit, platelet count, RBC), biochemistry and electrolytes (including ALT/AST, total and direct bilirubin, albumin, creatinine, total calcium, potassium and magnesium).

#### 9.13 Definition of clinical signs, symptoms, evaluations

These include: weight/height, TB signs and symptoms, functional status (ECOG), mental health (anxiety by GAD-7, depression by PHQ-9), neurologic (subjective sensory neuropathy grade), ophthalmologic (visual acuity, color blindness), audiometry assessments, chest radiograph, and at least duplicate electrocardiogram (Qt interval, heart rate, respiratory rate; mean of QTcF is used for analysis).

Reporting includes presence/absence and grading according to the MSF Severity Scale.

##### 9.14 Definition of severe linezolid-related toxicity

Any of the following events established as linezolid related:
1) Grade 3 or higher leukopenia, anemia, thrombocytopenia, peripheral neuropathy, or optic neuropathy;
2) SAEs; or
3) AEs requiring linezolid discontinuation.

#### 10. STATISTICAL METHODS AND DATA ANALYSIS

##### 10.1 General rules

Epicentre will perform final statistical analysis after the database lock and the final data review.

Data will be presented in summary tables by treatment arm or as a total group and by visit if applicable. Continuous variables will be summarized for each treatment arm using the number of observations available (N), number of missing observations, mean, standard deviation (SD), minimum, maximum, median and 25th (Q1) and 75th (Q3) percentiles. Categorical data will be summarized for each regimen group using counts of non-missing and missing data and percentages.
All statistical tests will be two-sided unless otherwise specified.

Statistical analyses will be performed using STATA version 17, R version 4.2 or higher versions of either.

##### 10.2 Data summaries/description

###### *10.2.1 Patient/Participant disposition*

The total number of patients/participants for each of the following categories will be presented in a table and in a study flow diagram (See in appendix 12):
- Screened patients and reason for non-inclusion
- Randomized participants
- Safety population (exposed participants)
- All-culture mITT population
- All-DST population
- mITT population
- PP population
- Assessable population

Although exclusion can be attributed to multiple reasons, only the primary reason will be displayed, in the following order of priority:

1. FQ resistance/RIF susceptibility or no MTB
    a. MTB not detected in 1st-line molecular test
    b. RIF susceptible

   c. RIF indeterminate
   d. MTB not detected in $2^{nd}$-line molecular test
   e. FQ resistant
   f. FQ indeterminate
   g. $1^{st}$ or $2^{nd}$-line molecular tests results missing

2. Age <inclusion age approved in the country
3. Cardiac risk factor
4. Lab value outside acceptable range
5. Contraindicated drug
6. Prior exposure/resistance
7. Drug allergy
8. Pregnant or breastfeeding
9. Unwilling to use contraception
10. Did not consent to study participation or withdrew before inclusion
11. Investigator discretion
12. Participation in other trial
13. Screening not complete in 14 days

Frequency and reason for discontinuation of treatment and/or study will be presented in a table.

##### *10.2.2 Demographic and baseline characteristics*

Demographic characteristics:

- Sex
- Age (years) at baseline

Clinical laboratory baseline (or screening) characteristics:

- Complete blood cell count at screening
- ALT/AST, total and direct bilirubin, albumin, creatinine, electrolyte testing at screening
- TSH
- HbA1C or 2 fasting blood glucose tests if HbA1C not done
- CD4 cell count and Viral load for HIV+ participants
- Hepatitis serology
- Pregnancy test

Microbiological laboratory baseline (or screening) characteristics:

- Sputum smear microscopy (Negative, Scanty, 1+, 2+, 3+, missing) at screening-collapsed
- Genotypic and phenotypic drug susceptibility testing results
- Sputum culture (Positive for MTB, Negative, Contaminated (contaminated + AFB/contaminated + positive for NTM)) at screening-collapsed across MGIT & LJ
- TB culture on MGIT and LJ separately in sites where both were used at screening
- Time to positivity (TTP) in MGIT at screening

Clinical and other baseline (or screening) characteristics:

- Medical history at screening
- TB history at screening
- TB symptom assessment
- Vital signs (body weight, height, BMI, oxygen saturation, temperature, pulse, respiratory rates and blood pressure)
- Physical exam (Symptoms/signs) (Grade 0-4)
- ECOG assessment

- Neurologic exam results (Grade 0-3)
- Ophthalmologic exam results (Grade 0-4)
- Mental health assessment results (Anxiety grade 0-4, Depression grade 0-4)
- Audiometry results (Grade 0-4)
- Concomitant medication (None, any; anti-retroviral therapy among HIV co-infected; other major immunomodulators [e.g. corticosteroids])
- Chest X ray (normal in both lungs/abnormal, presence of cavity and other lung lesions, extent of disease limited/moderate/extensive)
- ECG (Mean of the 2 highest QTcF intervals, Normal/Abnormal, Grade)
- Risk factors (history of homeless, unemployment, drugs use, cigarette smoking and alcohol use)

Descriptive statistics will be displayed to summarize the demographic and baseline characteristics data on the safety population. A priori no statistical test will be performed at baseline between treatment arms.

###### *10.2.3 Regimen characteristics (control arm)*

Composition and duration of control arm regimens will be described. The proportion of control arm participants who received a shortened (6-12 month) regimen will be reported.

###### *10.2.4 Adherence*

Percentage of adherence will be summarized descriptively by treatment arm overall for the mITT and safety populations.

#### 10.3 Efficacy analyses

Analyses for the efficacy endpoints will be performed in the mITT and PP populations.

##### *10.3.1 Analysis of the primary efficacy endpoint (PO)*

Analysis of the primary endpoint will separately compare the proportions of participants with a favorable outcome at Week 73 between each experimental arm and the control.

For all pairwise comparisons, the absolute, unadjusted difference in proportions (risk difference) will be estimated with corresponding 2-sided 95% confidence interval (CI) using a binomial regression model (generalized linear model for a binomial outcome with an identity link function). In case of convergence failure, risk difference will be estimated using a modified Poisson regression model with robust standard errors. A 2-sided 95% CI of the proportion of participants with a favorable outcome will also be estimated for all regimens.

The non-inferiority of an experimental arm compared to the control will be established if the difference in proportion with favorable outcome (proportion of participants with favorable outcome in the experimental arm minus proportion of participants with favorable outcome in the control arm) at Week 73 is greater than the lower non-inferiority margin, i.e. if the lower bound of the one-sided 97.5% CI (which correspond to the lower bound of the 2-sided 95% CI) is greater than or equal to -12% in both mITT and PP populations.

Multiple experimental regimens will be compared to one active control. The type I error will be controlled by ordering the non-inferiority comparisons, according to a fixed sequence approach: the regimen with the highest proportion of favorable outcomes will be compared to the control first. If non-inferiority is established, then a comparison between the control and the experimental regimen having the second highest proportion of favorable outcomes will be performed, and so on as long as non-inferiority is concluded. Once the non-inferiority is not demonstrated, the non-inferiority comparisons of all remaining experimental regimens will stop. All completed comparisons will be done at the full, one-sided alpha level of 2.5%.

For each experimental regimen for which non-inferiority is demonstrated, superiority compared to the control will then be tested at the 5% level of significance.

No adjustments will be made to control the type I error.

Adjusted estimate of risk difference with corresponding 95% confidence interval will be calculated using a binomial regression model. Country, BMI, pyrazinamide resistance, injectable resistance, age, sex, presence of each comorbidity at baseline or during study participation (including HIV, Hepatitis B and C status, diabetes, SARS-Cov-2 infection and COVID-19), smear result (negative/scanty, 1-2+, 3+, at screening), cavitation (presence or absence, at baseline), prior exposure to TB treatment, extent of disease (limited/moderate/extensive, based on chest X-ray at baseline) will be considered as covariates. Any covariate that is significant ($p<0.10$) in the univariate analysis will be introduced into a multivariable model and through a process of backward elimination only significant variables with $p<0.05$ will be retain in the final model.

*10.3.1.1 Subgroup analyses of the primary efficacy endpoint (PO)*

Sub-group analyses of the primary efficacy endpoint will be performed (in both mITT and PP populations) for regimens that are shown to be non-inferior. Subgroups will be established on the following baseline characteristics:

- Country (Georgia, India, Kazakhstan, Lesotho, Pakistan, Peru, South Africa)
- Presence of comorbidities (presence/absence of: HIV, hepatitis C, diabetes, and SARS-CoV-2 infection/COVID-19)
- BMI (<18.5 kg/m2, 18.5-24.9 kg/m2, ≥25 kg/m2)
- Pyrazinamide resistance (sensitive/resistant)
- Age (<18, 18-45, ≥45)
- Sex (male/female)
- Smear result (at screening) (positive/negative)
- Cavitation (presence/absence)
- Prior exposure to TB treatment (none/first line only/other)
- WHO recommendations implemented (by implementation date).

*10.3.1.2 Sensitivity analyses of the primary efficacy endpoint (PO)*

The primary unadjusted and adjusted efficacy analyses will be repeated:

- in the following populations (described in section 7.1.3): assessable, all-culture mITT and all-DST mITT populations.
- in the assessable population reclassifying outcomes of “LTFU” or “Unfavorable because unassessable” (as defined in section 8.1) as the outcome that had been assigned at Week 39.

- excluding participants from the control arm who received a shortened regimen, if the proportion exceeds 10%.
- using multiple imputation to impute missing data after examination of the missing data pattern, if missing values in the final multivariable model reduce the relevant analysis population by more than 20%.

##### *10.3.2 Analysis of the secondary efficacy endpoints*

For the analysis of the secondary efficacy endpoints, proportions will be calculated by treatment group and corresponding 95% confidence intervals will be presented.

For all pairwise comparison of proportions (between experimental groups and the control group), the risk difference will be estimated with corresponding 95% confidence interval using a binomial regression model.

In all adjusted models, country, BMI, pyrazinamide resistance, injectable resistance, age, sex, presence of each comorbidity at baseline or during study participation (including HIV, Hepatitis B and C status, diabetes, SARS-Cov-2 infection and COVID-19), smear result (negative/scanty, 1-2+, 3+, at screening), cavitation (presence or absence, at baseline), prior exposure to TB treatment, extent of disease (limited/moderate/extensive, based on chest X-ray at baseline) will be considered as covariates. Those significant at $p<0.10$ in the univariate analysis will be introduced into a multivariable model and through a process of backward elimination only significant variables with $p<0.05$ will be retain in the final model.

No adjustments in significance levels will be made for multiple comparisons.

Analyses will be performed for:

1) SEO1: Proportion of participants with a favorable outcome at Week 104.

2) SEO2: Sputum culture conversion (as defined in Section 8.1.2):
   a. Proportion of participants with initial sputum culture conversion by Week 8,
   b. Time to initial sputum culture conversion assessed in MGIT system will be separately compared between each experimental arm and the control using Kaplan-Meier analysis. Data for participants who discontinued treatment will be censored at the last assessment. The logrank test will be used for the comparison of the median time to sputum culture conversion. Kaplan-Meier curves will be presented. A cox proportional hazards model will be used to estimate hazard ratios with corresponding two-sided 95% confidence intervals and p-value. Adjusted hazard ratios with corresponding 95% confidence interval will be calculated using a Cox proportional hazards model, with adjustment performed as described above.
   c. Mean change from baseline in time to positivity (TTP) in MGIT over 8 weeks will be compared between each experimental arm and the control using a linear mixed-effects regression model, with change from baseline as the outcome. Visit, treatment group, interaction visit*treatment group and baseline TTP value will be fixed effects. Study participant will be treated as a random effect to account for both heterogeneity among participants and correlation among measurements taken on the same participant over time. To evaluate the approach of managing negative culture, this will also be analyzed in a mixed-effects tobit regression model where negative culture will

be right-censored.

3) SEO3: Proportion of participants with a favorable outcome at Week 39.

4) SEO4: Proportion of participants with a treatment failure or recurrence at Week 73 and at Week 104.

*Sensitivity analyses of the secondary efficacy endpoints*

Analysis of the risk differences in proportions of participants with a favorable outcome at Week 104 (SEO1) and participants with treatment failure or recurrence at Week 104 (SEO4) will be repeated:

- in the following populations (described in section 7.1.3): assessable, all-culture mITT and all-DST mITT populations.
- in the assessable population reclassifying outcomes of "LTFU" or "Unfavorable because unassessable" (as defined in section 8.1) as the outcome that had been assigned at Week 73.
- excluding participants from the control arm who received a shortened regimen, if the proportion exceeds 10%.

Adjusted analyses will repeated using mutiple imputation to impute missing data after examination of the missing data pattern, if missing values reduce the relevant analysis population by more than 20%.

##### *10.3.3 Analysis of the exploratory efficacy endpoints*

Analyses of exploratory endpoints will be performed in the mITT population. No adjustments in significance levels will be made for multiple comparisons.

1) EEO1: Comparison between experimental arms: For all pairwise comparisons across experimental arms (regimen 1 vs. 2, 1 vs. 3, 2 vs. 3, 2 vs. 4, 3 vs. 4, 4 vs. 5), the risk difference in proportion of participants with a favorable outcome at Week 73 will be estimated with corresponding 95% confidence interval using a binomial regression model.
2) EEO3: Sputum culture conversion as a marker of early treatment response: Three logistic regression analyses will be performed with unfavorable outcome at Week 73 as the dependent variable and each MGIT-based early treatment response outcome (1. Proportion of participants with culture conversion achieved within 8 weeks; 2. Time to culture conversion; 3. Change in time to positivity) and potential covariates as independent variables. Odds ratios and corresponding 95% confidence intervals will be estimated. Performance characteristics (sensitivity, specificity, negative and positive predictive value, area under the curve) of culture conversion by 8 weeks will also be calculated.
3) EEO4: Proportion of participants with a favorable outcome at Week 73: Pairwise comparisons will be performed between participants with strains that are resistant to pyrazinamide and those with strains that are susceptible to pyrazinamide.. The risk difference will be estimated with corresponding 95% confidence interval using a binomial regression model. An interaction term between treatment arm and strain type (resistant/susceptible) will be added to the model to test if a differential effect of treatment on the proportion of participants with a

favorable outcome at Week 73 was present for strains that are resistant versus strains that are susceptible.

4) EEO5: Proportion of participants with drug resistance amplification among participants with failure or relapse outcome at Week 104: Proportion of participants with drug resistance amplification among participants with failure or relapse outcome at Week 104 will be calculated by treatment arm as number of participants with failure or relapse at Week 104 (denominator) and number with failure or relapse at Week 104 with drug resistance amplification (numerator). The risk difference between each experimental arm and the control (and between experimental arms) will be estimated with corresponding 95% confidence interval using a binomial regression model.
5) EEO6: Proportion of adherent participants: For all pairwise comparisons between experimental arms and the control and across experimental arms, the risk difference will be estimated with corresponding 95% confidence interval using a binomial regression model.
6) EEO8: Proportion of participants with a favorable outcome at Week 73 and 104 compared between participants in experimental arms randomized to linezolid dose-reduction strategies: the risk difference will be estimated with corresponding 95% confidence interval using a binomial regression model.

*Sensitivity analyses of the exploratory efficacy endpoints*

Analysis for EEO8 will be repeated in participants in experimental arms who received linezolid dose-reduction strategies, irrespective of randomization[4].

#### 10.4 Safety analyses

The safety analysis will be based on the reported adverse events (AEs) in the case report forms, and other safety information, such as laboratory evaluations, clinical evaluations including neurological assessments, physical exams, mental health, X-ray, and ECG performed during the follow-up of the study.

The analyses of safety data will be performed on the safety population during the intervals of 73 weeks and 104 weeks post-randomization.

The Pharmacovigilance Database (PVDB) will be consulted as necessary to supplement safety data captured in the Clinical Database, OpenClinica (OC).

Missing data will not be imputed for safety analyses.

[4] Prior to protocol v. 3.0, dose-reduction assignment was selected by investigator. This sensitivity analysis will include all participants who experienced dose reduction, rather than only those in whom it was assigned through randomization.

##### 10.4.1 Analysis of adverse events

Each AE is coded to a "Preferred Term" (PT) and primary associated "System-Organ Class" (SOC) according to the MedDRA dictionary (Medical Dictionary for Regulatory Activities, version 19.1). AE grade 3 or greater, SAEs and AESIs are also coded to a category, as described in appendix 12.2.

The summaries of AE by treatment group and grade will include:

- The number and percentage of participants with at least one AE
- The number and percentage of participants with at least one AE in a specific SOC/PT.

Number and percentage of participants with at least one of the following events will also be provided, by SOC, PT and category:

- AEs of Grade 3 or higher
- Serious AEs regardless of grade
- AEs leading to death of any cause (overall) regardless of grade
- AE of special interest.

Counts will be provided by regimen for each PT within each SOC, and by category, and regardless of relationship to the study drug.

For all pairwise comparisons between experimental arms and the control group, the risk difference will be estimated with corresponding 95% confidence interval using a binomial regression model for:

- SSO5: the proportion of participants who died of any cause up to Week 73 / Week 104
- SSO6: a) the proportion of participants with AEs of Grade 3 or higher by Week 73 / Week 104;
  b) the proportion of participants with SAE up to Week 73 / Week 104; and
  c) the proportion of participants with AEs grade 3 or higher or SAEs of any grade up to Week 73 / Week 104
- SSO7: the proportion of participants with AESI up to Week 73 / Week 104.

##### 10.4.2 Analysis of laboratory variables

Descriptive statistics of laboratory parameters (including white blood cells, neutrophils, hemoglobin, platelet count, ALT, AST, total and direct bilirubin, creatinine, potassium, TSH, HbA1C, CD4 count, HIV1HIV-1 RNA) grades and changes in grades from baseline will be presented by regimen at each time point they were measured, following the schedule of events. Results from unscheduled visits will not be reported in the descriptive summaries or in tables of changes from baseline. Results from unscheduled visits will be included in the reporting frequency, time, and grade of AEs, SAEs, and AESIs and in listings of abnormal values.

Figures will present the mean change of grade over time by treatment arm.

##### 10.4.3 Analyses of clinical signs and assessments

Descriptive statistics of parameters and changes from baseline will be presented by regimen at each time point they were measured, following the schedule of events. When available, grades will be used rather than parameter value (physical exam: cough, hemoptysis, chest pain, fever (from severity scale), dyspnea, ECOG, mental health assessment (anxiety and depression grades), neurological, ophthalmological, neurological, ophthalmological and audiometry assessments, BMI, chest X-ray (including normal/abnormal, cavitation, extent of disease). For ECG, mean of the two highest QTcF intervals will be presented as well as grade. For other clinical signs, we do not plan to report descriptively.

Results from unscheduled visits will not be reported in the descriptive summaries or in tables of changes from baseline. Results from unscheduled visits will be included in the reporting frequency, time, and grade of AEs, SAEs, and AESIs and in listings of abnormal values.

Figures will present the mean change overtime by treatment arm.

###### *10.4.4 Analysis of the exploratory safety endpoints*

1) ESO2: Comparison between experimental arms: For all pairwise comparisons across experimental arms (regimen 1 vs. 2, 1 vs. 3, 2 vs. 3, 2 vs. 4, 3 vs. 4, 4 vs. 5), the risk difference in proportion of participants with grade 3 or higher AEs or SAEs of any grade by Week 73 will be estimated with corresponding 95% confidence interval using a binomial regression model.
2) ESO7: The risk difference in proportions of participants with severe linezolid-related toxicity occurring after linezolid dose-reduction between participants randomized to each linezolid dose-reduction strategy (300 mg daily or 600 mg thrice weekly) will be estimated with corresponding 95% confidence interval using a binomial regression model.

   Median time from linezolid dose reduction randomization to first severe linezolid-related toxicity occurring after linezolid dose-reduction will be compared between participants randomized to linezolid dose-reduction strategies using a two-sided log-rank test at the 5% significance level. Kaplan-Meier curves will be presented. A Cox proportional hazards model will be used to estimate hazard ratios with corresponding two-sided 95% confidence intervals and p-value. Adjusted hazard ratios with corresponding 95% confidence interval will be calculated using a cox proportional hazards model. Country, BMI, age, sex, presence of comorbidities (including HIV and receipt of ART, Hepatitis B and C status, diabetes, SARS-Cov-2 infection and COVID-19), substance use, companion drugs, and concomitant medications will be considered as covariates. Estimates and 95% confidence intervals will be obtained and presented. Those significant at $p<0.10$ in the univariate analysis will be introduced into a multivariable model and through a process of backward elimination only significant variables with $p<0.05$ will be retained in the final model. In adjusted analysis, if missing values reduce the relevant analysis population by more than 20%, we will consider imputing the missing data, using multiple imputation after examination of the missing data pattern. No adjustments in significance levels will be made for multiple comparisons.

##### 10.5 Major protocol deviations

Major protocol deviations (as defined in SOP SM-005-CT) will be listed by site with information such as type of deviation, date of occurrence/detection and reason. The percent of subjects with major deviations by type will also be summarized by country and study arms.

#### 11. INTERIM ANALYSIS

Bayesian interim analyses for randomization adaptation were performed every four weeks by an external and independent statistician.

A data safety and monitoring board (DSMB) independently assessed the safety and risk/benefit semi-annually during recruitment and delivery of experimental treatment. Analyses were performed by an external statistician.

#### 12. APPENDIX

##### 12.1 Study flow diagram

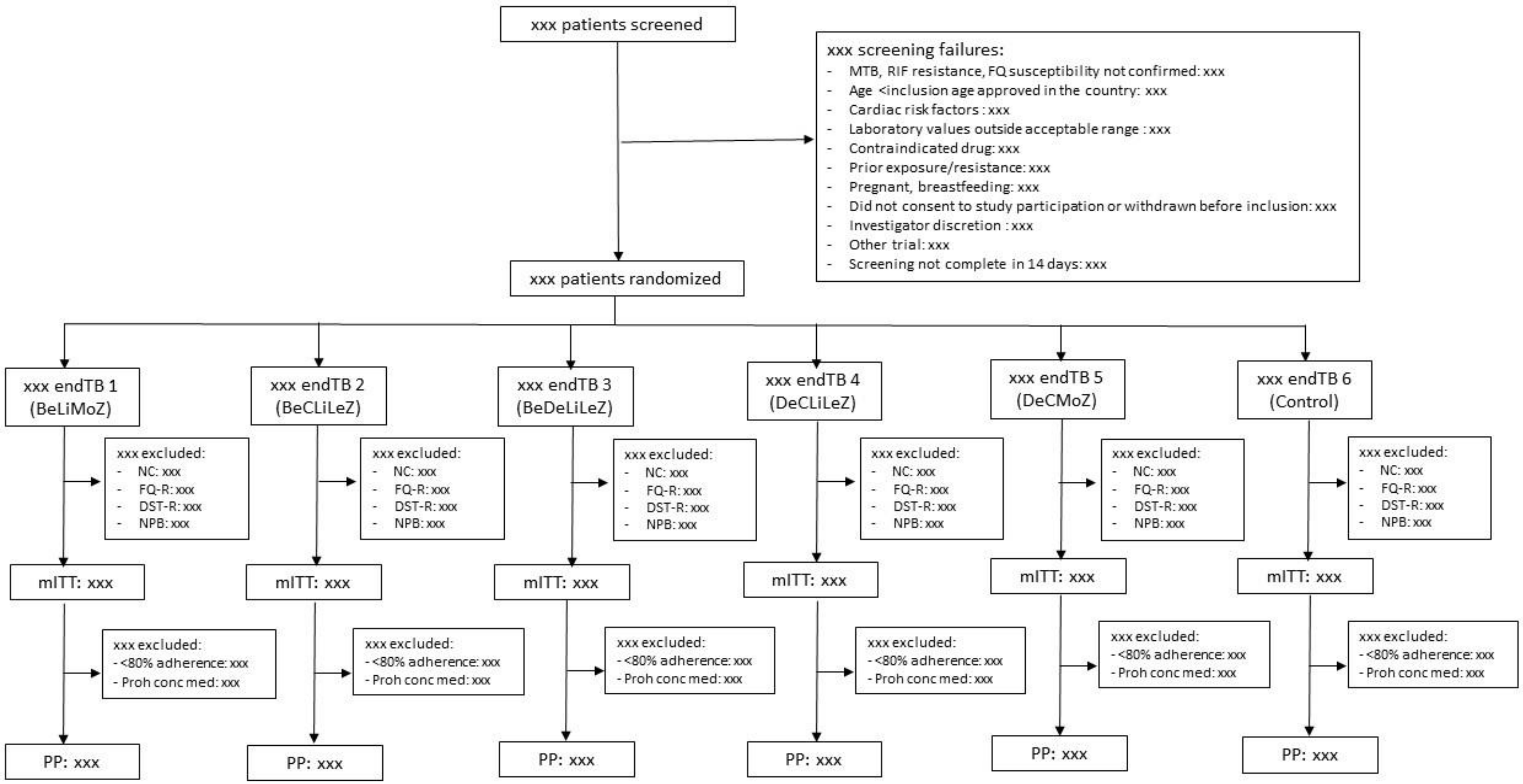


NC = Negative culture; FQ-R = FQ resistance; DST-R = Screening/Baseline DST resistant result to any drug from regimen; NPB = No post-baseline data (erroneous inclusion in trial, …);
<80% adherence = <80% adherence other than death and LTFU; Proh conc med = >7 days of prohibited concomitant medication other than death and LTFU

#### 12.2 AE categories

In addition to “Preferred Term” (PT) and primary associated “System-Organ Class” (SOC) according to the MedDRA dictionary (Medical Dictionary for Regulatory Activities, version 19.1), AE grade 3 or higher, SAE and AESI will be coded to one of the following categories, partially based on the Standardized MedDRA Queries:

- Blood glucose abnormalities
- Breast conditions
- Cardiac abnormalities
- Death of unknown cause
- Diabetes-related events
- Electrolytes abnormalities
- Eye/vision abnormalities
- Female reproductive system abnormalities
- Fractures
- Gastrointestinal abnormalities
- General signs and symptoms
- General system disorders
- Hearing abnormalities
- Hematologic abnormalities
- Immune reactions
- Injuries
- Joint disorders
- Liver-related abnormalities
- Male reproductive conditions
- Musculoskeletal symptoms
- Neoplasms
- Optic nerve abnormalities
- Other ear signs and symptoms
- Other infections
- Other investigations
- Other lab abnormalities
- Other metabolic abnormalities
- Other nervous system abnormalities
- Pancreatic abnormalities
- Peripheral neuropathy and related symptoms
- Pneumonias
- Pregnancy and Pregnancy-related events
- Psychiatric events
- Renal abnormalities
- Reproductive system abnormalities
- Respiratory events
- Seizure disorders
- Skin conditions
- Substance abuses
- TB/TB progression
- Tendon and ligaments abnormalities
- Thyroid conditions
- Urinary conditions

- Vascular events

---

[1] Cellamare M, Ventz S, Baudin E, Mitnick CD, Trippa L. A Bayesian response-adaptive trial in tuberculosis: The endTB trial. Clin Trials. 2017 Feb;14(1):17-28. doi: 10.1177/1740774516665090.

[2] Lee, M. et al. Linezolid for Treatment of Chronic Extensively Drug-Resistant Tuberculosis. N. Engl. J. Med. 367, 1508–1518 (2012).

[3] Cox, H. & Ford, N. Linezolid for the treatment of complicated drug-resistant tuberculosis: a systematic review and meta-analysis. Int. J. Tuberc. Lung Dis. 16, 447–454 (2012).
