## Supplemental Appendix for "Nine-month, all-oral regimens for rifampin-resistant tuberculosis"

#### 1 List of endTB Collaborators/Contributors (and study sites)

***Aundh Chest Hospital, India (study site):*** Gurpreet Arora, Valay Ashara, Sonali Bhattacharjee, Rakshanda Borkar, Namita Chincholikar, Nikita Choudhary, Praveen Davuluri, Neha Desai, Dipika, Sunil Dhongade, Drishti Dulani, Sheetal Ghule, Nilma Hirani, Prakash Hurali, Reeana Irani, Kiran Jadhav, Pritesh Jadhav, Radhika Jadhav, Nidhin Joseph, Akshay Joshi, Pranav Joshi, Akhilraj K, Sagar K, Madhavi Kamble, Rashmi Kate, Tanvi Keer, Madhuri S Kenche, Harsha Khandare, Pranesha Koli, Apurva Kulkarni, Sagar Kumar Patra, Abhishek Kuwar, Prakash Lande, Nana Mane, Priya Masalge, Priya Mishra, Rishabh Mitkari, Ajit More, Pracheta Munot, Ramya Murgesh, Sagar Muthe, Nupur Nakashe, Dwaragesh Natarajan, Sharad Paril, Supriya Patil, Vaishali Patil, Vaijayanti Patwardhan, Vivek Posture, Anil Pradeshi, Vanya Prasad, Bhavya Pundir, Yogesh Randhit, Michael Reckart, Pradnya Samant, Hanif Shaikh, Ashutosh Sharma, Neela Sharma, Sharda Sharma, Nilesh Shinde, Suvarna Shinde, Gauri Shingare, Abhay Shulka, Aniket Shriwas, Lokesh Sonawane, Urmila Sudhindra, Dipali Suryavanshi, Akshay Thakare, Danashree Udare, Gayatri Vaidya, Rani Wagh

***City Centre of Phthisiopulmonology, Kazakhstan (study site):*** Lyazzat Isenova, Gulbakhar Konigratbayeva, Gulnar Nurtayeva, Nurlan Zekeshov

***Epicentre, France*** **:** Romain Chenu, Souna Garba, Daiane Tozzi, Francis-Jaudel Yuya-Septoh

***Harvard Medical School, USA****:* Eva Chang, Julia Coit, Jacquelyn-My Do, Allison LaHood, Kelsey O'Brien, Enoma Okunbor, Elna Osso

***Hospital Nacional Dos de Mayo, Peru (study site):*** Victor Chávez, Piero Larco, Daniel Mendoza, Christian Robles, Angelo Rojas, Janet Salazar, Gabriela Santos, Juan Sosa, Cesar Ticona

***Hospital Nacional Hipólito Unanue, Peru (study site):*** Shirley Carranza, Silvana de la Gala, Diego Delgado, Cemir Galarza, Gladys Murga, Elizabeth Ramos

***Hospital Nacional Sergio Bernales, Peru (study site):*** Kelly Cantaro, Andrely Huerta, Jonathan Maldonado, Roberto Nuñez, Katherine Panduro, Ruth Rafael

***The Indus Hospital & Health Network, Pakistan (study site):*** Amaan Abdul, Sara Abedin, Eraj Afreen, Sohail Ahmed, Ubaid Ahmed, Jinsar Ali Shah, Azka Ashraf, Faisal Aslam, Zahida Bashir, Saba Fayyaz, Kiren Hafeez, Meherunissa Hamid, Basmah Hassan, Muzamil Hussain, Muhammad Irfan, Maria Omar, Shafiqua Parveen, Sharoon Sunail, Muhammad Farooq, Batool Mazhar, Sana Munir, Neelofer Sonia, Muhammad Yaseen, Muhammad Zubair, Fatima Zehra, Iqra Zulfiqar

***Institute of Chest Diseases, Pakistan (study site):*** Tazeen Abbasi, Ghulam Akbar Abro, Mushtaque Ahmed, Nazir Ahmed, Muhammad Hammad Ali, Nazakat Ali, Rao Danish, Ayesha Farhat, Sunaina Gill, Muhammad Hafeez, Ghulam Hussain, Habib Inayat, Sylvia Johnson, Farees Kamal, Adil Kamran, Sanjeet Kumar, Mahveer Maheshwari, Shahid Mamsa, Sumera Massey, Asif Mehmood, Aaisha Memon, Ayaz Ali Mirani, Shafaque Naz, Tanzeel Rafique Qureshi, Yash Roop Moazzam Sheikh, Muhammad Rafi Siddiqui, Bhom Singh, Mustajab Soomro

***Institute of Tropical Medicine, Belgium:*** Elisa Ardizzoni, Leen Rigouts, Praha Rupasinghe

***Interactive Development and Research, Pakistan:*** Annum Aftab, Saman Ahmed, Muhammad Adnan Alamgir, Ayaz Ali, Azhar Ali, Shahzaib Ahmed Ali, Sakhawat Ali, Sanam Altaf, Samreen Amjad, Mahnoor Sahiba Arshad, Sadia Ausim, Abdul Basit, Haroon Ezik, Hira Hussain, Sibtain Hyder, Shahzad Inayat, Aleeza Janmohammed, Komal Jaseem, Aqsa Jawed, Lamis Maniar, Pardeep Kumar, Susheel Kumar, Reema Latif, Sara Liaqat, Muhammad Ammar Nasser, Muhammad Shakeel, Shahzaib Shaikh, Anique Siddiqui, Jetmal Singh, Koushalia Sivan, Momal Taimoor, Nabeel Zia

***National Center for Tuberculosis and Lung Diseases, Georgia (study site):*** Tinatin Amirejibi, Rusudan Aspindzelashvili, Zaza Avaliani, Nino Bablishvili, Mariam Bichiashvili, Teona Bichashvili, Lasha Darchia, Marika Eristavi, Marine

Gachechiladze, Irine Keinashvili, Nino Kiria, Nani Kristesiashvili, Valerie Marechal, Darejan Meskhishvili, Lali Mikiashvili, Lia Trapaidze, Dali Zhizhilashvili, Maia Zhorzholiani

***National Center for Tuberculosis Problems, Kazakhstan (study site):*** Inessa Li, Merei Otepbergenova

***National Scientific Center of Phthisiopulmonology, Kazakhstan (study site):*** Gaukhar Abildayeva, Amanzhan Abubakirov, Sagit Bektasov, Orazbayeva Maryash, Elena Telegina, Lyailya Tursynbekova

***Partners In Health Kazakhstan, Kazakhstan:*** Zhanelya Amanzholova, Perizat Bektassova, Anel Belgozhanova, Nazerke Birimkulova, Nurgul Dyusebayeva, Nilay Erkut, Arnur Gusmanov, Banu Kassenova, Aigerim Kassym, Kanat Khazhidinov, Tatyana Lee, Azhar Magzumova, Moldir Mussabayeva, Gulnar Omarova, Nadezhda Ryapolova, Lyazzat Sagyndykova, Asel Serekbay, Assel Stambekova, Gulmira Tanatarova, Zinatdin Uaisov, Zhanel Zhantuarova, Gulnara Zhumakairova

***Partners In Health, USA:*** Nataliya Arlyapova, Joshua Bogus, Merida Carmona, Clare Fanagan, Sarah McAnaw, Alyssa Scharff, Sonya Soni, Megan Striplin

***Partners In Health Lesotho, Botsabelo MDR-TB Hospital, Lesotho (study site):*** Seyfu Abebe, Johnson Alakaye, Atang Bulane, Michael Custodio, Precious Hajison, Malefetsane Hetsa, David Holtzman, Matlotliso Khesa, Makatleho Lethola, Talime Lebitsa, Mpho Mahamo, Joalane Makaka, Mokenyakenya Matoko, Stephane Mpinda, Polo Mohoang, Daniel Monyaesa, Patrick Nkundayirazo, Mabohlokoa Ntsane (Sesomo Mohale), Seabata Ntsibane, Sebakeng Phate, Mathemba Radebe (Retsepile Tlali), Moliehi Rakhetsi, Tello Ranoosi, Meseret Tamirat, Mahooe Thokoana

***Partners In Health Peru (Socios En Salud), Peru:*** Ericka Alegre, Christian Aguilar, Lourdes Armuto, Nadia Barreda, Maria Barreto, Stephany Cabrera, Roger Calderon, Milagros Castro, Jelina Chavez, Laura Chavez, Haydee Cheje, Lesli Cori, Daniel Dávalos, Karen Delgadillo, Hannely Diaz, Ximena Flores, Jimmy Galarza, Dalicxa García, Fanny Garcia, Miriam Gaspar, Roselena Godos, Pamela Gomez, Luz Gonzales, Lizsery Guerrero, Sadith Inga, Judith Jasaycucho, Ana Pro Martinez, Raquel Molina, Raquel Mugruza, Merilyn Nuahan, Kevin Ore, Liliana Panduro, Olga Peña, Sara Perea, Cynthia Pinedo, Yhomay Ponce, Maritza Quiñones, Alicia Ramos, Elmer Ramirez, Rosina Reinoso, Cynthia Reyes, Jessica Reyes, Vanessa Riccio, Bryan Robles, Karen Rojas, Oswaldo Sanabria, Maria Saravia, Susana Saravia, Liz Senador, Jose Soberon, Edith Soncco, Maribel Soto, Catherine Suarez, Milagros Suarez, Veronica Suarez, Erika Torpoco, Oscar Torres, Claudia Torrez, Gabriela Tunque, Aida Ugarte, Gissela Vaderrama, William Valdivia, Isabel Valverde, Cynthia Vargas, Janet Vargas, Elvia Vasquez, Juan Veliz, Dioselinda Villa, Sheyla Villafuerte, Geraldine Villar, Stephanie Villegas, Milagros Wong, Rosa Yataco

***Médecins Sans Frontières Khayelitsha, South Africa (study site):*** Neide Cossa, Stobdan Kalon, Christine Knel, Nopinky Matinse, Vuyokazi Mbanjwa, Axole Ndayi, Tasanya Nomaliso, Nelisiwe Ntali, Tabassum Rashid, Allan Taylor, Hamza van der Ross, Brenda Wright

***Médecins Sans Frontières OCG, Switzerland*** **:** Sylvine Coutisson

***Médecins Sans Frontières OCP, France :*** Melchior Atger, Sandra Nadia Baya, Marwa Bekhiet, Veronique Boissière, Roberta Caboclo, Marhaba Chaudhry, Sandrine Cloez, Sandra Collin, Céline Delifer, Shana Desmaisons, Vanessa Ducher, Cathy Hewison, Mohamed Ibrahim, Kristen Lebeau, Merry Mazmanian, Rada Mirzayeva, Mathilde Moreau, Monica Moschioni, Audrey Pâquet, Christophe Perrin, Laura Pichon, Jeanne Roussel, Martina Scotton

### 2 Methods

#### 2.1 Objectives

##### 2.1.1 Primary Objective

The primary objective is to assess whether the efficacy of experimental regimens at Week 73 is non-inferior to that of the control.

##### 2.1.2 Secondary Objectives

1. To compare the efficacy of experimental regimens at Week 104 to that of the control
2. To compare the frequency of and time to early treatment response (culture conversion) in experimental regimens to that of the control
3. To compare the efficacy of experimental regimens at Week 39 to that of the control
4. To compare, at Week 73 and Week 104, the proportion of patients who experienced failure or relapse in the experimental arms to that in the control arm
5. To compare, at Week 73 and Week 104, the proportion of patients who died of any cause in the experimental arms to that in the control arm
6. To compare, at Week 73 and Week 104, the proportion of patients who experience grade 3 or higher AEs or SAEs of any grade in the experimental arms to that in the control arm
7. To compare, at Week 73 and Week 104, the proportion of patients who experience AESIs in experimental regimens to that in the control arm

##### 2.1.3 Exploratory Objectives

1. To compare safety and efficacy endpoints across experimental arms, e.g., arms containing bedaquiline vs. delamanid (regimen 2 vs. 4); arms containing clofazimine & levofloxacin vs. arms containing moxifloxacin (regimen 2 vs. 1); arm containing bedaquiline and delamanid vs. arms containing one of the drugs and clofazimine (regimen 3 vs. 4 and regimen 3 vs. 2)
2. To evaluate conversion endpoints as potential surrogate markers for unfavorable outcome
3. To compare the activity of regimens between patients with strains that are resistant to important drugs in those regimens (e.g., pyrazinamide) and patients with strains that are sensitive
4. To estimate the frequency of drug resistance amplification among treatment failures and relapses occurring by Week 104
5. To compare treatment adherence in the experimental arms to that in the control and across regimens
6. To compare the frequency of and time to severe linezolid-related toxicity between linezolid dose-reduction strategies.
7. To compare, at Week 73 and Week 104, efficacy endpoints across linezolid dose-reduction strategies: 300 mg daily or 600 mg thrice weekly.

#### 2.2 Design and oversight

The endTB protocol and statistical analysis plan are available at NEJM.org.

**Other Committees**

*endTB Laboratory Advisory Group (LAG)*

The endTB LAG provided scientific oversight of issues that involved research on specimens and/or isolates collected from participants. They advised on the relevance and scientific validity of laboratory procedures and potential ancillary laboratory studies.

LAG members: Martina Casenghi (Elizabeth Glaser Pediatric AIDS Foundation), Daniella Cirillo (San Raffaele Scientific Institute), Ted Cohen (Yale School of Public Health), Claudia Denkinger (University Hospital Heidelberg), Kathleen Eisenach (TB or NOT TB Consulting), Megan Murray (Harvard Medical School)

*Pharmacovigilance and Safety*

For pharmacovigilance and safety reporting, a pharmacovigilance (PV) Unit was established. Safety data collection and transmission to the PV unit was the responsibility of the Site Principal Investigators and Site Co-Investigators. The PV Unit performed the medical review of each SAE and AESI reports. They evaluated each adverse event-investigational medicinal product pair: the expectedness and the Sponsor's assessment of the causality. Cases considered as potential Suspected Unexpected Serious Adverse Reactions (SUSAR) were directed to the Medical Review Board (MRB). The MRB comprised international DR-TB experts appointed by the PV Unit to review and confirm all potential SUSARs and assess the overall impact on participants. The PV Unit also provided support to the Clinical Advisory Committee (CAC) for safety-related eligibility questions, case management, medical monitoring, and permanent treatment discontinuation.

CAC members: Catherine Berry (Médecins Sans Frontières), Jennifer Furin (Case Western Reserve University), Ohanna Kirakosyan (Médecins Sans Frontières), Ilaria Motta (Médecins Sans Frontières)

#### 2.3 Participants

##### 2.3.1 Full Inclusion & Exclusion Criteria

**Inclusion Criteria**

1. Has documented pulmonary tuberculosis due to strains of *M. tuberculosis* resistant to rifampin and susceptible to fluoroquinolones, diagnosed by validated rapid molecular test;
2. Is ≥ 15 years of age;
3. Is willing to use contraception: pre-menopausal women or women whose last menstrual period was within the preceding year, who have not been sterilized, must agree to use contraception unless their partner has had a vasectomy; men who have not had a vasectomy must agree to use condoms;
4. Provides informed consent for study participation; additionally, a legal representative of patients considered minor per local laws should also provide consent;
5. Lives in a dwelling that can be located by study staff and expects to remain in the area for the duration of the study.

**Exclusion Criteria**

1. Has known allergies or hypersensitivity to any of the investigational drugs;
2. Is known to be pregnant or is unwilling or unable to stop breast-feeding an infant;
3. Is unable to comply with treatment or follow-up schedule;
4. Has any condition (social or medical) which, in the opinion of the site principal investigator, would make study participant unsafe;
5. a. Has had exposure (intake of the drug for 30 days or more) in the past five years to bedaquiline, delamanid, linezolid, or clofazimine, or has proven or likely resistance to bedaquiline, delamanid, linezolid, or clofazimine (e.g., household contact of a DR-TB index case who died or experienced treatment failure after treatment containing bedaquiline, delamanid, linezolid, or clofazimine or had resistance to one of the listed drugs); exposure to other anti-TB drugs is not a reason for exclusion.
   b. Has received second-line drugs for 15 days or more prior to screening visit date in the current MDR/RR-TB treatment episode. Exceptions include: (1) patients whose treatment has failed according to the WHO 2013

definition and who are being considered for a new treatment regimen; (2) patients starting a new treatment regimen after having been "lost to follow-up" according to the WHO 2013 definition and, (3) patients in whom treatment failure is suspected (but not confirmed according to WHO 2013 definition), who are being considered for a new treatment regimen, and for whom the Clinical Advisory Committee (CAC) consultation establishes eligibility.

6. Has one or more of the following:
    - Hemoglobin ≤ 7.9 g/dL;
    - Uncorrectable electrolytes disorders: o Total Calcium < 7.0 mg/dL (1.75 mmol/L);
        - Potassium < 3.0 mEq/L (3.0 mmol/L) or ≥6.0 mEq/L (6.0 mmol/L);
        - Magnesium < 0.9 mEq/L (0.45 mmol/L);
    - Serum creatinine > 3 x ULN;
    - Aspartate Aminotransferase (AST) or Alanine Aminotransferase (ALT) ≥ 3 x ULN;
    - Total bilirubin ≥ 3 x ULN;
    - Unless otherwise specified, Grade 4 result as defined by the MSF Severity Scale on any of the screening laboratory tests.
7. Has cardiac risk factors defined as:
    - An arithmetic average of the two ECGs with highest QTcF intervals of greater than or equal to 450 ms. Retesting to reassess eligibility will be allowed using an unscheduled visit during the screening phase;
    - Evidence of ventricular pre-excitation (e.g., Wolff Parkinson White syndrome);
    - Electrocardiographic evidence of either:
        - Complete left bundle branch block or right bundle branch block; OR
        - Incomplete left bundle branch block or right bundle branch block and QRS complex duration greater than or equal to 120 msec on at least one ECG;
    - Having a pacemaker implant;
    - Congestive heart failure;
    - Evidence of second- or third-degree heart block;
    - Bradycardia as defined by sinus rate less than 50 bpm;
    - Personal or family history of Long QT Syndrome;
    - Personal history of arrhythmic cardiac disease with the exception of sinus arrhythmia;
    - Personal history of syncope (i.e. cardiac syncope not including syncope due to vasovagal or epileptic causes).
8. Concurrent participation in another trial of any medication used or being studied for TB treatment, as defined in cited documents.
9. Is taking any medication that is contraindicated with the medicines in the trial regimen which cannot be stopped (with or without replacement) or requires a wash-out period longer than 2 weeks.

##### 2.3.2 Retention of pregnant women in the endTB clinical trial

Women who became pregnant during study participation and whose pregnancy was not terminated could remain on study treatment if all of the following conditions were met:

- The clinical trial insurance policy subscribed in the country covered pregnant participants, including damage to and loss of the fetus;
- The local authorities and ethics committee(s) approved;
- The participant was at least 18 years of age;
- The Site PI considered that the expected benefits of continued treatment outweighed the risks of ongoing fetal exposure;
- The CAC reviewed the Site PI's recommendation
- The participant was informed of the therapeutic options, potential risks and expected benefits and sperate specific consent was obtained.

#### 2.4 Randomization and treatment

##### 2.4.1 Bayesian response-adaptive randomization

The endTB trial employed Bayesian response-adaptive randomization because of a possible efficiency advantage in studies with more than one effective regimen.[1] Since endTB was exploring 5 experimental regimens and aimed to identify all of those that are effective, a design that permitted doing this as efficiently as possible was chosen.

Fixed, balanced randomization across the 6 treatment groups was planned for the first 180 participants. We used a single randomization list, which was generated in Stata by the trial statistician and uploaded to the central Web-based system (Venn Life Sciences, Dublin, Ireland, v2.5), which site staff used to assign treatment group. Bayesian, responsive-adaptive randomization started after 185 patients were randomized (see Figure S2 ) and used blocks of varying size. The adaptations were binding. The probability of assignment to each experimental arm then varied according to interim and end-of-treatment outcomes of participants progressing through the trial. The probability of assignment to the control arm was also controlled to approximately match the sample size of the highest enrolling experimental arm. The final sample size proportions were unknown prior to completion of randomization since they depended on the outcomes observed during the trial and changed over time. It was necessary to estimate such proportions anytime a new randomization list was generated.

Data used for adaptations (every 4 weeks):

- Week 8 culture results: culture negativity (TS8)
- Treatment outcome at Week 39: favorable outcome (TS39)

The algorithm used TS8 and TS39 to compute the posterior probability of non-inferiority to the control arm. The randomization probabilities favored allocation to the most promising regimens. For the control, the randomization probability was defined to approximately match the experimental arm with the highest number of enrolled patients.

TS73 (Favorable outcomes at week 73) was used to assess futility and/or superiority per pre-specified thresholds. Stopping an arm early would have been considered if the posterior probability of a regimen being non-inferior to the control at Week 73 fell below a threshold of 5%. Conversely, if the posterior probability was at least 95% that a regimen was superior to the control at Week 73, stopping for efficacy would have been considered. Results (whether or not the thresholds were met) were shared with data safety and monitoring board (DSMB). Had either threshold been met, the protocol specified that DSMB could further review safety and efficacy data and make a (non-binding) recommendation to stop an arm or the whole study. Neither threshold was ever met.

For TS8, any positive culture was dominant. The culture result was coded as follows:

- positive if either/both of 2 collected samples was positive
- negative if no positive and one sample was negative
- contaminated if at least one sample was contaminated and no sample was positive or negative culture
- missing, unable to produce sputum – if not able to produce sputum for either sample and there was no positive or negative culture
  missing sputum – not collected for other reasons

For combined results across 2 visits (e.g., Week 39 treatment outcome used Week 36 and Week 39 culture results), for each visit: the first sample collected was dominant if positive or negative; otherwise, the second collected sample was used; the result was contaminated in the absence of positive or negative cultures in the visit samples if at least one of them was contaminated. Then, the results of the two visits were combined as described below (for TS39 and imputation):

- positive if W36 (first in series) or W39 (second in series) was positive
- negative: if no positive and ≥1 negative
- contaminated: at least one contaminated and the other is contaminated or missing
- missing, unable to produce sputum – if not able to produce sputum for any sample at week 36 or week 39 and no positive or negative culture
- missing sputum – not collected for other reasons

Imputation was performed for TS8 and TS39 when culture results were contaminated or missing, unable to produce sputum. In addition, TS8 was imputed when TS8 result was missing and TS39 was not missing. We imputed missing Week 8 culture results to combined results at weeks 2 and 4. We imputed W36-39 culture result to combined results at weeks 32 and 36.

The trial statistician, who was responsible for analysis after database lock, did not perform the interim adaptive analysis. Instead, these were performed by the Bayesian statistician who updated the priors and the resulting randomization probabilities and communicated them to VennLife, from which site staff continued to assign treatment group. No central or site staff were informed of the randomization probabilities. They Bayesian statistician had no involvement in analysis after database lock.

##### 2.4.2 Treatment

Experimental arm treatment was prescribed for 9 months (39 weeks), 7 days/week. Weight-based dosing is in Table S6. One drug could be discontinued from experimental arm treatments with participants still considered to be on study treatment. No drugs could be added or substituted in experimental arms. Discontinuation of more than one drug or replacement/addition of any drugs in the experimental arms resulted in study treatment discontinuation (see *Permanent Discontinuation of Study Drug/Treatment*). For the control arm, regimen composition was guided by a study SOP, which in turn reflected current WHO guidance and local variations; these included the possibility of using of 9-month, all-oral "modified Bangladesh" regimens in India and South Africa in those whose prior exposure and known susceptibility were consistent with recommendations for its use. Otherwise, the longer regimen was used. No drugs could be added or substituted to the shorter, control regimen; one drug could be added or substituted to the longer regimen. The differences were because the longer, conventional regimen is considered to be an individualized regimen while experimental regimens and the modified Bangladesh regimens were considered to be standardized regimens and deviations from those regimens considered to be relevant to their evaluation.

Control regimen treatment guidance was changed twice, in conformity with WHO Guideline updates. When new guidance took effect, starting 18 Dec 2018, endTB trial study procedures were updated according to the following principles:

1. Participants on treatment for less than one month: initiate new regimen constructed according to WHO 2018 recommendations
2. Participants on treatment for more than one moth with good treatment response: replace any injectables (amikacin, capreomycin, or kanamycin) with a Group A drug whenever possible. If Group A and B drugs were not appropriate, replace injectables kanamycin and capreomycin with amikacin.
3. Participants on treatment for more than on month with signs of slow or no treatment response: consider restarting a new treatment according to current recommendations

On 31 March 2020, further recommended changes were made according to the following principles for the shorter, all-oral MDR/RR-TB regimen:

1. Participants at any point of treatment with signs of good treatment response: if still taking an injectable, consider replacing amikacin with bedaquiline whenever possible.
2. Participants at any point of treatment with signs of slow or no treatment response: consider restarting a new conventional treatment according to 2020 WHO recommendations (taking note that resistance may have been developed to some of the drugs used in the regimen).

Changes within a class—fluoroquinolone, thioamide or aminoglycoside/polypeptide—did not count as replacements.

As noted, the shorter MDR/RR-TB regimen was only offered as the control in countries where it was being used as the choice in the National TB Program for MDR/RR-TB.

Weight-based drug dosing was prescribed as recommended by WHO recommendations (see Table S7).

#### 2.5 Procedures

Study visits entailed: physical examination, symptom assessment, adverse event assessment, electrocardiograms, and blood and sputum (at least two per visit) sample collection at designated study facilities at each site. Treatment delivery/administration and sputum specimen collection could also occur in participants' homes or other place acceptable to participants and staff. Other study procedures could occur at these places or through remote visits during the COVID-19 pandemic or other in-country emergencies that limited movement of participants and staff. Hematology and biochemistry analyses were performed at designated clinical laboratories. Unscheduled laboratory tests were performed in other clinical laboratories as needed for participant safety. Phenotypic drug susceptibility testing was performed in Mycobacteria Growth Indicator Tube (MGIT) at baseline and on late positive cultures at Week 16 or later at designated microbiology laboratories at each study site. Specific tests are described in more detail below. Adverse events were reviewed at every study visit, with investigators soliciting reports of signs or symptoms; daily treatment support visits provided another opportunity to ask participants about any new or worsening conditions. If reported, treatment supporters communicated the information to investigators and facilitated referrals for evaluation.

*Sputum Specimen Testing*

At least 2 sputa (spot or early morning) were collected at each visit at which sputum collection was indicated. When cultures were indicated, at least one was performed in MGIT; the other was performed in Löwenstein-Jensen medium except at the study lab in South Africa (National Institute for Communicable Diseases, which tested for both South Africa and Lesotho sites). During implementation of the endTB trial, this lab did not perform culture *M. tuberculosis* in Löwenstein-Jensen and therefore both cultures were performed in MGIT.

*Recording and Central Overreading of ECGs*

At screening and baseline visits (Visits 1 and 2), three ECGs were taken five minutes apart for each participant and recorded. At subsequent study visits (Visit 3-24; 27, and Early Termination, if applicable), two ECGs were taken five minutes apart and recorded. At screening and baseline, the ECG with the longest QT interval was sent to Clario (formerly eResearch Technologies) for central reading. For follow-up visit ECGs (Visits 3, 5, 7, 10, 15, 17, 19, 21, and 27), one ECG of best quality was sent for routine central reading. Clario overreads were also available for ECGs taken at unscheduled visits, as needed for safety.

#### 2.6 Follow-up and Outcomes

##### 2.6.1 Follow-up

Trial participation in all arms lasted until at least Week 73 and up to Week 104. Not all participants would have a study visit after Week 73. Sites conducted as many visits as possible until the end of the Week 73 window of the last participant randomized into the study. This "hybrid" follow-up option was chosen to balance the benefit of 2-year follow-up to capture data on relapse and the benefit of reporting trial results sooner.[2]

##### 2.6.2 Primary Efficacy Outcome

The primary efficacy outcome was the proportion of participants with favorable outcome at Week 73.

A participant's outcome was classified as favorable at Week 73 if the outcome as not classified as unfavorable, and one of the following was true:

- The last two culture results are negative. These two cultures must be taken from sputum samples collected on separate visits, the latest between Week 65 and Week 73;
- The last culture result (from a sputum sample collected between Weeks 65 and 73) is negative; and either there is no other post-baseline culture result or the penultimate culture result is positive due to laboratory cross contamination; and bacteriological, radiological and clinical evolution is favorable;

- There is no culture result from a sputum sample collected between Week 65 and Week 73 or the result of that culture is positive due to laboratory cross contamination; and the most recent culture result is negative; and bacteriological, radiological and clinical evolution is favorable.

A participant's outcome was classified as unfavorable at Week 73 in case of any of the following:

1. Replacement or addition of one or more investigational drugs in an experimental arm or in the control arm if using the shortened regimen (failure);
2. Replacement or addition of two or more investigational drugs in the control arm if using the conventional regimen (failure);
3. Initiation of a new MDR-TB treatment regimen after the end of the allocated study regimen and before Week 73 (relapse);
4. Death from any cause;
5. At least one of the last two cultures, the latest being from a sputum sample collected between Week 65 and Week 73, is positive in the absence of evidence of laboratory cross contamination (failure/relapse);
6. The last culture result (from a sputum sample collected between Week 65 and Week 73) is negative; AND
    - there is no other post-baseline culture result or the penultimate culture is positive due to laboratory cross contamination; and bacteriological, radiological or clinical evolution is unfavorable (failure/relapse);
7. There is no culture result from a sputum sample collected between Week 65 and Week 73 or it is positive due to laboratory cross contamination; AND
    - the most recent culture is negative; and bacteriological, radiological or clinical evolution is unfavorable (failure/relapse); or
    - -the most recent culture result is positive in the absence of laboratory cross contamination
8. The outcome is not assessable because there is no culture result from a sputum sample collected between Week 65 and Week 73 or it is positive due to laboratory cross contamination; AND
    - there is no other post-baseline culture result or the most recent culture is positive due to laboratory cross contamination; or,
    - the most recent culture is negative and bacteriological, radiological and clinical evolution is not assessable;
9. Previously classified as unfavorable in the present study[i]

[i] Exception: A participant whose outcome was unfavorable because it was unassessable at Week 39 is eligible for re-evaluation at Week 73.

##### 2.6.3 Secondary Efficacy Outcome

The secondary efficacy outcome was the proportion of participants with favorable outcome at Week 104.

A participant's outcome was classified as favorable at Week 104 if the outcome was not classified as unfavorable, and one of the following was true:

a) The last two cultures are negative. These two cultures must be from sputum samples collected on separate visits, the latest between Week 97 and Week 104;
b) The last culture result (from a sputum sample collected between Week 97 and Week 104) is negative; and either there is no other post-baseline culture result or the penultimate culture result is positive due to laboratory cross contamination; and bacteriological, radiological and clinical evolution is favorable;
c) There is no culture result from a sputum sample collected between Week 97 and Week 104 or the result of that culture is positive due to laboratory cross contamination; and the most recent culture result is negative; and bacteriological, radiological and clinical evolution is favorable.

A participant's outcome was classified as unfavorable at Week 104 in case of any of the following:

a) Replacement or addition of one or more investigational drugs in the experimental arm or in the control arm if using the shortened regimen (failure);
b) Replacement or addition of two or more investigational drugs in the control arm if using the conventional regimen (failure);
c) Initiation of a new MDR-TB treatment regimen after the end of the allocated study regimen and before Week 104 (relapse);
d) Death from any cause;
e) At least one of the last two cultures, the latest being from a sputum sample collected between Week 97 and Week 104, is positive in the absence of evidence of laboratory cross contamination (failure/relapse);
f) The last culture result (from a sputum sample collected between Week 97 and Week 104) is negative; ***AND***
   - there is no other post-baseline culture result or the penultimate culture is positive due to laboratory cross contamination; and bacteriological, radiological or clinical evolution is unfavorable (failure/relapse).
g) There is no culture result from a sputum sample collected between Week 97 and Week 104 or it is positive due to laboratory cross contamination; ***AND***
   - the most recent culture is negative; and bacteriological, radiological or clinical evolution is unfavorable (failure/relapse);
h) There is no culture result from a sputum sample collected between Week 97 and Week 104 or it is positive due to laboratory cross contamination; ***AND***
   - there is no other post-baseline culture or it is positive; or,
   - the most recent culture is negative and bacteriological, radiological and clinical evolution is not assessable.
i) Previously classified as unfavorable in the present study;[ii]
j) Loss to follow-up[iii]

*Event Adjudication*

In order to ensure the accuracy of the treatment outcome endpoints in the study and limit the variability across sites, an event adjudication process was established by which the CAC reviewed and validated the treatment outcomes assigned by the site investigators. An automated assignment algorithm, corresponding to the treatment outcome definitions, was programmed in Stata. During event adjudication, the CAC reviewed the treatment outcomes that were discordant between the site investigator and the algorithm as well as all those that had been established based on bacteriological,

[ii] Exception: a participant whose outcome is unfavorable because it is unassessable at Week 73 is eligible for re-evaluation at Week 104.

[iii] Loss to follow up is defined as participants for whom study staff have no information and whom they have been unable to contact for more than 14 weeks before last study visit per study schedule.

radiological and clinical evolution. Discrepancies between the treatment outcome emerging from CAC validation and site-investigator-assigned treatment outcome were discussed with site investigators. If the discrepancy was determined to be due data incorrectly entered into the database or the site investigator changed their opinion to agree with the algorithm, data was modified, and the final outcome was reported as concordant.

*Severity grading*

The MSF Severity Scale was derived primarily from Division of Microbiology and Infectious Disease Adult Toxicity Table (Nov 2007) and Common Terminology Criteria for Adverse Events (v.4.03 14-Jun-2010).

*Adverse Events of Special Interest*

Adverse Events of Special Interest (AESI) could be serious or non-serious and were defined as a medically significant event that had the potential to be causally associated with a study drug and needed to be carefully monitored by the Sponsor. For the endTB trial, pre-determined AESI were grade 3 or higher of the following: leukopenia, anemia, thrombocytopenia, prolonged QT (average QTcF >500 ms or >60 ms change from baseline together with signs/symptoms of serious arrhythmia), optic neuritis, peripheral neuropathy, or hepatic toxicity.

*Permanent Discontinuation of Study Drug/Treatment (see also Treatment)*

Permanent discontinuation of study drug(s) was advised if a participant insisted on stopping study treatment. Additionally, permanent discontinuation of study drug(s) was considered on a case-by-case basis depending on the following circumstances: a) requirement for prohibited concomitant medications; b) any condition (social or medical) which, in the opinion of the site PI, made the study participant unsafe; c) indications of treatment failure including positive culture at Week 16 or later; or d) enrolled participant became pregnant (see *Retention in study of pregnant women* in Inclusion/Exclusion section). Permanent discontinuation of study drug(s) could also occur if a grade 3 or higher adverse event was considered related to the study drug(s) and did not resolve within 14 days. Any permanent discontinuation of a drug or treatment was discussed with the CAC and classification of permanent discontinuation required prior discussion with, and input from, the CAC.

#### 2.7 Analysis

##### 2.7.1 Secondary analysis populations

All-culture mITT includes participants without pre-randomization cultures positive for *M. tuberculosis*. All-DST mITT includes participants with isolates resistant to bedaquiline, clofazimine, delamanid, fluoroquinolone, and/or linezolid. The assessable population includes participants from the PP population whose outcomes were classified as favorable or unfavorable and excludes those whose outcomes were unfavorable because they were not assessable. Also excluded from the assessable population are participants who experienced voluntary withdrawal, loss to follow up (LTFU), or confirmed reinfection. A fourth analysis population was created for sensitivity analyses. This “Assessable Reclassified” population is defined as the assessable population in which outcomes of “LTFU” or “Unfavorable because unassessable” are assigned the outcome that had been assigned at Week 39. Distinct populations were created for analyses of Week 73 and Week 104 endpoints.

*Accounting for Multiplicity*

We prespecified the hierarchical testing or fixed-sequence approach. The regimen with the highest proportion of favorable outcomes in the mITT population was compared to the control first; if non-inferiority was established, then a comparison between the control and the experimental regimen having the second highest proportion of favorable outcomes was performed, and continued as long as non-inferiority was concluded in the mITT population. The same process was conducted in the PP population. All comparisons were done at the full alpha level.

##### 2.7.2 Pre-specified adjusted, subgroup, sensitivity, and post-hoc adjusted analyses

Poisson regression model with robust standard errors was used for adjusted and subgroup analyses in light of convergence failures with binomial regression models. Adjusted estimate of risk difference with corresponding 95% confidence interval was calculated. In prespecified analyses, country, BMI, pyrazinamide resistance, injectable resistance, age, sex, presence of each comorbidity at baseline or during study participation (including HIV, hepatitis B and C status, diabetes, SARS-Cov-2 infection and COVID-19), smear result, lung cavitation presence at baseline, prior exposure to TB treatment, extent of disease (limited/moderate/extensive, based on chest X-ray at baseline) were considered as covariates. Any covariate that was significant ($p<0.10$) in the univariate analysis was introduced into a multivariable model and through a process of backward elimination only significant variables with $p<0.05$ were retained in the final model.

*Subgroup analyses*

Sub-group analyses of the primary efficacy endpoint were performed (in both mITT and PP populations) for regimens that were shown to be non-inferior. Subgroups were established on the following baseline characteristics:

- Country (Georgia, India, Kazakhstan, Lesotho, Pakistan, Peru, South Africa)
- Presence of comorbidities (presence/absence of: HIV, hepatitis C, diabetes, and SARS-CoV-2 infection/COVID-19)
- BMI (<18.5 kg/m2, 18.5-24.9 kg/m2, ≥25 kg/m2)
- Pyrazinamide resistance (sensitive/resistant)
- Age (<18, 18-45, ≥45)
- Sex (male/female)
- Smear result (at screening) (positive/negative)
- Cavitation (presence/absence)
- Prior exposure to TB treatment (none/first line only/other)
- WHO recommendations implemented (by implementation date).

*Sensitivity analyses*

The primary unadjusted and adjusted efficacy analyses were performed:

- in the following populations: 1) assessable, 2) all-culture mITT, and 3) all-DST mITT populations.
- in the assessable population reclassifying outcomes of "LTFU" or "unfavorable because unassessable" as the outcome that had been assigned at Week 39

We did not perform the following planned sensitivity analyses:

- excluding participants from the control arm who received a shortened regimen, because the proportion did not exceed the pre-defined threshold of 10%
- using multiple imputation to impute missing data after examination of the missing data pattern, because the proportion did not exceed the pre-defined threshold of 20%.

*Post-hoc adjusted analyses*

Because there was some variability in distribution of covariates observed, we performed a second set of adjusted analyses to evaluate confounding. Covariates included in the multivariable model were pyrazinamide resistance, HIV status, sputum smear microscopy grade, and cavitation on chest X-ray.

### 3 Results

First patient, first visit occurred on 22 February 2017; last patient, last visit was completed on 08 April 2023 when the last participant completed 73 weeks of post-randomization follow-up.

We report the distribution of the use of individual drugs and regimens in the control group by country (Table S10). Small differences can be observed in the use or choice of injectables, delamanid, and ethionamide/prothionamide. However, almost two-thirds (77 out of 119) of the participants received regimens containing bedaquiline, clofazimine, linezolid, and levofloxacin and another 5 (4.2%) received regimens containing bedaquiline, clofazimine, linezolid, and moxifloxacin. That a large number of participants received at least 3 Group A and 1 Group B drugs reinforces both the strength of the control arm and its relative uniformity. Changes over time can be observed in Table S11. Control group participants received regimens containing an injectable agent during only the first WHO Guideline period (prior to 18 Dec 2018). These 22 participants (18.5% of all control group participants) were the only ones to receive regimens that were not compliant with 2022 Guidelines. (See *Treatment*)

##### 3.1.1 Safety

*Deaths*

The most common causes of death were disease progression (5 participants) and sepsis (3 participants) [Table S26]. Two participants were found dead at home: sudden death could not be excluded although it was considered unlikely by study investigators and medical review committee.

*Permanent Drug Stops due to adverse events*

Permanent drug stops due to AEs occurred in 42.9% of participants in the control and in between 18.3% (9DCMZ) and 32.3% (9BDLLfxZ) in the experimental groups (Table S27). The most frequently stopped drugs were pyrazinamide (121 [16.2%] participants at median 92 [IQR: 53-188] days post randomization) and linezolid (84 [11.3%] participants at median 155.5 [IQR: 83.5-224.5] days post randomization).

*Figure S1. endTB Trial Design*

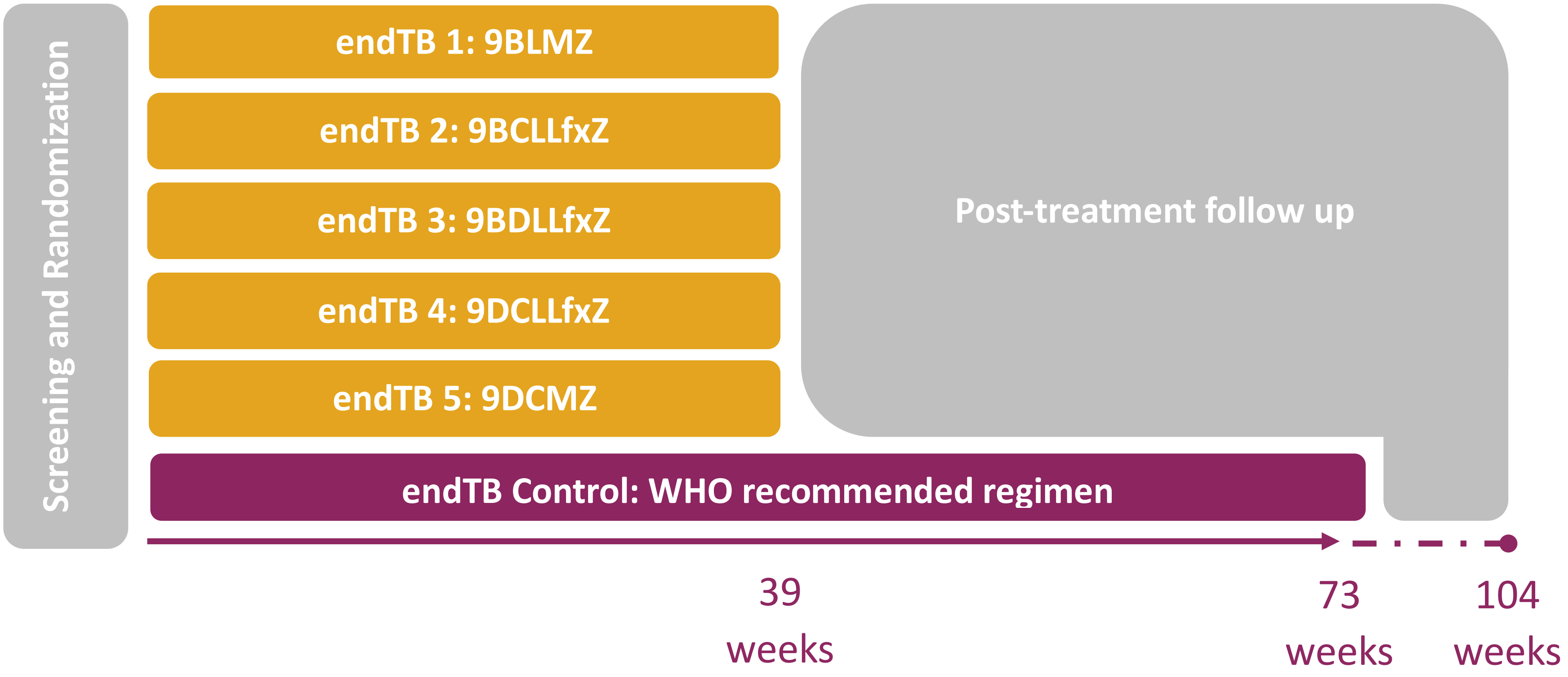

***Figure S2. Allocation ratio through recruitment***

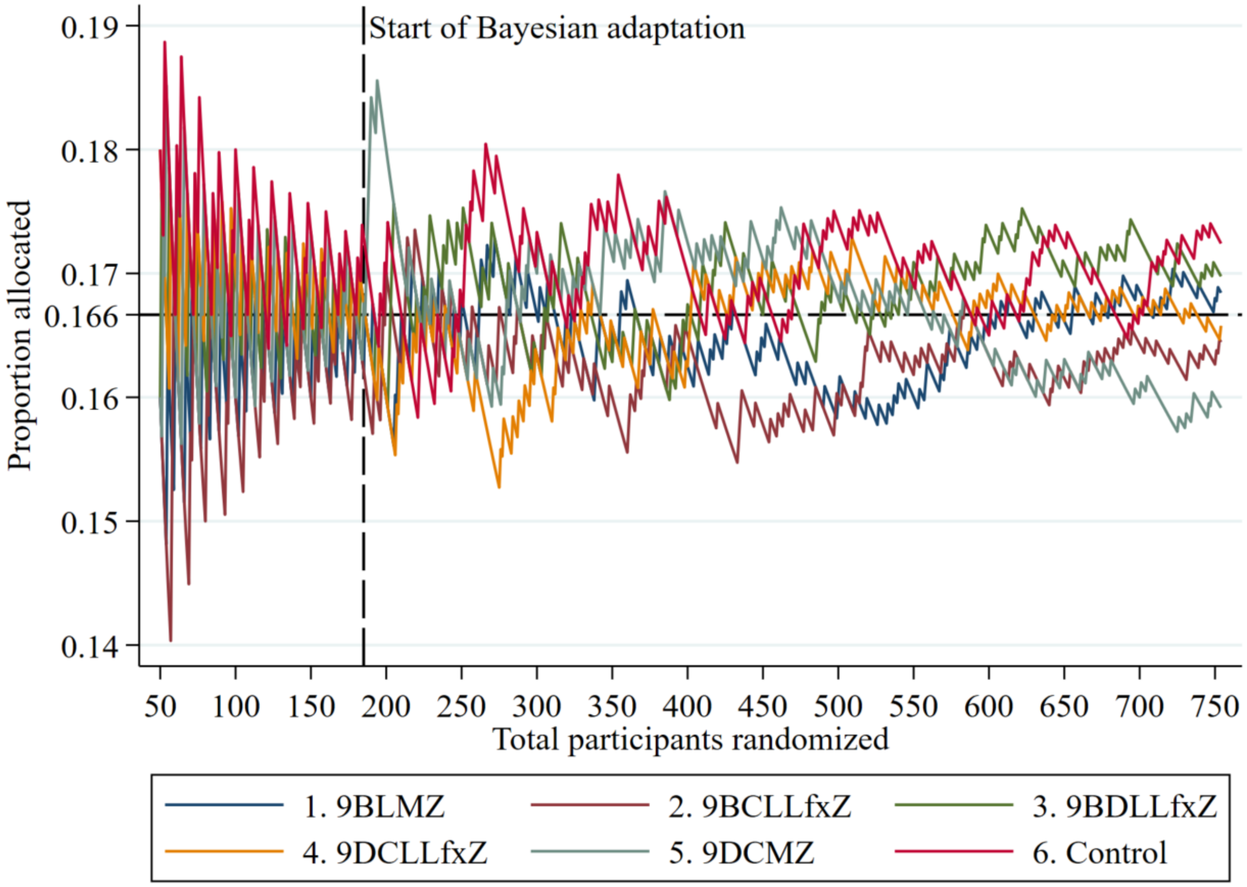

*Figure S3.  CONSORT Group allocation during fixed and adaptive periods*

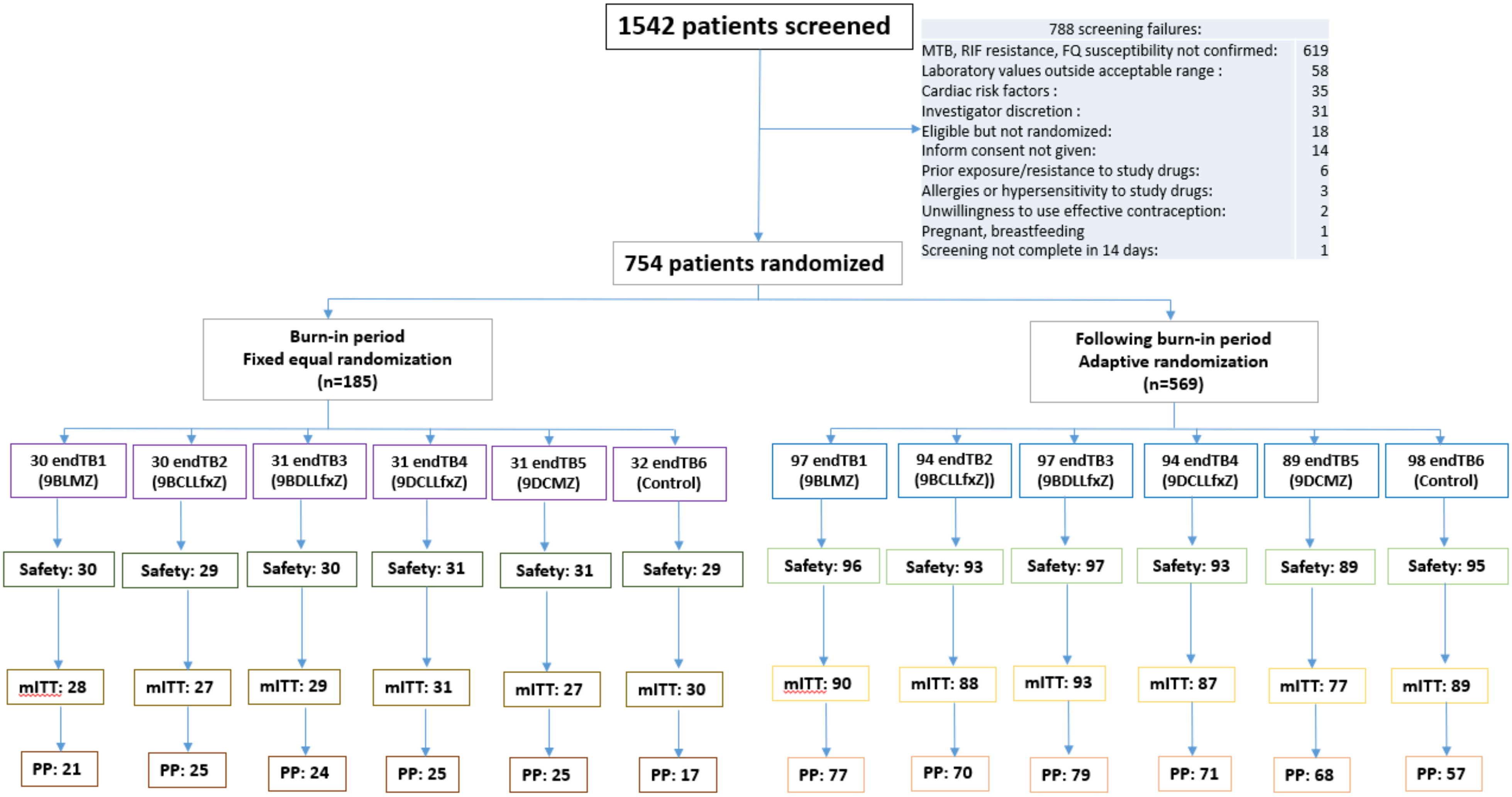

*Figure S4a. Subgroup Analysis of Efficacy of endTB Trial Regimen 9BLMZ vs Control at Week 73 (mITT population)*

| | 9BLMZ | Control | RD (95% CI) |
|---|---|---|---|
| **mITT** | **105/118 (89.0%)** | **96/119 (80.7%)** | **8.3% [-0.8%;17.4%]** |
| **Country** | | | |
| Georgia | 1/2 (50.0%) | 2/3 (66.7%) | -16.7% [-114.4%;81.1%] |
| India | 7/8 (87.5%) | 3/4 (75.0%) | 12.5% [-37.9%;62.9%] |
| Kazakhstan | 27/30 (90.0%) | 19/23 (82.6%) | 7.4% [-11.6%;26.4%] |
| Lesotho | 14/14 (100.0%) | 12/12 (100.0%) | – |
| Pakistan | 17/18 (94.4%) | 17/18 (94.4%) | 0.0% [-15.2%;15.2%] |
| Peru | 35/38 (92.1%) | 36/51 (70.6%) | 21.5% [6.3%;36.8%] |
| South Africa | 4/8 (50.0%) | 7/8 (87.5%) | -37.5% [-80.4%;5.4%] |
| **HIV** | | | |
| Negative | 91/103 (88.3%) | 78/99 (78.8%) | 9.6% [-0.6%;19.7%] |
| Positive | 14/15 (93.3%) | 17/19 (89.5%) | 3.9% [-15.1%;22.8%] |
| **Hepatitis C** | | | |
| Negative | 102/113 (90.3%) | 93/113 (82.3%) | 8.0% [-1.0%;16.9%] |
| Positive | 3/5 (60.0%) | 3/6 (50.0%) | 10.0% [-51.6%;71.6%] |
| **Diabetes** | | | |
| No | 86/99 (86.9%) | 86/104 (82.7%) | 4.2% [-5.7%;14.1%] |
| Yes | 19/19 (100.0%) | 10/15 (66.7%) | 33.3% [9.1%;57.5%] |
| **BMI** | | | |
| <18.5 | 36/41 (87.8%) | 35/39 (89.7%) | -1.9% [-15.8%;12.0%] |
| 18.5-24.9 | 57/65 (87.7%) | 47/62 (75.8%) | 11.9% [-1.5%;25.3%] |
| ≥25 | 12/12 (100.0%) | 14/18 (77.8%) | 22.2% [2.7%;41.8%] |
| **Sex** | | | |
| Female | 36/41 (87.8%) | 42/48 (87.5%) | 0.3% [-13.5%;14.1%] |
| Male | 69/77 (89.6%) | 54/71 (76.1%) | 13.6% [1.5%;25.6%] |
| **Age** | | | |
| <18 | 1/2 (50.0%) | 5/6 (83.3%) | -33.3% [-114.0%;47.3%] |
| 18-44 | 84/93 (90.3%) | 75/91 (82.4%) | 7.9% [-2.0%;17.8%] |
| ≥45 | 20/23 (87.0%) | 16/22 (72.7%) | 14.2% [-9.2%;37.6%] |
| **Pyrazinamide** | | | |
| Sensitive | 57/61 (93.4%) | 44/57 (77.2%) | 16.2% [3.7%;28.8%] |
| Resistant | 48/57 (84.2%) | 49/59 (83.1%) | 1.2% [-12.4%;14.7%] |
| **Smear results** | | | |
| Negative/scanty | 16/20 (80.0%) | 13/19 (68.4%) | 11.6% [-16.1%;39.2%] |
| 1-2+ | 54/57 (94.7%) | 43/52 (82.7%) | 12.0% [0.2%;23.9%] |
| 3+ | 35/41 (85.4%) | 40/48 (83.3%) | 2.0% [-13.2%;17.2%] |
| **Cavitation** | | | |
| Absence | 43/50 (86.0%) | 31/43 (72.1%) | 13.9% [-2.7%;30.5%] |
| Presence | 62/68 (91.2%) | 64/75 (85.3%) | 5.8% [-4.7%;16.3%] |
| **Prior exposure TB treatment** | | | |
| None | 68/76 (89.5%) | 59/74 (79.7%) | 9.7% [-1.8%;21.2%] |
| First line only | 20/20 (100.0%) | 24/31 (77.4%) | 22.6% [7.7%;37.4%] |
| Second line | 10/15 (66.7%) | 10/11 (90.9%) | -24.2% [-54.1%;5.6%] |
| **WHO recommendations** | | | |
| <18/12/2018 | 24/30 (80.0%) | 22/31 (71.0%) | 9.0% [-12.6%;30.7%] |
| 18/12/2018-31/03/2020 | 33/36 (91.7%) | 28/35 (80.0%) | 11.7% [-4.5%;27.8%] |
| ≥ 01/04/2020 | 48/52 (92.3%) | 46/53 (86.8%) | 5.5% [-6.2%;17.2%] |

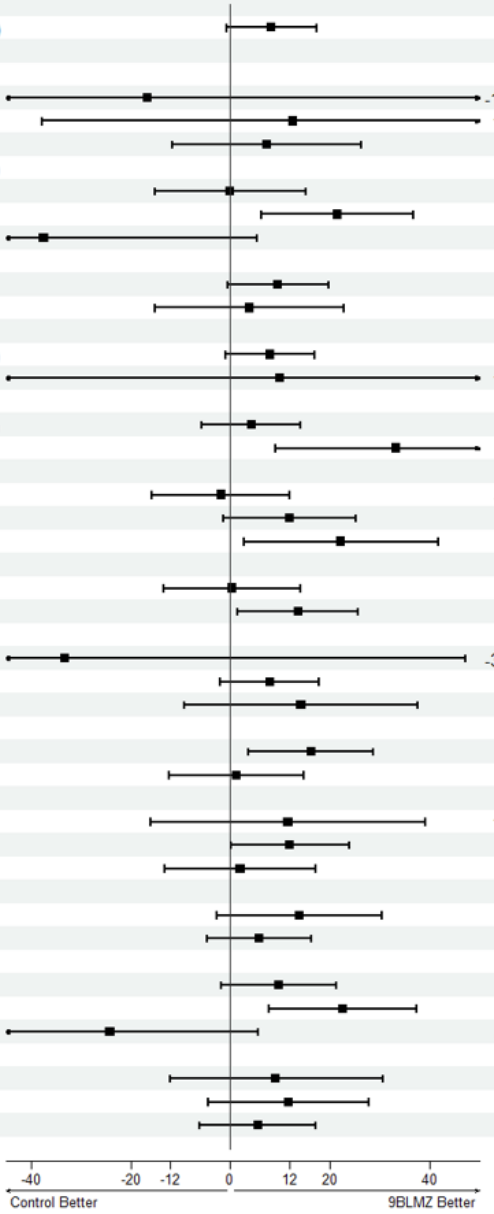

***Figure S4b. Subgroup Analysis of Efficacy of endTB Trial Regimen 9BCLLfxZ vs Control at Week 73 (mITT Population)***

| | 9BCLLfxZ | Control | RD (95% CI) |
|---|---|---|---|
| **mITT** | 104/115 (90.4%) | 96/119 (80.7%) | 9.8% [0.9%;18.7%] |
| **Country** | | | |
| Georgia | 2/2 (100.0%) | 2/3 (66.7%) | 33.3% [-26.3%;93.0%] |
| India | 4/4 (100.0%) | 3/4 (75.0%) | 25.0% [-20.4%;70.4%] |
| Kazakhstan | 31/35 (88.6%) | 19/23 (82.6%) | 6.0% [-12.9%;24.9%] |
| Lesotho | 11/11 (100.0%) | 12/12 (100.0%) | |
| Pakistan | 15/16 (93.8%) | 17/18 (94.4%) | -0.7% [-16.8%;15.4%] |
| Peru | 37/39 (94.9%) | 36/51 (70.6%) | 24.3% [9.9%;38.7%] |
| South Africa | 4/8 (50.0%) | 7/8 (87.5%) | -37.5% [-80.4%;5.4%] |
| **HIV** | | | |
| Negative | 90/101 (89.1%) | 78/99 (78.8%) | 10.3% [0.2%;20.4%] |
| Positive | 14/14 (100.0%) | 17/19 (89.5%) | 10.5% [-3.5%;24.5%] |
| **Hepatitis C** | | | |
| Negative | 100/109 (91.7%) | 93/113 (82.3%) | 9.4% [0.7%;18.2%] |
| Positive | 3/5 (60.0%) | 3/6 (50.0%) | 10.0% [-51.6%;71.6%] |
| **Diabetes** | | | |
| No | 86/96 (89.6%) | 86/104 (82.7%) | 6.9% [-2.6%;16.4%] |
| Yes | 17/18 (94.4%) | 10/15 (66.7%) | 27.8% [1.3%;54.3%] |
| **BMI** | | | |
| <18.5 | 27/33 (81.8%) | 35/39 (89.7%) | -7.9% [-24.3%;8.4%] |
| 18.5-24.9 | 57/61 (93.4%) | 47/62 (75.8%) | 17.6% [5.2%;30.0%] |
| ≥25 | 20/21 (95.2%) | 14/18 (77.8%) | 17.5% [-4.1%;39.0%] |
| **Sex** | | | |
| Female | 34/37 (91.9%) | 42/48 (87.5%) | 4.4% [-8.5%;17.3%] |
| Male | 70/78 (89.7%) | 54/71 (76.1%) | 13.7% [1.7%;25.7%] |
| **Age** | | | |
| <18 | 3/3 (100.0%) | 5/6 (83.3%) | 16.7% [-15.0%;48.3%] |
| 18-44 | 65/71 (91.5%) | 75/91 (82.4%) | 9.1% [-1.1%;19.3%] |
| ≥45 | 36/41 (87.8%) | 16/22 (72.7%) | 15.1% [-6.2%;36.4%] |
| **Pyrazinamide** | | | |
| Sensitive | 48/52 (92.3%) | 44/57 (77.2%) | 15.1% [2.0%;28.3%] |
| Resistant | 56/63 (88.9%) | 49/59 (83.1%) | 5.8% [-6.5%;18.2%] |
| **Smear results** | | | |
| Negative/scanty | 18/19 (94.7%) | 13/19 (68.4%) | 26.3% [2.8%;49.8%] |
| 1-2+ | 55/59 (93.2%) | 43/52 (82.7%) | 10.5% [-1.6%;22.7%] |
| 3+ | 31/37 (83.8%) | 40/48 (83.3%) | 0.5% [-15.5%;16.4%] |
| **Cavitation** | | | |
| Absence | 43/46 (93.5%) | 31/43 (72.1%) | 21.4% [6.1%;36.7%] |
| Presence | 61/69 (88.4%) | 64/75 (85.3%) | 3.1% [-8.0%;14.1%] |
| **Prior exposure TB treatment** | | | |
| None | 62/67 (92.5%) | 59/74 (79.7%) | 12.8% [1.7%;24.0%] |
| First line only | 20/23 (87.0%) | 24/31 (77.4%) | 9.5% [-10.8%;29.9%] |
| Second line | 18/19 (94.7%) | 10/11 (90.9%) | 3.8% [-16.2%;23.9%] |
| **WHO recommendations** | | | |
| <18/12/2018 | 24/28 (85.7%) | 22/31 (71.0%) | 14.7% [-6.0%;35.5%] |
| 18/12/2018-31/03/2020 | 31/33 (93.9%) | 28/35 (80.0%) | 13.9% [-1.7%;29.6%] |
| ≥ 01/04/2020 | 49/54 (90.7%) | 46/53 (86.8%) | 3.9% [-8.1%;16.0%] |

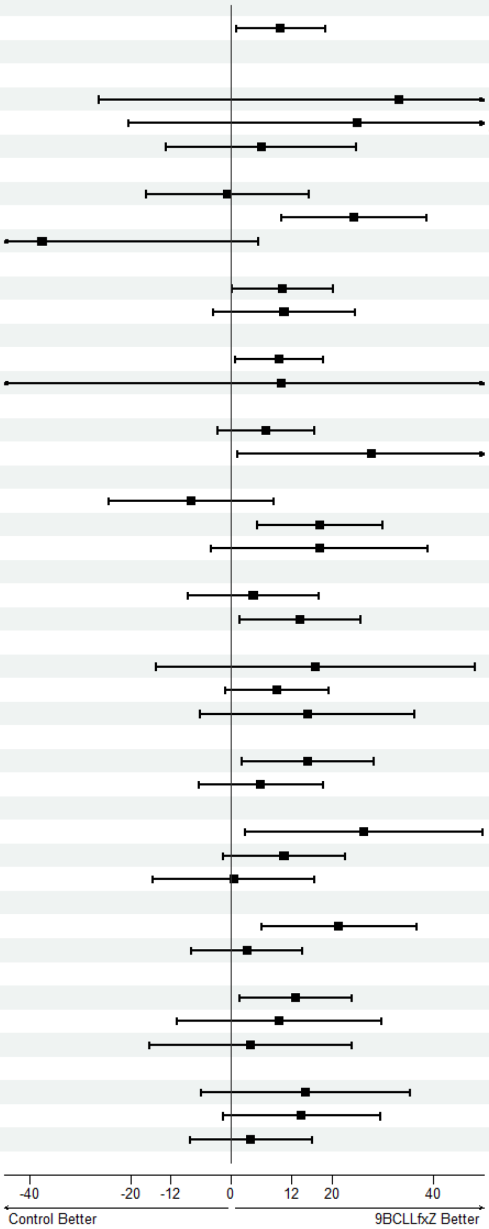

***Figure S4c. Subgroup Analysis of Efficacy of endTB Trial Regimen 9BDLLfxZ vs Control at Week 73 (mITT Population***

| | 9BDLLfxZ | Control | RD (95% CI) |
|---|---|---|---|
| **mITT** | **104/122 (85.2%)** | **96/119 (80.7%)** | **4.6% [-4.9%;14.1%]** |
| **Country** | | | |
| Georgia | 1/1 (100.0%) | 2/3 (66.7%) | 33.3% [-28.3%;94.9%] |
| India | 2/3 (66.7%) | 3/4 (75.0%) | -8.3% [-82.0%;65.3%] |
| Kazakhstan | 29/33 (87.9%) | 19/23 (82.6%) | 5.3% [-14.0%;24.5%] |
| Lesotho | 11/15 (73.3%) | 12/12 (100.0%) | -26.7% [-49.5%;-3.9%] |
| Pakistan | 10/13 (76.9%) | 17/18 (94.4%) | -17.5% [-43.2%;8.1%] |
| Peru | 44/49 (89.8%) | 36/51 (70.6%) | 19.2% [4.0%;34.4%] |
| South Africa | 7/8 (87.5%) | 7/8 (87.5%) | 0.0% [-33.5%;33.5%] |
| **HIV** | | | |
| Negative | 92/105 (87.6%) | 78/99 (78.8%) | 8.8% [-1.4%;19.1%] |
| Positive | 12/17 (70.6%) | 17/19 (89.5%) | -18.9% [-44.9%;7.2%] |
| **Hepatitis C** | | | |
| Negative | 102/119 (85.7%) | 93/113 (82.3%) | 3.4% [-6.0%;12.9%] |
| Positive | 2/3 (66.7%) | 3/6 (50.0%) | 16.7% [-54.1%;87.4%] |
| **Diabetes** | | | |
| No | 87/101 (86.1%) | 86/104 (82.7%) | 3.4% [-6.5%;13.4%] |
| Yes | 16/20 (80.0%) | 10/15 (66.7%) | 13.3% [-16.7%;43.4%] |
| **BMI** | | | |
| <18.5 | 20/28 (71.4%) | 35/39 (89.7%) | -18.3% [-37.7%;1.1%] |
| 18.5-24.9 | 72/81 (88.9%) | 47/62 (75.8%) | 13.1% [0.4%;25.8%] |
| ≥25 | 12/13 (92.3%) | 14/18 (77.8%) | 14.5% [-9.9%;39.0%] |
| **Sex** | | | |
| Female | 46/55 (83.6%) | 42/48 (87.5%) | -3.9% [-17.5%;9.7%] |
| Male | 58/67 (86.6%) | 54/71 (76.1%) | 10.5% [-2.4%;23.4%] |
| **Age** | | | |
| <18 | 5/5 (100.0%) | 5/6 (83.3%) | 16.7% [-14.6%;47.9%] |
| 18-44 | 71/86 (82.6%) | 75/91 (82.4%) | 0.1% [-11.1%;11.4%] |
| ≥45 | 28/31 (90.3%) | 16/22 (72.7%) | 17.6% [-3.9%;39.1%] |
| **Pyrazinamide** | | | |
| Sensitive | 50/56 (89.3%) | 44/57 (77.2%) | 12.1% [-1.5%;25.7%] |
| Resistant | 54/66 (81.8%) | 49/59 (83.1%) | -1.2% [-14.6%;12.2%] |
| **Smear results** | | | |
| Negative/scanty | 28/31 (90.3%) | 13/19 (68.4%) | 21.9% [-1.7%;45.5%] |
| 1-2+ | 47/58 (81.0%) | 43/52 (82.7%) | -1.7% [-16.1%;12.8%] |
| 3+ | 29/33 (87.9%) | 40/48 (83.3%) | 4.5% [-10.9%;20.0%] |
| **Cavitation** | | | |
| Absence | 38/48 (79.2%) | 31/43 (72.1%) | 7.1% [-10.7%;24.8%] |
| Presence | 65/73 (89.0%) | 64/75 (85.3%) | 3.7% [-7.1%;14.5%] |
| **Prior exposure TB treatment** | | | |
| None | 70/78 (89.7%) | 59/74 (79.7%) | 10.0% [-1.4%;21.4%] |
| First line only | 22/27 (81.5%) | 24/31 (77.4%) | 4.1% [-16.9%;25.0%] |
| Second line | 11/15 (73.3%) | 10/11 (90.9%) | -17.6% [-46.2%;11.1%] |
| **WHO recommendations** | | | |
| <18/12/2018 | 25/30 (83.3%) | 22/31 (71.0%) | 12.4% [-8.6%;33.4%] |
| 18/12/2018-31/03/2020 | 34/38 (89.5%) | 28/35 (80.0%) | 9.5% [-7.1%;26.0%] |
| ≥ 01/04/2020 | 45/54 (83.3%) | 46/53 (86.8%) | -3.5% [-17.0%;10.1%] |

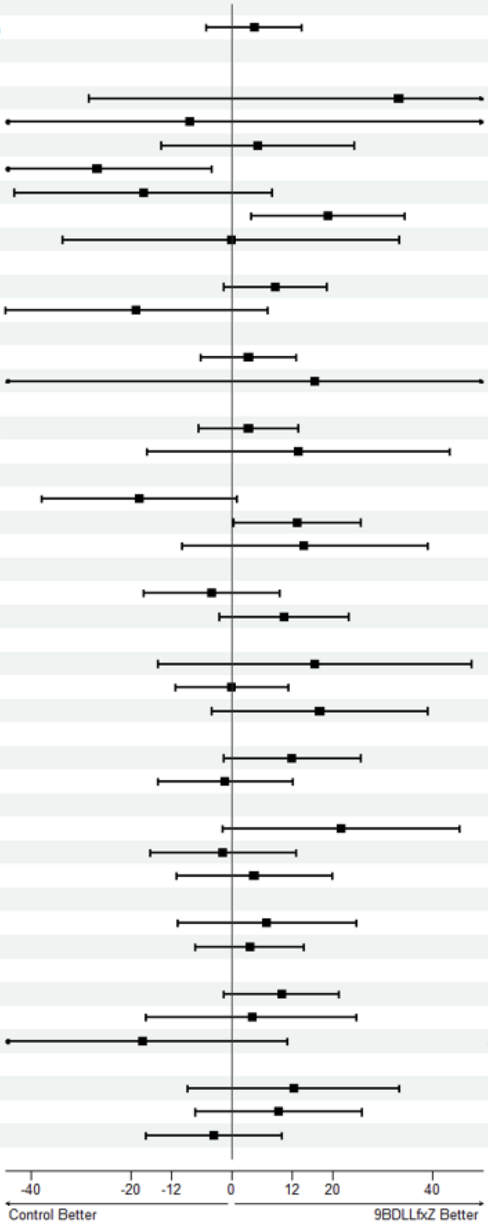

*Figure S4d. Subgroup Analysis of Efficacy of endTB Trial Regimen 9DCLLfxZ vs Control at Week 73 (mITT Population)*

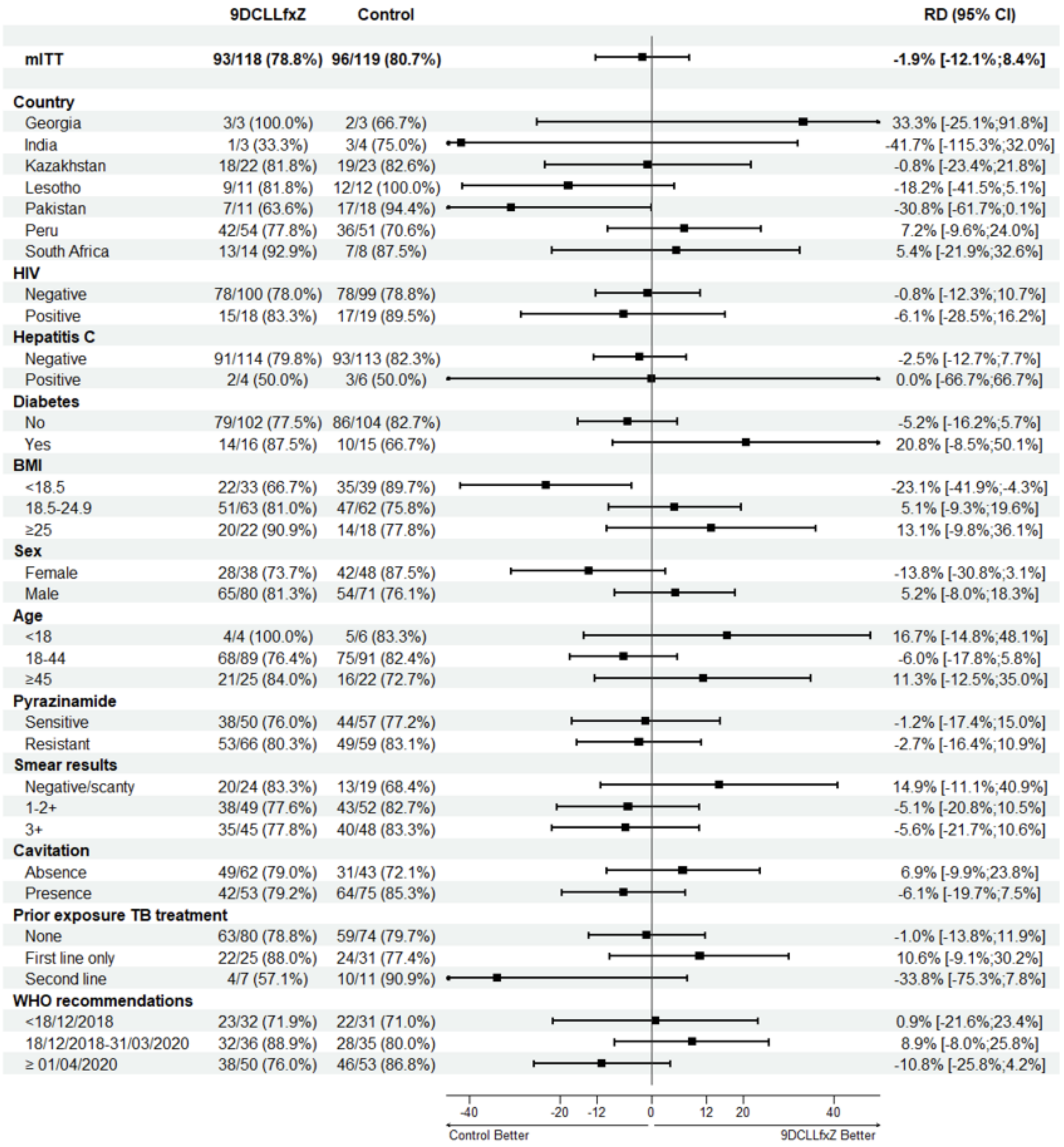

*Figure S4e. Subgroup Analysis of Efficacy of endTB Trial Regimen 9DCMZ vs Control at Week 73 (mITT Population)*

| | 9DCMZ | Control | RD (95% CI) |
|---|---|---|---|
| **mITT** | **89/104 (85.6%)** | **96/119 (80.7%)** | **4.9% [-4.9%;14.7%]** |
| **Country** | | | |
| Georgia | 1/1 (100.0%) | 2/3 (66.7%) | 33.3% [-28.3%;94.9%] |
| India | 1/1 (100.0%) | 3/4 (75.0%) | 25.0% [-22.4%;72.4%] |
| Kazakhstan | 20/24 (83.3%) | 19/23 (82.6%) | 0.7% [-21.0%;22.5%] |
| Lesotho | 13/14 (92.9%) | 12/12 (100.0%) | -7.1% [-20.9%;6.6%] |
| Pakistan | 14/16 (87.5%) | 17/18 (94.4%) | -6.9% [-26.6%;12.7%] |
| Peru | 36/42 (85.7%) | 36/51 (70.6%) | 15.1% [-1.3%;31.6%] |
| South Africa | 4/6 (66.7%) | 7/8 (87.5%) | -20.8% [-66.6%;25.0%] |
| **HIV** | | | |
| Negative | 78/89 (87.6%) | 78/99 (78.8%) | 8.9% [-1.7%;19.4%] |
| Positive | 11/15 (73.3%) | 17/19 (89.5%) | -16.1% [-42.8%;10.5%] |
| **Hepatitis C** | | | |
| Negative | 87/101 (86.1%) | 93/113 (82.3%) | 3.8% [-5.9%;13.6%] |
| Positive | 2/3 (66.7%) | 3/6 (50.0%) | 16.7% [-54.1%;87.4%] |
| **Diabetes** | | | |
| No | 77/88 (87.5%) | 86/104 (82.7%) | 4.8% [-5.2%;14.9%] |
| Yes | 12/16 (75.0%) | 10/15 (66.7%) | 8.3% [-24.1%;40.8%] |
| **BMI** | | | |
| <18.5 | 29/35 (82.9%) | 35/39 (89.7%) | -6.9% [-22.7%;8.9%] |
| 18.5-24.9 | 47/55 (85.5%) | 47/62 (75.8%) | 9.6% [-4.6%;23.9%] |
| ≥25 | 13/14 (92.9%) | 14/18 (77.8%) | 15.1% [-8.8%;38.9%] |
| **Sex** | | | |
| Female | 42/45 (93.3%) | 42/48 (87.5%) | 5.8% [-6.1%;17.8%] |
| Male | 47/59 (79.7%) | 54/71 (76.1%) | 3.6% [-10.7%;17.9%] |
| **Age** | | | |
| <18 | 5/5 (100.0%) | 5/6 (83.3%) | 16.7% [-14.6%;47.9%] |
| 18-44 | 62/70 (88.6%) | 75/91 (82.4%) | 6.2% [-4.7%;17.0%] |
| ≥45 | 22/29 (75.9%) | 16/22 (72.7%) | 3.1% [-21.4%;27.6%] |
| **Pyrazinamide** | | | |
| Sensitive | 36/41 (87.8%) | 44/57 (77.2%) | 10.6% [-4.3%;25.5%] |
| Resistant | 53/63 (84.1%) | 49/59 (83.1%) | 1.1% [-12.1%;14.3%] |
| **Smear results** | | | |
| Negative/scanty | 16/18 (88.9%) | 13/19 (68.4%) | 20.5% [-5.3%;46.3%] |
| 1-2+ | 38/43 (88.4%) | 43/52 (82.7%) | 5.7% [-8.4%;19.8%] |
| 3+ | 35/43 (81.4%) | 40/48 (83.3%) | -1.9% [-17.7%;13.8%] |
| **Cavitation** | | | |
| Absence | 42/46 (91.3%) | 31/43 (72.1%) | 19.2% [3.4%;35.0%] |
| Presence | 47/58 (81.0%) | 64/75 (85.3%) | -4.3% [-17.2%;8.6%] |
| **Prior exposure TB treatment** | | | |
| None | 60/70 (85.7%) | 59/74 (79.7%) | 6.0% [-6.4%;18.3%] |
| First line only | 18/22 (81.8%) | 24/31 (77.4%) | 4.4% [-17.6%;26.4%] |
| Second line | 11/11 (100.0%) | 10/11 (90.9%) | 9.1% [-8.3%;26.5%] |
| **WHO recommendations** | | | |
| <18/12/2018 | 25/32 (78.1%) | 22/31 (71.0%) | 7.2% [-14.5%;28.8%] |
| 18/12/2018-31/03/2020 | 28/33 (84.8%) | 28/35 (80.0%) | 4.8% [-13.3%;23.0%] |
| ≥ 01/04/2020 | 36/39 (92.3%) | 46/53 (86.8%) | 5.5% [-6.9%;18.0%] |

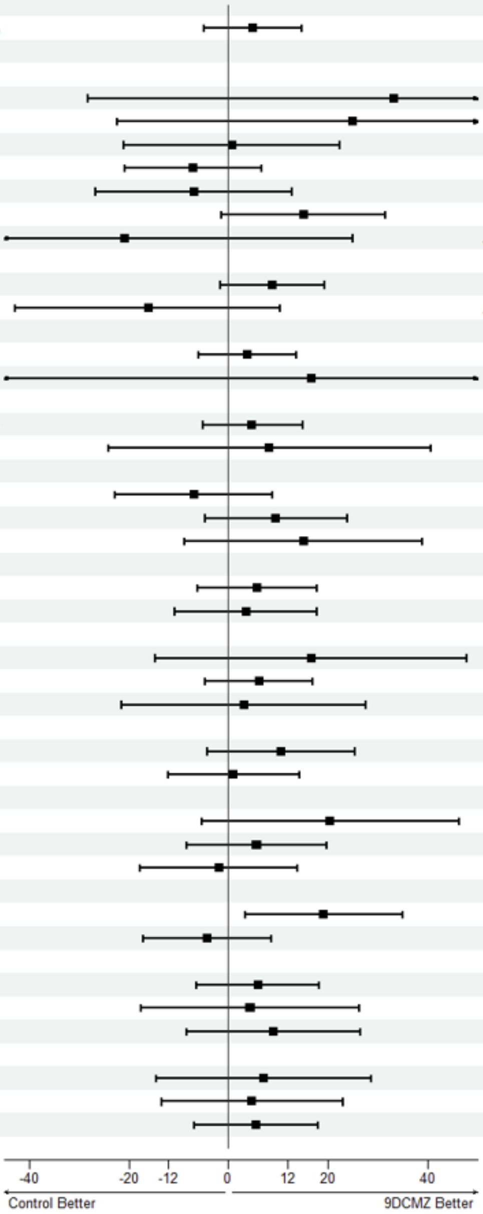

***Figure S5a. Kaplan Meier Curve of Time to Unfavorable Outcome by Week 73: endTB Experimental Regimen 9BLMZ vs. Control (mITT population)***

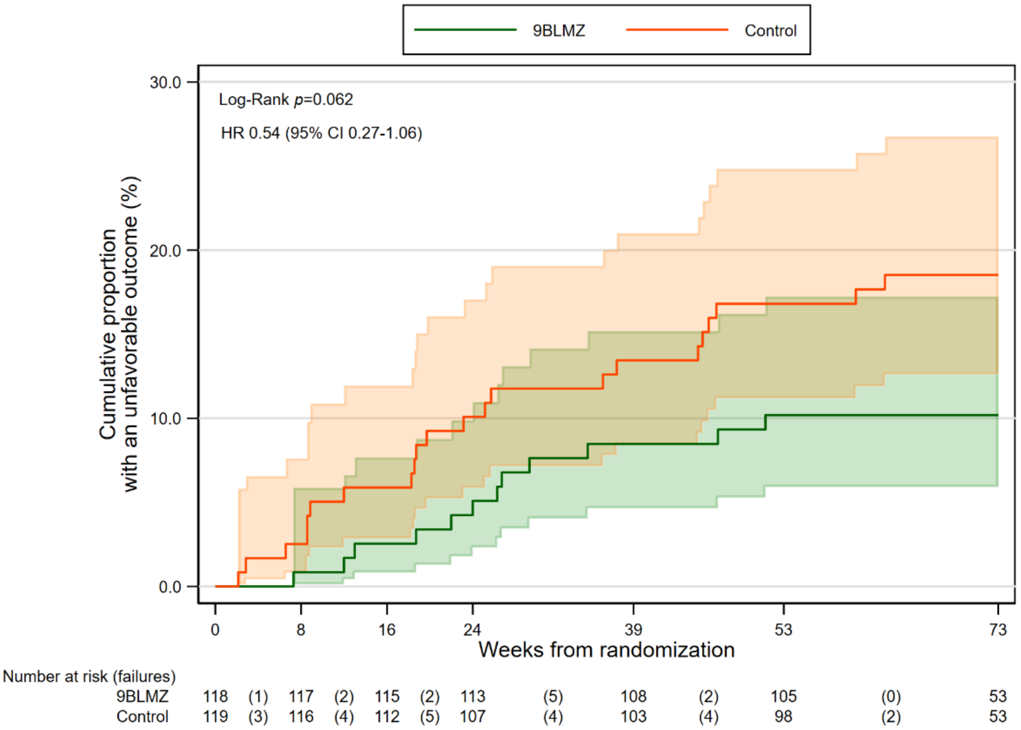

Shaded areas are 95% CI around estimates; Control: orange, Experimental: light green, Control and Experimental: overlapped color

***Figure S5b. Kaplan Meier Curve of Time to Unfavorable Outcome by Week 73: endTB Experimental Regimen 9BCLLfxZ vs. Control (mITT population)***

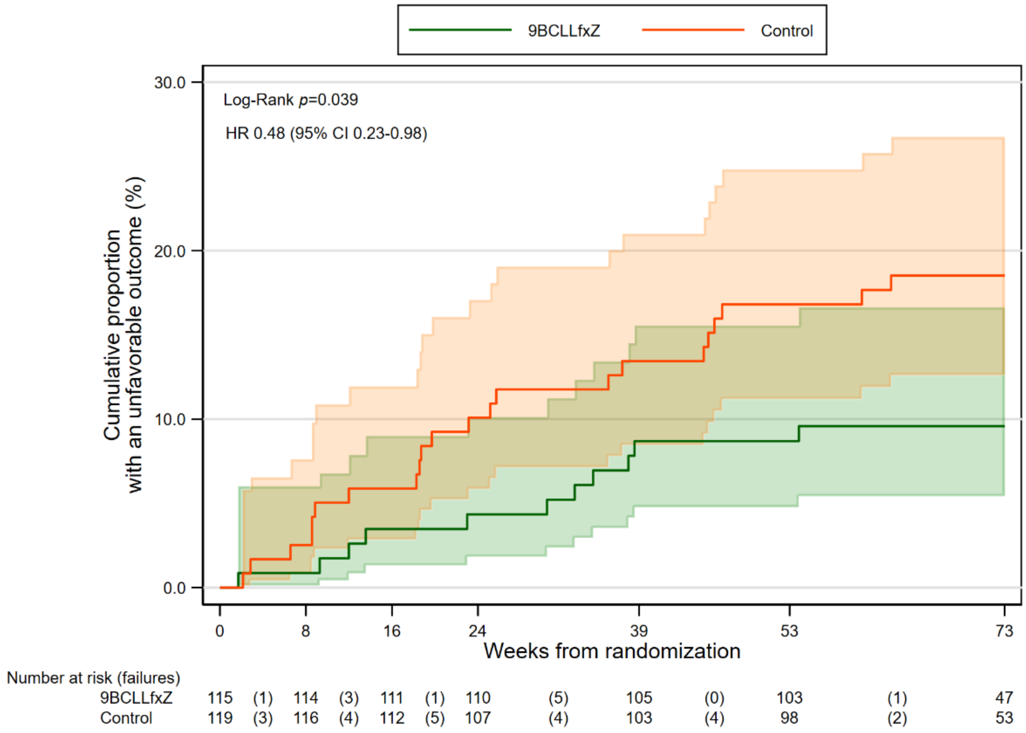

Shaded areas are 95% CI around estimates; Control: orange, Experimental: light green, Control and Experimental: overlapped color

***Figure S5c. Kaplan Meier Curve of Time to Unfavorable Outcome by Week 73: endTB Experimental Regimen 9BDLLfxZ vs. Control (mITT population)***

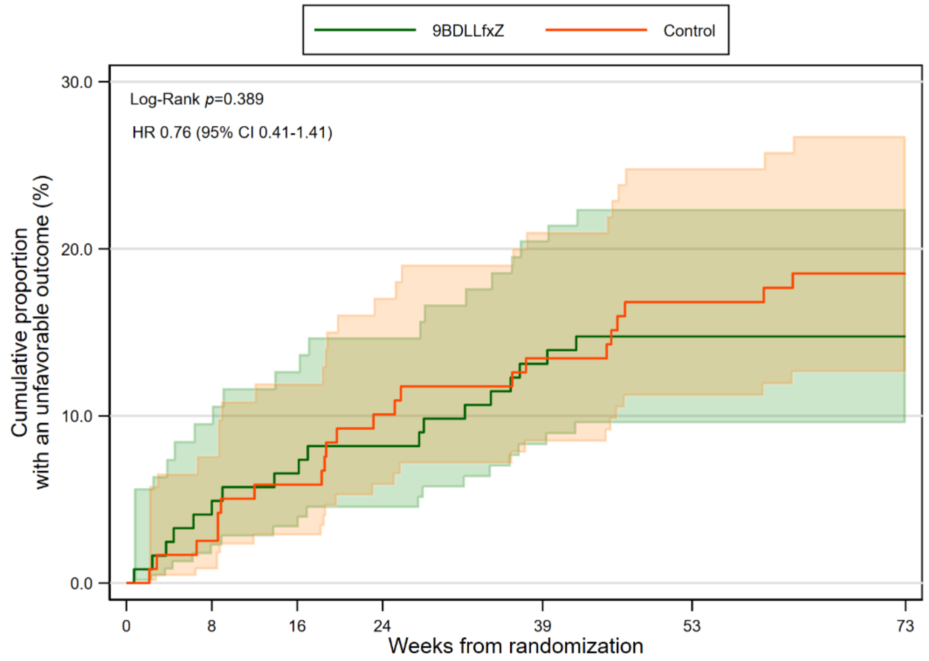

Number at risk (failures)

| | | | | | | | | | | | | | |
|---|---|---|---|---|---|---|---|---|---|---|---|---|---|
| 9BDLLfxZ | 122 | (5) | 117 | (3) | 114 | (2) | 112 | (6) | 106 | (2) | 104 | (0) | 51 |
| Control | 119 | (3) | 116 | (4) | 112 | (5) | 107 | (4) | 103 | (4) | 98 | (2) | 53 |

Shaded areas are 95% CI around estimates; Control: orange, Experimental: light green, Control and Experimental: overlapped color

***Figure S5d. Kaplan Meier Curve of Time to Unfavorable Outcome by Week 73: endTB Experimental Regimen 9DCLLfxZ vs. Control (mITT population)***

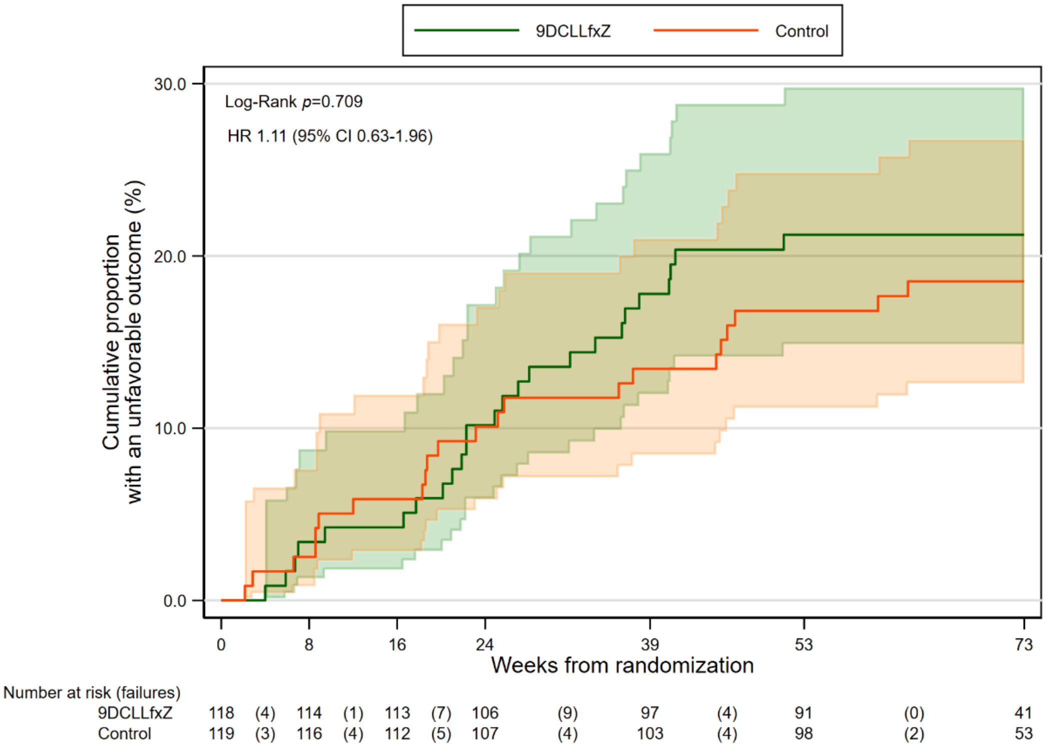

Shaded areas are 95% CI around estimates; Control: orange, Experimental: light green, Control and Experimental: overlapped color

***Figure S5e. Kaplan Meier Curve of Time to Unfavorable Outcome by Week 73: endTB Experimental Regimen 9DCMZ vs. Control (mITT population)***

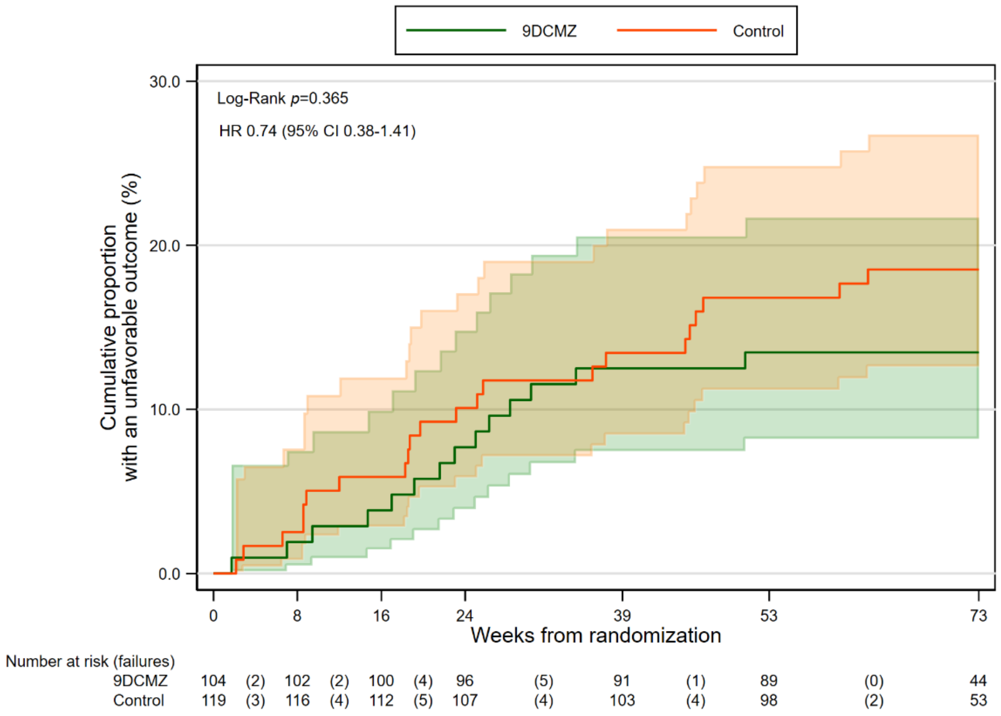

Shaded areas are 95% CI around estimates; Control: orange; Experimental: light green; Control and Experimental: overlapped color

***Table S1. endTB Trial Protocol Writing Committee***

| Name | Role | Institution |
|---|---|---|
| Elisabeth Baudin | Trial Statistician | Epicentre, France |
| Elmira Berikova | Site representative Kazakhstan | National Scientific Center of Phthisiopulmonology, Kazakhstan |
| Maryline Bonnet | TB Epidemiologist | Translational Research on HIV and Endemic and Emerging Infectious Diseases, France |
| Bouke de Jong | Mycobacteriologist | Institute of Tropical Medicine, Belgium |
| Gabriella Ferlazzo | MDR-TB Clinician | Médecins Sans Frontières, France |
| Lorenzo Guglielmetti | MDR-TB Clinician | Médecins Sans Frontières, France |
| Uzma Khan | MDR-TB Clinician | Interactive Research and Development, Singapore |
| Nana Kiria | Site representative Georgia | National Center for Tuberculosis and Lung Diseases, Georgia |
| Nathalie Lachenal | PV expert | Médecins Sans Frontières, Switzerland |
| Leonid Lecca | Site representative Peru | Socios En Salud Sucursal Perú, Peru |
| Helen McIlleron | Pharmacologist | University of Cape Town, South Africa |
| Carole Mitnick | TB Epidemiologist | Harvard Medical School, USA |
| Lawrence Oyewusi | Site representative Lesotho | Partners In Health, Lesotho |
| Samiran Panda | Site representative India | Indian Council of Medical Research-National AIDS Research Center, India |
| Patrick Phillips | TB Statistician | University of California San Francisco, USA |
| Michael Rich | MDR-TB Clinician | Partners In Health, USA |
| Nassem Salahuddin | Site representative Pakistan | The Indus Hospital and Health Network, Pakistan |
| Kwonjune Seung | MDR-TB Clinician | Partners In Health, USA |
| Oleksandr Telnov | MDR-TB Clinician | Hopitaux Universitaires de Genève, Switzerland |
| Lorenzo Trippa | Bayesian Statistician | Dana-Farber Cancer Institute, USA |
| Francis Varaine | MDR-TB Expert | Médecins Sans Frontières, France |
| Gustavo Velásquez | MDR-TB Clinician | University of California San Francisco, USA |
| Sean Wasserman | Site representative South Africa | Institute for Infection and Immunity at St. George’s, United Kingdom |
| Peter Zimetbaum | Electrophysiologist | Beth Israel Deaconess Medical Center, USA |

***Table S2. endTB Implementing Partners by Country***

| Country | Implementing Partners |
|---|---|
| Georgia | National Center for Tuberculosis and Lung Diseases |
| India | Doctors Without Borders India<br>Aundh Chest Hospital |
| Kazakhstan | Partners In Health Kazakhstan<br>National Scientific Center of Phthisiopulmonology<br>National Center for Tuberculosis Problems<br>City Centre of Phthisiopulmonology |
| Lesotho | Partners In Health Lesotho<br>Botsabelo MDR-TB Hospital |
| Pakistan | Interactive Research and Development<br>The Indus Hospital & Health Network<br>Institute of Chest Disease |
| Peru | Socios En Salud (Partners In Health Peru)<br>Hospital Nacional Hipólito Unanue<br>Hospital Nacional Sergio Bernales<br>Hospital Nacional Dos de Mayo |
| South Africa | Médecins Sans Frontières Belgium<br>Town 2 Community Day Centre |

***Table S3. endTB Data Safety and Monitoring Board***

| Name | Institution |
|---|---|
| D. Stephen Coad | Queen Mary University of London, United Kingdom |
| Kelly Dooley | Vanderbilt University, United States |
| Mathilde Fréchet-Jachym | Sanatorium, Centre Hospitalier de Bligny, France |
| Michael Hoelscher | Ludwig Maximilians Universitat, Germany |
| Kevin Monahan | Boston University, United States |

***Table S4. endTB Scientific Advisory Committee***

| Name | Institution |
|---|---|
| Kathleen Eisenach | TB or NOT TB Consulting, United States |
| Robert Horsburgh | Boston University, United States |
| Christoph Lange | Research Center Borstel, Germany |
| Christian Lienhardt | National Institutes of Health, United States |
| Graeme Meintjes | University of Cape Town, South Africa |
| Eric Nuermberger | Johns Hopkins University School of Medicine, United States |
| Andrew Nunn | University College London, United Kingdom |

***Table S5. Global TB Community Advisory Board***

| Name | Institution | Country |
|---|---|---|
| Albert S. Makone | Shiloah Zimbabwe | Zimbabwe |
| Ani Herna Sari | Perhimpunan Organisasi Pasien TB (POP-TB) and Perkumpulan Rekat Surabaya | Indonesia |
| Diptendu Bhattacharya | Survivors Against TB | India |
| Gloria Kerubo Moses | Amref Health Africa | Kenya |
| Jimmy Galarza Castillo | University of Texas Health [formerly Socios En Salud, Peru] | USA |
| Jonathan Stillo | TB Europe Coalition; Eurasian Community for Access to Treatment; Wayne State University | Romania; Moldova; USA |
| Kseniia Schenina | TB People, TB Europe Coalition | Russia |
| Ketholelie Angami | Access to Rights and Knowledge Foundation; Nagaland Users’ Network | India |
| Lindsay McKenna | Treatment Action Group | USA |
| Oxana Rucsineanu | Society of Moldova against Tuberculosis (SMIT); TB Patients Association in Moldova | Moldova |
| Paran Sarmita Winarni | Pejuang Tangguh; Youth Movement Against TB Indonesia | Indonesia |
| Patrick Agbassi | VillageReach | Ivory Coast |
| Rosa Herrera | Americas TB Coalition; Instituto de Servicios de Salud Publica del Estado de Baja California | Mexico |
| Sergiy Kondratyuk | International Treatment Preparedness Coalition | Ukraine |
| Wim Vandevelde | Global Network for and by People Living with HIV; European AIDS Treatment Group; AfroCAB | South Africa; Belgium |

***Table S6. Weight-based dosing and frequency of administration of endTB trial study drugs (experimental arms)***

<table>
<tr><th rowspan="2">Drug*</th><th colspan="6">Weight Band (kg)</th></tr>
<tr><th>24-30</th><th>>30-35</th><th>>35-45</th><th>>45-55</th><th>>55-70</th><th>>70</th></tr>
<tr><td>Bedaquiline (B)</td><td>200 mg QD for 2 weeks** followed by 100 mg 3x/week</td><td colspan="5">400 mg QD x 2 weeks*** followed by 200 mg 3x/week</td></tr>
<tr><td>Delamanid (D)</td><td>50 mg BID</td><td colspan="5">100 mg BID</td></tr>
<tr><td>Moxifloxacin (M)</td><td colspan="6">400 mg</td></tr>
<tr><td>Levofloxacin (Lfx)</td><td colspan="2">750 mg</td><td colspan="4">1000 mg</td></tr>
<tr><td>Linezolid (L)****</td><td>300 mg QD for up to Week 16 (followed by 300 mg QD or 600 mg 3x/week)</td><td colspan="5">600 mg QD for up to Week 16 (followed by 300 mg QD or 600 mg 3x/week)</td></tr>
<tr><td>Clofazimine (C)</td><td colspan="6">100 mg</td></tr>
<tr><td>Pyrazinamide (Z)</td><td colspan="2">800 mg</td><td>1200 mg</td><td colspan="2">1600 mg</td><td>2000 mg</td></tr>
</table>

*Dosing was once a day unless otherwise indicated.

**The intensive phase of bedaquiline treatment for the 24-30 kg weight band comprised 2 weeks of 200 mg QD. If the patient was already receiving bedaquiline treatment, bedaquiline within the experimental regimen should have been dosed as a continuation of that treatment (i.e., only remaining doses of 2-week intensive phase administered and intensive phase was not restarted if it had already been completed at time of study treatment initiation).

*** The intensive phase of bedaquiline treatment for participants weighing more than 30 kg comprised 2 weeks of 400 mg QD. If the patient was already receiving bedaquiline treatment, bedaquiline within the experimental regimen should have been dosed as a continuation of that treatment (i.e., only remaining doses of 2-week intensive phase administered and intensive phase was not restarted if it had already been completed at time of study treatment initiation).

**** Linezolid dosing was routinely modified at Week 16 or sooner if necessary to reduce toxicity related to linezolid. The modification entailed either decreased (300 mg daily) or intermittent (600 mg 3x/week) dosing.

*Table S7. Weight-based dosing and frequency of administration of endTB trial study drugs (control arm)*

<table>
<tr><th rowspan="2">Drug*</th><th colspan="6">Weight Band (kg)</th></tr>
<tr><th>24-30</th><th>>30-35</th><th>>35-45</th><th>>45-55</th><th>>55-70</th><th>>70</th></tr>
<tr><td>Levofloxacin (Lfx)</td><td>500 mg</td><td colspan="2">750 mg</td><td colspan="3">1000 mg</td></tr>
<tr><td>Moxifloxacin (M)</td><td colspan="6">400 mg</td></tr>
<tr><td>Bedaquiline (B)</td><td>200 mg daily for 2 weeks, followed by 100 mg 3 times/week</td><td colspan="5">400 mg daily for 2 weeks, followed by 200 mg 3 times/week</td></tr>
<tr><td>Linezolid (L)</td><td>300 mg</td><td colspan="5">600 mg</td></tr>
<tr><td>Clofazimine (C)</td><td colspan="6">100mg</td></tr>
<tr><td>Cycloserine (Cs)/ Terizidone (Tzd)</td><td colspan="3">500 mg</td><td colspan="3">750 mg</td></tr>
<tr><td>Ethambutol (E)</td><td>400 or 600 mg</td><td colspan="2">800 mg</td><td colspan="3">1200 mg</td></tr>
<tr><td>Delamanid (D)</td><td>50 mg BID</td><td colspan="5">100 mg BID</td></tr>
<tr><td>Pyrazinamide (Z)</td><td>1000 mg</td><td>1200 mg</td><td colspan="3">1600 mg</td><td>2000 mg</td></tr>
<tr><td>Amikacin (Am)</td><td>500 mg</td><td>625 mg</td><td>750 mg</td><td>750 or 1000 mg</td><td colspan="2">1000 mg</td></tr>
<tr><td>Ethionamide (Eto)/ Prothionamide (Pto)</td><td colspan="3">500 mg</td><td colspan="2">750 mg</td><td>1000 mg</td></tr>
<tr><td>Para-aminosalicylic acid (PAS)</td><td>3 to 3,5 g BID</td><td colspan="4">4 g BID</td><td>4 or 6 g BID</td></tr>
<tr><td>Isoniazid (H)</td><td>150 mg</td><td>200 mg</td><td colspan="4">300 mg</td></tr>
<tr><td>High-dose isoniazid ($H^H$)</td><td>400 mg</td><td colspan="2">450 mg</td><td colspan="3">600 mg</td></tr>
</table>

*Dosing was once a day unless otherwise indicated.

#### *Table S8. endTB trial Schedule of Events*

| | | Treatment | | | | | | | | | | | | | | | | | | | | Follow-Up | | | | | | | | | | | | |
|---|---|---|---|---|---|---|---|---|---|---|---|---|---|---|---|---|---|---|---|---|---|---|---|---|---|---|---|---|---|---|---|---|---|---|
| | Screening | Baseline | W1 | W2 | W3 | W4 | W5 | W6 | W7 | W8 | W9 | W 10 | W 11 | W 12 | W 16 | W 20 | W 24 | W 28 | W 32 | W 36 | W39 | W43 | W47 | W53 | W59 | W65 | W73 | W81 | W89 | W97 | W 104 | Sub total (minimal count) | Early Termination | Post-termination follow-up |
| Visit | 1 | 2 | 3 | 4 | 5 | 6 | 7 | 8 | 9 | 10 | 11 | 12 | 13 | 14 | 15 | 16 | 17 | 18 | 19 | 20 | 21 | 22 | 23 | 24 | 25 | 26 | 27 | 28 | 29 | 30 | 31 | | | |
| Window Period | | | +/- 2 | +/- 2 | +/- 2 | +/- 2 | +/- 2 | +/- 2 | +/- 2 | +/- 2[#] | +/- 2 | +/- 2 | +/- 2 | +/- 2 | +/-7 | +/- 7 | +/- 7 | +/- 7 | +/- 7 | +/- 7 | +/- 14 | +/- 14 | +/- 14 | +/- 14 | +/- 14 | +/- 14 | +/- 30 | +/- 14 | +/- 14 | +/- 14 | +/- 30 | | | |
| Eligibility | | | | | | | | | | | | | | | | | | | | | | | | | | | | | | | | | | |
| Subject Consent | X | X | | | | | | | | | | | | | | | | | | | | | | | | | | | | | | 2 | | |
| Demographics | X | X | | | | | | | | | | | | | | | | | | | | | | | | | | | | | | 2 | | |
| Medical History | X | | | | | | | | | | | | | | | | | | | | | | | | | | | | | | | 1 | | |
| Inclusion/ Exclusion | X | X | | | | | | | | | | | | | | | | | | | | | | | | | | | | | | 2 | | |
| Clinical Evaluation | | | | | | | | | | | | | | | | | | | | | | | | | | | | | | | | | | |
| Vital Signs | X | X | X | X | X | X | X | X | X | X | X | X | X | X | X | X | X | X | X | X | X | X | X | X | X | X | X | X | X | X | X | 31 | X | X |
| Interval Medical History | | X | X | X | X | X | X | X | X | X | X | X | X | X | X | X | X | X | X | X | X | X | X | X | X | X | X | X | X | X | X | 30 | X | X |
| Physical Exam | X | X | X | X | X | X | X | X | X | X | X | X | X | X | X | X | X | X | X | X | X | X | X | X | X | X | X | X | X | X | X | 31 | X | X |
| TB Symptom Assessment | | X | X | X | X | X | X | X | X | X | X | X | X | X | X | X | X | X | X | X | X | X | X | X | X | X | X | X | X | X | X | 30 | X | X |
| ECOG Assessment | | X | | | | | | | | | | | | | | | | | | | X | | | | | | X | | | | X | 4 | X | X |
| Neurologic Exam | | X | | | | X | | | | X | | | | X | X | X | X | X | X | X | X | X | X | | | | X | | | | X | 15 | X | |
| Ophthalmologic Exam | | X | | | | X | | | | X | | | | X | X | X | X | X | X | X | X | X | X | | | | X | | | | X | 15 | X | |
| Mental Health Assessment | | X | | | | | | | | | | | | | | | | | | | | | | | | | X | | | | X | 3 | X | |
| Treatment | | | | | | | | | | | | | | | | | | | | | | | | | | | | | | | | | | |
| Randomization | | X* | | | | | | | | | | | | | X** | | | | | | | | | | | | | | | | | (1) | | |
| Treatment Compliance | | | X | X | X | X | X | X | X | X | X | X | X | X | X | X | X | X | X | X | X | X[C] | X[C] | X[C] | X[C] | X[C] | X[C] | X[C] | X[C] | X[C] | X[C] | 19 | X[C] | |
| Adherence Counseling | | X | X | X | X | X | X | X | X | X | X | X | X | X | X | X | X | X | X | X | X | X[C] | X[C] | X[C] | X[C] | X[C] | X[C] | X[C] | X[C] | X[C] | | 20 | | |
| Concomitant Medication Review | X | X | X | X | X | X | X | X | X | X | X | X | X | X | X | X | X | X | X | X | X | X | X | X | X | X | X | X | X | X | X | 31 | X | X |
| AE Review | | | X | X | X | X | X | X | X | X | X | X | X | X | X | X | X | X | X | X | X | X | X | X | X | X | X | X | X | X | X | 29 | X | X |
| Laboratory Testing | | | | | | | | | | | | | | | | | | | | | | | | | | | | | | | | | | |
| CBC | X[1] | | | | | X | | | | X | | | | X | X | X | X | X | X | X | X | X | X | | | | | | | | X | 14 | X | |
| Total Ca++, K+, Mg++ | X[1] | | | | | X | | | | X | | | | X | X | X | X | X | X | X | X | X | X | | | | | | | | | 13 | X | |

| | Screening | Baseline | W1 | W2 | W3 | W4 | W5 | W6 | W7 | W8 | W 9 | W 10 | W 11 | W 12 | W 16 | W 20 | W 24 | W 28 | W 32 | W 36 | W39 | W 43 | W 47 | W 53 | W 59 | W 65 | W 73 | W 81 | W 89 | W 97 | W 104 | Sub total (minimal count) | Early Termination | Post-termination follow-up |
|---|---|---|---|---|---|---|---|---|---|---|---|---|---|---|---|---|---|---|---|---|---|---|---|---|---|---|---|---|---|---|---|---|---|---|
| Visit | 1 | 2 | 3 | 4 | 5 | 6 | 7 | 8 | 9 | 10 | 11 | 12 | 13 | 14 | 15 | 16 | 17 | 18 | 19 | 20 | 21 | 22 | 23 | 24 | 25 | 26 | 27 | 28 | 29 | 30 | 31 | | | |
| Window Period | | | +/- 2 | +/- 2 | +/- 2 | +/- 2 | +/- 2 | +/- 2 | +/- 2 | +/- 2# | +/- 2 | +/- 2 | +/- 2 | +/- 2 | +/- 7 | +/- 7 | +/- 7 | +/- 7 | +/- 7 | +/- 7 | +/- 14 | +/- 14 | +/- 14 | +/- 14 | +/- 14 | +/- 14 | +/- 30 | +/- 14 | +/- 14 | +/- 14 | +/- 30 | | | |
| Creatinine | X[1] | | | | | X | | | | X | | | | X | X | X | X | X | X | X | X | X | X | | | | | | | | | 13 | X | |
| AST and ALT[2] | X[1] | | | | | X | | | | X | | | | X | X | X | X | X | X | X | X | X | X | | | | | | | | | 13 | X | |
| Total and Direct Bilirubin | X[1] | | | | | X | | | | X | | | | X | X | X | X | X | X | X | X | X | X | | | | | | | | | 13 | X | |
| Albumin | X[1] | | | | | | | | | | | | | | | | | | | | | | | | | | | | | | | 1 | | |
| TSH | X[3] | X[8] | | | | | | | | | | | | | | | X | | | | | | X | | | | | | | | | 3 | X | |
| HbA1c | X[4] | X[8] | | | | | | | | | | | | | | | X[5] | | | | | | X[5] | | | | X[5] | | | | | (1) | X[5, 17] | X[5, 17] |
| HIV test | X[6] | | | | | | | | | | | | | | | | | | | | | | | | | | | | | | | 1 | | |
| CD4 count and viral load (in HIV+) | X[7] | X[8] | | | | | | | | | | | | | | | X[9] | | | | | | X[9] | | | | X[9] | | | | | (0) | X[9,17] | X[9,17] |
| Hepatitis B serology (anti-HBc total and HbsAg) | X | X[8] | | | | | | | | | | | | | | | | | | | | | | | | | | | | | | 1 | | |
| Hepatitis C Serology: HCVAb | X | X[8] | | | | | | | | | | | | | | | | | | | | | | | | | | | | | | 1 | | |
| Pregnancy test[11] | X | X[10] | | | | | | | | | | | | | | | | | | | | | | | | | | | | | | (2) | X | |
| Sputum Specimen Testing | | | | | | | | | | | | | | | | | | | | | | | | | | | | | | | | | | |
| Smear Microscopy | X | | | X | | X | | | | X | | | | X | X | X | X | X | X | X | X | X | X | X | X | X | X | X | X | X | X | 22 | X | X |
| Culture (LJ & MGIT) | X | | | X | | X | | | | X | | | | X | X | X | X | X | X | X | X | X | X | X | X | X | X | X | X | X | X | 22 | X | X |
| Rapid Molecular Test for RIF Resistance | X[12] | X[8] | | | | | | | | | | | | | | | | | | | | | | | | | | | | | | 1 | | |
| Rapid molecular test for FQ resistance | X[12] | X[8] | | | | | | | | | | | | | X[13] | | | | | | | | | | | | | | | | | (1) | | |
| DST (conventional 1st and 2nd line) | X | | | | | | | | | | | | | | X[13,14] | | | | | | | | | | | | | | | | | (1) | X[17] | X[17] |
| DST (new drugs) | X[14] | | | | | | | | | | | | | | X[14] | | | | | | | | | | | | | | | | | (0) | X[17] | X[17] |
| Genotyping[14] | X | | | | | | | | | | | | | | X | | | | | | | | | | | | | | | | | (0) | X[17] | X[17] |
| Specimen Storage | X[15] | | | | | | | | | | | | | | X[15] | | | | | | | | | | | | | | | | | (1) | X | |
| Special Assays or Procedures | | | | | | | | | | | | | | | | | | | | | | | | | | | | | | | | | | |
| Audiometry | | X | | | | X | | | | X | | | | X | X | X | X | X | X | X | X | X | X | | | | X | | | | X | 15 | X | |
| ECG[1] | X[1] | X | X | X | X | X | X | X | X | X | X | X | X | X | X | X | X | X | X | X | X | X | X | X | | | X | | | | | 25 | X | |
| Chest X-ray | | X[16] | | | | | | | | X | | | | | | | | | | | X | | | | | | X | | | | X | 5 | X[17] | X[17] |

***Legend: endTB Trial Schedule of Events***

#: W8 visit window was to occur within 8 weeks (+/-2 days) of study treatment initiation.

*: Randomization for treatment regimen assignment.

**: Only for participants receiving linezolid, randomization to linezolid dose reduction strategy could occur at W16 or earlier (concomitant to dose reduction decision).

C: Only for participants still on prescription.

1. Test with abnormal result at screening visit could be repeated within screening window.
2. ALT/AST tests were to be symptom-driven after Week 47.
3. Optionally, documented results of TSH (performed less than 4 weeks prior to screening visit date) could substitute for screening/baseline test. Investigators were encouraged to repeat TSH if they deemed it useful for patient management or to have a reliable pre-treatment result.
4. Optionally, documented results of HbA1c (performed less than 3 months prior to screening visit date) could substitute for screening/baseline test. Investigators were encouraged to repeat HbA1c if they deemed it useful for patient management or to have a reliable pre-treatment result.
5. HbA1c test (or 2 fasting blood glucose tests if HbA1c could not be done) to be repeated every 6 months only if abnormal at screening/baseline.
6. HIV serology was to be offered, unless patient was known to be positive or a documented negative result was available from less than one month before screening visit date.
7. CD4 count and HIV viral load were performed if the patient was known or found to be HIV positive. Optionally, documented results of CD4 count (performed less than six months prior to screening visit date) and viral load (performed less than 4 weeks prior to screening visit date) could substitute for screening test. Investigators were encouraged to repeat CD4 count/viral load if they deemed it useful for patient management or to have a reliable pre-treatment result.
8. Completed only if not done at screening.
9. CD4 count and HIV viral load were monitored every 6 months in HIV-positive patients.
10. Serum pregnancy test at baseline visit could be repeated if the blood specimen had not been drawn within 72 hours prior to treatment start.
11. Pregnancy tests could be performed on urine or serum samples after baseline visit until the end of study treatment.
12. Rapid molecular test results for RIF and FQ resistance, performed at designated study lab not more than 3 weeks prior to screening visit date had to be available before randomization.
13. Done locally on first positive culture or any culture reversion at or after Week 16.
14. Done at ITM on first positive culture or any culture reversion at or after Week 16 and on corresponding screening/baseline strain.
15. Stored the screening/baseline sample and isolate from the corresponding culture. When possible, stored samples and isolates from subsequent positive cultures for study (and future research) purposes.
16. Was not necessary if previous adequate imaging results within 3 weeks prior to baseline visit date were available.
17. Could be indicated, based on availability of previous results.

*Table S9. Other Baseline Characteristics of mITT Population*

| Baseline characteristics | 9BLMZ (n = 118) | 9BCLLfxZ (n = 115) | 9BDLLfxZ (n = 122) | 9DCLLfxZ (n = 118) | 9DCMZ (n = 104) | Control (n = 119) | Total (N = 696) |
|---|---|---|---|---|---|---|---|
| **Baseline QTcF* (ms)** | | | | | | | |
| Median [IQR] | 400.5 [384.0;415.0] | 399.5 [387.5;416.5] | 398.0 [382.5;420.0] | 401.3 [380.0;417.0] | 399.3 [383.8;414.5] | 397.5 [376.5;417.0] | 399.5 [383.0;417.5] |
| Min-Max | 340.0 - 449.0 | 340.0 - 448.0 | 344.0 - 448.0 | 335.5 - 448.0 | 340.0 - 444.5 | 348.0 - 443.0 | 335.5 - 449.0 |
| **Sensory neuropathy** | | | | | | | |
| Normal | 88 (74.6%) | 77 (67.0%) | 101 (82.8%) | 97 (82.2%) | 74 (71.2%) | 93 (78.2%) | 530 (76.2%) |
| Grade 1 | 17 (14.4%) | 21 (18.3%) | 14 (11.5%) | 6 (5.1%) | 9 (8.7%) | 15 (12.6%) | 82 (11.8%) |
| Grade 2 | 6 (5.1%) | 15 (13.0%) | 3 (2.5%) | 12 (10.2%) | 15 (14.4%) | 4 (3.4%) | 55 (7.9%) |
| Grade 3 | 7 (5.9%) | 2 (1.7%) | 4 (3.3%) | 3 (2.5%) | 6 (5.8%) | 7 (5.9%) | 29 (4.2%) |
| **Visual acuity** | | | | | | | |
| Grade 1 | 23 (19.5%) | 28 (24.4%) | 42 (34.4%) | 24 (20.3%) | 23 (22.1%) | 32 (27.1%) | 172 (24.8%) |
| Grade 2 | 2 (1.7%) | 7 (6.1%) | 6 (4.9%) | 4 (3.4%) | 3 (2.9%) | 6 (5.1%) | 28 (4.0%) |
| Grade 3 | 22 (18.6%) | 26 (22.6%) | 18 (14.8%) | 19 (16.1%) | 18 (17.3%) | 17 (14.4%) | 120 (17.3%) |
| Grade 4 | 1 (0.9%) | 3 (2.6%) | 2 (1.6%) | 3 (2.5%) | 4 (3.9%) | 1 (0.9%) | 14 (2.0%) |
| **Complete blood cell count** | | | | | | | |
| **WBC count ($10^9$/L)** | | | | | | | |
| Median [IQR] | 8.2 [6.5;10.9] | 8.7 [7.2;11.3] | 8.6 [6.7;11.0] | 8.3 [6.4;10.0] | 8.2 [6.2;10.3] | 8.6 [7.0;10.6] | 8.5 [6.7;10.8] |
| Min-Max | 3.1 - 23.5 | 2.9 - 18.5 | 3.0 - 17.8 | 3.5 - 17.3 | 2.5 - 19.9 | 2.1 - 16.6 | 2.1 - 23.5 |
| **Neutrophils ($10^9$/L)** | | | | | | | |
| Median [IQR] | 5.8 [4.5;7.7] | 6.2 [5.0;8.5] | 6.1 [4.4;8.0] | 5.5 [4.0;7.2] | 5.6 [4.1;7.5] | 6.0 [4.6;7.5] | 5.8 [4.5;7.9] |
| Min-Max | 1.8 - 20.7 | 1.1 - 15.9 | 1.2 - 15.6 | 1.8 - 14.5 | 0.9 - 16.5 | 0.9 - 14.1 | 0.9 - 20.7 |
| **Lymphocytes ($10^9$/L)** | | | | | | | |
| Median [IQR] | 1.6 [1.3;2.1] | 1.6 [1.3;2.1] | 1.7 [1.3;2.1] | 1.6 [1.3;2.2] | 1.6 [1.3;2.1] | 1.6 [1.2;2.1] | 1.6 [1.3;2.1] |
| Min-Max | 0.3 - 12.0 | 0.4 - 4.7 | 0.5 - 3.9 | 0.3 - 3.9 | 0.3 - 5.4 | 0.5 - 6.2 | 0.3 - 12.0 |
| **Eosinophils (%)** | | | | | | | |
| Median [IQR] | 0.2 [0.1;0.2] | 0.2 [0.1;0.3] | 0.2 [0.1;0.3] | 0.2 [0.1;0.3] | 0.1 [0.1;0.2] | 0.1 [0.1;0.3] | 0.2 [0.1;0.3] |
| Min-Max | 0.0 - 3.0 | 0.0 - 1.5 | 0.0 - 4.0 | 0.0 - 0.9 | 0.0 - 1.3 | 0.0 - 2.8 | 0.0 - 4.0 |
| **Hemoglobin (g/dL)** | | | | | | | |
| Median [IQR] | 12.2 [10.6;13.9] | 12.7 [11.0;14.1] | 12.3 [11.0;13.4] | 12.6 [11.2;14.3] | 12.5 [11.0;13.6] | 12.4 [10.7;13.7] | 12.5 [10.9;13.8] |
| Min-Max | 8.1 - 16.2 | 8.2 - 17.1 | 8.1 - 17.9 | 8.3 - 17.1 | 8.6 - 16.3 | 8.1 - 16.6 | 8.1 - 17.9 |
| **Hematocrit (L/L)** | | | | | | | |
| Median [IQR] | 37.6 [33.8;41.7] | 37.4 [34.6;41.1] | 37.0 [34.0;40.8] | 38.1 [34.6;42.0] | 37.8 [34.0;40.5] | 37.8 [33.8;41.2] | 37.7 [34.0;41.0] |
| Min-Max | 25.1 - 49.5 | 26.0 - 48.0 | 25.0 - 56.4 | 25.7 - 50.8 | 26.7 - 53.1 | 23.7 - 50.0 | 23.7 - 56.4 |
| **Platelet count ($10^9$/L)** | | | | | | | |

| Baseline characteristics | 9BLMZ (n = 118) | 9BCLLfxZ (n = 115) | 9BDLLfxZ (n = 122) | 9DCLLfxZ (n = 118) | 9DCMZ (n = 104) | Control (n = 119) | Total (N = 696) |
|---|---|---|---|---|---|---|---|
| Median [IQR] | 409.0 [295.0;538.0] | 387.0 [299.0;474.0] | 406.0 [304.0;480.0] | 359.0 [279.0;443.0] | 373.5 [282.0;459.0] | 369.0 [299.0;483.0] | 382.5 [294.0;479.0] |
| Min-Max | 146.0 - 854.0 | 80.0 - 810.0 | 121.0 - 729.0 | 140.0 - 747.0 | 131.0 - 1166.0 | 30.0 - 856.0 | 30.0 - 1166.0 |
| **RBC ($10^{12}$/L)** | | | | | | | |
| Median [IQR] | 4.6 [4.1;4.9] | 4.5 [4.2;4.8] | 4.5 [4.1;4.9] | 4.5 [4.1;4.9] | 4.4 [4.1;4.8] | 4.5 [4.1;4.9] | 4.5 [4.1;4.9] |
| Min-Max | 2.8 - 5.8 | 2.8 - 6.0 | 3.0 - 6.4 | 2.8 - 6.0 | 2.8 - 6.5 | 2.3 - 5.9 | 2.3 - 6.5 |
| **Biochemistry** | | | | | | | |
| **ALT (U/L)** | | | | | | | |
| Median [IQR] | 19.0 [12.0;36.9] | 18.0 [11.0;27.0] | 18.0 [12.0; 27.0] | 22.0 [13.0;29.0] | 16.0 [11.0;25.0] | 18.0 [11.0;28.0] | 19.0 [11.9;29.0] |
| Min-Max | 2.4-81.0 | 2.9-115.0 | 2.6-103.0 | 3.5-103.0 | 1.5-99.0 | 5.0-123.0 | 1.5-123.0 |
| **AST (U/L)** | | | | | | | |
| Median [IQR] | 22.0 [15.3;31.0] | 20.0 [15.2;28.9] | 22.0 [15.0;29.2] | 21.9 [17.0;30.6] | 21.4 [14.0;29.0] | 21.0 [15.0;30.0] | 21.0 [15.0;29.7] |
| Min-Max | 5.0-96.0 | 4.0-85.0 | 3.0-96.4 | 2.0-125.0 | 2.0-109.0 | 2.0-79.3 | 2.0-125.0 |
| **Bilirubin (mg/dL)** | | | | | | | |
| Median [IQR] | 0.40 [0.23;0.53] | 0.40 [0.29;0.60] | 0.30 [0.20;0.48] | 0.38 [0.23;0.60] | 0.38 [0.29;0.60] | 0.38 [0.29;0.60] | 0.38 [0.24;0.60] |
| Min-Max | 0.14-1.40 | 0.00-1.60 | 0.08-1.18 | 0.15-2.09 | 0.13-1.40 | 0.11-1.10 | 0.0-2.09 |

*median of the highest QT intervals corrected according to the Fridericia formula

***Table S10. endTB Trial Control Regimen composition and duration at initiation of treatment, by country (mITT Population)***

| | Georgia | India | Kazakhstan | Lesotho | Peru | Pakistan | South Africa | Total |
|---|---|---|---|---|---|---|---|---|
| N (%) | 3 (2.5%) | 4 (3.4%) | 23 (19.3%) | 12 (10.1%) | 51 (42.9%) | 18 (15.1%) | 8 (6.7%) | 119 (100.0%) |
| **Number of drugs in regimen** | | | | | | | | |
| 4 | 0 (0.0%) | 0 (0.0%) | 0 (0.0%) | 0 (0.0%) | 0 (0.0%) | 1 (5.6%) | 0 (0.0%) | 1 (0.8%) |
| 5 | 2 (66.7%) | 1 (25.0%) | 16 (69.6%) | 8 (66.7%) | 39 (76.5%) | 16 (88.9%) | 5 (62.5%) | 87 (73.1%) |
| 6 | 1 (33.3%) | 0 (0.0%) | 5 (21.7%) | 4 (33.3%) | 12 (23.5%) | 0 (0.0%) | 2 (25.0%) | 24 (20.2%) |
| 7 | 0 (0.0%) | 3 (75.0%) | 2 (8.7%) | 0 (0.0%) | 0 (0.0%) | 1 (5.6%) | 1 (12.5%) | 7 (5.9%) |
| **Drugs** | | | | | | | | |
| *WHO Group A* | | | | | | | | |
| Bedaquiline (B) | 0 (0.0%) | 4 (100.0%) | 18 (78.3%) | 9 (75.0%) | 39 (76.5%) | 18 (100.0%) | 8 (100.0%) | 96 (80.7%) |
| Levofloxacin (Lfx) | 2 (66.7%) | 4 (100.0%) | 22 (95.7%) | 12 (100.0%) | 51 (100.0%) | 14 (77.8%) | 8 (100.0%) | 113 (95.0%) |
| Moxifloxacin (M) | 1 (33.3%) | 0 (0.0%) | 1 (4.3%) | 0 (0.0%) | 0 (0.0%) | 4 (22.2%) | 0 (0.0%) | 6 (5.0%) |
| Linezolid (L) | 0 (0.0%) | 1 (25.0%) | 18 (78.3%) | 7 (58.3%) | 39 (76.5%) | 17 (94.4%) | 4 (50.0%) | 86 (72.3%) |
| *WHO Group B* | | | | | | | | |
| Clofazimine (Cfz) | 0 (0.0%) | 4 (100.0%) | 18 (78.3%) | 10 (83.3%) | 36 (70.6%) | 18 (100.0%) | 8 (100.0%) | 94 (79.0%) |
| Cycloserine/Terizidone (Cs-Tzd) | 3 (100.0%) | 1 (25.0%) | 21 (91.3%) | 7 (58.3%) | 35 (68.6%) | 14 (77.8%) | 4 (50.0%) | 85 (71.4%) |
| *WHO Group C* | | | | | | | | |
| Ethambutol (E) | 0 (0.0%) | 3 (75.0%) | 4 (17.4%) | 0 (0.0%) | 12 (23.5%) | 1 (5.6%) | 1 (12.5%) | 21 (17.6%) |
| Isoniazid (H) | 0 (0.0%) | 3 (75.0%) | 0 (0.0%) | 0 (0.0%) | 0 (0.0%) | 1 (5.6%) | 2 (25.0%) | 6 (5.0%) |
| Pyrazinamide (Z) | 2 (66.7%) | 3 (75.0%) | 8 (34.8%) | 4 (33.3%) | 31 (60.8%) | 1 (5.6%) | 6 (75.0%) | 55 (46.2%) |
| Delamanid (D) | 2 (66.7%) | 0 (0.0%) | 1 (4.3%) | 7 (58.3%) | 0 (0.0%) | 2 (11.1%) | 0 (0.0%) | 12 (10.1%) |
| Ethionamide/Prothionamide (Eto-Pto) | 2 (66.7%) | 3 (75.0%) | 5 (21.7%) | 4 (33.3%) | 12 (23.5%) | 1 (5.6%) | 3 (37.5%) | 30 (25.2%) |
| Para-aminosalicylic acid (PAS) | 1 (33.3%) | 0 (0.0%) | 3 (13.0%) | 2 (16.7%) | 0 (0.0%) | 0 (0.0%) | 0 (0.0%) | 6 (5.0%) |
| Capreomycin (CM) | 3 (100.0%) | 0 (0.0%) | 5 (21.7%) | 0 (0.0%) | 5 (9.8%) | 0 (0.0%) | 0 (0.0%) | 13 (10.9%) |
| Kanamycin (KM) | 0 (0.0%) | 0 (0.0%) | 0 (0.0%) | 2 (16.7%) | 7 (13.7%) | 0 (0.0%) | 0 (0.0%) | 9 (7.6%) |
| **Regimen** | | | | | | | | |
| B/Cfz/E/Eto-Pto/H/Lfx/Z | 0 (0.0%) | 3 (75.0%) | 0 (0.0%) | 0 (0.0%) | 0 (0.0%) | 1 (5.6%) | 1 (12.5%) | 5 (4.2%) |
| B/Cfz/Cs-Tzd/L/M | 0 (0.0%) | 0 (0.0%) | 0 (0.0%) | 0 (0.0%) | 0 (0.0%) | 2 (11.1%) | 0 (0.0%) | 2 (1.7%) |
| B/Cfz/Cs-Tzd/L/Lfx | 0 (0.0%) | 1 (25.0%) | 14 (60.9%) | 3 (25.0%) | 20 (39.2%) | 12 (66.7%) | 2 (25.0%) | 52 (43.7%) |
| B/Cfz/Cs-Tzd/D/Lfx | 0 (0.0%) | 0 (0.0%) | 0 (0.0%) | 2 (16.7%) | 0 (0.0%) | 0 (0.0%) | 0 (0.0%) | 2 (1.7%) |
| B/Cfz/D/L/Lfx | 0 (0.0%) | 0 (0.0%) | 1 (4.4%) | 3 (25.0%) | 0 (0.0%) | 0 (0.0%) | 0 (0.0%) | 4 (3.4%) |
| B/Cfz/D/L/M | 0 (0.0%) | 0 (0.0%) | 0 (0.0%) | 0 (0.0%) | 0 (0.0%) | 2 (11.1%) | 0 (0.0%) | 2 (1.7%) |
| B/Cfz/Eto-Pto/Lfx/Z | 0 (0.0%) | 0 (0.0%) | 0 (0.0%) | 0 (0.0%) | 0 (0.0%) | 0 (0.0%) | 2 (25.0%) | 2 (1.7%) |
| B/Cfz/H/Lfx/Z | 0 (0.0%) | 0 (0.0%) | 0 (0.0%) | 0 (0.0%) | 0 (0.0%) | 0 (0.0%) | 1 (12.5%) | 1 (0.8%) |
| B/Cfz/L/Lfx/Z | 0 (0.0%) | 0 (0.0%) | 1 (4.4%) | 0 (0.0%) | 16 (31.4%) | 0 (0.0%) | 0 (0.0%) | 17 (14.3%) |
| B/Cfz/L/Lfx | 0 (0.0%) | 0 (0.0%) | 0 (0.0%) | 0 (0.0%) | 0 (0.0%) | 1 (5.6%) | 0 (0.0%) | 1 (0.8%) |
| B/Cfz/Cs-Tzd/L/M/Z | 0 (0.0%) | 0 (0.0%) | 1 (4.4%) | 0 (0.0%) | 0 (0.0%) | 0 (0.0%) | 0 (0.0%) | 1 (0.8%) |
| B/Cfz/Cs-Tzd/L/Lfx/Z | 0 (0.0%) | 0 (0.0%) | 1 (4.4%) | 0 (0.0%) | 0 (0.0%) | 0 (0.0%) | 2 (25.0%) | 3 (2.5%) |

| | Georgia | India | Kazakhstan | Lesotho | Peru | Pakistan | South Africa | Total |
|---|---|---|---|---|---|---|---|---|
| B/Cfz/D/Eto-Pto/Lfx/Z | 0 (0.0%) | 0 (0.0%) | 0 (0.0%) | 1 (8.3%) | 0 (0.0%) | 0 (0.0%) | 0 (0.0%) | 1 (0.8%) |
| B/Cs-Tzd/L/Lfx/Z | 0 (0.0%) | 0 (0.0%) | 0 (0.0%) | 0 (0.0%) | 3 (5.9%) | 0 (0.0%) | 0 (0.0%) | 3 (2.5%) |
| CM/Cs-Tzd/D/Lfx/PAS | 1 (33.3%) | 0 (0.0%) | 0 (0.0%) | 0 (0.0%) | 0 (0.0%) | 0 (0.0%) | 0 (0.0%) | 1 (0.8%) |
| CM/Cs-Tzd/Eto-Pto/M/Z | 1 (33.3%) | 0 (0.0%) | 0 (0.0%) | 0 (0.0%) | 0 (0.0%) | 0 (0.0%) | 0 (0.0%) | 1 (0.8%) |
| CM/Cs-Tzd/E/Eto-Pto/Lfx/PAS /Z | 0 (0.0%) | 0 (0.0%) | 2 (8.7%) | 0 (0.0%) | 0 (0.0%) | 0 (0.0%) | 0 (0.0%) | 2 (1.7%) |
| CM/Cs-Tzd/D/Eto-Pto/Lfx/Z | 1 (33.3%) | 0 (0.0%) | 0 (0.0%) | 0 (0.0%) | 0 (0.0%) | 0 (0.0%) | 0 (0.0%) | 1 (0.8%) |
| CM/Cs-Tzd/E/Eto-Pto/Lfx/Z | 0 (0.0%) | 0 (0.0%) | 2 (8.7%) | 0 (0.0%) | 5 (9.8%) | 0 (0.0%) | 0 (0.0%) | 7 (5.9%) |
| CM/Cs-Tzd/Eto-Pto/Lfx/PAS/Z | 0 (0.0%) | 0 (0.0%) | 1 (4.4%) | 0 (0.0%) | 0 (0.0%) | 0 (0.0%) | 0 (0.0%) | 1 (0.8%) |
| Cs-Tzd/Eto-Pto/KM/Lfx/PAS /Z | 0 (0.0%) | 0 (0.0%) | 0 (0.0%) | 2 (16.7%) | 0 (0.0%) | 0 (0.0%) | 0 (0.0%) | 2 (1.7%) |
| Cs-Tzd/D/Eto-Pto/L/Lfx/Z | 0 (0.0%) | 0 (0.0%) | 0 (0.0%) | 1 (8.3%) | 0 (0.0%) | 0 (0.0%) | 0 (0.0%) | 1 (0.8%) |
| Cs-Tzd/E/Eto-Pto/KM/Lfx /Z | 0 (0.0%) | 0 (0.0%) | 0 (0.0%) | 0 (0.0%) | 7 (13.7%) | 0 (0.0%) | 0 (0.0%) | 7 (5.9%) |
| **Other regimen descriptors** | | | | | | | | |
| Shorter 9-12 month regimen^ | 0 (0.0%) | 3 (75.0%) | 0 (0.0%) | 0 (0.0%) | 0 (0.0%) | 1 (5.6%) | 1 (12.5%) | 5 (4.2%) |
| Treatment duration (weeks) * | | | | | | | | |
| N | 2 | 1 | 18 | 10 | 34 | 17 | 6 | 88 |
| Median [IQR] | 89.9 [87.0;92.7] | 62.7 [62.7;62.7] | 78.3 [78.1;79.0] | 80.8 [80.1;81.0] | 78.0 [78.0;79.7] | 78.0 [78.0;78.3] | 72.0 [71.9;74.9] | 78.1 [78.0;80.0] |
| Min-Max | 87.0 - 92.7 | 62.7 - 62.7 | 71.1 - 95.9 | 79.1 - 81.3 | 72.0 - 100.6 | 78.0 - 82.7 | 71.9 - 76.9 | 62.7 - 100.6 |

^Shorter all-oral bedaquiline-containing treatment regimen of 9-12 months, as recommended by WHO (2019 rapid communication, 2020 consolidated guidelines)

*excluding shorter 9-12 regimen and early treatment discontinuations

*Table S11.* ***Distribution of*** ***endTB Control Group Regimens by WHO Guideline Period and Conformity with Guidelines***

| **Latest WHO Guidelines to which regimen conformed** | **WHO Guideline Period during which Control Group Participant initiated Treatment** | | | |
|---|---|---|---|---|
| | **<18/12/2018** | **18/12/2018-31/03/2020** | **> 31/03/2020** | **All periods** |
| | **(n=31)** | **(n=35)** | **(n=53)** | **(n=119)** |
| **2022 Guidelines** | | | | **97 (81.5%)** |
| 3 Group A/1-2 Group B/± Group C | 4 (12.9%) | 34 (97.1%) | 47 (88.7%) | 85 (71.4%) |
| Shorter regimen | 0 (0.0%) | 0 (0.0%) | 5 (9.4%) | 5 (4.2%) |
| 2 Group A/1-2 Group B/+ Group C | 5 (16.1%) | 1 (2.9%) | 1 (1.9%) | 7 (5.9%) |
| **earlier WHO Guidelines** | | | | **22 (18.5%)** |
| Injectable/FQ/+ Group C | 22 (71.0%) | 0 (0.0%) | 0 (0.0%) | 22 (18.5%) |

**WHO Drug Group Classification**
Group A: Fluoroquinolone (FQ): levofloxacin, moxifloxacin; bedaquiline, linezolid
Group B: Clofazimine, cycloserine/terizidone
Group C: Ethambutol, isoniazid, pyrazinamide, delamanid, ethionamide-prothionamide, PAS, capreomycin, kanamycin

***Table S12. Unadjusted Analyses of Efficacy of endTB Trial Regimens at 39 Weeks (mITT Population)***

| Week 39 treatment outcome | 9BLMZ (n= 118) | 9BCLLfxZ (n= 115) | 9BDLLfxZ (n= 122) | 9DCLLFxZ (n= 118) | 9DCMZ (n= 104) | Control (n= 119) | Total (N = 696) |
|---|---|---|---|---|---|---|---|
| **Total Favorable** | 106 (89.8%) | 105 (91.3%) | 103 (84.4%) | 94 (79.7%) | 87 (83.7%) | 104 (87.4%) | 599 (86.1%) |
| 95% CI | [82.9%;94.6%] | [84.6%;95.8%] | [76.8%;90.4%] | [71.3%;86.5%] | [75.1%;90.2%] | [80.1%;92.8%] | [83.3%;88.6%] |

***Table S13. Adjusted (pre-specified) Analyses of Efficacy of endTB Trial Regimens at 39 Weeks (mITT Population)***

| W39 treatment outcome | 9BLMZ (n= 118) | 9BCLLfxZ (n= 115) | 9BDLLfxZ (n= 122) | 9DCLLFxZ (n= 118) | 9DCMZ (n= 104) | Control (n= 119) | Total (N = 696) |
|---|---|---|---|---|---|---|---|
| **Total Favorable** | 106 (89.8%) | 105 (91.3%) | 103 (84.4%) | 94 (79.7%) | 87 (83.7%) | 104 (87.4%) | 599 (86.1%) |
| 95% CI | [82.9%;94.6%] | [84.6%;95.8%] | [76.8%;90.4%] | [71.3%;86.5%] | [75.1%;90.2%] | [80.1%;92.8%] | [83.3%;88.6%] |
| Adjusted risk differences* | 4.1% | 5.3% | -2.2% | -6.8% | -2.3% | - | |
| 95% CI around the difference | [-3.9%;12.1%] | [-2.1%;12.7%] | [-10.8%;6.4%] | [-16.0%;2.5%] | [-11.5%;7.0%] | | |

Backward selection: initial model includes significant variables at baseline with p<0.10: BMI, Hepatitis C status, extent of disease.

* significant covariates in final model: BMI. (n=696)

^*No convergence: Poisson regression*

***Table S14. Unadjusted Analyses of Efficacy of endTB Trial Regimens at 39 Weeks (PP Population)***

| W39 treatment outcome | 9BLMZ (n = 98) | 9BCLLfxZ (n = 95) | 9BDLLfxZ (n = 103) | 9DCLLfxZ (n = 96) | 9DCMZ (n = 93) | Control (n = 74) | Total (N = 559) |
|---|---|---|---|---|---|---|---|
| **Total Favorable**^ | 94 (95.9%) | 92 (96.8%) | 96 (93.2%) | 85 (88.5%) | 80 (86.0%) | 71 (95.9%) | 518 (92.7%) |
| 95% CI | [89.9%;98.9%] | [91.0%;99.3%] | [86.5%;97.2%] | [80.4%;94.1%] | [77.3%;92.3%] | [88.6%;99.2%] | [90.2%;94.7%] |

^ All cultures negative

***Table S15. Adjusted (pre-specified) Analyses of Efficacy of endTB Trial Regimens at 39 Weeks (PP Population)***

| W39 treatment outcome | 9BLMZ (n = 98) | 9BCLLfxZ(n = 95) | 9BDLLfxZ (n = 103) | 9DCLLfxZ (n = 96) | 9DCMZ (n = 93) | Control (n = 74) | Total (N = 559) |
|---|---|---|---|---|---|---|---|
| **Total Favorable** | 94 (95.9%) | 92 (96.8%) | 96 (93.2%) | 85 (88.5%) | 80 (86.0%) | 71 (95.9%) | 518 (92.7%) |
| 95% CI | [89.9%;98.9%] | [91.0%;99.3%] | [86.5%;97.2%] | [80.4%;94.1%] | [77.3%;92.3%] | [88.6%;99.2%] | [90.2%;94.7%] |
| Adjusted risk differences*^ | 0.0% | 1.2% | -4.1% | -8.9% | -10.3% | - | |
| 95% CI around the difference | [-6.3%;6.2%] | [-4.9%;7.3%] | [-11.5%;3.3%] | [-17.1%;0.6%] | [-18.9%;-1.7%] | | |

Backward selection: initial model includes significant variables at baseline with p<0.10: BMI, age, HIV, smear result, extent of disease
* significant covariates in final model: BMI, age. (n=559)
*^No convergence: Poisson regression*

***Table S16. Adjusted Analyses of Efficacy of endTB Trial Regimens at 73 Weeks (mITT Population)***

| W73 treatment outcome | 9BLMZ (n = 118) | 9BCLLfxZ (n = 115) | 9BDLLfxZ (n = 122) | 9DCLLfxZ (n = 118) | 9DCMZ (n = 104) | Control (n = 119) | Total (N = 696) |
|---|---|---|---|---|---|---|---|
| **Total Favorable** | 105 (89.0%) | 104 (90.4%) | 104 (85.2%) | 93 (78.8%) | 89 (85.6%) | 96 (80.7%) | 591 (84.9%) |
| 95% CI | [81.9%;94.0%] | [83.5%;95.1%] | [77.7%;91.0%] | [70.3%;85.8%] | [77.3%;91.7%] | [72.4%;87.3%] | [82.0%;87.5%] |
| **Adjusted risk differences-pre-specified analysis*^** | **8.8%** | **9.5%** | **3.9%** | **-1.9%** | **4.4%** | **-** | |
| 95% CI | [-0.6%;18.2%] | [0.4%;18.6%] | [-5.8%;13.6%] | [-12.5%;8.7%] | [-5.7%;14.5%] | | |

Backward selection: initial model includes significant variables at baseline with p<0.10: BMI, age, Hepatitis C status, extent of disease
* significant covariates in final, pre-specified model: Hepatitis C status, extent of disease. (n=691)
*^No convergence: Poisson regression*

| | | | | | | |
|---|---|---|---|---|---|---|
| **Adjusted risk differences-post-hoc analysis**^# | **10.0%** | **10.4%** | **5.3%** | **-0.7%** | **6.3%** | **-** |
| 95% CI | [0.7%;19.2%] | [1.3%;19.6%] | [-4.6%;15.2%] | [-11.3%;9.8%] | [-3.9%;16.4%] | |

#Final, post-hoc model includes: PZA res, HIV, smear res, cavitation (n=685)
*^No convergence: Poisson regression*

***Table S17. Adjusted Analyses of Efficacy of endTB Trial Regimens at 73 Weeks (PP Population)***

| W73 treatment outcome | 9BLMZ (n = 98) | 9BCLLfxZ (n = 95) | 9BDLLfxZ (n = 103) | 9DCLLfxZ (n = 96) | 9DCMZ (n = 93) | Control (n = 74) | Total (N = 559) |
|---|---|---|---|---|---|---|---|
| **Total Favorable** | 94 (95.9%) | 91 (95.8%) | 97 (94.2%) | 82 (85.4%) | 82 (88.2%) | 71 (95.9%) | 517 (92.5%) |
| 95% CI | [89.9%;98.9%] | [89.6%;98.8%] | [87.8%;97.8%] | [76.7%;91.8%] | [79.8%;93.9%] | [88.6%;99.2%] | [90.0%;94.5%] |
| **Adjusted risk differences-pre-specified analysis*^** | **0.1%** | **0.3%** | **-2.9%** | **-12.1%** | **-8.0%** | **-** | |
| 95% CI | [-6.2%; 6.4%] | [-6.1%; 6.7%] | [-10.2%; 4.4%] | [-21.0%; -3.2%] | [-16.2%; 0.2%] | | |
| Backward selection: initial model includes significant variables at baseline with p<0.10: BMI, age, Injectable resistance, smear, extent of disease.<br>* significant covariates in final pre-specified model: age, BMI. (n=559)<br>*^No convergence: Poisson regression* | | | | | | | |
| **Adjusted risk differences-post-hoc analyses[#]^** | **0.0%** | **-0.8%** | **-2.4%** | **-10.2%** | **-8.2%** | **-** | |
| 95% CI | [-6.3%;6.3%] | [-7.3%;5.7%] | [-9.3%;4.5%] | [-19.1%;-1.3%] | [-16.6%;0.1%] | | |

[#]Final, post-hoc model includes: PZA res, HIV, smear res, cavitation (n=559)
*^No convergence: Poisson regression*

***Table S18. Sensitivity Analyses of Efficacy of endTB Trial Regimens at 73 Weeks (Assessable, all-culture mITT, all-DST mITT, and Assessable Reclassified populations)***

| Population | W73 treatment outcome | 9BLMZ | 9BCLLfxZ | 9BDLLfxZ | 9DCLLfxZ | 9DCMZ | Control | Total |
|---|---|---|---|---|---|---|---|---|
| **1. Assessable** | **Total in population** | 97 (100.0%) | 94 (100.0%) | 101 (100.0%) | 94 (100.0%) | 91 (100.0%) | 74 (100.0%) | 551 (100.0%) |
| | **Total Favorable** | 94 (96.9%) | 91 (96.8%) | 97 (96.0%) | 82 (87.2%) | 82 (90.1%) | 71 (95.9%) | 517 (93.8%) |
| | 95% CI | [91.2%; 99.4%] | [91.0%; 99.3%] | [90.2%; 98.9%] | [78.8%; 93.2%] | [82.1%; 95.4%] | [88.6%; 99.2%] | [91.5%; 95.7%] |
| | Difference from control | 1.0% | 0.9% | 0.1% | -8.7% | -5.8% | - | |
| | 95% CI of the difference | [-4.7%; 6.6%] | [-4.9 ; 6.6%] | [-5.8%; 6.0%] | [-16.8%; -0.6%] | [-13.4%; 1.8%] | | |
| **2. all-culture mITT** | **Total in population** | 124 (100.0%) | 119 (100.0%) | 125 (100.0%) | 121 (100.0%) | 113 (100.0%) | 121 (100.0%) | 723 (100.0%) |
| | **Total Favorable** | 110 (88.7%) | 107 (89.9%) | 107 (85.6%) | 96 (79.3%) | 96 (85.0%) | 98 (81.0%) | 614 (84.9%) |
| | 95% CI | [81.8%; 93.7%] | [83.0%; 94.7%] | [78.2%; 91.2%] | [71.0%; 86.2%] | [77.0%; 91.0%] | [72.9%; 87.6%] | [82.1%; 87.5%] |
| | Difference from control | 7.7% | 8.9% | 4.6% | -1.7% | 4.0% | - | |
| | 95% CI of the difference | [-1.2%;16.7%] | [0.1; 17.8%] | [-4.7%; 13.9%] | [-11.7%; 8.4%] | [-5.6%; 13.6%] | | |
| **3. all-DST mITT** | **Total in population** | 118 (100.0%) | 115 (100.0%) | 122 (100.0%) | 118 (100.0%) | 105 (100.0%) | 119 (100.0%) | 697 (100.0%) |
| | **Total Favorable** | 105 (89.0%) | 104 (90.4%) | 104 (85.3%) | 93 (78.8%) | 90 (85.7%) | 96 (80.7%) | 592 (84.9%) |
| | 95% CI | [81.9%; 94.0%] | [83.5%; 95.1%] | [77.7%; 91.0%] | [70.3%; 85.8%] | [77.5%; 91.8%] | [72.4%; 87.3%] | [82.1%; 87.5%] |
| | Difference from control | 8.3% | 9.8% | 4.6% | -1.9% | 5.0% | - | |
| | 95% CI of the difference | [-0.8%;17.4%] | [0.9; 18.7%] | [-4.9%; 14.1%] | [-12.1%; 8.4%] | [-4.7%; 14.8%] | | |
| **4. Assessable reclassifying "Unfavorable because unassessable" to Week 39 outcome** | **Total in population** | 98 (100.0%) | 95 (100.0%) | 103 (100.0%) | 96 (100.0%) | 93 (100.0%) | 74 (100.0%) | 559 (100.0%) |
| | **Total Favorable** | 95 (96.9%) | 92 (96.8%) | 98 (95.2%) | 83 (86.5%) | 82 (88.2%) | 71 (96.0%) | 521 (93.2%) |
| | 95% CI | [91.3%; 99.4%] | [91.0%; 99.3%] | [89.0%; 98.4%] | [78.0%; 92.6%] | [79.8%; 93.9%] | [88.6%; 99.2%] | [90.8%; 95.1%] |
| | Difference from control | 1.0% | 0.9% | -0.8% | -9.5% | -7.8% | - | |
| | 95% CI of the difference | [-4.6%; 6.6%] | [-4.8 ; 6.6%] | [-6.9%; 5.3%] | [-17.7%; -1.3%] | [-15.7%; 0.2%] | | |

Note: Sensitivity analysis excluding participants from the control arm who received a shortened regimen, if the proportion exceeds 10% was not performed as the proportion of shortened regimen received in the control is 4.8% (6/126). Multiple imputation to impute missing data was not performed as missing values in the final multivariable model reduce the relevant analysis population by only 0.7% (<20%).

***Table S19. Sensitivity Analyses of Efficacy of endTB Trial Regimens at 104 Weeks (Assessable, all-culture mITT, all-DST mITT, and Assessable Reclassified populations)***

| N° | W104 treatment outcome | 9BLMZ | 9BCLLfxZ | 9BDLLfxZ | 9DCLLfxZ | 9DCMZ | Control | Total |
|---|---|---|---|---|---|---|---|---|
| **1. Assessable** | **Total in population** | 94 (100.0%) | 92 (100.0%) | 101 (100.0%) | 91 (100.0%) | 91 (100.0%) | 73 (100.0%) | 542 (100.0%) |
| | **Total Favorable** | 91 (96.8%) | 90 (97.8%) | 97 (96.0%) | 79 (86.8%) | 81 (89.0%) | 69 (94.5%) | 507 (93.5%) |
| | 95% CI | [91.0%; 99.3%] | [92.4%; 99.7%] | [90.2%; 98.9%] | [78.1%; 93.0%] | [80.7%; 94.6%] | [86.6%; 98.5%] | [91.1%; 95.5%] |
| | Difference from control | 2.3% | 3.3% | 1.5% | -7.7% | -5.5% | - | |
| | 95% CI of the difference | [-4.0%; 8.6%] | [-2.7%; 9.3%] | [-4.9%; 8.0%] | [-16.4%; 1.0%] | [-13.8%; 2.8%] | | |
| **2. all-culture mITT** | **Total in population** | 124 (100.0%) | 119 (100.0%) | 125 (100.0%) | 121 (100.0%) | 113 (100.0%) | 121 (100.0%) | 723 (100.0%) |
| | **Total Favorable** | 106 (85.5%) | 105 (88.2%) | 106 (84.8%) | 91 (75.2%) | 95 (84.1%) | 94 (77.7%) | 597 (82.6%) |
| | 95% CI | [78.0%; 91.2%] | [81.0%; 93.4%] | [77.3%; 90.6%] | [66.5%; 82.6%] | [76.0%; 90.3%] | [69.2%; 84.8%] | [79.6%; 85.3%] |
| | Difference from control | 7.8% | 10.5% | 7.1% | -2.5% | 6.4% | - | |
| | 95% CI of the difference | [-1.9%; 17.5%] | [1.1%; 20.0%] | [-2.6%; 16.8%] | [-13.2%; 8.2%] | [-3.6%; 16.4%] | | |
| **3. all-DST mITT** | **Total in population** | 118 (100.0%) | 115 (100.0%) | 122 (100.0%) | 118 (100.0%) | 105 (100.0%) | 119 (100.0%) | 697 (100.0%) |
| | **Total Favorable** | 101 (85.6%) | 102 (88.7%) | 103 (84.4%) | 88 (74.6%) | 89 (84.8%) | 92 (77.3%) | 575 (82.5%) |
| | 95% CI | [77.9%; 91.4%] | [81.4%; 93.8%] | [76.8%; 90.4%] | [65.7%; 82.1%] | [76.4%; 91.0%] | [68.7%; 84.5%] | [79.5%; 85.2%] |
| | Difference from control | 8.3% | 11.4% | 7.1% | -2.7% | 7.5% | - | |
| | 95% CI of the difference | [-1.6%;18.1%] | [1.9; 20.9%] | [-2.8%; 17.0%] | [-13.6%; 8.1%] | [-2.7%; 17.6%] | | |
| **4. Assessable reclassifying “LTFU” or “unfavorable because unassessable” to Week 73 outcome** | **Total in population** | 98 (100.0%) | 95 (100.0%) | 103 (100.0%) | 96 (100.0%) | 93 (100.0%) | 74 (100.0%) | 559 (100.0%) |
| | **Total Favorable** | 91 (92.9%) | 90 (94.7%) | 97 (94.2%) | 79 (82.3%) | 81 (87.1%) | 69 (93.2%) | 507 (90.7%) |
| | 95% CI | [85.8%; 97.1%] | [88.1%; 98.3%] | [87.8%; 97.8%] | [73.2%; 89.3%] | [78.5%; 93.2%] | [84.9%; 97.8%] | [88.0%; 93.0%] |
| | Difference from control | -0.4% | 1.5% | 0.9% | -11.0% | -6.1% | - | |
| | 95% CI of the difference | [-8.0%; 7.3%] | [-5.8%; 8.8%] | [-6.4% ; 8.2%] | [-20.5%; -0.1%] | [-15.0%; 2.7%] | | |

Note: Sensitivity analysis excluding participants from the control arm who received a shortened regimen, if the proportion exceeds 10% was not performed as the proportion of shortened regimen received in the control is 4.8% (6/126). Multiple imputation to impute missing data was not performed as missing values in the final multivariable model reduce the relevant analysis population by only 0.7% (<20%).

*Table S20. Number (%) of participants experiencing adverse events (AEs) of grade 3 or higher by SOC and PT by Week 73 (Safety Population)*

| *Event*<br>**System Organ Class (SOC)**<br>- Preferred Term (PT) | **9BLMZ (n = 126)** | **9BCLLfxZ (n = 122)** | **9BDLLfxZ (n = 127)** | **9DCLLfxZ (n = 124)** | **9DCMZ (n = 120)** | **Control (n = 126)** | **Total* (N = 745)** |
|---|---|---|---|---|---|---|---|
| **Investigations** | **43 (34.1%)** | **43 (35.2%)** | **39 (30.7%)** | **46 (37.1%)** | **50 (41.7%)** | **39 (31.0%)** | **260 (34.9%)** |
| - Creatinine renal clearance decreased | 21 (16.7%) | 23 (18.9%) | 29 (22.8%) | 37 (29.8%) | 36 (30.0%) | 23 (18.3%) | 169 (22.7%) |
| - Alanine aminotransferase increased | 14 (11.1%) | 10 (8.2%) | 5 (3.9%) | 8 (6.5%) | 7 (5.8%) | 3 (2.4%) | 47 (6.3%) |
| - Blood magnesium increased | 6 (4.8%) | 6 (4.9%) | 4 (3.1%) | 1 (0.8%) | 5 (4.2%) | 3 (2.4%) | 25 (3.4%) |
| - Aspartate aminotransferase increased | 4 (3.2%) | 7 (5.7%) | 4 (3.1%) | 3 (2.4%) | 3 (2.5%) | 4 (3.2%) | 25 (3.4%) |
| - Bilirubin conjugated increased | 2 (1.6%) | 3 (2.5%) | 3 (2.4%) | 1 (0.8%) | 1 (0.8%) | 4 (3.2%) | 14 (1.9%) |
| - Lymphocyte count decreased | 4 (3.2%) | 1 (0.8%) | 2 (1.6%) | 0 (0.0%) | 2 (1.7%) | 1 (0.8%) | 10 (1.3%) |
| - Electrocardiogram QT prolonged | 0 (0.0%) | 4 (3.3%) | 0 (0.0%) | 0 (0.0%) | 5 (4.2%) | 0 (0.0%) | 9 (1.2%) |
| - Transaminases increased | 1 (0.8%) | 1 (0.8%) | 1 (0.8%) | 3 (2.4%) | 3 (2.5%) | 0 (0.0%) | 9 (1.2%) |
| - Gamma-glutamyltransferase increased | 1 (0.8%) | 1 (0.8%) | 0 (0.0%) | 1 (0.8%) | 1 (0.8%) | 2 (1.6%) | 6 (0.8%) |
| - Neutrophil count decreased | 1 (0.8%) | 0 (0.0%) | 0 (0.0%) | 0 (0.0%) | 1 (0.8%) | 2 (1.6%) | 4 (0.5%) |
| - Blood bilirubin increased | 0 (0.0%) | 1 (0.8%) | 1 (0.8%) | 0 (0.0%) | 1 (0.8%) | 1 (0.8%) | 4 (0.5%) |
| - Glycosylated haemoglobin increased | 2 (1.6%) | 1 (0.8%) | 0 (0.0%) | 0 (0.0%) | 1 (0.8%) | 0 (0.0%) | 4 (0.5%) |
| - Hepatic enzyme increased | 2 (1.6%) | 1 (0.8%) | 0 (0.0%) | 0 (0.0%) | 0 (0.0%) | 0 (0.0%) | 3 (0.4%) |
| - Blood potassium increased | 0 (0.0%) | 0 (0.0%) | 0 (0.0%) | 0 (0.0%) | 2 (1.7%) | 1 (0.8%) | 3 (0.4%) |
| - White blood cell count decreased | 0 (0.0%) | 0 (0.0%) | 0 (0.0%) | 0 (0.0%) | 1 (0.8%) | 2 (1.6%) | 3 (0.4%) |
| - Weight increased | 0 (0.0%) | 0 (0.0%) | 0 (0.0%) | 0 (0.0%) | 0 (0.0%) | 2 (1.6%) | 2 (0.3%) |
| - Haemoglobin increased | 2 (1.6%) | 0 (0.0%) | 0 (0.0%) | 0 (0.0%) | 0 (0.0%) | 0 (0.0%) | 2 (0.3%) |
| - Eosinophil count increased | 0 (0.0%) | 1 (0.8%) | 0 (0.0%) | 0 (0.0%) | 0 (0.0%) | 0 (0.0%) | 1 (0.1%) |
| - Blood creatinine decreased | 0 (0.0%) | 0 (0.0%) | 0 (0.0%) | 0 (0.0%) | 1 (0.8%) | 0 (0.0%) | 1 (0.1%) |
| - Blood creatinine increased | 0 (0.0%) | 0 (0.0%) | 1 (0.8%) | 0 (0.0%) | 0 (0.0%) | 0 (0.0%) | 1 (0.1%) |
| - Blood calcium decreased | 0 (0.0%) | 1 (0.8%) | 0 (0.0%) | 0 (0.0%) | 0 (0.0%) | 0 (0.0%) | 1 (0.1%) |
| - Electrocardiogram QRS complex prolonged | 0 (0.0%) | 0 (0.0%) | 0 (0.0%) | 0 (0.0%) | 0 (0.0%) | 1 (0.8%) | 1 (0.1%) |
| - Blood uric acid increased | 0 (0.0%) | 1 (0.8%) | 0 (0.0%) | 0 (0.0%) | 0 (0.0%) | 0 (0.0%) | 1 (0.1%) |
| - Blood urea increased | 0 (0.0%) | 0 (0.0%) | 1 (0.8%) | 0 (0.0%) | 0 (0.0%) | 0 (0.0%) | 1 (0.1%) |
| - Psychiatric evaluation abnormal | 0 (0.0%) | 0 (0.0%) | 0 (0.0%) | 0 (0.0%) | 1 (0.8%) | 0 (0.0%) | 1 (0.1%) |
| **Nervous system disorders** | **8 (6.3%)** | **11 (9.0%)** | **16 (12.6%)** | **11 (8.9%)** | **4 (3.3%)** | **14 (11.1%)** | **64 (8.6%)** |
| - Neuropathy peripheral | 3 (2.4%) | 5 (4.1%) | 7 (5.5%) | 2 (1.6%) | 1 (0.8%) | 6 (4.8%) | 24 (3.2%) |
| - Paraesthesia | 2 (1.6%) | 3 (2.5%) | 4 (3.1%) | 4 (3.2%) | 1 (0.8%) | 6 (4.8%) | 20 (2.7%) |
| - Areflexia | 0 (0.0%) | 0 (0.0%) | 1 (0.8%) | 2 (1.6%) | 3 (2.5%) | 1 (0.8%) | 7 (0.9%) |
| - Polyneuropathy | 1 (0.8%) | 0 (0.0%) | 2 (1.6%) | 1 (0.8%) | 2 (1.7%) | 0 (0.0%) | 6 (0.8%) |

| *Event*<br>**System Organ Class (SOC)**<br>- Preferred Term (PT) | **9BLMZ (n = 126)** | **9BCLLfxZ (n = 122)** | **9BDLLfxZ (n = 127)** | **9DCLLfxZ (n = 124)** | **9DCMZ (n = 120)** | **Control (n = 126)** | **Total* (N = 745)** |
|---|---|---|---|---|---|---|---|
| - Optic neuritis | 0 (0.0%) | 0 (0.0%) | 0 (0.0%) | 1 (0.8%) | 0 (0.0%) | 1 (0.8%) | 2 (0.3%) |
| - Hyporeflexia | 2 (1.6%) | 0 (0.0%) | 0 (0.0%) | 0 (0.0%) | 0 (0.0%) | 0 (0.0%) | 2 (0.3%) |
| - Headache | 0 (0.0%) | 1 (0.8%) | 1 (0.8%) | 0 (0.0%) | 0 (0.0%) | 0 (0.0%) | 2 (0.3%) |
| - Sciatica | 0 (0.0%) | 0 (0.0%) | 1 (0.8%) | 0 (0.0%) | 0 (0.0%) | 0 (0.0%) | 1 (0.1%) |
| - Facial paralysis | 0 (0.0%) | 0 (0.0%) | 1 (0.8%) | 0 (0.0%) | 0 (0.0%) | 0 (0.0%) | 1 (0.1%) |
| - Idiopathic intracranial hypertension | 0 (0.0%) | 0 (0.0%) | 0 (0.0%) | 0 (0.0%) | 0 (0.0%) | 1 (0.8%) | 1 (0.1%) |
| - Syncope | 0 (0.0%) | 0 (0.0%) | 0 (0.0%) | 1 (0.8%) | 0 (0.0%) | 0 (0.0%) | 1 (0.1%) |
| - Epilepsy | 0 (0.0%) | 1 (0.8%) | 0 (0.0%) | 0 (0.0%) | 0 (0.0%) | 0 (0.0%) | 1 (0.1%) |
| - Transient ischaemic attack | 0 (0.0%) | 1 (0.8%) | 0 (0.0%) | 0 (0.0%) | 0 (0.0%) | 0 (0.0%) | 1 (0.1%) |
| **Metabolism and nutrition disorders** | **10 (7.9%)** | **6 (4.9%)** | **10 (7.9%)** | **8 (6.5%)** | **10 (8.3%)** | **11 (8.7%)** | **55 (7.4%)** |
| - Hypermagnesaemia | 6 (4.8%) | 3 (2.5%) | 5 (3.9%) | 3 (2.4%) | 3 (2.5%) | 7 (5.6%) | 27 (3.6%) |
| - Hyperglycaemia | 2 (1.6%) | 2 (1.6%) | 0 (0.0%) | 2 (1.6%) | 2 (1.7%) | 1 (0.8%) | 9 (1.2%) |
| - Hyperkalaemia | 2 (1.6%) | 0 (0.0%) | 0 (0.0%) | 2 (1.6%) | 1 (0.8%) | 1 (0.8%) | 6 (0.8%) |
| - Hypoalbuminaemia | 1 (0.8%) | 1 (0.8%) | 0 (0.0%) | 0 (0.0%) | 1 (0.8%) | 0 (0.0%) | 3 (0.4%) |
| - Hyperuricaemia | 0 (0.0%) | 0 (0.0%) | 1 (0.8%) | 0 (0.0%) | 1 (0.8%) | 0 (0.0%) | 2 (0.3%) |
| - Obesity | 0 (0.0%) | 0 (0.0%) | 0 (0.0%) | 0 (0.0%) | 1 (0.8%) | 1 (0.8%) | 2 (0.3%) |
| - Diabetes mellitus inadequate control | 0 (0.0%) | 0 (0.0%) | 0 (0.0%) | 0 (0.0%) | 2 (1.7%) | 0 (0.0%) | 2 (0.3%) |
| - Hypomagnesaemia | 0 (0.0%) | 0 (0.0%) | 1 (0.8%) | 0 (0.0%) | 0 (0.0%) | 1 (0.8%) | 2 (0.3%) |
| - Hypercalcaemia | 0 (0.0%) | 0 (0.0%) | 1 (0.8%) | 0 (0.0%) | 0 (0.0%) | 0 (0.0%) | 1 (0.1%) |
| - Hypokalaemia | 0 (0.0%) | 0 (0.0%) | 1 (0.8%) | 0 (0.0%) | 0 (0.0%) | 0 (0.0%) | 1 (0.1%) |
| - Hypertriglyceridaemia | 0 (0.0%) | 0 (0.0%) | 0 (0.0%) | 0 (0.0%) | 1 (0.8%) | 0 (0.0%) | 1 (0.1%) |
| - Decreased appetite | 0 (0.0%) | 0 (0.0%) | 1 (0.8%) | 0 (0.0%) | 0 (0.0%) | 0 (0.0%) | 1 (0.1%) |
| - Hyponatraemia | 0 (0.0%) | 0 (0.0%) | 0 (0.0%) | 1 (0.8%) | 0 (0.0%) | 0 (0.0%) | 1 (0.1%) |
| - Hypocalcaemia | 0 (0.0%) | 0 (0.0%) | 0 (0.0%) | 1 (0.8%) | 0 (0.0%) | 0 (0.0%) | 1 (0.1%) |
| **Eye disorders** | **4 (3.2%)** | **12 (9.8%)** | **10 (7.9%)** | **8 (6.5%)** | **7 (5.8%)** | **11 (8.7%)** | **52 (7.0%)** |
| - Visual acuity reduced | 3 (2.4%) | 6 (4.9%) | 7 (5.5%) | 4 (3.2%) | 4 (3.3%) | 7 (5.6%) | 31 (4.2%) |
| - Visual impairment | 0 (0.0%) | 2 (1.6%) | 3 (2.4%) | 2 (1.6%) | 2 (1.7%) | 2 (1.6%) | 11 (1.5%) |
| - Cataract | 1 (0.8%) | 1 (0.8%) | 0 (0.0%) | 1 (0.8%) | 0 (0.0%) | 2 (1.6%) | 5 (0.7%) |
| - Refraction disorder | 0 (0.0%) | 1 (0.8%) | 0 (0.0%) | 1 (0.8%) | 0 (0.0%) | 0 (0.0%) | 2 (0.3%) |
| - Retinopathy | 0 (0.0%) | 1 (0.8%) | 0 (0.0%) | 0 (0.0%) | 1 (0.8%) | 0 (0.0%) | 2 (0.3%) |
| - Myopia | 0 (0.0%) | 0 (0.0%) | 0 (0.0%) | 0 (0.0%) | 1 (0.8%) | 0 (0.0%) | 1 (0.1%) |
| - Presbyopia | 0 (0.0%) | 1 (0.8%) | 0 (0.0%) | 0 (0.0%) | 0 (0.0%) | 0 (0.0%) | 1 (0.1%) |
| - Vision blurred | 0 (0.0%) | 0 (0.0%) | 0 (0.0%) | 0 (0.0%) | 0 (0.0%) | 1 (0.8%) | 1 (0.1%) |
| - Optic neuropathy | 0 (0.0%) | 1 (0.8%) | 0 (0.0%) | 0 (0.0%) | 0 (0.0%) | 0 (0.0%) | 1 (0.1%) |
| - Hypermetropia | 0 (0.0%) | 1 (0.8%) | 0 (0.0%) | 0 (0.0%) | 0 (0.0%) | 0 (0.0%) | 1 (0.1%) |

| *Event*<br>**System Organ Class (SOC)**<br>- Preferred Term (PT) | **9BLMZ (n = 126)** | **9BCLLfxZ (n = 122)** | **9BDLLfxZ (n = 127)** | **9DCLLfxZ (n = 124)** | **9DCMZ (n = 120)** | **Control (n = 126)** | **Total* (N = 745)** |
|---|---|---|---|---|---|---|---|
| - Eye haemorrhage | 0 (0.0%) | 1 (0.8%) | 0 (0.0%) | 0 (0.0%) | 0 (0.0%) | 0 (0.0%) | 1 (0.1%) |
| **Blood and lymphatic system disorders** | **6 (4.8%)** | **8 (6.6%)** | **9 (7.1%)** | **13 (10.5%)** | **5 (4.2%)** | **10 (7.9%)** | **51 (6.8%)** |
| - Anaemia | 4 (3.2%) | 6 (4.9%) | 9 (7.1%) | 9 (7.3%) | 3 (2.5%) | 7 (5.6%) | 38 (5.1%) |
| - Lymphopenia | 1 (0.8%) | 0 (0.0%) | 0 (0.0%) | 4 (3.2%) | 1 (0.8%) | 3 (2.4%) | 9 (1.2%) |
| - Neutropenia | 0 (0.0%) | 1 (0.8%) | 0 (0.0%) | 1 (0.8%) | 1 (0.8%) | 1 (0.8%) | 4 (0.5%) |
| - Leukopenia | 0 (0.0%) | 1 (0.8%) | 0 (0.0%) | 1 (0.8%) | 0 (0.0%) | 1 (0.8%) | 3 (0.4%) |
| - Bicytopenia | 0 (0.0%) | 0 (0.0%) | 0 (0.0%) | 0 (0.0%) | 1 (0.8%) | 1 (0.8%) | 2 (0.3%) |
| - Thrombocytopenia | 1 (0.8%) | 0 (0.0%) | 0 (0.0%) | 0 (0.0%) | 0 (0.0%) | 1 (0.8%) | 2 (0.3%) |
| - Pancytopenia | 0 (0.0%) | 0 (0.0%) | 0 (0.0%) | 1 (0.8%) | 0 (0.0%) | 0 (0.0%) | 1 (0.1%) |
| - Bone marrow failure | 0 (0.0%) | 1 (0.8%) | 0 (0.0%) | 0 (0.0%) | 0 (0.0%) | 0 (0.0%) | 1 (0.1%) |
| - Leukocytosis | 0 (0.0%) | 0 (0.0%) | 0 (0.0%) | 0 (0.0%) | 0 (0.0%) | 1 (0.8%) | 1 (0.1%) |
| - Myelosuppression | 0 (0.0%) | 0 (0.0%) | 0 (0.0%) | 1 (0.8%) | 0 (0.0%) | 0 (0.0%) | 1 (0.1%) |
| **Ear and labyrinth disorders** | **10 (7.9%)** | **11 (9.0%)** | **6 (4.7%)** | **4 (3.2%)** | **9 (7.5%)** | **9 (7.1%)** | **49 (6.6%)** |
| - Hypoacusis | 8 (6.3%) | 9 (7.4%) | 4 (3.1%) | 4 (3.2%) | 7 (5.8%) | 8 (6.3%) | 40 (5.4%) |
| - Deafness | 2 (1.6%) | 2 (1.6%) | 1 (0.8%) | 0 (0.0%) | 2 (1.7%) | 1 (0.8%) | 8 (1.1%) |
| - Deafness unilateral | 0 (0.0%) | 0 (0.0%) | 1 (0.8%) | 0 (0.0%) | 0 (0.0%) | 0 (0.0%) | 1 (0.1%) |
| - Tinnitus | 0 (0.0%) | 1 (0.8%) | 0 (0.0%) | 0 (0.0%) | 0 (0.0%) | 0 (0.0%) | 1 (0.1%) |
| **Infections and infestations** | **12 (9.5%)** | **3 (2.5%)** | **6 (4.7%)** | **6 (4.8%)** | **6 (5.0%)** | **4 (3.2%)** | **37 (5.0%)** |
| - Pneumonia | 4 (3.2%) | 0 (0.0%) | 3 (2.4%) | 0 (0.0%) | 1 (0.8%) | 2 (1.6%) | 10 (1.3%) |
| - COVID-19 | 2 (1.6%) | 0 (0.0%) | 0 (0.0%) | 0 (0.0%) | 1 (0.8%) | 1 (0.8%) | 4 (0.5%) |
| - Hepatitis A | 1 (0.8%) | 0 (0.0%) | 0 (0.0%) | 2 (1.6%) | 0 (0.0%) | 0 (0.0%) | 3 (0.4%) |
| - COVID-19 pneumonia | 0 (0.0%) | 2 (1.6%) | 0 (0.0%) | 0 (0.0%) | 1 (0.8%) | 0 (0.0%) | 3 (0.4%) |
| - Sepsis | 0 (0.0%) | 1 (0.8%) | 0 (0.0%) | 1 (0.8%) | 1 (0.8%) | 0 (0.0%) | 3 (0.4%) |
| - Pneumocystis jirovecii pneumonia | 1 (0.8%) | 0 (0.0%) | 0 (0.0%) | 1 (0.8%) | 0 (0.0%) | 0 (0.0%) | 2 (0.3%) |
| - Pneumonia bacterial | 0 (0.0%) | 0 (0.0%) | 1 (0.8%) | 0 (0.0%) | 0 (0.0%) | 0 (0.0%) | 1 (0.1%) |
| - Infective glossitis | 0 (0.0%) | 0 (0.0%) | 0 (0.0%) | 1 (0.8%) | 0 (0.0%) | 0 (0.0%) | 1 (0.1%) |
| - Pulmonary tuberculoma | 0 (0.0%) | 0 (0.0%) | 1 (0.8%) | 0 (0.0%) | 0 (0.0%) | 0 (0.0%) | 1 (0.1%) |
| - Respiratory tract infection | 0 (0.0%) | 0 (0.0%) | 1 (0.8%) | 0 (0.0%) | 0 (0.0%) | 0 (0.0%) | 1 (0.1%) |
| - Wound infection | 1 (0.8%) | 0 (0.0%) | 0 (0.0%) | 0 (0.0%) | 0 (0.0%) | 0 (0.0%) | 1 (0.1%) |
| - Tonsillitis | 0 (0.0%) | 0 (0.0%) | 0 (0.0%) | 0 (0.0%) | 0 (0.0%) | 1 (0.8%) | 1 (0.1%) |
| - Abscess soft tissue | 0 (0.0%) | 0 (0.0%) | 0 (0.0%) | 0 (0.0%) | 1 (0.8%) | 0 (0.0%) | 1 (0.1%) |
| - Pyelonephritis | 0 (0.0%) | 0 (0.0%) | 0 (0.0%) | 0 (0.0%) | 1 (0.8%) | 0 (0.0%) | 1 (0.1%) |
| - Tuberculosis | 1 (0.8%) | 0 (0.0%) | 0 (0.0%) | 0 (0.0%) | 0 (0.0%) | 0 (0.0%) | 1 (0.1%) |
| - Appendicitis | 0 (0.0%) | 0 (0.0%) | 0 (0.0%) | 0 (0.0%) | 0 (0.0%) | 1 (0.8%) | 1 (0.1%) |
| - Gastroenteritis | 0 (0.0%) | 0 (0.0%) | 0 (0.0%) | 1 (0.8%) | 0 (0.0%) | 0 (0.0%) | 1 (0.1%) |

| *Event*<br>**System Organ Class (SOC)**<br>- Preferred Term (PT) | **9BLMZ (n = 126)** | **9BCLLfxZ (n = 122)** | **9BDLLfxZ (n = 127)** | **9DCLLfxZ (n = 124)** | **9DCMZ (n = 120)** | **Control (n = 126)** | **Total* (N = 745)** |
|---|---|---|---|---|---|---|---|
| - Herpes zoster | 0 (0.0%) | 0 (0.0%) | 0 (0.0%) | 0 (0.0%) | 1 (0.8%) | 0 (0.0%) | 1 (0.1%) |
| - Skin infection | 1 (0.8%) | 0 (0.0%) | 0 (0.0%) | 0 (0.0%) | 0 (0.0%) | 0 (0.0%) | 1 (0.1%) |
| - HIV infection | 0 (0.0%) | 0 (0.0%) | 0 (0.0%) | 0 (0.0%) | 0 (0.0%) | 1 (0.8%) | 1 (0.1%) |
| - Cellulitis | 0 (0.0%) | 0 (0.0%) | 1 (0.8%) | 0 (0.0%) | 0 (0.0%) | 0 (0.0%) | 1 (0.1%) |
| - Septic shock | 1 (0.8%) | 0 (0.0%) | 0 (0.0%) | 0 (0.0%) | 0 (0.0%) | 0 (0.0%) | 1 (0.1%) |
| - Pneumonia staphylococcal | 0 (0.0%) | 0 (0.0%) | 1 (0.8%) | 0 (0.0%) | 0 (0.0%) | 0 (0.0%) | 1 (0.1%) |
| - Post procedural sepsis | 0 (0.0%) | 0 (0.0%) | 0 (0.0%) | 1 (0.8%) | 0 (0.0%) | 0 (0.0%) | 1 (0.1%) |
| - Dengue fever | 1 (0.8%) | 0 (0.0%) | 0 (0.0%) | 0 (0.0%) | 0 (0.0%) | 0 (0.0%) | 1 (0.1%) |
| **Hepatobiliary disorders** | **6 (4.8%)** | **4 (3.3%)** | **2 (1.6%)** | **7 (5.6%)** | **5 (4.2%)** | **3 (2.4%)** | **27 (3.6%)** |
| - Hepatotoxicity | 5 (4.0%) | 0 (0.0%) | 1 (0.8%) | 3 (2.4%) | 1 (0.8%) | 1 (0.8%) | 11 (1.5%) |
| - Drug-induced liver injury | 1 (0.8%) | 3 (2.5%) | 1 (0.8%) | 3 (2.4%) | 0 (0.0%) | 0 (0.0%) | 8 (1.1%) |
| - Hepatitis | 0 (0.0%) | 1 (0.8%) | 0 (0.0%) | 0 (0.0%) | 2 (1.7%) | 2 (1.6%) | 5 (0.7%) |
| - Hyperbilirubinaemia | 0 (0.0%) | 0 (0.0%) | 0 (0.0%) | 1 (0.8%) | 1 (0.8%) | 0 (0.0%) | 2 (0.3%) |
| - Acute on chronic liver failure | 0 (0.0%) | 0 (0.0%) | 0 (0.0%) | 0 (0.0%) | 1 (0.8%) | 0 (0.0%) | 1 (0.1%) |
| **Musculoskeletal and connective tissue disorders** | **1 (0.8%)** | **6 (4.9%)** | **4 (3.1%)** | **4 (3.2%)** | **2 (1.7%)** | **5 (4.0%)** | **22 (3.0%)** |
| - Arthralgia | 0 (0.0%) | 4 (3.3%) | 3 (2.4%) | 2 (1.6%) | 1 (0.8%) | 5 (4.0%) | 15 (2.0%) |
| - Myalgia | 0 (0.0%) | 2 (1.6%) | 1 (0.8%) | 0 (0.0%) | 0 (0.0%) | 1 (0.8%) | 4 (0.5%) |
| - Back pain | 1 (0.8%) | 0 (0.0%) | 0 (0.0%) | 1 (0.8%) | 0 (0.0%) | 1 (0.8%) | 3 (0.4%) |
| - Rotator cuff syndrome | 0 (0.0%) | 1 (0.8%) | 0 (0.0%) | 1 (0.8%) | 0 (0.0%) | 0 (0.0%) | 2 (0.3%) |
| - Osteoarthritis | 0 (0.0%) | 0 (0.0%) | 0 (0.0%) | 0 (0.0%) | 1 (0.8%) | 0 (0.0%) | 1 (0.1%) |
| - Arthropathy | 0 (0.0%) | 1 (0.8%) | 0 (0.0%) | 0 (0.0%) | 0 (0.0%) | 0 (0.0%) | 1 (0.1%) |
| **Gastrointestinal disorders** | **1 (0.8%)** | **2 (1.6%)** | **3 (2.4%)** | **5 (4.0%)** | **2 (1.7%)** | **3 (2.4%)** | **16 (2.1%)** |
| - Vomiting | 0 (0.0%) | 1 (0.8%) | 0 (0.0%) | 3 (2.4%) | 0 (0.0%) | 3 (2.4%) | 7 (0.9%) |
| - Nausea | 0 (0.0%) | 1 (0.8%) | 2 (1.6%) | 1 (0.8%) | 1 (0.8%) | 0 (0.0%) | 5 (0.7%) |
| - Gastritis | 0 (0.0%) | 0 (0.0%) | 0 (0.0%) | 0 (0.0%) | 1 (0.8%) | 0 (0.0%) | 1 (0.1%) |
| - Dyspepsia | 0 (0.0%) | 0 (0.0%) | 1 (0.8%) | 0 (0.0%) | 0 (0.0%) | 0 (0.0%) | 1 (0.1%) |
| - Acute abdomen | 0 (0.0%) | 0 (0.0%) | 1 (0.8%) | 0 (0.0%) | 0 (0.0%) | 0 (0.0%) | 1 (0.1%) |
| - Pancreatitis | 0 (0.0%) | 0 (0.0%) | 0 (0.0%) | 0 (0.0%) | 0 (0.0%) | 1 (0.8%) | 1 (0.1%) |
| - Abdominal pain | 0 (0.0%) | 0 (0.0%) | 0 (0.0%) | 1 (0.8%) | 0 (0.0%) | 0 (0.0%) | 1 (0.1%) |
| - Small intestinal obstruction | 0 (0.0%) | 0 (0.0%) | 0 (0.0%) | 1 (0.8%) | 0 (0.0%) | 0 (0.0%) | 1 (0.1%) |
| - Upper gastrointestinal haemorrhage | 0 (0.0%) | 0 (0.0%) | 0 (0.0%) | 0 (0.0%) | 1 (0.8%) | 0 (0.0%) | 1 (0.1%) |
| - Abdominal hernia | 1 (0.8%) | 0 (0.0%) | 0 (0.0%) | 0 (0.0%) | 0 (0.0%) | 0 (0.0%) | 1 (0.1%) |
| **Respiratory, thoracic and mediastinal disorders** | **0 (0.0%)** | **2 (1.6%)** | **4 (3.1%)** | **2 (1.6%)** | **3 (2.5%)** | **4 (3.2%)** | **15 (2.0%)** |
| - Haemoptysis | 0 (0.0%) | 0 (0.0%) | 0 (0.0%) | 0 (0.0%) | 1 (0.8%) | 2 (1.6%) | 3 (0.4%) |

| *Event*<br>**System Organ Class (SOC)**<br>- Preferred Term (PT) | **9BLMZ (n = 126)** | **9BCLLfxZ (n = 122)** | **9BDLLfxZ (n = 127)** | **9DCLLfxZ (n = 124)** | **9DCMZ (n = 120)** | **Control (n = 126)** | **Total* (N = 745)** |
|---|---|---|---|---|---|---|---|
| - Pneumothorax | 0 (0.0%) | 0 (0.0%) | 0 (0.0%) | 1 (0.8%) | 1 (0.8%) | 1 (0.8%) | 3 (0.4%) |
| - Dyspnoea | 0 (0.0%) | 1 (0.8%) | 1 (0.8%) | 1 (0.8%) | 0 (0.0%) | 0 (0.0%) | 3 (0.4%) |
| - Cough | 0 (0.0%) | 0 (0.0%) | 1 (0.8%) | 0 (0.0%) | 2 (1.7%) | 0 (0.0%) | 3 (0.4%) |
| - Respiratory failure | 0 (0.0%) | 0 (0.0%) | 1 (0.8%) | 0 (0.0%) | 0 (0.0%) | 1 (0.8%) | 2 (0.3%) |
| - Acute pulmonary oedema | 0 (0.0%) | 1 (0.8%) | 0 (0.0%) | 0 (0.0%) | 0 (0.0%) | 0 (0.0%) | 1 (0.1%) |
| - Pulmonary cavitation | 0 (0.0%) | 0 (0.0%) | 0 (0.0%) | 0 (0.0%) | 0 (0.0%) | 1 (0.8%) | 1 (0.1%) |
| - Acute respiratory failure | 0 (0.0%) | 0 (0.0%) | 1 (0.8%) | 0 (0.0%) | 0 (0.0%) | 0 (0.0%) | 1 (0.1%) |
| **Skin and subcutaneous tissue disorders** | **1 (0.8%)** | **2 (1.6%)** | **2 (1.6%)** | **3 (2.4%)** | **3 (2.5%)** | **1 (0.8%)** | **12 (1.6%)** |
| - Rash | 0 (0.0%) | 1 (0.8%) | 1 (0.8%) | 2 (1.6%) | 0 (0.0%) | 0 (0.0%) | 4 (0.5%) |
| - Pruritus | 0 (0.0%) | 1 (0.8%) | 0 (0.0%) | 0 (0.0%) | 2 (1.7%) | 0 (0.0%) | 3 (0.4%) |
| - Drug reaction with eosinophilia and systemic symptoms | 0 (0.0%) | 0 (0.0%) | 0 (0.0%) | 1 (0.8%) | 1 (0.8%) | 0 (0.0%) | 2 (0.3%) |
| - Dermatitis exfoliative | 0 (0.0%) | 0 (0.0%) | 0 (0.0%) | 0 (0.0%) | 1 (0.8%) | 0 (0.0%) | 1 (0.1%) |
| - Eczema | 0 (0.0%) | 0 (0.0%) | 0 (0.0%) | 0 (0.0%) | 0 (0.0%) | 1 (0.8%) | 1 (0.1%) |
| - Fixed eruption | 1 (0.8%) | 0 (0.0%) | 0 (0.0%) | 0 (0.0%) | 0 (0.0%) | 0 (0.0%) | 1 (0.1%) |
| - Dry skin | 0 (0.0%) | 0 (0.0%) | 1 (0.8%) | 0 (0.0%) | 0 (0.0%) | 0 (0.0%) | 1 (0.1%) |
| **General disorders and administration site conditions** | **1 (0.8%)** | **1 (0.8%)** | **2 (1.6%)** | **2 (1.6%)** | **2 (1.7%)** | **4 (3.2%)** | **12 (1.6%)** |
| - Fatigue | 0 (0.0%) | 1 (0.8%) | 0 (0.0%) | 0 (0.0%) | 0 (0.0%) | 2 (1.6%) | 3 (0.4%) |
| - Chest pain | 0 (0.0%) | 0 (0.0%) | 1 (0.8%) | 0 (0.0%) | 0 (0.0%) | 1 (0.8%) | 2 (0.3%) |
| - Malaise | 0 (0.0%) | 1 (0.8%) | 0 (0.0%) | 0 (0.0%) | 0 (0.0%) | 1 (0.8%) | 2 (0.3%) |
| - Pyrexia | 0 (0.0%) | 0 (0.0%) | 0 (0.0%) | 0 (0.0%) | 1 (0.8%) | 1 (0.8%) | 2 (0.3%) |
| - Death | 0 (0.0%) | 0 (0.0%) | 0 (0.0%) | 1 (0.8%) | 0 (0.0%) | 0 (0.0%) | 1 (0.1%) |
| - Sudden death | 0 (0.0%) | 0 (0.0%) | 0 (0.0%) | 0 (0.0%) | 1 (0.8%) | 0 (0.0%) | 1 (0.1%) |
| - Generalised oedema | 1 (0.8%) | 0 (0.0%) | 0 (0.0%) | 0 (0.0%) | 0 (0.0%) | 0 (0.0%) | 1 (0.1%) |
| - Disease progression | 0 (0.0%) | 0 (0.0%) | 0 (0.0%) | 1 (0.8%) | 0 (0.0%) | 0 (0.0%) | 1 (0.1%) |
| - Asthenia | 0 (0.0%) | 0 (0.0%) | 0 (0.0%) | 0 (0.0%) | 0 (0.0%) | 1 (0.8%) | 1 (0.1%) |
| - Mucosal inflammation | 0 (0.0%) | 0 (0.0%) | 1 (0.8%) | 0 (0.0%) | 0 (0.0%) | 0 (0.0%) | 1 (0.1%) |
| **Injury, poisoning and procedural complications** | **0 (0.0%)** | **1 (0.8%)** | **2 (1.6%)** | **2 (1.6%)** | **2 (1.7%)** | **4 (3.2%)** | **11 (1.5%)** |
| - Alcohol poisoning | 0 (0.0%) | 1 (0.8%) | 0 (0.0%) | 0 (0.0%) | 0 (0.0%) | 2 (1.6%) | 3 (0.4%) |
| - Femur fracture | 0 (0.0%) | 0 (0.0%) | 0 (0.0%) | 0 (0.0%) | 1 (0.8%) | 0 (0.0%) | 1 (0.1%) |
| - Thermal burn | 0 (0.0%) | 0 (0.0%) | 1 (0.8%) | 0 (0.0%) | 0 (0.0%) | 0 (0.0%) | 1 (0.1%) |
| - Abortion induced incomplete | 0 (0.0%) | 0 (0.0%) | 1 (0.8%) | 0 (0.0%) | 0 (0.0%) | 0 (0.0%) | 1 (0.1%) |
| - Hip fracture | 0 (0.0%) | 0 (0.0%) | 0 (0.0%) | 0 (0.0%) | 1 (0.8%) | 0 (0.0%) | 1 (0.1%) |
| - Ankle fracture | 0 (0.0%) | 0 (0.0%) | 0 (0.0%) | 0 (0.0%) | 0 (0.0%) | 1 (0.8%) | 1 (0.1%) |
| - Stab wound | 0 (0.0%) | 0 (0.0%) | 0 (0.0%) | 0 (0.0%) | 0 (0.0%) | 1 (0.8%) | 1 (0.1%) |

| *Event*<br>**System Organ Class (SOC)**<br>-Preferred Term (PT) | **9BLMZ (n = 126)** | **9BCLLfxZ (n = 122)** | **9BDLLfxZ (n = 127)** | **9DCLLfxZ (n = 124)** | **9DCMZ (n = 120)** | **Control (n = 126)** | **Total* (N = 745)** |
|---|---|---|---|---|---|---|---|
| - Thoracic vertebral fracture | 0 (0.0%) | 0 (0.0%) | 0 (0.0%) | 1 (0.8%) | 0 (0.0%) | 0 (0.0%) | 1 (0.1%) |
| - Hand fracture | 0 (0.0%) | 0 (0.0%) | 0 (0.0%) | 1 (0.8%) | 0 (0.0%) | 0 (0.0%) | 1 (0.1%) |
| **Psychiatric disorders** | **0 (0.0%)** | **2 (1.6%)** | **1 (0.8%)** | **1 (0.8%)** | **3 (2.5%)** | **3 (2.4%)** | **10 (1.3%)** |
| - Suicide attempt | 0 (0.0%) | 1 (0.8%) | 0 (0.0%) | 0 (0.0%) | 3 (2.5%) | 1 (0.8%) | 5 (0.7%) |
| - Depression | 0 (0.0%) | 1 (0.8%) | 0 (0.0%) | 1 (0.8%) | 0 (0.0%) | 2 (1.6%) | 4 (0.5%) |
| - Anxiety | 0 (0.0%) | 0 (0.0%) | 0 (0.0%) | 1 (0.8%) | 0 (0.0%) | 1 (0.8%) | 2 (0.3%) |
| - Anxiety disorder | 0 (0.0%) | 0 (0.0%) | 1 (0.8%) | 0 (0.0%) | 0 (0.0%) | 0 (0.0%) | 1 (0.1%) |
| - Drug dependence | 0 (0.0%) | 1 (0.8%) | 0 (0.0%) | 0 (0.0%) | 0 (0.0%) | 0 (0.0%) | 1 (0.1%) |
| **Vascular disorders** | **1 (0.8%)** | **1 (0.8%)** | **1 (0.8%)** | **2 (1.6%)** | **0 (0.0%)** | **0 (0.0%)** | **5 (0.7%)** |
| - Hypotension | 0 (0.0%) | 0 (0.0%) | 1 (0.8%) | 1 (0.8%) | 0 (0.0%) | 0 (0.0%) | 2 (0.3%) |
| - Diabetic vascular disorder | 1 (0.8%) | 0 (0.0%) | 0 (0.0%) | 0 (0.0%) | 0 (0.0%) | 0 (0.0%) | 1 (0.1%) |
| - Hypertension | 0 (0.0%) | 1 (0.8%) | 0 (0.0%) | 0 (0.0%) | 0 (0.0%) | 0 (0.0%) | 1 (0.1%) |
| - Deep vein thrombosis | 0 (0.0%) | 0 (0.0%) | 0 (0.0%) | 1 (0.8%) | 0 (0.0%) | 0 (0.0%) | 1 (0.1%) |
| **Cardiac disorders** | **1 (0.8%)** | **2 (1.6%)** | **0 (0.0%)** | **2 (1.6%)** | **0 (0.0%)** | **0 (0.0%)** | **5 (0.7%)** |
| - Acute myocardial infarction | 0 (0.0%) | 1 (0.8%) | 0 (0.0%) | 2 (1.6%) | 0 (0.0%) | 0 (0.0%) | 3 (0.4%) |
| - Cardiac failure congestive | 1 (0.8%) | 1 (0.8%) | 0 (0.0%) | 0 (0.0%) | 0 (0.0%) | 0 (0.0%) | 2 (0.3%) |
| - Cardiac failure | 0 (0.0%) | 1 (0.8%) | 0 (0.0%) | 1 (0.8%) | 0 (0.0%) | 0 (0.0%) | 2 (0.3%) |
| **Renal and urinary disorders** | **1 (0.8%)** | **0 (0.0%)** | **1 (0.8%)** | **0 (0.0%)** | **0 (0.0%)** | **1 (0.8%)** | **3 (0.4%)** |
| - Proteinuria | 1 (0.8%) | 0 (0.0%) | 1 (0.8%) | 0 (0.0%) | 0 (0.0%) | 0 (0.0%) | 2 (0.3%) |
| - Chronic kidney disease | 0 (0.0%) | 0 (0.0%) | 0 (0.0%) | 0 (0.0%) | 0 (0.0%) | 1 (0.8%) | 1 (0.1%) |
| - Acute kidney injury | 1 (0.8%) | 0 (0.0%) | 0 (0.0%) | 0 (0.0%) | 0 (0.0%) | 0 (0.0%) | 1 (0.1%) |
| **Immune system disorders** | **0 (0.0%)** | **0 (0.0%)** | **2 (1.6%)** | **0 (0.0%)** | **0 (0.0%)** | **0 (0.0%)** | **2 (0.3%)** |
| - Hypersensitivity | 0 (0.0%) | 0 (0.0%) | 2 (1.6%) | 0 (0.0%) | 0 (0.0%) | 0 (0.0%) | 2 (0.3%) |
| **Pregnancy, puerperium and perinatal conditions** | **0 (0.0%)** | **0 (0.0%)** | **1 (0.8%)** | **0 (0.0%)** | **0 (0.0%)** | **1 (0.8%)** | **2 (0.3%)** |
| - Abortion threatened | 0 (0.0%) | 0 (0.0%) | 1 (0.8%) | 0 (0.0%) | 0 (0.0%) | 0 (0.0%) | 1 (0.1%) |
| - Abortion spontaneous | 0 (0.0%) | 0 (0.0%) | 0 (0.0%) | 0 (0.0%) | 0 (0.0%) | 1 (0.8%) | 1 (0.1%) |
| **Neoplasms benign, malignant and unspecified (incl cysts and polyps)** | **0 (0.0%)** | **0 (0.0%)** | **0 (0.0%)** | **0 (0.0%)** | **0 (0.0%)** | **1 (0.8%)** | **1 (0.1%)** |
| - Lung neoplasm malignant | 0 (0.0%) | 0 (0.0%) | 0 (0.0%) | 0 (0.0%) | 0 (0.0%) | 1 (0.8%) | 1 (0.1%) |

**Total by preferred term (PT) represents the sum of participants who experienced at least one event corresponding to that PT. If they experienced more than one event of the same PT, it is counted only once. If a participant experienced more than one unique PT within a system organ class (SOC), each unique PT counts toward the PT total, but only one event counts toward the SOC.*

*Table S21.* ***Number (%) of participants experiencing adverse events (AEs) of grade 3 or higher by SOC and PT by Week 104 (Safety Population)***

| *Event*<br>**System Organ Class (SOC)**<br>- Preferred Term (PT) | **9BLMZ (n = 126)** | **9BCLLfxZ (n = 122)** | **9BDLLfxZ (n = 127)** | **9DCLLfxZ (n = 124)** | **9DCMZ (n = 120)** | **Control (n = 126)** | **Total* (N = 745)** |
|---|---|---|---|---|---|---|---|
| **Investigations** | **43 (34.1%)** | **47 (38.5%)** | **43 (33.9%)** | **47 (37.9%)** | **51 (42.5%)** | **40 (31.7%)** | **271 (36.4%)** |
| - Creatinine renal clearance decreased | 21 (16.7%) | 24 (19.7%) | 30 (23.6%) | 37 (29.8%) | 37 (30.8%) | 23 (18.3%) | 172 (23.1%) |
| - Alanine aminotransferase increased | 14 (11.1%) | 10 (8.2%) | 5 (3.9%) | 8 (6.5%) | 7 (5.8%) | 3 (2.4%) | 47 (6.3%) |
| - Aspartate aminotransferase increased | 4 (3.2%) | 7 (5.7%) | 5 (3.9%) | 3 (2.4%) | 3 (2.5%) | 6 (4.8%) | 28 (3.8%) |
| - Blood magnesium increased | 6 (4.8%) | 6 (4.9%) | 4 (3.1%) | 1 (0.8%) | 5 (4.2%) | 3 (2.4%) | 25 (3.4%) |
| - Bilirubin conjugated increased | 2 (1.6%) | 3 (2.5%) | 3 (2.4%) | 1 (0.8%) | 1 (0.8%) | 4 (3.2%) | 14 (1.9%) |
| - Transaminases increased | 1 (0.8%) | 2 (1.6%) | 2 (1.6%) | 3 (2.4%) | 3 (2.5%) | 0 (0.0%) | 11 (1.5%) |
| - Lymphocyte count decreased | 4 (3.2%) | 1 (0.8%) | 2 (1.6%) | 0 (0.0%) | 2 (1.7%) | 1 (0.8%) | 10 (1.3%) |
| - Electrocardiogram QT prolonged | 0 (0.0%) | 4 (3.3%) | 0 (0.0%) | 0 (0.0%) | 5 (4.2%) | 0 (0.0%) | 9 (1.2%) |
| - Gamma-glutamyltransferase increased | 1 (0.8%) | 1 (0.8%) | 1 (0.8%) | 1 (0.8%) | 1 (0.8%) | 2 (1.6%) | 7 (0.9%) |
| - Glycosylated haemoglobin increased | 2 (1.6%) | 2 (1.6%) | 0 (0.0%) | 1 (0.8%) | 1 (0.8%) | 0 (0.0%) | 6 (0.8%) |
| - Neutrophil count decreased | 1 (0.8%) | 1 (0.8%) | 0 (0.0%) | 0 (0.0%) | 1 (0.8%) | 3 (2.4%) | 6 (0.8%) |
| - Blood bilirubin increased | 0 (0.0%) | 1 (0.8%) | 1 (0.8%) | 0 (0.0%) | 1 (0.8%) | 1 (0.8%) | 4 (0.5%) |
| - Hepatic enzyme increased | 2 (1.6%) | 1 (0.8%) | 1 (0.8%) | 0 (0.0%) | 0 (0.0%) | 0 (0.0%) | 4 (0.5%) |
| - Weight increased | 0 (0.0%) | 0 (0.0%) | 1 (0.8%) | 0 (0.0%) | 0 (0.0%) | 2 (1.6%) | 3 (0.4%) |
| - White blood cell count decreased | 0 (0.0%) | 0 (0.0%) | 0 (0.0%) | 0 (0.0%) | 1 (0.8%) | 2 (1.6%) | 3 (0.4%) |
| - Blood potassium increased | 0 (0.0%) | 0 (0.0%) | 0 (0.0%) | 0 (0.0%) | 2 (1.7%) | 1 (0.8%) | 3 (0.4%) |
| - Haemoglobin increased | 2 (1.6%) | 0 (0.0%) | 0 (0.0%) | 0 (0.0%) | 0 (0.0%) | 0 (0.0%) | 2 (0.3%) |
| - Blood uric acid increased | 0 (0.0%) | 1 (0.8%) | 0 (0.0%) | 0 (0.0%) | 0 (0.0%) | 0 (0.0%) | 1 (0.1%) |
| - Eosinophil count increased | 0 (0.0%) | 1 (0.8%) | 0 (0.0%) | 0 (0.0%) | 0 (0.0%) | 0 (0.0%) | 1 (0.1%) |
| - Eastern Cooperative Oncology Group performance status worsened | 0 (0.0%) | 0 (0.0%) | 0 (0.0%) | 1 (0.8%) | 0 (0.0%) | 0 (0.0%) | 1 (0.1%) |
| - Blood creatinine increased | 0 (0.0%) | 0 (0.0%) | 1 (0.8%) | 0 (0.0%) | 0 (0.0%) | 0 (0.0%) | 1 (0.1%) |
| - Blood creatinine decreased | 0 (0.0%) | 0 (0.0%) | 0 (0.0%) | 0 (0.0%) | 1 (0.8%) | 0 (0.0%) | 1 (0.1%) |
| - Psychiatric evaluation abnormal | 0 (0.0%) | 0 (0.0%) | 0 (0.0%) | 0 (0.0%) | 1 (0.8%) | 0 (0.0%) | 1 (0.1%) |
| - Blood urea increased | 0 (0.0%) | 0 (0.0%) | 1 (0.8%) | 0 (0.0%) | 0 (0.0%) | 0 (0.0%) | 1 (0.1%) |
| - Electrocardiogram QRS complex prolonged | 0 (0.0%) | 0 (0.0%) | 0 (0.0%) | 0 (0.0%) | 0 (0.0%) | 1 (0.8%) | 1 (0.1%) |
| - Blood calcium decreased | 0 (0.0%) | 1 (0.8%) | 0 (0.0%) | 0 (0.0%) | 0 (0.0%) | 0 (0.0%) | 1 (0.1%) |
| **Nervous system disorders** | **8 (6.3%)** | **11 (9.0%)** | **17 (13.4%)** | **13 (10.5%)** | **4 (3.3%)** | **15 (11.9%)** | **68 (9.1%)** |
| - Neuropathy peripheral | 3 (2.4%) | 5 (4.1%) | 7 (5.5%) | 2 (1.6%) | 1 (0.8%) | 6 (4.8%) | 24 (3.2%) |
| - Paraesthesia | 2 (1.6%) | 3 (2.5%) | 4 (3.1%) | 4 (3.2%) | 1 (0.8%) | 7 (5.6%) | 21 (2.8%) |
| - Areflexia | 0 (0.0%) | 0 (0.0%) | 1 (0.8%) | 2 (1.6%) | 3 (2.5%) | 1 (0.8%) | 7 (0.9%) |
| - Polyneuropathy | 1 (0.8%) | 0 (0.0%) | 2 (1.6%) | 1 (0.8%) | 2 (1.7%) | 0 (0.0%) | 6 (0.8%) |

| *Event*<br>**System Organ Class (SOC)**<br>- Preferred Term (PT) | **9BLMZ (n = 126)** | **9BCLLfxZ (n = 122)** | **9BDLLfxZ (n = 127)** | **9DCLLfxZ (n = 124)** | **9DCMZ (n = 120)** | **Control (n = 126)** | **Total* (N = 745)** |
|---|---|---|---|---|---|---|---|
| - Hyporeflexia | 2 (1.6%) | 0 (0.0%) | 1 (0.8%) | 0 (0.0%) | 0 (0.0%) | 0 (0.0%) | 3 (0.4%) |
| - Transient ischaemic attack | 0 (0.0%) | 1 (0.8%) | 0 (0.0%) | 1 (0.8%) | 0 (0.0%) | 0 (0.0%) | 2 (0.3%) |
| - Headache | 0 (0.0%) | 1 (0.8%) | 1 (0.8%) | 0 (0.0%) | 0 (0.0%) | 0 (0.0%) | 2 (0.3%) |
| - Optic neuritis | 0 (0.0%) | 0 (0.0%) | 0 (0.0%) | 1 (0.8%) | 0 (0.0%) | 1 (0.8%) | 2 (0.3%) |
| - Facial paralysis | 0 (0.0%) | 0 (0.0%) | 1 (0.8%) | 0 (0.0%) | 0 (0.0%) | 0 (0.0%) | 1 (0.1%) |
| - Sciatica | 0 (0.0%) | 0 (0.0%) | 1 (0.8%) | 0 (0.0%) | 0 (0.0%) | 0 (0.0%) | 1 (0.1%) |
| - Syncope | 0 (0.0%) | 0 (0.0%) | 0 (0.0%) | 1 (0.8%) | 0 (0.0%) | 0 (0.0%) | 1 (0.1%) |
| - Brain injury | 0 (0.0%) | 0 (0.0%) | 0 (0.0%) | 1 (0.8%) | 0 (0.0%) | 0 (0.0%) | 1 (0.1%) |
| - Idiopathic intracranial hypertension | 0 (0.0%) | 0 (0.0%) | 0 (0.0%) | 0 (0.0%) | 0 (0.0%) | 1 (0.8%) | 1 (0.1%) |
| - Epilepsy | 0 (0.0%) | 1 (0.8%) | 0 (0.0%) | 0 (0.0%) | 0 (0.0%) | 0 (0.0%) | 1 (0.1%) |
| **Metabolism and nutrition disorders** | **11 (8.7%)** | **7 (5.7%)** | **11 (8.7%)** | **10 (8.1%)** | **10 (8.3%)** | **13 (10.3%)** | **62 (8.3%)** |
| - Hypermagnesaemia | 7 (5.6%) | 3 (2.5%) | 5 (3.9%) | 3 (2.4%) | 3 (2.5%) | 8 (6.3%) | 29 (3.9%) |
| - Hyperglycaemia | 2 (1.6%) | 2 (1.6%) | 0 (0.0%) | 3 (2.4%) | 2 (1.7%) | 1 (0.8%) | 10 (1.3%) |
| - Hyperkalaemia | 2 (1.6%) | 0 (0.0%) | 0 (0.0%) | 2 (1.6%) | 1 (0.8%) | 1 (0.8%) | 6 (0.8%) |
| - Hypocalcaemia | 0 (0.0%) | 1 (0.8%) | 0 (0.0%) | 2 (1.6%) | 0 (0.0%) | 1 (0.8%) | 4 (0.5%) |
| - Diabetes mellitus inadequate control | 0 (0.0%) | 0 (0.0%) | 1 (0.8%) | 0 (0.0%) | 2 (1.7%) | 0 (0.0%) | 3 (0.4%) |
| - Hypoalbuminaemia | 1 (0.8%) | 1 (0.8%) | 0 (0.0%) | 0 (0.0%) | 1 (0.8%) | 0 (0.0%) | 3 (0.4%) |
| - Hyperuricaemia | 0 (0.0%) | 0 (0.0%) | 1 (0.8%) | 0 (0.0%) | 1 (0.8%) | 0 (0.0%) | 2 (0.3%) |
| - Hypomagnesaemia | 0 (0.0%) | 0 (0.0%) | 1 (0.8%) | 0 (0.0%) | 0 (0.0%) | 1 (0.8%) | 2 (0.3%) |
| - Obesity | 0 (0.0%) | 0 (0.0%) | 0 (0.0%) | 0 (0.0%) | 1 (0.8%) | 1 (0.8%) | 2 (0.3%) |
| - Hyponatraemia | 0 (0.0%) | 0 (0.0%) | 0 (0.0%) | 1 (0.8%) | 0 (0.0%) | 0 (0.0%) | 1 (0.1%) |
| - Hypokalaemia | 0 (0.0%) | 0 (0.0%) | 1 (0.8%) | 0 (0.0%) | 0 (0.0%) | 0 (0.0%) | 1 (0.1%) |
| - Decreased appetite | 0 (0.0%) | 0 (0.0%) | 1 (0.8%) | 0 (0.0%) | 0 (0.0%) | 0 (0.0%) | 1 (0.1%) |
| - Hypercalcaemia | 0 (0.0%) | 0 (0.0%) | 1 (0.8%) | 0 (0.0%) | 0 (0.0%) | 0 (0.0%) | 1 (0.1%) |
| - Hypertriglyceridaemia | 0 (0.0%) | 0 (0.0%) | 0 (0.0%) | 0 (0.0%) | 1 (0.8%) | 0 (0.0%) | 1 (0.1%) |
| **Eye disorders** | **5 (4.0%)** | **13 (10.7%)** | **10 (7.9%)** | **10 (8.1%)** | **8 (6.7%)** | **12 (9.5%)** | **58 (7.8%)** |
| - Visual acuity reduced | 4 (3.2%) | 7 (5.7%) | 7 (5.5%) | 5 (4.0%) | 5 (4.2%) | 7 (5.6%) | 35 (4.7%) |
| - Visual impairment | 0 (0.0%) | 2 (1.6%) | 3 (2.4%) | 2 (1.6%) | 2 (1.7%) | 2 (1.6%) | 11 (1.5%) |
| - Cataract | 1 (0.8%) | 1 (0.8%) | 0 (0.0%) | 1 (0.8%) | 0 (0.0%) | 2 (1.6%) | 5 (0.7%) |
| - Refraction disorder | 0 (0.0%) | 1 (0.8%) | 0 (0.0%) | 1 (0.8%) | 0 (0.0%) | 0 (0.0%) | 2 (0.3%) |
| - Retinopathy | 0 (0.0%) | 1 (0.8%) | 0 (0.0%) | 0 (0.0%) | 1 (0.8%) | 0 (0.0%) | 2 (0.3%) |
| - Hypermetropia | 0 (0.0%) | 1 (0.8%) | 0 (0.0%) | 0 (0.0%) | 0 (0.0%) | 0 (0.0%) | 1 (0.1%) |
| - Myopia | 0 (0.0%) | 0 (0.0%) | 0 (0.0%) | 0 (0.0%) | 1 (0.8%) | 0 (0.0%) | 1 (0.1%) |
| - Cataract subcapsular | 0 (0.0%) | 0 (0.0%) | 0 (0.0%) | 0 (0.0%) | 0 (0.0%) | 1 (0.8%) | 1 (0.1%) |

| *Event*<br>**System Organ Class (SOC)**<br>- Preferred Term (PT) | **9BLMZ (n = 126)** | **9BCLLfxZ (n = 122)** | **9BDLLfxZ (n = 127)** | **9DCLLfxZ (n = 124)** | **9DCMZ (n = 120)** | **Control (n = 126)** | **Total* (N = 745)** |
|---|---|---|---|---|---|---|---|
| - Vision blurred | 0 (0.0%) | 0 (0.0%) | 0 (0.0%) | 0 (0.0%) | 0 (0.0%) | 1 (0.8%) | 1 (0.1%) |
| - Optic neuropathy | 0 (0.0%) | 1 (0.8%) | 0 (0.0%) | 0 (0.0%) | 0 (0.0%) | 0 (0.0%) | 1 (0.1%) |
| - Visual field defect | 0 (0.0%) | 0 (0.0%) | 0 (0.0%) | 1 (0.8%) | 0 (0.0%) | 0 (0.0%) | 1 (0.1%) |
| - Presbyopia | 0 (0.0%) | 1 (0.8%) | 0 (0.0%) | 0 (0.0%) | 0 (0.0%) | 0 (0.0%) | 1 (0.1%) |
| - Eye haemorrhage | 0 (0.0%) | 1 (0.8%) | 0 (0.0%) | 0 (0.0%) | 0 (0.0%) | 0 (0.0%) | 1 (0.1%) |
| **Ear and labyrinth disorders** | **10 (7.9%)** | **14 (11.5%)** | **6 (4.7%)** | **4 (3.2%)** | **11 (9.2%)** | **9 (7.1%)** | **54 (7.2%)** |
| - Hypoacusis | 8 (6.3%) | 12 (9.8%) | 4 (3.1%) | 4 (3.2%) | 9 (7.5%) | 8 (6.3%) | 45 (6.0%) |
| - Deafness | 2 (1.6%) | 2 (1.6%) | 1 (0.8%) | 0 (0.0%) | 2 (1.7%) | 1 (0.8%) | 8 (1.1%) |
| - Deafness unilateral | 0 (0.0%) | 0 (0.0%) | 1 (0.8%) | 0 (0.0%) | 0 (0.0%) | 0 (0.0%) | 1 (0.1%) |
| - Tinnitus | 0 (0.0%) | 1 (0.8%) | 0 (0.0%) | 0 (0.0%) | 0 (0.0%) | 0 (0.0%) | 1 (0.1%) |
| **Blood and lymphatic system disorders** | **6 (4.8%)** | **8 (6.6%)** | **9 (7.1%)** | **13 (10.5%)** | **5 (4.2%)** | **10 (7.9%)** | **51 (6.8%)** |
| - Anaemia | 4 (3.2%) | 6 (4.9%) | 9 (7.1%) | 9 (7.3%) | 3 (2.5%) | 7 (5.6%) | 38 (5.1%) |
| - Lymphopenia | 1 (0.8%) | 0 (0.0%) | 0 (0.0%) | 4 (3.2%) | 1 (0.8%) | 3 (2.4%) | 9 (1.2%) |
| - Neutropenia | 0 (0.0%) | 1 (0.8%) | 0 (0.0%) | 0 (0.0%) | 1 (0.8%) | 1 (0.8%) | 3 (0.4%) |
| - Leukopenia | 0 (0.0%) | 1 (0.8%) | 0 (0.0%) | 0 (0.0%) | 0 (0.0%) | 1 (0.8%) | 2 (0.3%) |
| - Bicytopenia | 0 (0.0%) | 0 (0.0%) | 0 (0.0%) | 0 (0.0%) | 1 (0.8%) | 1 (0.8%) | 2 (0.3%) |
| - Thrombocytopenia | 1 (0.8%) | 0 (0.0%) | 0 (0.0%) | 0 (0.0%) | 0 (0.0%) | 1 (0.8%) | 2 (0.3%) |
| - Pancytopenia | 0 (0.0%) | 0 (0.0%) | 0 (0.0%) | 1 (0.8%) | 0 (0.0%) | 0 (0.0%) | 1 (0.1%) |
| - Leukocytosis | 0 (0.0%) | 0 (0.0%) | 0 (0.0%) | 0 (0.0%) | 0 (0.0%) | 1 (0.8%) | 1 (0.1%) |
| - Bone marrow failure | 0 (0.0%) | 1 (0.8%) | 0 (0.0%) | 0 (0.0%) | 0 (0.0%) | 0 (0.0%) | 1 (0.1%) |
| - Myelosuppression | 0 (0.0%) | 0 (0.0%) | 0 (0.0%) | 1 (0.8%) | 0 (0.0%) | 0 (0.0%) | 1 (0.1%) |
| **Infections and infestations** | **13 (10.3%)** | **6 (4.9%)** | **6 (4.7%)** | **6 (4.8%)** | **7 (5.8%)** | **4 (3.2%)** | **42 (5.6%)** |
| - Pneumonia | 4 (3.2%) | 1 (0.8%) | 3 (2.4%) | 0 (0.0%) | 1 (0.8%) | 2 (1.6%) | 11 (1.5%) |
| - COVID-19 | 2 (1.6%) | 0 (0.0%) | 0 (0.0%) | 0 (0.0%) | 2 (1.7%) | 1 (0.8%) | 5 (0.7%) |
| - COVID-19 pneumonia | 0 (0.0%) | 4 (3.3%) | 0 (0.0%) | 0 (0.0%) | 1 (0.8%) | 0 (0.0%) | 5 (0.7%) |
| - Sepsis | 0 (0.0%) | 1 (0.8%) | 0 (0.0%) | 1 (0.8%) | 1 (0.8%) | 0 (0.0%) | 3 (0.4%) |
| - Hepatitis A | 1 (0.8%) | 0 (0.0%) | 0 (0.0%) | 2 (1.6%) | 0 (0.0%) | 0 (0.0%) | 3 (0.4%) |
| - Appendicitis | 1 (0.8%) | 0 (0.0%) | 0 (0.0%) | 0 (0.0%) | 0 (0.0%) | 1 (0.8%) | 2 (0.3%) |
| - Pneumocystis jirovecii pneumonia | 1 (0.8%) | 0 (0.0%) | 0 (0.0%) | 1 (0.8%) | 0 (0.0%) | 0 (0.0%) | 2 (0.3%) |
| - Post procedural sepsis | 0 (0.0%) | 0 (0.0%) | 0 (0.0%) | 1 (0.8%) | 0 (0.0%) | 0 (0.0%) | 1 (0.1%) |
| - Pyelonephritis | 0 (0.0%) | 0 (0.0%) | 0 (0.0%) | 0 (0.0%) | 1 (0.8%) | 0 (0.0%) | 1 (0.1%) |
| - Diabetic foot infection | 1 (0.8%) | 0 (0.0%) | 0 (0.0%) | 0 (0.0%) | 0 (0.0%) | 0 (0.0%) | 1 (0.1%) |
| - Osteomyelitis | 1 (0.8%) | 0 (0.0%) | 0 (0.0%) | 0 (0.0%) | 0 (0.0%) | 0 (0.0%) | 1 (0.1%) |
| - Infective glossitis | 0 (0.0%) | 0 (0.0%) | 0 (0.0%) | 1 (0.8%) | 0 (0.0%) | 0 (0.0%) | 1 (0.1%) |

| *Event*<br>**System Organ Class (SOC)**<br>- Preferred Term (PT) | **9BLMZ (n = 126)** | **9BCLLfxZ (n = 122)** | **9BDLLfxZ (n = 127)** | **9DCLLfxZ (n = 124)** | **9DCMZ (n = 120)** | **Control (n = 126)** | **Total* (N = 745)** |
|---|---|---|---|---|---|---|---|
| - Tonsillitis | 0 (0.0%) | 0 (0.0%) | 0 (0.0%) | 0 (0.0%) | 0 (0.0%) | 1 (0.8%) | 1 (0.1%) |
| - Herpes zoster | 0 (0.0%) | 0 (0.0%) | 0 (0.0%) | 0 (0.0%) | 1 (0.8%) | 0 (0.0%) | 1 (0.1%) |
| - Septic shock | 1 (0.8%) | 0 (0.0%) | 0 (0.0%) | 0 (0.0%) | 0 (0.0%) | 0 (0.0%) | 1 (0.1%) |
| - Pulmonary tuberculoma | 0 (0.0%) | 0 (0.0%) | 1 (0.8%) | 0 (0.0%) | 0 (0.0%) | 0 (0.0%) | 1 (0.1%) |
| - HIV infection | 0 (0.0%) | 0 (0.0%) | 0 (0.0%) | 0 (0.0%) | 0 (0.0%) | 1 (0.8%) | 1 (0.1%) |
| - Dengue fever | 1 (0.8%) | 0 (0.0%) | 0 (0.0%) | 0 (0.0%) | 0 (0.0%) | 0 (0.0%) | 1 (0.1%) |
| - Pneumonia bacterial | 0 (0.0%) | 0 (0.0%) | 1 (0.8%) | 0 (0.0%) | 0 (0.0%) | 0 (0.0%) | 1 (0.1%) |
| - Cellulitis | 0 (0.0%) | 0 (0.0%) | 1 (0.8%) | 0 (0.0%) | 0 (0.0%) | 0 (0.0%) | 1 (0.1%) |
| - Pneumonia staphylococcal | 0 (0.0%) | 0 (0.0%) | 1 (0.8%) | 0 (0.0%) | 0 (0.0%) | 0 (0.0%) | 1 (0.1%) |
| - Respiratory tract infection | 0 (0.0%) | 0 (0.0%) | 1 (0.8%) | 0 (0.0%) | 0 (0.0%) | 0 (0.0%) | 1 (0.1%) |
| - Tuberculosis | 1 (0.8%) | 0 (0.0%) | 0 (0.0%) | 0 (0.0%) | 0 (0.0%) | 0 (0.0%) | 1 (0.1%) |
| - Skin infection | 1 (0.8%) | 0 (0.0%) | 0 (0.0%) | 0 (0.0%) | 0 (0.0%) | 0 (0.0%) | 1 (0.1%) |
| - Abscess soft tissue | 0 (0.0%) | 0 (0.0%) | 0 (0.0%) | 0 (0.0%) | 1 (0.8%) | 0 (0.0%) | 1 (0.1%) |
| - Wound infection | 1 (0.8%) | 0 (0.0%) | 0 (0.0%) | 0 (0.0%) | 0 (0.0%) | 0 (0.0%) | 1 (0.1%) |
| - Gastroenteritis | 0 (0.0%) | 0 (0.0%) | 0 (0.0%) | 1 (0.8%) | 0 (0.0%) | 0 (0.0%) | 1 (0.1%) |
| **Hepatobiliary disorders** | **6 (4.8%)** | **5 (4.1%)** | **2 (1.6%)** | **7 (5.6%)** | **5 (4.2%)** | **3 (2.4%)** | **28 (3.8%)** |
| - Hepatotoxicity | 5 (4.0%) | 1 (0.8%) | 1 (0.8%) | 3 (2.4%) | 1 (0.8%) | 1 (0.8%) | 12 (1.6%) |
| - Drug-induced liver injury | 1 (0.8%) | 3 (2.5%) | 1 (0.8%) | 3 (2.4%) | 0 (0.0%) | 0 (0.0%) | 8 (1.1%) |
| - Hepatitis | 0 (0.0%) | 1 (0.8%) | 0 (0.0%) | 0 (0.0%) | 2 (1.7%) | 2 (1.6%) | 5 (0.7%) |
| - Hyperbilirubinaemia | 0 (0.0%) | 0 (0.0%) | 0 (0.0%) | 1 (0.8%) | 1 (0.8%) | 0 (0.0%) | 2 (0.3%) |
| - Acute on chronic liver failure | 0 (0.0%) | 0 (0.0%) | 0 (0.0%) | 0 (0.0%) | 1 (0.8%) | 0 (0.0%) | 1 (0.1%) |
| **Musculoskeletal and connective tissue disorders** | **1 (0.8%)** | **6 (4.9%)** | **4 (3.1%)** | **4 (3.2%)** | **2 (1.7%)** | **5 (4.0%)** | **22 (3.0%)** |
| - Arthralgia | 0 (0.0%) | 4 (3.3%) | 3 (2.4%) | 2 (1.6%) | 1 (0.8%) | 5 (4.0%) | 15 (2.0%) |
| - Myalgia | 0 (0.0%) | 2 (1.6%) | 1 (0.8%) | 0 (0.0%) | 0 (0.0%) | 1 (0.8%) | 4 (0.5%) |
| - Back pain | 1 (0.8%) | 0 (0.0%) | 0 (0.0%) | 1 (0.8%) | 0 (0.0%) | 1 (0.8%) | 3 (0.4%) |
| - Rotator cuff syndrome | 0 (0.0%) | 1 (0.8%) | 0 (0.0%) | 1 (0.8%) | 0 (0.0%) | 0 (0.0%) | 2 (0.3%) |
| - Osteoarthritis | 0 (0.0%) | 0 (0.0%) | 0 (0.0%) | 0 (0.0%) | 1 (0.8%) | 0 (0.0%) | 1 (0.1%) |
| - Arthropathy | 0 (0.0%) | 1 (0.8%) | 0 (0.0%) | 0 (0.0%) | 0 (0.0%) | 0 (0.0%) | 1 (0.1%) |
| **Gastrointestinal disorders** | **1 (0.8%)** | **2 (1.6%)** | **3 (2.4%)** | **5 (4.0%)** | **3 (2.5%)** | **5 (4.0%)** | **19 (2.6%)** |
| - Vomiting | 0 (0.0%) | 1 (0.8%) | 0 (0.0%) | 3 (2.4%) | 0 (0.0%) | 3 (2.4%) | 7 (0.9%) |
| - Nausea | 0 (0.0%) | 1 (0.8%) | 2 (1.6%) | 1 (0.8%) | 1 (0.8%) | 0 (0.0%) | 5 (0.7%) |
| - Mallory-Weiss syndrome | 0 (0.0%) | 0 (0.0%) | 0 (0.0%) | 0 (0.0%) | 0 (0.0%) | 1 (0.8%) | 1 (0.1%) |
| - Dyspepsia | 0 (0.0%) | 0 (0.0%) | 1 (0.8%) | 0 (0.0%) | 0 (0.0%) | 0 (0.0%) | 1 (0.1%) |
| - Upper gastrointestinal haemorrhage | 0 (0.0%) | 0 (0.0%) | 0 (0.0%) | 0 (0.0%) | 1 (0.8%) | 0 (0.0%) | 1 (0.1%) |

| *Event*<br>**System Organ Class (SOC)**<br>- Preferred Term (PT) | **9BLMZ (n = 126)** | **9BCLLfxZ (n = 122)** | **9BDLLfxZ (n = 127)** | **9DCLLfxZ (n = 124)** | **9DCMZ (n = 120)** | **Control (n = 126)** | **Total* (N = 745)** |
|---|---|---|---|---|---|---|---|
| - Abdominal hernia | 1 (0.8%) | 0 (0.0%) | 0 (0.0%) | 0 (0.0%) | 0 (0.0%) | 0 (0.0%) | 1 (0.1%) |
| - Abdominal pain | 0 (0.0%) | 0 (0.0%) | 0 (0.0%) | 1 (0.8%) | 0 (0.0%) | 0 (0.0%) | 1 (0.1%) |
| - Pancreatitis | 0 (0.0%) | 0 (0.0%) | 0 (0.0%) | 0 (0.0%) | 0 (0.0%) | 1 (0.8%) | 1 (0.1%) |
| - Abdominal pain upper | 0 (0.0%) | 0 (0.0%) | 0 (0.0%) | 0 (0.0%) | 1 (0.8%) | 0 (0.0%) | 1 (0.1%) |
| - Pancreatitis acute | 0 (0.0%) | 0 (0.0%) | 0 (0.0%) | 0 (0.0%) | 0 (0.0%) | 1 (0.8%) | 1 (0.1%) |
| - Acute abdomen | 0 (0.0%) | 0 (0.0%) | 1 (0.8%) | 0 (0.0%) | 0 (0.0%) | 0 (0.0%) | 1 (0.1%) |
| - Small intestinal obstruction | 0 (0.0%) | 0 (0.0%) | 0 (0.0%) | 1 (0.8%) | 0 (0.0%) | 0 (0.0%) | 1 (0.1%) |
| - Gastritis | 0 (0.0%) | 0 (0.0%) | 0 (0.0%) | 0 (0.0%) | 1 (0.8%) | 0 (0.0%) | 1 (0.1%) |
| **Psychiatric disorders** | **1 (0.8%)** | **4 (3.3%)** | **2 (1.6%)** | **2 (1.6%)** | **4 (3.3%)** | **5 (4.0%)** | **18 (2.4%)** |
| - Depression | 0 (0.0%) | 3 (2.5%) | 0 (0.0%) | 1 (0.8%) | 1 (0.8%) | 3 (2.4%) | 8 (1.1%) |
| - Anxiety | 0 (0.0%) | 1 (0.8%) | 1 (0.8%) | 2 (1.6%) | 0 (0.0%) | 2 (1.6%) | 6 (0.8%) |
| - Suicide attempt | 0 (0.0%) | 1 (0.8%) | 0 (0.0%) | 0 (0.0%) | 3 (2.5%) | 1 (0.8%) | 5 (0.7%) |
| - Drug dependence | 0 (0.0%) | 1 (0.8%) | 0 (0.0%) | 0 (0.0%) | 0 (0.0%) | 0 (0.0%) | 1 (0.1%) |
| - Brief psychotic disorder without marked stressors | 1 (0.8%) | 0 (0.0%) | 0 (0.0%) | 0 (0.0%) | 0 (0.0%) | 0 (0.0%) | 1 (0.1%) |
| - Anxiety disorder | 0 (0.0%) | 0 (0.0%) | 1 (0.8%) | 0 (0.0%) | 0 (0.0%) | 0 (0.0%) | 1 (0.1%) |
| **Respiratory, thoracic and mediastinal disorders** | **0 (0.0%)** | **3 (2.5%)** | **4 (3.1%)** | **2 (1.6%)** | **3 (2.5%)** | **4 (3.2%)** | **16 (2.1%)** |
| - Pneumothorax | 0 (0.0%) | 0 (0.0%) | 0 (0.0%) | 1 (0.8%) | 1 (0.8%) | 1 (0.8%) | 3 (0.4%) |
| - Haemoptysis | 0 (0.0%) | 0 (0.0%) | 0 (0.0%) | 0 (0.0%) | 1 (0.8%) | 2 (1.6%) | 3 (0.4%) |
| - Dyspnoea | 0 (0.0%) | 1 (0.8%) | 1 (0.8%) | 1 (0.8%) | 0 (0.0%) | 0 (0.0%) | 3 (0.4%) |
| - Cough | 0 (0.0%) | 0 (0.0%) | 1 (0.8%) | 0 (0.0%) | 2 (1.7%) | 0 (0.0%) | 3 (0.4%) |
| - Respiratory failure | 0 (0.0%) | 0 (0.0%) | 1 (0.8%) | 0 (0.0%) | 0 (0.0%) | 1 (0.8%) | 2 (0.3%) |
| - Pulmonary cavitation | 0 (0.0%) | 0 (0.0%) | 0 (0.0%) | 0 (0.0%) | 0 (0.0%) | 1 (0.8%) | 1 (0.1%) |
| - Acute pulmonary oedema | 0 (0.0%) | 1 (0.8%) | 0 (0.0%) | 0 (0.0%) | 0 (0.0%) | 0 (0.0%) | 1 (0.1%) |
| - Pulmonary haemorrhage | 0 (0.0%) | 1 (0.8%) | 0 (0.0%) | 0 (0.0%) | 0 (0.0%) | 0 (0.0%) | 1 (0.1%) |
| - Acute respiratory failure | 0 (0.0%) | 0 (0.0%) | 1 (0.8%) | 0 (0.0%) | 0 (0.0%) | 0 (0.0%) | 1 (0.1%) |
| **Skin and subcutaneous tissue disorders** | **1 (0.8%)** | **2 (1.6%)** | **2 (1.6%)** | **3 (2.4%)** | **3 (2.5%)** | **1 (0.8%)** | **12 (1.6%)** |
| - Rash | 0 (0.0%) | 1 (0.8%) | 1 (0.8%) | 2 (1.6%) | 0 (0.0%) | 0 (0.0%) | 4 (0.5%) |
| - Pruritus | 0 (0.0%) | 1 (0.8%) | 0 (0.0%) | 0 (0.0%) | 2 (1.7%) | 0 (0.0%) | 3 (0.4%) |
| - Drug reaction with eosinophilia and systemic symptoms | 0 (0.0%) | 0 (0.0%) | 0 (0.0%) | 1 (0.8%) | 1 (0.8%) | 0 (0.0%) | 2 (0.3%) |
| - Eczema | 0 (0.0%) | 0 (0.0%) | 0 (0.0%) | 0 (0.0%) | 0 (0.0%) | 1 (0.8%) | 1 (0.1%) |
| - Dry skin | 0 (0.0%) | 0 (0.0%) | 1 (0.8%) | 0 (0.0%) | 0 (0.0%) | 0 (0.0%) | 1 (0.1%) |
| - Fixed eruption | 1 (0.8%) | 0 (0.0%) | 0 (0.0%) | 0 (0.0%) | 0 (0.0%) | 0 (0.0%) | 1 (0.1%) |
| - Dermatitis exfoliative | 0 (0.0%) | 0 (0.0%) | 0 (0.0%) | 0 (0.0%) | 1 (0.8%) | 0 (0.0%) | 1 (0.1%) |

| *Event*<br>**System Organ Class (SOC)**<br>- Preferred Term (PT) | **9BLMZ (n = 126)** | **9BCLLfxZ (n = 122)** | **9BDLLfxZ (n = 127)** | **9DCLLfxZ (n = 124)** | **9DCMZ (n = 120)** | **Control (n = 126)** | **Total* (N = 745)** |
|---|---|---|---|---|---|---|---|
| **General disorders and administration site conditions** | **1 (0.8%)** | **1 (0.8%)** | **2 (1.6%)** | **2 (1.6%)** | **2 (1.7%)** | **4 (3.2%)** | **12 (1.6%)** |
| - Fatigue | 0 (0.0%) | 1 (0.8%) | 0 (0.0%) | 0 (0.0%) | 0 (0.0%) | 2 (1.6%) | 3 (0.4%) |
| - Malaise | 0 (0.0%) | 1 (0.8%) | 0 (0.0%) | 0 (0.0%) | 0 (0.0%) | 1 (0.8%) | 2 (0.3%) |
| - Chest pain | 0 (0.0%) | 0 (0.0%) | 1 (0.8%) | 0 (0.0%) | 0 (0.0%) | 1 (0.8%) | 2 (0.3%) |
| - Pyrexia | 0 (0.0%) | 0 (0.0%) | 0 (0.0%) | 0 (0.0%) | 1 (0.8%) | 1 (0.8%) | 2 (0.3%) |
| - Generalised oedema | 1 (0.8%) | 0 (0.0%) | 0 (0.0%) | 0 (0.0%) | 0 (0.0%) | 0 (0.0%) | 1 (0.1%) |
| - Asthenia | 0 (0.0%) | 0 (0.0%) | 0 (0.0%) | 0 (0.0%) | 0 (0.0%) | 1 (0.8%) | 1 (0.1%) |
| - Mucosal inflammation | 0 (0.0%) | 0 (0.0%) | 1 (0.8%) | 0 (0.0%) | 0 (0.0%) | 0 (0.0%) | 1 (0.1%) |
| - Death | 0 (0.0%) | 0 (0.0%) | 0 (0.0%) | 1 (0.8%) | 0 (0.0%) | 0 (0.0%) | 1 (0.1%) |
| - Sudden death | 0 (0.0%) | 0 (0.0%) | 0 (0.0%) | 0 (0.0%) | 1 (0.8%) | 0 (0.0%) | 1 (0.1%) |
| - Disease progression | 0 (0.0%) | 0 (0.0%) | 0 (0.0%) | 1 (0.8%) | 0 (0.0%) | 0 (0.0%) | 1 (0.1%) |
| **Injury, poisoning and procedural complications** | **0 (0.0%)** | **1 (0.8%)** | **2 (1.6%)** | **2 (1.6%)** | **2 (1.7%)** | **4 (3.2%)** | **11 (1.5%)** |
| - Alcohol poisoning | 0 (0.0%) | 1 (0.8%) | 0 (0.0%) | 0 (0.0%) | 0 (0.0%) | 2 (1.6%) | 3 (0.4%) |
| - Thermal burn | 0 (0.0%) | 0 (0.0%) | 1 (0.8%) | 0 (0.0%) | 0 (0.0%) | 0 (0.0%) | 1 (0.1%) |
| - Hand fracture | 0 (0.0%) | 0 (0.0%) | 0 (0.0%) | 1 (0.8%) | 0 (0.0%) | 0 (0.0%) | 1 (0.1%) |
| - Thoracic vertebral fracture | 0 (0.0%) | 0 (0.0%) | 0 (0.0%) | 1 (0.8%) | 0 (0.0%) | 0 (0.0%) | 1 (0.1%) |
| - Femur fracture | 0 (0.0%) | 0 (0.0%) | 0 (0.0%) | 0 (0.0%) | 1 (0.8%) | 0 (0.0%) | 1 (0.1%) |
| - Stab wound | 0 (0.0%) | 0 (0.0%) | 0 (0.0%) | 0 (0.0%) | 0 (0.0%) | 1 (0.8%) | 1 (0.1%) |
| - Hip fracture | 0 (0.0%) | 0 (0.0%) | 0 (0.0%) | 0 (0.0%) | 1 (0.8%) | 0 (0.0%) | 1 (0.1%) |
| - Abortion induced incomplete | 0 (0.0%) | 0 (0.0%) | 1 (0.8%) | 0 (0.0%) | 0 (0.0%) | 0 (0.0%) | 1 (0.1%) |
| - Ankle fracture | 0 (0.0%) | 0 (0.0%) | 0 (0.0%) | 0 (0.0%) | 0 (0.0%) | 1 (0.8%) | 1 (0.1%) |
| **Cardiac disorders** | **1 (0.8%)** | **2 (1.6%)** | **0 (0.0%)** | **2 (1.6%)** | **0 (0.0%)** | **1 (0.8%)** | **6 (0.8%)** |
| - Acute myocardial infarction | 0 (0.0%) | 1 (0.8%) | 0 (0.0%) | 2 (1.6%) | 0 (0.0%) | 0 (0.0%) | 3 (0.4%) |
| - Cardiac failure congestive | 1 (0.8%) | 1 (0.8%) | 0 (0.0%) | 0 (0.0%) | 0 (0.0%) | 0 (0.0%) | 2 (0.3%) |
| - Cardiac failure | 0 (0.0%) | 1 (0.8%) | 0 (0.0%) | 1 (0.8%) | 0 (0.0%) | 0 (0.0%) | 2 (0.3%) |
| - Cardiomyopathy alcoholic | 0 (0.0%) | 0 (0.0%) | 0 (0.0%) | 0 (0.0%) | 0 (0.0%) | 1 (0.8%) | 1 (0.1%) |
| **Vascular disorders** | **1 (0.8%)** | **1 (0.8%)** | **1 (0.8%)** | **2 (1.6%)** | **0 (0.0%)** | **0 (0.0%)** | **5 (0.7%)** |
| - Hypotension | 0 (0.0%) | 0 (0.0%) | 1 (0.8%) | 1 (0.8%) | 0 (0.0%) | 0 (0.0%) | 2 (0.3%) |
| - Deep vein thrombosis | 0 (0.0%) | 0 (0.0%) | 0 (0.0%) | 1 (0.8%) | 0 (0.0%) | 0 (0.0%) | 1 (0.1%) |
| - Diabetic vascular disorder | 1 (0.8%) | 0 (0.0%) | 0 (0.0%) | 0 (0.0%) | 0 (0.0%) | 0 (0.0%) | 1 (0.1%) |
| - Hypertension | 0 (0.0%) | 1 (0.8%) | 0 (0.0%) | 0 (0.0%) | 0 (0.0%) | 0 (0.0%) | 1 (0.1%) |
| **Renal and urinary disorders** | **1 (0.8%)** | **0 (0.0%)** | **1 (0.8%)** | **0 (0.0%)** | **0 (0.0%)** | **1 (0.8%)** | **3 (0.4%)** |
| - Proteinuria | 1 (0.8%) | 0 (0.0%) | 1 (0.8%) | 0 (0.0%) | 0 (0.0%) | 0 (0.0%) | 2 (0.3%) |
| - Acute kidney injury | 1 (0.8%) | 0 (0.0%) | 0 (0.0%) | 0 (0.0%) | 0 (0.0%) | 0 (0.0%) | 1 (0.1%) |

| *Event*<br>**System Organ Class (SOC)**<br>- Preferred Term (PT) | **9BLMZ (n = 126)** | **9BCLLfxZ (n = 122)** | **9BDLLfxZ (n = 127)** | **9DCLLfxZ (n = 124)** | **9DCMZ (n = 120)** | **Control (n = 126)** | **Total* (N = 745)** |
|---|---|---|---|---|---|---|---|
| - Chronic kidney disease | 0 (0.0%) | 0 (0.0%) | 0 (0.0%) | 0 (0.0%) | 0 (0.0%) | 1 (0.8%) | 1 (0.1%) |
| **Immune system disorders** | **0 (0.0%)** | **0 (0.0%)** | **2 (1.6%)** | **0 (0.0%)** | **0 (0.0%)** | **0 (0.0%)** | **2 (0.3%)** |
| - Hypersensitivity | 0 (0.0%) | 0 (0.0%) | 2 (1.6%) | 0 (0.0%) | 0 (0.0%) | 0 (0.0%) | 2 (0.3%) |
| **Pregnancy, puerperium and perinatal conditions** | **0 (0.0%)** | **0 (0.0%)** | **1 (0.8%)** | **0 (0.0%)** | **0 (0.0%)** | **1 (0.8%)** | **2 (0.3%)** |
| - Abortion spontaneous | 0 (0.0%) | 0 (0.0%) | 0 (0.0%) | 0 (0.0%) | 0 (0.0%) | 1 (0.8%) | 1 (0.1%) |
| - Abortion threatened | 0 (0.0%) | 0 (0.0%) | 1 (0.8%) | 0 (0.0%) | 0 (0.0%) | 0 (0.0%) | 1 (0.1%) |
| **Neoplasms benign, malignant and unspecified (incl cysts and polyps)** | **0 (0.0%)** | **0 (0.0%)** | **0 (0.0%)** | **0 (0.0%)** | **1 (0.8%)** | **1 (0.8%)** | **2 (0.3%)** |
| - Lung neoplasm malignant | 0 (0.0%) | 0 (0.0%) | 0 (0.0%) | 0 (0.0%) | 0 (0.0%) | 1 (0.8%) | 1 (0.1%) |
| - Pancreatic carcinoma | 0 (0.0%) | 0 (0.0%) | 0 (0.0%) | 0 (0.0%) | 1 (0.8%) | 0 (0.0%) | 1 (0.1%) |

**Total by preferred term (PT) represents the sum of participants who experienced at least one event corresponding to that PT. If they experienced more than one event of the same PT, it is counted only once. If a participant experienced more than one unique PT within a system organ class (SOC), each unique PT counts toward the PT total, but only one event counts toward the SOC.*

***Table S22. Number (%) of participants experiencing serious adverse events (SAEs) by SOC and PT by Week 73 (Safety Population)***

| *Event*<br>**System Organ Class (SOC)**<br>- Preferred Term (PT) | **9BLMZ (n = 126)** | **9BCLLfxZ (n = 122)** | **9BDLLfxZ (n = 127)** | **9DCLLfxZ (n = 124)** | **9DCMZ (n = 120)** | **Control (n = 126)** | **Total* (N = 745)** |
|---|---|---|---|---|---|---|---|
| **Infections and infestations** | **11 (8.7%)** | **3 (2.5%)** | **6 (4.7%)** | **3 (2.4%)** | **6 (5.0%)** | **4 (3.2%)** | **33 (4.4%)** |
| - Pneumonia | 4 (3.2%) | 0 (0.0%) | 3 (2.4%) | 0 (0.0%) | 1 (0.8%) | 2 (1.6%) | 10 (1.3%) |
| - COVID-19 | 2 (1.6%) | 0 (0.0%) | 0 (0.0%) | 0 (0.0%) | 1 (0.8%) | 1 (0.8%) | 4 (0.5%) |
| - COVID-19 pneumonia | 0 (0.0%) | 2 (1.6%) | 0 (0.0%) | 0 (0.0%) | 1 (0.8%) | 0 (0.0%) | 3 (0.4%) |
| - Sepsis | 0 (0.0%) | 1 (0.8%) | 0 (0.0%) | 1 (0.8%) | 1 (0.8%) | 0 (0.0%) | 3 (0.4%) |
| - Pneumocystis jirovecii pneumonia | 1 (0.8%) | 0 (0.0%) | 0 (0.0%) | 1 (0.8%) | 0 (0.0%) | 0 (0.0%) | 2 (0.3%) |
| - Abscess soft tissue | 0 (0.0%) | 0 (0.0%) | 0 (0.0%) | 0 (0.0%) | 1 (0.8%) | 0 (0.0%) | 1 (0.1%) |
| - Skin infection | 1 (0.8%) | 0 (0.0%) | 0 (0.0%) | 0 (0.0%) | 0 (0.0%) | 0 (0.0%) | 1 (0.1%) |
| - Tonsillitis | 0 (0.0%) | 0 (0.0%) | 0 (0.0%) | 0 (0.0%) | 0 (0.0%) | 1 (0.8%) | 1 (0.1%) |
| - Pneumonia staphylococcal | 0 (0.0%) | 0 (0.0%) | 1 (0.8%) | 0 (0.0%) | 0 (0.0%) | 0 (0.0%) | 1 (0.1%) |
| - Appendicitis | 0 (0.0%) | 0 (0.0%) | 0 (0.0%) | 0 (0.0%) | 0 (0.0%) | 1 (0.8%) | 1 (0.1%) |
| - Gastroenteritis | 0 (0.0%) | 0 (0.0%) | 0 (0.0%) | 1 (0.8%) | 0 (0.0%) | 0 (0.0%) | 1 (0.1%) |
| - Septic shock | 1 (0.8%) | 0 (0.0%) | 0 (0.0%) | 0 (0.0%) | 0 (0.0%) | 0 (0.0%) | 1 (0.1%) |
| - Cellulitis | 0 (0.0%) | 0 (0.0%) | 1 (0.8%) | 0 (0.0%) | 0 (0.0%) | 0 (0.0%) | 1 (0.1%) |

| *Event*<br>**System Organ Class (SOC)**<br>- Preferred Term (PT) | **9BLMZ**<br>**(n = 126)** | **9BCLLfxZ**<br>**(n = 122)** | **9BDLLfxZ**<br>**(n = 127)** | **9DCLLfxZ**<br>**(n = 124)** | **9DCMZ**<br>**(n = 120)** | **Control**<br>**(n = 126)** | **Total***<br>**(N = 745)** |
|---|---|---|---|---|---|---|---|
| - Herpes zoster | 0 (0.0%) | 0 (0.0%) | 0 (0.0%) | 0 (0.0%) | 1 (0.8%) | 0 (0.0%) | 1 (0.1%) |
| - Wound infection | 1 (0.8%) | 0 (0.0%) | 0 (0.0%) | 0 (0.0%) | 0 (0.0%) | 0 (0.0%) | 1 (0.1%) |
| - Pulmonary tuberculoma | 0 (0.0%) | 0 (0.0%) | 1 (0.8%) | 0 (0.0%) | 0 (0.0%) | 0 (0.0%) | 1 (0.1%) |
| - Respiratory tract infection | 0 (0.0%) | 0 (0.0%) | 1 (0.8%) | 0 (0.0%) | 0 (0.0%) | 0 (0.0%) | 1 (0.1%) |
| - Pneumonia bacterial | 0 (0.0%) | 0 (0.0%) | 1 (0.8%) | 0 (0.0%) | 0 (0.0%) | 0 (0.0%) | 1 (0.1%) |
| - Post procedural sepsis | 0 (0.0%) | 0 (0.0%) | 0 (0.0%) | 1 (0.8%) | 0 (0.0%) | 0 (0.0%) | 1 (0.1%) |
| - Tuberculosis | 1 (0.8%) | 0 (0.0%) | 0 (0.0%) | 0 (0.0%) | 0 (0.0%) | 0 (0.0%) | 1 (0.1%) |
| - Dengue fever | 1 (0.8%) | 0 (0.0%) | 0 (0.0%) | 0 (0.0%) | 0 (0.0%) | 0 (0.0%) | 1 (0.1%) |
| **Blood and lymphatic system disorders** | **1 (0.8%)** | **3 (2.5%)** | **3 (2.4%)** | **3 (2.4%)** | **2 (1.7%)** | **4 (3.2%)** | **16 (2.1%)** |
| - Anaemia | 1 (0.8%) | 3 (2.5%) | 3 (2.4%) | 2 (1.6%) | 2 (1.7%) | 4 (3.2%) | 15 (2.0%) |
| - Pancytopenia | 0 (0.0%) | 0 (0.0%) | 0 (0.0%) | 1 (0.8%) | 0 (0.0%) | 0 (0.0%) | 1 (0.1%) |
| **Investigations** | **3 (2.4%)** | **4 (3.3%)** | **3 (2.4%)** | **0 (0.0%)** | **2 (1.7%)** | **3 (2.4%)** | **15 (2.0%)** |
| - Alanine aminotransferase increased | 3 (2.4%) | 1 (0.8%) | 2 (1.6%) | 0 (0.0%) | 0 (0.0%) | 0 (0.0%) | 6 (0.8%) |
| - Aspartate aminotransferase increased | 1 (0.8%) | 1 (0.8%) | 2 (1.6%) | 0 (0.0%) | 0 (0.0%) | 1 (0.8%) | 5 (0.7%) |
| - Electrocardiogram QT prolonged | 0 (0.0%) | 2 (1.6%) | 0 (0.0%) | 0 (0.0%) | 2 (1.7%) | 0 (0.0%) | 4 (0.5%) |
| - Bilirubin conjugated increased | 0 (0.0%) | 0 (0.0%) | 1 (0.8%) | 0 (0.0%) | 0 (0.0%) | 1 (0.8%) | 2 (0.3%) |
| - Lymphocyte count decreased | 0 (0.0%) | 0 (0.0%) | 1 (0.8%) | 0 (0.0%) | 0 (0.0%) | 0 (0.0%) | 1 (0.1%) |
| - Neutrophil count decreased | 0 (0.0%) | 0 (0.0%) | 0 (0.0%) | 0 (0.0%) | 0 (0.0%) | 1 (0.8%) | 1 (0.1%) |
| - Transaminases increased | 0 (0.0%) | 1 (0.8%) | 0 (0.0%) | 0 (0.0%) | 0 (0.0%) | 0 (0.0%) | 1 (0.1%) |
| **Respiratory, thoracic and mediastinal disorders** | **0 (0.0%)** | **1 (0.8%)** | **3 (2.4%)** | **2 (1.6%)** | **2 (1.7%)** | **5 (4.0%)** | **13 (1.7%)** |
| - Haemoptysis | 0 (0.0%) | 0 (0.0%) | 0 (0.0%) | 0 (0.0%) | 1 (0.8%) | 3 (2.4%) | 4 (0.5%) |
| - Pneumothorax | 0 (0.0%) | 0 (0.0%) | 0 (0.0%) | 1 (0.8%) | 1 (0.8%) | 1 (0.8%) | 3 (0.4%) |
| - Respiratory failure | 0 (0.0%) | 0 (0.0%) | 1 (0.8%) | 0 (0.0%) | 0 (0.0%) | 1 (0.8%) | 2 (0.3%) |
| - Dyspnoea | 0 (0.0%) | 0 (0.0%) | 1 (0.8%) | 1 (0.8%) | 0 (0.0%) | 0 (0.0%) | 2 (0.3%) |
| - Pulmonary cavitation | 0 (0.0%) | 0 (0.0%) | 0 (0.0%) | 0 (0.0%) | 0 (0.0%) | 1 (0.8%) | 1 (0.1%) |
| - Acute respiratory failure | 0 (0.0%) | 0 (0.0%) | 1 (0.8%) | 0 (0.0%) | 0 (0.0%) | 0 (0.0%) | 1 (0.1%) |
| - Acute pulmonary oedema | 0 (0.0%) | 1 (0.8%) | 0 (0.0%) | 0 (0.0%) | 0 (0.0%) | 0 (0.0%) | 1 (0.1%) |
| **Gastrointestinal disorders** | **1 (0.8%)** | **1 (0.8%)** | **1 (0.8%)** | **4 (3.2%)** | **1 (0.8%)** | **3 (2.4%)** | **11 (1.5%)** |
| - Vomiting | 0 (0.0%) | 1 (0.8%) | 0 (0.0%) | 3 (2.4%) | 0 (0.0%) | 2 (1.6%) | 6 (0.8%) |
| - Small intestinal obstruction | 0 (0.0%) | 0 (0.0%) | 0 (0.0%) | 1 (0.8%) | 0 (0.0%) | 0 (0.0%) | 1 (0.1%) |
| - Acute abdomen | 0 (0.0%) | 0 (0.0%) | 1 (0.8%) | 0 (0.0%) | 0 (0.0%) | 0 (0.0%) | 1 (0.1%) |
| - Pancreatitis | 0 (0.0%) | 0 (0.0%) | 0 (0.0%) | 0 (0.0%) | 0 (0.0%) | 1 (0.8%) | 1 (0.1%) |
| - Gastritis | 0 (0.0%) | 0 (0.0%) | 0 (0.0%) | 0 (0.0%) | 1 (0.8%) | 0 (0.0%) | 1 (0.1%) |
| - Upper gastrointestinal haemorrhage | 0 (0.0%) | 0 (0.0%) | 0 (0.0%) | 0 (0.0%) | 1 (0.8%) | 0 (0.0%) | 1 (0.1%) |
| - Abdominal hernia | 1 (0.8%) | 0 (0.0%) | 0 (0.0%) | 0 (0.0%) | 0 (0.0%) | 0 (0.0%) | 1 (0.1%) |

| *Event*<br>**System Organ Class (SOC)**<br>- Preferred Term (PT) | **9BLMZ**<br>**(n = 126)** | **9BCLLfxZ**<br>**(n = 122)** | **9BDLLfxZ**<br>**(n = 127)** | **9DCLLfxZ**<br>**(n = 124)** | **9DCMZ**<br>**(n = 120)** | **Control**<br>**(n = 126)** | **Total***<br>**(N = 745)** |
|---|---|---|---|---|---|---|---|
| - Nausea | 0 (0.0%) | 0 (0.0%) | 0 (0.0%) | 1 (0.8%) | 0 (0.0%) | 0 (0.0%) | 1 (0.1%) |
| **Injury, poisoning and procedural complications** | **0 (0.0%)** | **1 (0.8%)** | **2 (1.6%)** | **1 (0.8%)** | **2 (1.7%)** | **4 (3.2%)** | **10 (1.3%)** |
| - Alcohol poisoning | 0 (0.0%) | 1 (0.8%) | 0 (0.0%) | 0 (0.0%) | 0 (0.0%) | 2 (1.6%) | 3 (0.4%) |
| - Hip fracture | 0 (0.0%) | 0 (0.0%) | 0 (0.0%) | 0 (0.0%) | 1 (0.8%) | 0 (0.0%) | 1 (0.1%) |
| - Stab wound | 0 (0.0%) | 0 (0.0%) | 0 (0.0%) | 0 (0.0%) | 0 (0.0%) | 1 (0.8%) | 1 (0.1%) |
| - Thoracic vertebral fracture | 0 (0.0%) | 0 (0.0%) | 0 (0.0%) | 1 (0.8%) | 0 (0.0%) | 0 (0.0%) | 1 (0.1%) |
| - Thermal burn | 0 (0.0%) | 0 (0.0%) | 1 (0.8%) | 0 (0.0%) | 0 (0.0%) | 0 (0.0%) | 1 (0.1%) |
| - Ankle fracture | 0 (0.0%) | 0 (0.0%) | 0 (0.0%) | 0 (0.0%) | 0 (0.0%) | 1 (0.8%) | 1 (0.1%) |
| - Abortion induced incomplete | 0 (0.0%) | 0 (0.0%) | 1 (0.8%) | 0 (0.0%) | 0 (0.0%) | 0 (0.0%) | 1 (0.1%) |
| - Femur fracture | 0 (0.0%) | 0 (0.0%) | 0 (0.0%) | 0 (0.0%) | 1 (0.8%) | 0 (0.0%) | 1 (0.1%) |
| **Psychiatric disorders** | **0 (0.0%)** | **3 (2.5%)** | **1 (0.8%)** | **0 (0.0%)** | **3 (2.5%)** | **2 (1.6%)** | **9 (1.2%)** |
| - Suicide attempt | 0 (0.0%) | 1 (0.8%) | 0 (0.0%) | 0 (0.0%) | 3 (2.5%) | 1 (0.8%) | 5 (0.7%) |
| - Depression | 0 (0.0%) | 0 (0.0%) | 0 (0.0%) | 0 (0.0%) | 0 (0.0%) | 1 (0.8%) | 1 (0.1%) |
| - Anxiety disorder | 0 (0.0%) | 0 (0.0%) | 1 (0.8%) | 0 (0.0%) | 0 (0.0%) | 0 (0.0%) | 1 (0.1%) |
| - Drug dependence | 0 (0.0%) | 1 (0.8%) | 0 (0.0%) | 0 (0.0%) | 0 (0.0%) | 0 (0.0%) | 1 (0.1%) |
| - Psychotic disorder | 0 (0.0%) | 1 (0.8%) | 0 (0.0%) | 0 (0.0%) | 0 (0.0%) | 0 (0.0%) | 1 (0.1%) |
| **Hepatobiliary disorders** | **1 (0.8%)** | **1 (0.8%)** | **0 (0.0%)** | **1 (0.8%)** | **2 (1.7%)** | **2 (1.6%)** | **7 (0.9%)** |
| - Hepatotoxicity | 1 (0.8%) | 0 (0.0%) | 0 (0.0%) | 0 (0.0%) | 0 (0.0%) | 1 (0.8%) | 2 (0.3%) |
| - Drug-induced liver injury | 0 (0.0%) | 1 (0.8%) | 0 (0.0%) | 1 (0.8%) | 0 (0.0%) | 0 (0.0%) | 2 (0.3%) |
| - Hepatitis | 0 (0.0%) | 0 (0.0%) | 0 (0.0%) | 0 (0.0%) | 0 (0.0%) | 1 (0.8%) | 1 (0.1%) |
| - Acute on chronic liver failure | 0 (0.0%) | 0 (0.0%) | 0 (0.0%) | 0 (0.0%) | 1 (0.8%) | 0 (0.0%) | 1 (0.1%) |
| - Hyperbilirubinaemia | 0 (0.0%) | 0 (0.0%) | 0 (0.0%) | 0 (0.0%) | 1 (0.8%) | 0 (0.0%) | 1 (0.1%) |
| **Nervous system disorders** | **0 (0.0%)** | **2 (1.6%)** | **2 (1.6%)** | **2 (1.6%)** | **0 (0.0%)** | **1 (0.8%)** | **7 (0.9%)** |
| - Seizure | 0 (0.0%) | 1 (0.8%) | 0 (0.0%) | 1 (0.8%) | 0 (0.0%) | 0 (0.0%) | 2 (0.3%) |
| - Idiopathic intracranial hypertension | 0 (0.0%) | 0 (0.0%) | 0 (0.0%) | 0 (0.0%) | 0 (0.0%) | 1 (0.8%) | 1 (0.1%) |
| - Facial paralysis | 0 (0.0%) | 0 (0.0%) | 1 (0.8%) | 0 (0.0%) | 0 (0.0%) | 0 (0.0%) | 1 (0.1%) |
| - Optic neuritis | 0 (0.0%) | 0 (0.0%) | 0 (0.0%) | 1 (0.8%) | 0 (0.0%) | 0 (0.0%) | 1 (0.1%) |
| - Neuropathy peripheral | 0 (0.0%) | 0 (0.0%) | 1 (0.8%) | 0 (0.0%) | 0 (0.0%) | 0 (0.0%) | 1 (0.1%) |
| - Transient ischaemic attack | 0 (0.0%) | 1 (0.8%) | 0 (0.0%) | 0 (0.0%) | 0 (0.0%) | 0 (0.0%) | 1 (0.1%) |
| **General disorders and administration site conditions** | **2 (1.6%)** | **0 (0.0%)** | **0 (0.0%)** | **2 (1.6%)** | **2 (1.7%)** | **0 (0.0%)** | **6 (0.8%)** |
| - Sudden death | 0 (0.0%) | 0 (0.0%) | 0 (0.0%) | 0 (0.0%) | 1 (0.8%) | 0 (0.0%) | 1 (0.1%) |
| - Generalised oedema | 1 (0.8%) | 0 (0.0%) | 0 (0.0%) | 0 (0.0%) | 0 (0.0%) | 0 (0.0%) | 1 (0.1%) |
| - Pyrexia | 1 (0.8%) | 0 (0.0%) | 0 (0.0%) | 0 (0.0%) | 0 (0.0%) | 0 (0.0%) | 1 (0.1%) |
| - Disease progression | 0 (0.0%) | 0 (0.0%) | 0 (0.0%) | 1 (0.8%) | 0 (0.0%) | 0 (0.0%) | 1 (0.1%) |
| - Chest pain | 0 (0.0%) | 0 (0.0%) | 0 (0.0%) | 0 (0.0%) | 1 (0.8%) | 0 (0.0%) | 1 (0.1%) |

| *Event*<br>**System Organ Class (SOC)**<br>- Preferred Term (PT) | **9BLMZ**<br>**(n = 126)** | **9BCLLfxZ**<br>**(n = 122)** | **9BDLLfxZ**<br>**(n = 127)** | **9DCLLfxZ**<br>**(n = 124)** | **9DCMZ**<br>**(n = 120)** | **Control**<br>**(n = 126)** | **Total***<br>**(N = 745)** |
|---|---|---|---|---|---|---|---|
| - Death | 0 (0.0%) | 0 (0.0%) | 0 (0.0%) | 1 (0.8%) | 0 (0.0%) | 0 (0.0%) | 1 (0.1%) |
| **Cardiac disorders** | **1 (0.8%)** | **2 (1.6%)** | **0 (0.0%)** | **2 (1.6%)** | **0 (0.0%)** | **0 (0.0%)** | **5 (0.7%)** |
| - Cardiac failure congestive | 1 (0.8%) | 1 (0.8%) | 0 (0.0%) | 0 (0.0%) | 0 (0.0%) | 0 (0.0%) | 2 (0.3%) |
| - Cardiac failure | 0 (0.0%) | 1 (0.8%) | 0 (0.0%) | 1 (0.8%) | 0 (0.0%) | 0 (0.0%) | 2 (0.3%) |
| - Acute myocardial infarction | 0 (0.0%) | 1 (0.8%) | 0 (0.0%) | 1 (0.8%) | 0 (0.0%) | 0 (0.0%) | 2 (0.3%) |
| **Metabolism and nutrition disorders** | **1 (0.8%)** | **2 (1.6%)** | **0 (0.0%)** | **0 (0.0%)** | **2 (1.7%)** | **0 (0.0%)** | **5 (0.7%)** |
| - Diabetes mellitus inadequate control | 0 (0.0%) | 0 (0.0%) | 0 (0.0%) | 0 (0.0%) | 2 (1.7%) | 0 (0.0%) | 2 (0.3%) |
| - Hyperglycaemia | 0 (0.0%) | 2 (1.6%) | 0 (0.0%) | 0 (0.0%) | 0 (0.0%) | 0 (0.0%) | 2 (0.3%) |
| - Hypoalbuminaemia | 1 (0.8%) | 0 (0.0%) | 0 (0.0%) | 0 (0.0%) | 0 (0.0%) | 0 (0.0%) | 1 (0.1%) |
| **Skin and subcutaneous tissue disorders** | **0 (0.0%)** | **0 (0.0%)** | **0 (0.0%)** | **1 (0.8%)** | **3 (2.5%)** | **0 (0.0%)** | **4 (0.5%)** |
| - Drug reaction with eosinophilia and systemic symptoms | 0 (0.0%) | 0 (0.0%) | 0 (0.0%) | 1 (0.8%) | 1 (0.8%) | 0 (0.0%) | 2 (0.3%) |
| - Swelling face | 0 (0.0%) | 0 (0.0%) | 0 (0.0%) | 0 (0.0%) | 1 (0.8%) | 0 (0.0%) | 1 (0.1%) |
| - Dermatitis exfoliative | 0 (0.0%) | 0 (0.0%) | 0 (0.0%) | 0 (0.0%) | 1 (0.8%) | 0 (0.0%) | 1 (0.1%) |
| **Musculoskeletal and connective tissue disorders** | **0 (0.0%)** | **1 (0.8%)** | **0 (0.0%)** | **1 (0.8%)** | **2 (1.7%)** | **0 (0.0%)** | **4 (0.5%)** |
| - Arthralgia | 0 (0.0%) | 0 (0.0%) | 0 (0.0%) | 1 (0.8%) | 1 (0.8%) | 0 (0.0%) | 2 (0.3%) |
| - Arthropathy | 0 (0.0%) | 1 (0.8%) | 0 (0.0%) | 0 (0.0%) | 0 (0.0%) | 0 (0.0%) | 1 (0.1%) |
| - Osteoarthritis | 0 (0.0%) | 0 (0.0%) | 0 (0.0%) | 0 (0.0%) | 1 (0.8%) | 0 (0.0%) | 1 (0.1%) |
| **Eye disorders** | **1 (0.8%)** | **2 (1.6%)** | **0 (0.0%)** | **0 (0.0%)** | **0 (0.0%)** | **0 (0.0%)** | **3 (0.4%)** |
| - Optic neuropathy | 1 (0.8%) | 1 (0.8%) | 0 (0.0%) | 0 (0.0%) | 0 (0.0%) | 0 (0.0%) | 2 (0.3%) |
| - Eye haemorrhage | 0 (0.0%) | 1 (0.8%) | 0 (0.0%) | 0 (0.0%) | 0 (0.0%) | 0 (0.0%) | 1 (0.1%) |
| **Vascular disorders** | **1 (0.8%)** | **1 (0.8%)** | **0 (0.0%)** | **1 (0.8%)** | **0 (0.0%)** | **0 (0.0%)** | **3 (0.4%)** |
| - Deep vein thrombosis | 0 (0.0%) | 0 (0.0%) | 0 (0.0%) | 1 (0.8%) | 0 (0.0%) | 0 (0.0%) | 1 (0.1%) |
| - Hypertension | 0 (0.0%) | 1 (0.8%) | 0 (0.0%) | 0 (0.0%) | 0 (0.0%) | 0 (0.0%) | 1 (0.1%) |
| - Diabetic vascular disorder | 1 (0.8%) | 0 (0.0%) | 0 (0.0%) | 0 (0.0%) | 0 (0.0%) | 0 (0.0%) | 1 (0.1%) |
| **Renal and urinary disorders** | **1 (0.8%)** | **0 (0.0%)** | **1 (0.8%)** | **0 (0.0%)** | **0 (0.0%)** | **0 (0.0%)** | **2 (0.3%)** |
| - Proteinuria | 0 (0.0%) | 0 (0.0%) | 1 (0.8%) | 0 (0.0%) | 0 (0.0%) | 0 (0.0%) | 1 (0.1%) |
| - Acute kidney injury | 1 (0.8%) | 0 (0.0%) | 0 (0.0%) | 0 (0.0%) | 0 (0.0%) | 0 (0.0%) | 1 (0.1%) |
| **Immune system disorders** | **0 (0.0%)** | **0 (0.0%)** | **2 (1.6%)** | **0 (0.0%)** | **0 (0.0%)** | **0 (0.0%)** | **2 (0.3%)** |
| - Hypersensitivity | 0 (0.0%) | 0 (0.0%) | 2 (1.6%) | 0 (0.0%) | 0 (0.0%) | 0 (0.0%) | 2 (0.3%) |
| **Pregnancy, puerperium and perinatal conditions** | **0 (0.0%)** | **0 (0.0%)** | **1 (0.8%)** | **0 (0.0%)** | **0 (0.0%)** | **0 (0.0%)** | **1 (0.1%)** |
| - Abortion threatened | 0 (0.0%) | 0 (0.0%) | 1 (0.8%) | 0 (0.0%) | 0 (0.0%) | 0 (0.0%) | 1 (0.1%) |
| **Neoplasms benign, malignant and unspecified (incl cysts and polyps)** | **0 (0.0%)** | **0 (0.0%)** | **0 (0.0%)** | **0 (0.0%)** | **0 (0.0%)** | **1 (0.8%)** | **1 (0.1%)** |
| - Lung neoplasm malignant | 0 (0.0%) | 0 (0.0%) | 0 (0.0%) | 0 (0.0%) | 0 (0.0%) | 1 (0.8%) | 1 (0.1%) |

| *Event*<br>**System Organ Class (SOC)**<br>- Preferred Term (PT) | **9BLMZ**<br>**(n = 126)** | **9BCLLfxZ**<br>**(n = 122)** | **9BDLLfxZ**<br>**(n = 127)** | **9DCLLfxZ**<br>**(n = 124)** | **9DCMZ**<br>**(n = 120)** | **Control**<br>**(n = 126)** | **Total***<br>**(N = 745)** |
|---|---|---|---|---|---|---|---|
| **Product issues** | **1 (0.8%)** | **0 (0.0%)** | **0 (0.0%)** | **0 (0.0%)** | **0 (0.0%)** | **0 (0.0%)** | **1 (0.1%)** |
| - Device dislocation | 1 (0.8%) | 0 (0.0%) | 0 (0.0%) | 0 (0.0%) | 0 (0.0%) | 0 (0.0%) | 1 (0.1%) |

**Total by preferred term (PT) represents the sum of participants who experienced at least one event corresponding to that PT. If they experienced more than one event of the same PT, it is counted only once. If a participant experienced more than one unique PT within a system organ class (SOC), each unique PT counts toward the PT total, but only one event counts toward the SOC.*

*Table S23.* ***Number (%) of participants experiencing serious adverse events (SAEs) by SOC and PT by Week 104 (Safety Population)***

| *Event*<br>**System Organ Class (SOC)**<br>- Preferred Term (PT) | **9BLMZ**<br>**(n = 126)** | **9BCLLfxZ**<br>**(n = 122)** | **9BDLLfxZ**<br>**(n = 127)** | **9DCLLfxZ**<br>**(n = 124)** | **9DCMZ**<br>**(n = 120)** | **Control**<br>**(n = 126)** | **Total***<br>**(N = 745)** |
|---|---|---|---|---|---|---|---|
| **Infections and infestations** | **12 (9.5%)** | **6 (4.9%)** | **6 (4.7%)** | **3 (2.4%)** | **7 (5.8%)** | **4 (3.2%)** | **38 (5.1%)** |
| - Pneumonia | 4 (3.2%) | 1 (0.8%) | 3 (2.4%) | 0 (0.0%) | 1 (0.8%) | 2 (1.6%) | 11 (1.5%) |
| - COVID-19 | 2 (1.6%) | 0 (0.0%) | 0 (0.0%) | 0 (0.0%) | 2 (1.7%) | 1 (0.8%) | 5 (0.7%) |
| - COVID-19 pneumonia | 0 (0.0%) | 4 (3.3%) | 0 (0.0%) | 0 (0.0%) | 1 (0.8%) | 0 (0.0%) | 5 (0.7%) |
| - Sepsis | 0 (0.0%) | 1 (0.8%) | 0 (0.0%) | 1 (0.8%) | 1 (0.8%) | 0 (0.0%) | 3 (0.4%) |
| - Appendicitis | 1 (0.8%) | 0 (0.0%) | 0 (0.0%) | 0 (0.0%) | 0 (0.0%) | 1 (0.8%) | 2 (0.3%) |
| - Pneumocystis jirovecii pneumonia | 1 (0.8%) | 0 (0.0%) | 0 (0.0%) | 1 (0.8%) | 0 (0.0%) | 0 (0.0%) | 2 (0.3%) |
| - Pneumonia bacterial | 0 (0.0%) | 0 (0.0%) | 1 (0.8%) | 0 (0.0%) | 0 (0.0%) | 0 (0.0%) | 1 (0.1%) |
| - Respiratory tract infection | 0 (0.0%) | 0 (0.0%) | 1 (0.8%) | 0 (0.0%) | 0 (0.0%) | 0 (0.0%) | 1 (0.1%) |
| - Post procedural sepsis | 0 (0.0%) | 0 (0.0%) | 0 (0.0%) | 1 (0.8%) | 0 (0.0%) | 0 (0.0%) | 1 (0.1%) |
| - Gastroenteritis | 0 (0.0%) | 0 (0.0%) | 0 (0.0%) | 1 (0.8%) | 0 (0.0%) | 0 (0.0%) | 1 (0.1%) |
| - Diabetic foot infection | 1 (0.8%) | 0 (0.0%) | 0 (0.0%) | 0 (0.0%) | 0 (0.0%) | 0 (0.0%) | 1 (0.1%) |
| - Pneumonia staphylococcal | 0 (0.0%) | 0 (0.0%) | 1 (0.8%) | 0 (0.0%) | 0 (0.0%) | 0 (0.0%) | 1 (0.1%) |
| - Tonsillitis | 0 (0.0%) | 0 (0.0%) | 0 (0.0%) | 0 (0.0%) | 0 (0.0%) | 1 (0.8%) | 1 (0.1%) |
| - Skin infection | 1 (0.8%) | 0 (0.0%) | 0 (0.0%) | 0 (0.0%) | 0 (0.0%) | 0 (0.0%) | 1 (0.1%) |
| - Dengue fever | 1 (0.8%) | 0 (0.0%) | 0 (0.0%) | 0 (0.0%) | 0 (0.0%) | 0 (0.0%) | 1 (0.1%) |
| - Herpes zoster | 0 (0.0%) | 0 (0.0%) | 0 (0.0%) | 0 (0.0%) | 1 (0.8%) | 0 (0.0%) | 1 (0.1%) |
| - Septic shock | 1 (0.8%) | 0 (0.0%) | 0 (0.0%) | 0 (0.0%) | 0 (0.0%) | 0 (0.0%) | 1 (0.1%) |
| - Cellulitis | 0 (0.0%) | 0 (0.0%) | 1 (0.8%) | 0 (0.0%) | 0 (0.0%) | 0 (0.0%) | 1 (0.1%) |
| - Wound infection | 1 (0.8%) | 0 (0.0%) | 0 (0.0%) | 0 (0.0%) | 0 (0.0%) | 0 (0.0%) | 1 (0.1%) |
| - Tuberculosis | 1 (0.8%) | 0 (0.0%) | 0 (0.0%) | 0 (0.0%) | 0 (0.0%) | 0 (0.0%) | 1 (0.1%) |

| *Event*<br>**System Organ Class (SOC)**<br>- Preferred Term (PT) | **9BLMZ**<br>**(n = 126)** | **9BCLLfxZ**<br>**(n = 122)** | **9BDLLfxZ**<br>**(n = 127)** | **9DCLLfxZ**<br>**(n = 124)** | **9DCMZ**<br>**(n = 120)** | **Control**<br>**(n = 126)** | **Total***<br>**(N = 745)** |
|---|---|---|---|---|---|---|---|
| - Pulmonary tuberculoma | 0 (0.0%) | 0 (0.0%) | 1 (0.8%) | 0 (0.0%) | 0 (0.0%) | 0 (0.0%) | 1 (0.1%) |
| - Abscess soft tissue | 0 (0.0%) | 0 (0.0%) | 0 (0.0%) | 0 (0.0%) | 1 (0.8%) | 0 (0.0%) | 1 (0.1%) |
| - Osteomyelitis | 1 (0.8%) | 0 (0.0%) | 0 (0.0%) | 0 (0.0%) | 0 (0.0%) | 0 (0.0%) | 1 (0.1%) |
| **Blood and lymphatic system disorders** | **1 (0.8%)** | **3 (2.5%)** | **3 (2.4%)** | **4 (3.2%)** | **2 (1.7%)** | **4 (3.2%)** | **17 (2.3%)** |
| - Anaemia | 1 (0.8%) | 3 (2.5%) | 3 (2.4%) | 3 (2.4%) | 2 (1.7%) | 4 (3.2%) | 16 (2.1%) |
| - Pancytopenia | 0 (0.0%) | 0 (0.0%) | 0 (0.0%) | 1 (0.8%) | 0 (0.0%) | 0 (0.0%) | 1 (0.1%) |
| **Investigations** | **3 (2.4%)** | **4 (3.3%)** | **3 (2.4%)** | **0 (0.0%)** | **2 (1.7%)** | **3 (2.4%)** | **15 (2.0%)** |
| - Alanine aminotransferase increased | 3 (2.4%) | 1 (0.8%) | 2 (1.6%) | 0 (0.0%) | 0 (0.0%) | 0 (0.0%) | 6 (0.8%) |
| - Aspartate aminotransferase increased | 1 (0.8%) | 1 (0.8%) | 2 (1.6%) | 0 (0.0%) | 0 (0.0%) | 1 (0.8%) | 5 (0.7%) |
| - Electrocardiogram QT prolonged | 0 (0.0%) | 2 (1.6%) | 0 (0.0%) | 0 (0.0%) | 2 (1.7%) | 0 (0.0%) | 4 (0.5%) |
| - Bilirubin conjugated increased | 0 (0.0%) | 0 (0.0%) | 1 (0.8%) | 0 (0.0%) | 0 (0.0%) | 1 (0.8%) | 2 (0.3%) |
| - Lymphocyte count decreased | 0 (0.0%) | 0 (0.0%) | 1 (0.8%) | 0 (0.0%) | 0 (0.0%) | 0 (0.0%) | 1 (0.1%) |
| - Transaminases increased | 0 (0.0%) | 1 (0.8%) | 0 (0.0%) | 0 (0.0%) | 0 (0.0%) | 0 (0.0%) | 1 (0.1%) |
| - Neutrophil count decreased | 0 (0.0%) | 0 (0.0%) | 0 (0.0%) | 0 (0.0%) | 0 (0.0%) | 1 (0.8%) | 1 (0.1%) |
| **Respiratory, thoracic and mediastinal disorders** | **0 (0.0%)** | **2 (1.6%)** | **3 (2.4%)** | **2 (1.6%)** | **2 (1.7%)** | **5 (4.0%)** | **14 (1.9%)** |
| - Haemoptysis | 0 (0.0%) | 0 (0.0%) | 0 (0.0%) | 0 (0.0%) | 1 (0.8%) | 3 (2.4%) | 4 (0.5%) |
| - Pneumothorax | 0 (0.0%) | 0 (0.0%) | 0 (0.0%) | 1 (0.8%) | 1 (0.8%) | 1 (0.8%) | 3 (0.4%) |
| - Respiratory failure | 0 (0.0%) | 0 (0.0%) | 1 (0.8%) | 0 (0.0%) | 0 (0.0%) | 1 (0.8%) | 2 (0.3%) |
| - Dyspnoea | 0 (0.0%) | 0 (0.0%) | 1 (0.8%) | 1 (0.8%) | 0 (0.0%) | 0 (0.0%) | 2 (0.3%) |
| - Pulmonary haemorrhage | 0 (0.0%) | 1 (0.8%) | 0 (0.0%) | 0 (0.0%) | 0 (0.0%) | 0 (0.0%) | 1 (0.1%) |
| - Pulmonary cavitation | 0 (0.0%) | 0 (0.0%) | 0 (0.0%) | 0 (0.0%) | 0 (0.0%) | 1 (0.8%) | 1 (0.1%) |
| - Acute respiratory failure | 0 (0.0%) | 0 (0.0%) | 1 (0.8%) | 0 (0.0%) | 0 (0.0%) | 0 (0.0%) | 1 (0.1%) |
| - Acute pulmonary oedema | 0 (0.0%) | 1 (0.8%) | 0 (0.0%) | 0 (0.0%) | 0 (0.0%) | 0 (0.0%) | 1 (0.1%) |
| **Gastrointestinal disorders** | **1 (0.8%)** | **1 (0.8%)** | **1 (0.8%)** | **4 (3.2%)** | **2 (1.7%)** | **5 (4.0%)** | **14 (1.9%)** |
| - Vomiting | 0 (0.0%) | 1 (0.8%) | 0 (0.0%) | 3 (2.4%) | 0 (0.0%) | 2 (1.6%) | 6 (0.8%) |
| - Pancreatitis | 0 (0.0%) | 0 (0.0%) | 0 (0.0%) | 0 (0.0%) | 0 (0.0%) | 1 (0.8%) | 1 (0.1%) |
| - Abdominal hernia | 1 (0.8%) | 0 (0.0%) | 0 (0.0%) | 0 (0.0%) | 0 (0.0%) | 0 (0.0%) | 1 (0.1%) |
| - Acute abdomen | 0 (0.0%) | 0 (0.0%) | 1 (0.8%) | 0 (0.0%) | 0 (0.0%) | 0 (0.0%) | 1 (0.1%) |
| - Abdominal pain upper | 0 (0.0%) | 0 (0.0%) | 0 (0.0%) | 0 (0.0%) | 1 (0.8%) | 0 (0.0%) | 1 (0.1%) |
| - Mallory-Weiss syndrome | 0 (0.0%) | 0 (0.0%) | 0 (0.0%) | 0 (0.0%) | 0 (0.0%) | 1 (0.8%) | 1 (0.1%) |
| - Gastritis | 0 (0.0%) | 0 (0.0%) | 0 (0.0%) | 0 (0.0%) | 1 (0.8%) | 0 (0.0%) | 1 (0.1%) |
| - Upper gastrointestinal haemorrhage | 0 (0.0%) | 0 (0.0%) | 0 (0.0%) | 0 (0.0%) | 1 (0.8%) | 0 (0.0%) | 1 (0.1%) |
| - Pancreatitis acute | 0 (0.0%) | 0 (0.0%) | 0 (0.0%) | 0 (0.0%) | 0 (0.0%) | 1 (0.8%) | 1 (0.1%) |
| - Nausea | 0 (0.0%) | 0 (0.0%) | 0 (0.0%) | 1 (0.8%) | 0 (0.0%) | 0 (0.0%) | 1 (0.1%) |
| - Small intestinal obstruction | 0 (0.0%) | 0 (0.0%) | 0 (0.0%) | 1 (0.8%) | 0 (0.0%) | 0 (0.0%) | 1 (0.1%) |

| *Event*<br>**System Organ Class (SOC)**<br>- Preferred Term (PT) | **9BLMZ**<br>**(n = 126)** | **9BCLLfxZ**<br>**(n = 122)** | **9BDLLfxZ**<br>**(n = 127)** | **9DCLLfxZ**<br>**(n = 124)** | **9DCMZ**<br>**(n = 120)** | **Control**<br>**(n = 126)** | **Total***<br>**(N = 745)** |
|---|---|---|---|---|---|---|---|
| **Injury, poisoning and procedural complications** | **0 (0.0%)** | **1 (0.8%)** | **2 (1.6%)** | **1 (0.8%)** | **2 (1.7%)** | **4 (3.2%)** | **10 (1.3%)** |
| - Alcohol poisoning | 0 (0.0%) | 1 (0.8%) | 0 (0.0%) | 0 (0.0%) | 0 (0.0%) | 2 (1.6%) | 3 (0.4%) |
| - Thermal burn | 0 (0.0%) | 0 (0.0%) | 1 (0.8%) | 0 (0.0%) | 0 (0.0%) | 0 (0.0%) | 1 (0.1%) |
| - Hip fracture | 0 (0.0%) | 0 (0.0%) | 0 (0.0%) | 0 (0.0%) | 1 (0.8%) | 0 (0.0%) | 1 (0.1%) |
| - Ankle fracture | 0 (0.0%) | 0 (0.0%) | 0 (0.0%) | 0 (0.0%) | 0 (0.0%) | 1 (0.8%) | 1 (0.1%) |
| - Thoracic vertebral fracture | 0 (0.0%) | 0 (0.0%) | 0 (0.0%) | 1 (0.8%) | 0 (0.0%) | 0 (0.0%) | 1 (0.1%) |
| - Stab wound | 0 (0.0%) | 0 (0.0%) | 0 (0.0%) | 0 (0.0%) | 0 (0.0%) | 1 (0.8%) | 1 (0.1%) |
| - Femur fracture | 0 (0.0%) | 0 (0.0%) | 0 (0.0%) | 0 (0.0%) | 1 (0.8%) | 0 (0.0%) | 1 (0.1%) |
| - Abortion induced incomplete | 0 (0.0%) | 0 (0.0%) | 1 (0.8%) | 0 (0.0%) | 0 (0.0%) | 0 (0.0%) | 1 (0.1%) |
| **Nervous system disorders** | **0 (0.0%)** | **2 (1.6%)** | **2 (1.6%)** | **5 (4.0%)** | **0 (0.0%)** | **1 (0.8%)** | **10 (1.3%)** |
| - Seizure | 0 (0.0%) | 1 (0.8%) | 0 (0.0%) | 2 (1.6%) | 0 (0.0%) | 0 (0.0%) | 3 (0.4%) |
| - Transient ischaemic attack | 0 (0.0%) | 1 (0.8%) | 0 (0.0%) | 1 (0.8%) | 0 (0.0%) | 0 (0.0%) | 2 (0.3%) |
| - Idiopathic intracranial hypertension | 0 (0.0%) | 0 (0.0%) | 0 (0.0%) | 0 (0.0%) | 0 (0.0%) | 1 (0.8%) | 1 (0.1%) |
| - Facial paralysis | 0 (0.0%) | 0 (0.0%) | 1 (0.8%) | 0 (0.0%) | 0 (0.0%) | 0 (0.0%) | 1 (0.1%) |
| - Neuropathy peripheral | 0 (0.0%) | 0 (0.0%) | 1 (0.8%) | 0 (0.0%) | 0 (0.0%) | 0 (0.0%) | 1 (0.1%) |
| - Optic neuritis | 0 (0.0%) | 0 (0.0%) | 0 (0.0%) | 1 (0.8%) | 0 (0.0%) | 0 (0.0%) | 1 (0.1%) |
| - Brain injury | 0 (0.0%) | 0 (0.0%) | 0 (0.0%) | 1 (0.8%) | 0 (0.0%) | 0 (0.0%) | 1 (0.1%) |
| **Psychiatric disorders** | **1 (0.8%)** | **3 (2.5%)** | **1 (0.8%)** | **0 (0.0%)** | **3 (2.5%)** | **2 (1.6%)** | **10 (1.3%)** |
| - Suicide attempt | 0 (0.0%) | 1 (0.8%) | 0 (0.0%) | 0 (0.0%) | 3 (2.5%) | 1 (0.8%) | 5 (0.7%) |
| - Brief psychotic disorder without marked stressors | 1 (0.8%) | 0 (0.0%) | 0 (0.0%) | 0 (0.0%) | 0 (0.0%) | 0 (0.0%) | 1 (0.1%) |
| - Depression | 0 (0.0%) | 0 (0.0%) | 0 (0.0%) | 0 (0.0%) | 0 (0.0%) | 1 (0.8%) | 1 (0.1%) |
| - Drug dependence | 0 (0.0%) | 1 (0.8%) | 0 (0.0%) | 0 (0.0%) | 0 (0.0%) | 0 (0.0%) | 1 (0.1%) |
| - Psychotic disorder | 0 (0.0%) | 1 (0.8%) | 0 (0.0%) | 0 (0.0%) | 0 (0.0%) | 0 (0.0%) | 1 (0.1%) |
| - Anxiety disorder | 0 (0.0%) | 0 (0.0%) | 1 (0.8%) | 0 (0.0%) | 0 (0.0%) | 0 (0.0%) | 1 (0.1%) |
| **Hepatobiliary disorders** | **1 (0.8%)** | **1 (0.8%)** | **0 (0.0%)** | **1 (0.8%)** | **2 (1.7%)** | **2 (1.6%)** | **7 (0.9%)** |
| - Hepatotoxicity | 1 (0.8%) | 0 (0.0%) | 0 (0.0%) | 0 (0.0%) | 0 (0.0%) | 1 (0.8%) | 2 (0.3%) |
| - Drug-induced liver injury | 0 (0.0%) | 1 (0.8%) | 0 (0.0%) | 1 (0.8%) | 0 (0.0%) | 0 (0.0%) | 2 (0.3%) |
| - Hepatitis | 0 (0.0%) | 0 (0.0%) | 0 (0.0%) | 0 (0.0%) | 0 (0.0%) | 1 (0.8%) | 1 (0.1%) |
| - Acute on chronic liver failure | 0 (0.0%) | 0 (0.0%) | 0 (0.0%) | 0 (0.0%) | 1 (0.8%) | 0 (0.0%) | 1 (0.1%) |
| - Hyperbilirubinaemia | 0 (0.0%) | 0 (0.0%) | 0 (0.0%) | 0 (0.0%) | 1 (0.8%) | 0 (0.0%) | 1 (0.1%) |
| **Cardiac disorders** | **1 (0.8%)** | **2 (1.6%)** | **0 (0.0%)** | **2 (1.6%)** | **0 (0.0%)** | **1 (0.8%)** | **6 (0.8%)** |
| - Cardiac failure congestive | 1 (0.8%) | 1 (0.8%) | 0 (0.0%) | 0 (0.0%) | 0 (0.0%) | 0 (0.0%) | 2 (0.3%) |
| - Acute myocardial infarction | 0 (0.0%) | 1 (0.8%) | 0 (0.0%) | 1 (0.8%) | 0 (0.0%) | 0 (0.0%) | 2 (0.3%) |
| - Cardiac failure | 0 (0.0%) | 1 (0.8%) | 0 (0.0%) | 1 (0.8%) | 0 (0.0%) | 0 (0.0%) | 2 (0.3%) |
| - Cardiomyopathy alcoholic | 0 (0.0%) | 0 (0.0%) | 0 (0.0%) | 0 (0.0%) | 0 (0.0%) | 1 (0.8%) | 1 (0.1%) |

| *Event*<br>**System Organ Class (SOC)**<br>- Preferred Term (PT) | **9BLMZ**<br>**(n = 126)** | **9BCLLfxZ**<br>**(n = 122)** | **9BDLLfxZ**<br>**(n = 127)** | **9DCLLfxZ**<br>**(n = 124)** | **9DCMZ**<br>**(n = 120)** | **Control**<br>**(n = 126)** | **Total***<br>**(N = 745)** |
|---|---|---|---|---|---|---|---|
| **General disorders and administration site conditions** | **2 (1.6%)** | **0 (0.0%)** | **0 (0.0%)** | **2 (1.6%)** | **2 (1.7%)** | **0 (0.0%)** | **6 (0.8%)** |
| - Pyrexia | 1 (0.8%) | 0 (0.0%) | 0 (0.0%) | 0 (0.0%) | 0 (0.0%) | 0 (0.0%) | 1 (0.1%) |
| - Disease progression | 0 (0.0%) | 0 (0.0%) | 0 (0.0%) | 1 (0.8%) | 0 (0.0%) | 0 (0.0%) | 1 (0.1%) |
| - Death | 0 (0.0%) | 0 (0.0%) | 0 (0.0%) | 1 (0.8%) | 0 (0.0%) | 0 (0.0%) | 1 (0.1%) |
| - Chest pain | 0 (0.0%) | 0 (0.0%) | 0 (0.0%) | 0 (0.0%) | 1 (0.8%) | 0 (0.0%) | 1 (0.1%) |
| - Sudden death | 0 (0.0%) | 0 (0.0%) | 0 (0.0%) | 0 (0.0%) | 1 (0.8%) | 0 (0.0%) | 1 (0.1%) |
| - Generalised oedema | 1 (0.8%) | 0 (0.0%) | 0 (0.0%) | 0 (0.0%) | 0 (0.0%) | 0 (0.0%) | 1 (0.1%) |
| **Metabolism and nutrition disorders** | **1 (0.8%)** | **2 (1.6%)** | **0 (0.0%)** | **0 (0.0%)** | **2 (1.7%)** | **0 (0.0%)** | **5 (0.7%)** |
| - Diabetes mellitus inadequate control | 0 (0.0%) | 0 (0.0%) | 0 (0.0%) | 0 (0.0%) | 2 (1.7%) | 0 (0.0%) | 2 (0.3%) |
| - Hyperglycaemia | 0 (0.0%) | 2 (1.6%) | 0 (0.0%) | 0 (0.0%) | 0 (0.0%) | 0 (0.0%) | 2 (0.3%) |
| - Hypoalbuminaemia | 1 (0.8%) | 0 (0.0%) | 0 (0.0%) | 0 (0.0%) | 0 (0.0%) | 0 (0.0%) | 1 (0.1%) |
| **Skin and subcutaneous tissue disorders** | **0 (0.0%)** | **0 (0.0%)** | **0 (0.0%)** | **1 (0.8%)** | **3 (2.5%)** | **0 (0.0%)** | **4 (0.5%)** |
| - Drug reaction with eosinophilia and systemic symptoms | 0 (0.0%) | 0 (0.0%) | 0 (0.0%) | 1 (0.8%) | 1 (0.8%) | 0 (0.0%) | 2 (0.3%) |
| - Dermatitis exfoliative | 0 (0.0%) | 0 (0.0%) | 0 (0.0%) | 0 (0.0%) | 1 (0.8%) | 0 (0.0%) | 1 (0.1%) |
| - Swelling face | 0 (0.0%) | 0 (0.0%) | 0 (0.0%) | 0 (0.0%) | 1 (0.8%) | 0 (0.0%) | 1 (0.1%) |
| **Eye disorders** | **1 (0.8%)** | **2 (1.6%)** | **0 (0.0%)** | **0 (0.0%)** | **0 (0.0%)** | **1 (0.8%)** | **4 (0.5%)** |
| - Optic neuropathy | 1 (0.8%) | 1 (0.8%) | 0 (0.0%) | 0 (0.0%) | 0 (0.0%) | 0 (0.0%) | 2 (0.3%) |
| - Eye haemorrhage | 0 (0.0%) | 1 (0.8%) | 0 (0.0%) | 0 (0.0%) | 0 (0.0%) | 0 (0.0%) | 1 (0.1%) |
| - Cataract subcapsular | 0 (0.0%) | 0 (0.0%) | 0 (0.0%) | 0 (0.0%) | 0 (0.0%) | 1 (0.8%) | 1 (0.1%) |
| **Musculoskeletal and connective tissue disorders** | **0 (0.0%)** | **1 (0.8%)** | **0 (0.0%)** | **1 (0.8%)** | **2 (1.7%)** | **0 (0.0%)** | **4 (0.5%)** |
| - Arthralgia | 0 (0.0%) | 0 (0.0%) | 0 (0.0%) | 1 (0.8%) | 1 (0.8%) | 0 (0.0%) | 2 (0.3%) |
| - Arthropathy | 0 (0.0%) | 1 (0.8%) | 0 (0.0%) | 0 (0.0%) | 0 (0.0%) | 0 (0.0%) | 1 (0.1%) |
| - Osteoarthritis | 0 (0.0%) | 0 (0.0%) | 0 (0.0%) | 0 (0.0%) | 1 (0.8%) | 0 (0.0%) | 1 (0.1%) |
| **Vascular disorders** | **1 (0.8%)** | **1 (0.8%)** | **0 (0.0%)** | **1 (0.8%)** | **0 (0.0%)** | **0 (0.0%)** | **3 (0.4%)** |
| - Deep vein thrombosis | 0 (0.0%) | 0 (0.0%) | 0 (0.0%) | 1 (0.8%) | 0 (0.0%) | 0 (0.0%) | 1 (0.1%) |
| - Diabetic vascular disorder | 1 (0.8%) | 0 (0.0%) | 0 (0.0%) | 0 (0.0%) | 0 (0.0%) | 0 (0.0%) | 1 (0.1%) |
| - Hypertension | 0 (0.0%) | 1 (0.8%) | 0 (0.0%) | 0 (0.0%) | 0 (0.0%) | 0 (0.0%) | 1 (0.1%) |
| **Renal and urinary disorders** | **1 (0.8%)** | **0 (0.0%)** | **1 (0.8%)** | **0 (0.0%)** | **0 (0.0%)** | **0 (0.0%)** | **2 (0.3%)** |
| - Acute kidney injury | 1 (0.8%) | 0 (0.0%) | 0 (0.0%) | 0 (0.0%) | 0 (0.0%) | 0 (0.0%) | 1 (0.1%) |
| - Proteinuria | 0 (0.0%) | 0 (0.0%) | 1 (0.8%) | 0 (0.0%) | 0 (0.0%) | 0 (0.0%) | 1 (0.1%) |
| **Neoplasms benign, malignant and unspecified (incl cysts and polyps)** | **0 (0.0%)** | **0 (0.0%)** | **0 (0.0%)** | **0 (0.0%)** | **1 (0.8%)** | **1 (0.8%)** | **2 (0.3%)** |
| - Pancreatic carcinoma | 0 (0.0%) | 0 (0.0%) | 0 (0.0%) | 0 (0.0%) | 1 (0.8%) | 0 (0.0%) | 1 (0.1%) |
| - Lung neoplasm malignant | 0 (0.0%) | 0 (0.0%) | 0 (0.0%) | 0 (0.0%) | 0 (0.0%) | 1 (0.8%) | 1 (0.1%) |
| **Immune system disorders** | **0 (0.0%)** | **0 (0.0%)** | **2 (1.6%)** | **0 (0.0%)** | **0 (0.0%)** | **0 (0.0%)** | **2 (0.3%)** |

| Event<br>System Organ Class (SOC)<br>- Preferred Term (PT) | 9BLMZ<br>(n = 126) | 9BCLLfxZ<br>(n = 122) | 9BDLLfxZ<br>(n = 127) | 9DCLLfxZ<br>(n = 124) | 9DCMZ<br>(n = 120) | Control<br>(n = 126) | Total*<br>(N = 745) |
|---|---|---|---|---|---|---|---|
| - Hypersensitivity | 0 (0.0%) | 0 (0.0%) | 2 (1.6%) | 0 (0.0%) | 0 (0.0%) | 0 (0.0%) | 2 (0.3%) |
| **Product issues** | **1 (0.8%)** | **0 (0.0%)** | **0 (0.0%)** | **0 (0.0%)** | **0 (0.0%)** | **0 (0.0%)** | **1 (0.1%)** |
| - Device dislocation | 1 (0.8%) | 0 (0.0%) | 0 (0.0%) | 0 (0.0%) | 0 (0.0%) | 0 (0.0%) | 1 (0.1%) |
| **Pregnancy, puerperium and perinatal conditions** | **0 (0.0%)** | **0 (0.0%)** | **1 (0.8%)** | **0 (0.0%)** | **0 (0.0%)** | **0 (0.0%)** | **1 (0.1%)** |
| - Abortion threatened | 0 (0.0%) | 0 (0.0%) | 1 (0.8%) | 0 (0.0%) | 0 (0.0%) | 0 (0.0%) | 1 (0.1%) |

**Total by preferred term (PT) represents the sum of participants who experienced at least one event corresponding to that PT. If they experienced more than one event of the same PT, it is counted only once. If a participant experienced more than one unique PT within a system organ class (SOC), each unique PT counts toward the PT total, but only one event counts toward the SOC.*

***Table S24. Number (%) of participants with adverse events of special interest (AESI) of grade 3 or higher and time to onset by Week 73 (Safety population)***

| Adverse events (AEs) | 9BLMZ<br>(n = 126) | 9BCLLfxZ<br>(n = 122) | 9BDLLfxZ<br>(n = 127) | 9DCLLfxZ<br>(n = 124) | 9DCMZ<br>(n = 120) | Control<br>(n = 126) | Total<br>(N = 745) |
|---|---|---|---|---|---|---|---|
| - AESI: ≥ grade 3 hematologic toxicity | 11 (8.7%) | 9 (7.4%) | 10 (7.9%) | 13 (10.5%) | 9 (7.5%) | 13 (10.3%) | 65 (8.7%) |
| *Days to event, median [IQR]* | *29 [26-109]* | *80 [56-171]* | *67.5 [28-82]* | *83 [61.0-98.0]* | *145 [88-205]* | *112 [28-269]* | *83.0 [28-171.0]* |
| - AESI: ≥ grade 3 peripheral neuropathy | 4 (3.2%) | 5 (4.1%) | 9 (7.1%) | 3 (2.4%) | 3 (2.5%) | 6 (4.8%) | 30 (4.0%) |
| *Days to event, median [IQR]* | *162.5 [144-220]* | *83 [50-137]* | *166 [125-220]* | *119 [112-329]* | *57 [34-139]* | *218.5 [178-248]* | *162 [112-220]* |
| - AESI: ≥ grade 3 hepatotoxicity | 23 (18.3%) | 17 (13.9%) | 8 (6.3%) | 18 (14.5%) | 12 (10.0%) | 9 (7.1%) | 87 (11.7%) |
| *Days to event, median [IQR]* | *56 [41-154]* | *82 [55-139]* | *41 [25.5-56]* | *81.5 [40-200]* | *59.5 [26-167]* | *84 [40-269]* | *58 [40-166]* |
| - AESI: ≥ grade 3 optic neuropathy | 0 (0.0%) | 1 (0.8%) | 0 (0.0%) | 1 (0.8%) | 0 (0.0%) | 2 (1.6%) | 4 (0.5%) |
| *Days to event, median [IQR]* | - | *65 [65-65]* | - | *106 [106-106]* | - | *106.5 [57-156]* | *85.5 [61-131]* |
| - AESI: ≥ grade 3 QT prolongation | 0 (0.0%) | 4 (3.3%) | 0 (0.0%) | 0 (0.0%) | 5 (4.2%) | 0 (0.0%) | 9 (1.2%) |
| *Days to event, median [IQR]* | - | *169 [119-244.5]* | - | - | *222.0 [145-223]* | - | *177 [145-223]* |

***Table S25. Adverse events of special interest (AESI) of grade 3 or higher and time to onset by Week 104 (Safety population)***

| **Adverse events (AEs)** | **9BLMZ (n = 126)** | **9BCLLfxZ (n = 122)** | **9BDLLfxZ (n = 127)** | **9DCLLfxZ (n = 124)** | **9DCMZ (n = 120)** | **Control (n = 126)** | **Total (N = 745)** |
|---|---|---|---|---|---|---|---|
| - AESI: ≥ grade 3 hematologic toxicity | 11 (8.7%) | 10 (8.2%) | 10 (7.9%) | 13 (10.5%) | 9 (7.5%) | 14 (11.1%) | 67 (9.0%) |
| *Days to event, median [IQR]* | *29 [26-109]* | *123 [56-194]* | *67.5 [28-82]* | *83 [61.0-98.0]* | *145 [88-205]* | *148 [28-286]* | *83 [28-193]* |
| - AESI: ≥ grade 3 peripheral neuropathy | 4 (3.2%) | 5 (4.1%) | 9 (7.1%) | 3 (2.4%) | 3 (2.5%) | 6 (4.8%) | 30 (4.0%) |
| *Days to event, median [IQR]* | *162.5 [144-220]* | *83 [50-137]* | *166 [125-220]* | *119 [112-329]* | *57 [34-139]* | *218.5 [178-248]* | *162 [112-220]* |
| - AESI: ≥ grade 3 hepatotoxicity | 23 (18.3%) | 19 (15.6%) | 11 (8.7%) | 18 (14.5%) | 12 (10.0%) | 9 (7.1%) | 92 (12.4%) |
| *Days to event, median [IQR]* | *56 [41-154]* | *110 [55-221]* | *56 [26-509]* | *81.5 [40-200]* | *59.5 [26-167]* | *84 [40-269]* | *71 [40.5-185]* |
| - AESI: ≥ grade 3 optic neuropathy | 0 (0.0%) | 1 (0.8%) | 0 (0.0%) | 1 (0.8%) | 0 (0.0%) | 2 (1.6%) | 4 (0.5%) |
| *Days to event, median [IQR]* | - | *65 [65-65]* | - | *106 [106-106]* | - | *106.5 [57-156]* | *85.5 [61-131]* |
| - AESI: ≥ grade 3 QT prolongation | 0 (0.0%) | 4 (3.3%) | 0 (0.0%) | 0 (0.0%) | 5 (4.2%) | 0 (0.0%) | 9 (1.2%) |
| *Days to event, median [IQR]* | - | *169 [119-244.5]* | - | - | *222.0 [145-223]* | - | *177 [145-223]* |

***Table S26. Line Listing of Deaths in endTB Trial by Week 104 (Safety population)***

| N | endTB arm | Age Group | Sex | Preferred term | Causality according to investigators | Causality according to the sponsor |
|---|---|---|---|---|---|---|
| 1 | 9BLMZ | 30-39 | M | *Pneumocystis jirovecii* pneumonia | Not related | Not related |
| 2 | 9BLMZ | 50-59 | M | Disease progression | Not related | Not related |
| 3 | 9BLMZ | 50-59 | F | Septic shock | Not related | Not related |
| 4 | 9BCLLfxZ | 60-69 | M | Alcohol poisoning | Not related | Not related |
| 5 | 9BDLLfxZ | 30-39 | F | Respiratory failure | Not related | Not related |
| 6 | 9BDLLfxZ | 70+ | F | Respiratory failure | Not related | Not related |
| 7 | 9BDLLfxZ | 50-59 | M | Acute abdomen and acute respiratory failure | Not related | Not related |
| 8 | 9DCLLfxZ | 40-49 | M | Respiratory failure | Not related | Not related |
| 9 | 9DCLLfxZ | 20-29 | M | Disease progression | Not related | Not related |
| 10 | 9DCLLfxZ | 60-69 | M | Sepsis | Not related | Not related |
| 11 | 9DCLLfxZ | 50-59 | M | Death (unknown cause) | Not related | Not related |
| 12 | 9DCMZ | 50-59 | M | Sepsis | Not related | Not related |
| 13 | 9DCMZ | 30-39 | M | Death (unknown cause) | Not related | Related |
| 14 | 9DCMZ | 50-59 | M | Pancreatic carcinoma | Not related | Not related |
| 15 | Control | 30-39 | M | Alcohol poisoning | Not related | Not related |
| 16 | Control | 40-49 | M | Pneumonia | Not related | Not related |
| 17 | Control | 50-59 | M | Pancreatitis acute | Not related | Not related |
| 18 | Control | 30-39 | M | Cardiomyopathy alcoholic | Not related | Not related |

***Table S27. Frequency of endTB trial Participants Experiencing Adverse Events Leading to (and Time to) Permanent Discontinuation of at Least One Study Drug by Treatment Group by Week 73 (Safety Population)***

| Adverse events (AEs) | 9BLMZ (n = 126) | 9BCLLfxZ (n = 122) | 9BDLLfxZ (n = 127) | 9DCLLfxZ (n = 124) | 9DCMZ (n = 120) | Control (n = 126) | Total (N = 745) |
|---|---|---|---|---|---|---|---|
| Participants with at least one AE leading to permanent discontinuation ≥1 drug | 29 (23.0%) | 32 (26.2%) | 41 (32.3%) | 32 (25.8%) | 22 (18.3%) | 54 (42.9%) | 210 (28.2%) |
| *Days to drug discontinuation, median [IQR]* | 105 [77-215] | 134 [76-217] | 118 [61-182] | 115.5 [58-174] | 100 [41-217] | 158.5 [56-281] | 122.5 [61-215] |
| - **Pyrazinamide** | **24 (19.1%)** | **23 (18.9%)** | **24 (18.9%)** | **18 (14.5%)** | **18 (15.0%)** | **14 (11.1%)** | **121 (16.2%)** |
| *Days to drug discontinuation, median [IQR]* | 101 [67.5-225.5] | 112 [72-199] | 97.5 [48.5-178.5] | 84 [37-153] | 83 [41-173] | 56.5 [38-187] | 92 [53-188] |
| - **Linezolid** | **9 (7.1%)** | **11 (9.0%)** | **20 (15.8%)** | **20 (16.1%)** | **-** | **23 (18.3%)** | **83 (11.1%)** |
| *Days to drug discontinuation, median [IQR]* | 131 [73-171] | 213 [112-239] | 120.5 [59.5-190] | 114.5 [54-182.5] | - | 195 [129-319] | 155.0 [80-224] |
| - **Clofazimine** | **-** | **6 (4.9%)** | **-** | **9 (7.3%)** | **8 (6.7%)** | **10 (7.9%)** | **33 (4.4%)** |
| *Days to drug discontinuation, median [IQR]* | - | 214 [163-259] | - | 49 [31-168] | 76.5 [39.5-229.5] | 340.5 [217-440] | 192.0 [50-284] |
| - **Levofloxacin** | **-** | **5 (4.1%)** | **3 (2.4%)** | **9 (7.3%)** | **1 (0.8%)** | **16 (12.7%)** | **34 (4.6%)** |
| *Days to drug discontinuation, median [IQR]* | - | 236 [213-252] | 32 [26-69] | 49 [31-178] | 83 [83-83] | 223.5 [111-439] | 168 [49-273] |
| - **Bedaquiline** | **5 (4.0%)** | **4 (3.3%)** | **3 (2.4%)** | **-** | **-** | **8 (6.4%)** | **20 (2.7%)** |
| *Days to drug discontinuation, median [IQR]* | 91 [58-156] | 223 [153.5-279.5] | 32 [26-69] | - | - | 315.5 [110-440] | 162.5 [55.5-279.5] |
| - **Moxifloxacin** | **7 (5.6%)** | **-** | **-** | **-** | **10 (8.3%)** | **1 (0.8%)** | **18 (2.4%)** |
| *Days to drug discontinuation, median [IQR]* | 90 [53-156] | - | - | - | 100 [41-242] | 23 [23-23] | 93.5 [41-217] |
| - **Delamanid** | **-** | **-** | **3 (2.4%)** | **8 (6.5%)** | **7 (5.8%)** | **0 (0.0%)** | **18 (2.4%)** |

| Adverse events (AEs) | 9BLMZ (n = 126) | 9BCLLfxZ (n = 122) | 9BDLLfxZ (n = 127) | 9DCLLfxZ (n = 124) | 9DCMZ (n = 120) | Control (n = 126) | Total (N = 745) |
|---|---|---|---|---|---|---|---|
| *Days to drug discontinuation, median [IQR]* | - | - | 32 [26-69] | 47 [31-185] | 50 [38-217] | - | 47 [31-103] |

***Table S28. Participant pregnancies, exposure information, and treatment decision during the endTB Trial***

| N | Arm | Age Group | Exposure information & timing | Participant treatment/study decision | Time-to-onset days) |
|---|---|---|---|---|---|
| 1 | 9BDLLfxZ | 20-29 | Maternal exposure during pregnancy from 1st trimester | Early discontinued | 103 |
| 2 | 9BLMZ | 20-29 | Maternal exposure during pregnancy from 1st trimester | Early discontinued | 224 |
| 3 | 9BDLLfxZ | 20-29 | Maternal exposure during pregnancy from 1st trimester | Maintained in the study | 127 |
| 4 | 9BDLLfxZ | 20-29 | Maternal exposure during pregnancy from 1st trimester | Maintained in the study | 77 |
| 5 | 9BDLLfxZ | 20-29 | Maternal exposure during pregnancy from 1st trimester | Maintained in the study | 63 |
| 6 | 9BDLLfxZ | 30-39 | Maternal exposure before pregnancy | Maintained in the study | 497 |
| 7 | Control | 30-39 | Maternal exposure during pregnancy from 1st trimester | Maintained in the study | 66 |
| 8 | 9BLMZ | 30-39 | Maternal exposure before pregnancy | Study completed | ~665 |
| 9 | 9DCMZ | 30-39 | Maternal exposure before pregnancy | Maintained in the study | 566 |
| 10 | 9BCLlfxZ | 20-29 | Maternal exposure during pregnancy from 1st trimester | Maintained in the study | 280 |
| 11 | Control | 30-39 | Maternal exposure during pregnancy from 1st trimester | Maintained in the study | 129 |
| 12 | 9DCLLfxZ | 30-39 | Maternal exposure before pregnancy | Maintained in the study | 701 |
| 13 | 9DCLLfxZ | 20-29 | Maternal exposure during pregnancy from 1st trimester | Maintained in the study | 150 |
| 14 | 9DCLLfxZ | 20-29 | Maternal exposure before pregnancy | Maintained in the study | 607 |
| 15 | 9DCMZ | 20-29 | Maternal exposure before pregnancy | Maintained in the study | 418 |
| 16 | Control | 20-29 | Maternal exposure before pregnancy | Maintained in the study | 350 |
| 17 | Control | 20-29 | Maternal exposure before pregnancy | Maintained in the study | 473 |
| 18 | Control | 40-49 | Maternal exposure during pregnancy from 1st trimester | Maintained in the study | 422 |
| 19 | 9DCMZ | 20-29 | Maternal exposure before pregnancy | Maintained in the study | 569 |
| 20 | 9BDLLfxZ | 20-29 | Maternal exposure before pregnancy | Maintained in the study | 487 |
| 21 | 9BDLLfxZ | 20-29 | Maternal exposure before pregnancy | Maintained in the study | 582 |
| 22 | Control | 20-29 | Paternal exposure before pregnancy | Maintained in the study | 374 |
| 23 | 9DCLLfxZ | Unknown | Paternal exposure before pregnancy | Maintained in the study | Unknown |
| 24 | 9BDLLfxZ | Unknown | Paternal exposure before pregnancy | Maintained in the study | Unknown |
| 25 | 9DCMZ | Unknown | Paternal exposure before pregnancy | Maintained in the study | 652 |
| 26 | 9DCLLfxZ | 15-19 | Paternal exposure before pregnancy | Maintained in the study | 388 |
| 27 | 9DCLLfxZ | 15-19 | Paternal exposure before pregnancy | Maintained in the study | 146 |

| N | Arm | Age Group | Exposure information & timing | Participant treatment/study decision | Time-to-onset days) |
|---|---|---|---|---|---|
| 28 | 9BLMZ | Unknown | Paternal exposure before pregnancy | Maintained in the study | 256 |
| 29 | Control | Unknown | Paternal exposure before pregnancy | Maintained in the study | 78 |
| 30 | 9BDLLfxZ | Unknown | Paternal exposure before pregnancy | Maintained in the study | 11 |
| 31 | Control | Unknown | Paternal exposure before pregnancy | Maintained in the study | 444 |
| 32 | 9DCLLfxZ | 20-29 | Paternal exposure before pregnancy | Maintained in the study | 403 |
| 33 | 9BLMZ | Unknown | Paternal exposure before pregnancy | Maintained in the study | 582 |

[1] Wason, J.M.S. & Trippa, L. A comparison of Bayesian adaptive randomization and multi-stage designs for multi-arm clinical trials. *Stat. Med.* 33, 2206-2221 (2014).

[2] Nunn, A, Phillips, P. & Mitchison, D. Timing of relapse in short-course chemotherapy trials for tuberculosis. *Int. J. Tuberc. Lung Dis.* 14, 241-2 (2010).
